# Systematic review of the prevalence of Long Covid

**DOI:** 10.1101/2022.11.06.22281979

**Authors:** Mirembe Woodrow, Charles Carey, Nida Ziauddeen, Rebecca Thomas, Athena Akrami, Vittoria Lutje, Darren C Greenwood, Nisreen A Alwan

**Affiliations:** School of Primary Care, Population Sciences and Medical Education, Faculty of Medicine, University of Southampton, Southampton, UK; Manchester University NHS Foundation Trust and The University of Manchester, Manchester, UK; School of Primary Care, Population Sciences and Medical Education, Faculty of Medicine, University of Southampton, Southampton, UK and NIHR Applied Research Collaboration Wessex, Southampton, UK; University of Liverpool, Liverpool, UK; Sainsbury Wellcome Centre, University College London, London, UK and Patient-led Research Collaborative, Washington DC, USA; Cochrane Infectious Diseases Group, Liverpool, UK; School of Medicine, University of Leeds, UK; School of Primary Care, Population Sciences and Medical Education, Faculty of Medicine, University of Southampton, Southampton, UK, NIHR Applied Research Collaboration Wessex, Southampton, UK and NIHR Southampton Biomedical Research Centre, University of Southampton and University Hospital Southampton NHS Foundation Trust, Southampton, UK

**Author notes:** Correspondence to: Prof Nisreen A Alwan, School of Primary Care, Population Sciences and Medical, Education, Faculty of Medicine, University of Southampton, Southampton, UK,; Alternate corresponding author: Mirembe Woodrow, School of Primary Care, Population Sciences and Medical Education, Faculty of Medicine, University of Southampton, Southampton, UK. Equal contribution as senior authors (Darren C Greenwood, Nisreen A Alwan).

**Keywords:** Long Covid, Systematic Review, Prevalence, SARSCoV2

## Abstract

**Background:** Long Covid occurs in those infected with SARSCoV2 whose symptoms persist or develop beyond the acute phase. We conducted a systematic review to determine the prevalence of persistent symptoms, functional disability or pathological changes in adults or children at least 12 weeks post-infection.

**Methods:** We searched key registers and databases from 1^st^ January 2020 to 2^nd^ r 2021, limited to publications in English and studies with at least 100 participants. Studies where all participants were critically ill were excluded. Long Covid was extracted as prevalence of at least one symptom or pathology, or prevalence of the most common symptom or pathology, at 12 weeks or later. Heterogeneity was quantified in absolute terms and as a proportion of total variation and explored across pre-defined subgroups (PROSPERO ID CRD42020218351).

**Results:** 120 studies in 130 publications were included. Length of follow-up varied between 12 weeks - 12 months. Few studies had low risk of bias. All complete and subgroup analyses except one had I^2^ ≥ 90%, with prevalence of persistent symptoms range of 0% - 93% (pooled estimate 42.1%, 95% prediction interval : 6.8% to 87.9%). Studies using routine healthcare records tended to report lower prevalence of persistent symptoms/pathology than self-report. However, studies systematically investigating pathology in all participants at follow up tended to report the highest estimates of all three. Studies of hospitalised cases had generally higher estimates than community-based studies.

**Conclusions:** The way in which Long Covid is defined and measured affects prevalence estimation. Given the widespread nature of SARSCoV2 infection globally, the burden of chronic illness is likely to be substantial even using the most conservative estimates.

Funding this systematic review received no specific funding.

**Key points:** In a systematic review of 130 publications, prevalence estimates of Long Covid (>12 weeks) after SARSCoV2 infection differed according to how persistent symptoms/pathology were identified and measured, and ranged between 0% - 93% (pooled estimate 42.1%, 95% prediction interval: 6.8% to 87.9%).

## Introduction

Long Covid is the state of not fully recovering for many weeks, months or years after contracting SARSCoV2 infection. The World Health Organization (WHO) defines Post COVID-19 Condition (Long Covid) as the condition occurring in individuals with a history of probable or confirmed SARSCoV2 infection 3 months after the onset with symptoms that last at least 2 months, cannot be explained by an alternative diagnosis and generally impacts everyday functioning(1). These symptoms may be the same as the acute illness or new symptoms developing weeks or months after the acute phase. Clinical guidelines(2, 3) in the UK and the US consider Long Covid as symptoms ongoing for four weeks or more.

Long Covid can occur across the spectrum of severity of initial infection(4). A wide range of symptoms have been reported with exhaustion, breathlessness, muscle aches, cognitive dysfunction, headache, palpitations, dizziness and chest tightness or heaviness amongst the most common(5, 6). Patients are still struggling to access adequate recognition, support, medical assessment and treatment(7, 8).

Studies assessing the prevalence of Long Covid have produced wide-ranging results due to varying settings, case definitions, population denominators and methods of ascertainment. For the purposes of this review, we define Long Covid as persistent (constant, fluctuating or relapsing) symptoms and/or functional disability and/or the development of new pathology following SARSCoV2 infection for equal or more than 12 weeks from onset of symptoms or from time of diagnosis, in people where the infection is self-described, clinically diagnosed, and/or diagnosed through a laboratory test.

We aimed to systematically collate, appraise and synthesise studies that describe the prevalence of Long Covid and to characterise its typology including patient demographics, symptoms/function disability and pathology.

## Methods

### Search strategy and selection criteria

Included study designs were cohort, cross-sectional and case control studies with an estimate of the denominator where participants were followed-up/assessed at a minimum of 12 weeks post-infection. Studies were restricted to those published in English between 1^st^ January 2020 and 2^nd^ November 2021, including peer-reviewed articles, online reports, letters, and preprints. Only studies with a sample size of 100 or more participants (at the time of follow-up assessment if longitudinal study) were included (50 or more per subgroup).

Studies of adults and children with a confirmed or probable SARSCoV2 infection in any age group (as defined by each study) were included. The control group in studies that included one is individuals with a confirmed or probable case of SARSCoV2 infection (as defined by the study) who have recovered (duration as defined by study as long as under 12 weeks from symptom onset or confirmation of infection) and have no new pathology attributed to SARSCoV2 infection. Studies that compared population-based prevalence as the control arm were excluded from the control analysis.

Community-based, hospital-based, and mixed studies were all included, apart from studies that only reported outcomes for critically ill patients admitted to intensive care, because this review did not aim to estimate delayed recovery following ICU admission (post-ICU syndrome). Patients who were not hospitalised within two weeks of symptom onset but were subsequently hospitalised were counted as non-hospitalised for the purpose of this review.

A systematic search was conducted using MEDLINE (Ovid), Embase (Ovid), the Cochrane Covid-19 Study register (www.covid-19.cochrane.org; includes Cochrane Central Register of Controlled Trials (CENTRAL), WHO International Clinical Trials Registry Platform (ICTRP), medRxiv, Cochrane CENTRAL, MEDLINE (PubMed), ClinicalTrials.gov, and the WHO Global research on coronavirus disease (COVID- 19) database(9). The initial search was run on 13 November 2020 and updated on 2 November 2021, both by VL. An example of the search strategy applied to Medline is provided in the Supplementary material; it was adapted for other databases as needed.

The screening management software Covidence was used to screen for eligibility. All articles were screened independently by two reviewers at each stage (title, abstract, and full text) with any discrepancies resolved by NAA. This review is reported in line with PRISMA guidelines(10). The protocol was published on the international prospective register of international reviews, PROSPERO (CRD42020218351): https://www.crd.york.ac.uk/prospero/display_record.php?RecordID=218351.

### Data analysis

Data for each study was extracted independently by two of four reviewers (MW, DCG, CC, NZ). Any discrepancies were resolved by consensus between the two reviewers for each study or by a third reviewer (NAA). Where multiple publications were identified as originating from the same study, all data was extracted but each data point was only used once in the analysis. In addition to excluding duplicate reports, or duplicate results from the same study, a number of general decisions were made to cope with multiple publications from the same study, either focusing on different lengths of follow-up, different timepoints, or different subgroups. These were guided by principles of (1) avoiding double counting individuals, (2) using the most appropriate outcome, for example, general Long Covid definition, in the broadest group such as the widest population, largest sample, most recent update, (3) unless stratifying by length of follow-up, we took the earliest and/or most complete follow-up as the main result.

The primary outcome is Long Covid, defined as non-recovery from COVID-19, according to symptoms, functional ability or pathology. SARSCoV2 infection can be confirmed, probable or suspected with prolonged symptoms (including but not limited to those explicitly defined as ‘new onset’), functional disability or pathology for equal to or more than 12 weeks from onset of symptoms or positive test date (as defined by the study). Secondary outcomes included the demographics of people with Long Covid in relation to each study’s denominator, prevalence of specific persistent or relapsing symptoms, prevalence of functional disability, and the characterisation of post-COVID-19 pathology.

A Long Covid-specific risk of bias tool was developed, based on the Newcastle-Ottawa scale, but tailored to the relevant sources of bias. The domains used are reported in Supplementary Table 3. Risk of bias was particularly assessed in relation to the denominator, how the symptoms were assessed (active or passive elicitation of the symptoms) and hospital stay. Subgroup analysis by risk of bias was performed. In studies where follow-up was measured post-hospital admission or discharge, symptom onset was estimated to have been 7 or 14 days prior to discharge respectively and estimated as 21 days if follow-up was measured from a post-infection negative test.

The prevalence was extracted as cumulative incidence. In extracting the prevalence of persistent symptoms, we used either prevalence of at least one symptom or pathology, or the prevalence of the most common symptom/pathology, depending on the data reported by the study. Data for each symptom was extracted separately in studies that reported on the prevalence of individual symptoms but did not provide an overall estimate of prevalence of Long Covid. We used the symptom with the highest estimate as our best estimate of overall prevalence, though it is likely to be an underestimate of actual prevalence. In studies with controls, the prevalence of the same symptom was used for comparison. Where length of follow-up varied between study participants, we report a measure of average (e.g. mean or median) length of follow-up, or the midpoint of the reported range.

All analysis was conducted in Stata version 17(11). The distribution, prevalence estimates, numerators, denominators, and assessment time points in different populations was qualitatively summarised. We used random-effects meta-analysis on the logit of the proportions to ensure estimates and confidence limits did not go below 0% or over 100%, transforming back to the original scale for presentation.

The heterogeneity was quantified both in absolute terms (range of individual study estimates) and as a proportion of total variation (I^2^), and explored across pre-defined subgroups described below. In a variation to our protocol, we present pooled estimates alongside 95% prediction intervals to evaluate and incorporate uncertainty in the analysis, as recently recommended for prevalence studies, where true between-study heterogeneity is expected(12, 13) . Heterogeneity was explored by stratifying on pre-defined subgroups: outcome type (pathology, symptom, functional status), geographical region (China, Europe, North America, Mixed and other), source of sample (community, healthcare workers, outpatients, hospital inpatients), length of follow-up, study design, confirmed diagnosis, and other risk of bias domains. We also stratified by severity score based on the WHO Clinical Progression Scale [supplemental methods].

Potential small study effects such as publication bias were investigated using contour-enhanced funnel plots and Egger’s test of funnel plot asymmetry.

**Role of funding source** None

## Results

### Literature search

The searches found 11,518 studies in total. After deduplication and title and abstract screening, 457 full text studies were assessed for eligibility. Hand-searching sourced an additional 9 studies and in total 130 publications were included, 120 of which were discrete studies (Figure 1). 24 studies were conducted in China (including Hong Kong), 66 in Europe, 14 in North America and 16 in various other countries(14–143). Reasons for exclusion are listed in Supplementary Table 1.

**Figure 1:**
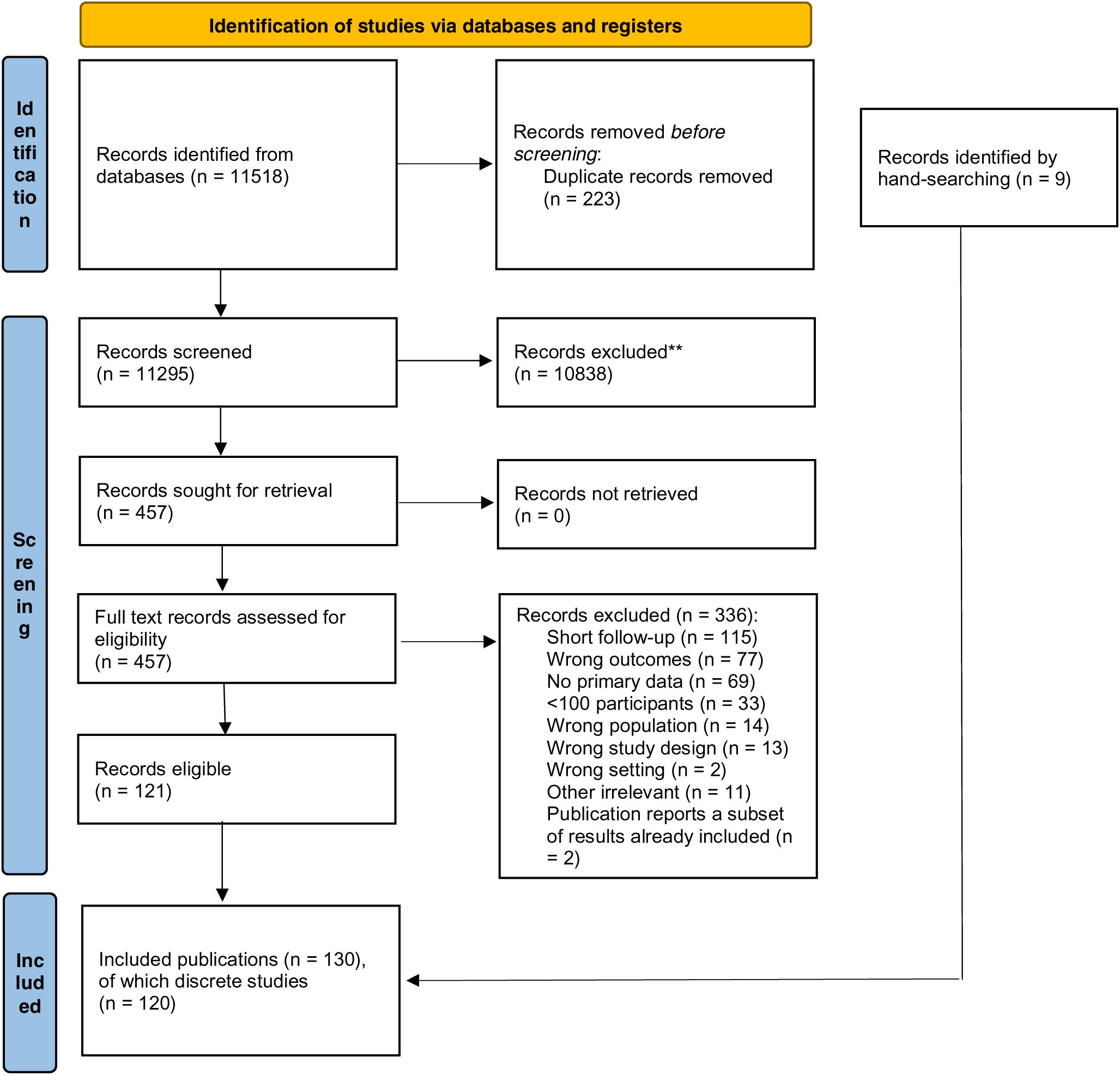
Study selection

Table 1 summarises the included studies’ key characteristics and primary outcome for the first follow-up. Study design was reported as described by each study or designated based on study description if not explicitly stated. Most studies were in adults and included patients who were hospitalised in the acute phase (24 studies with <10% of the sample hospitalised in the acute phase).

**Table 1.**
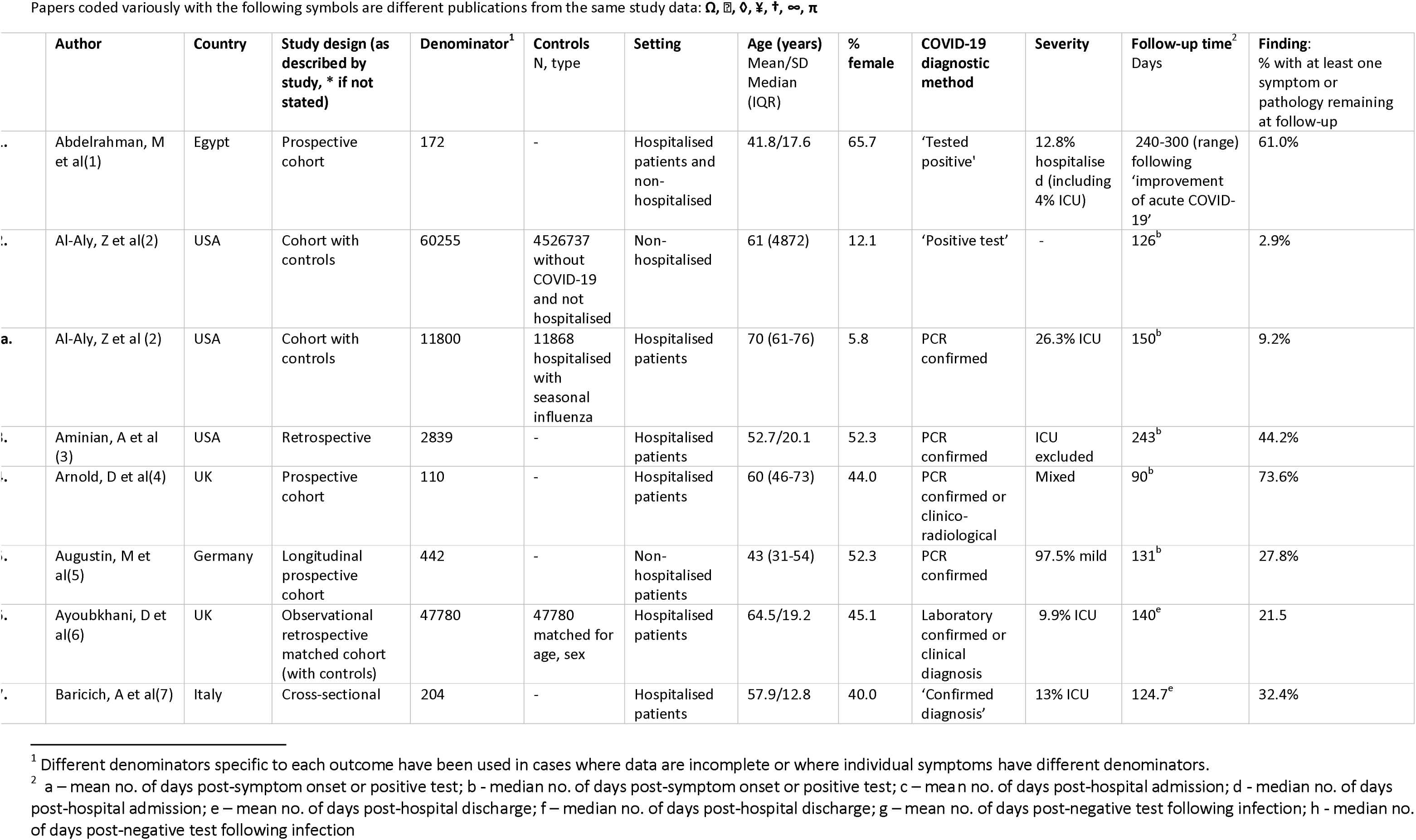
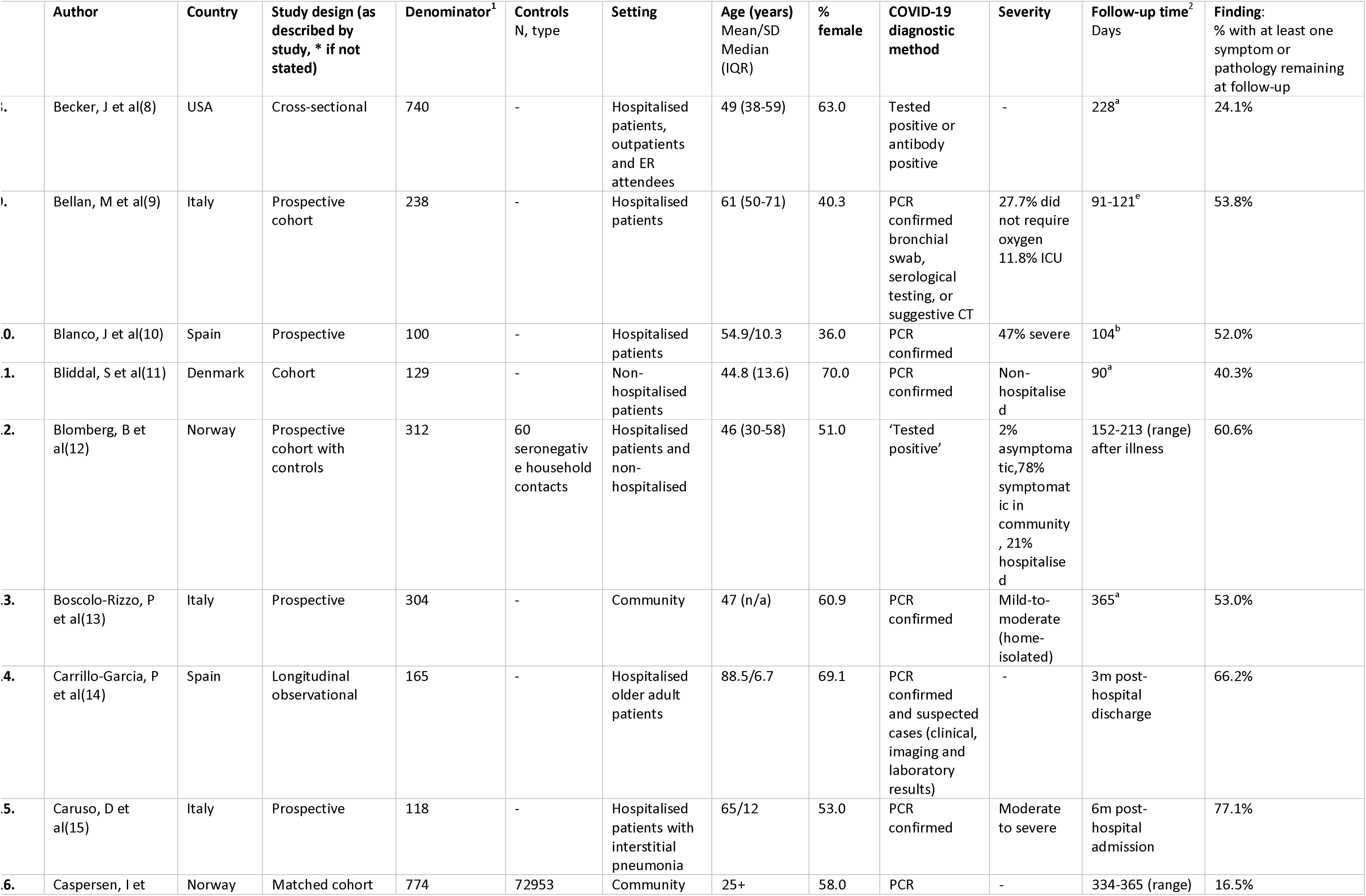
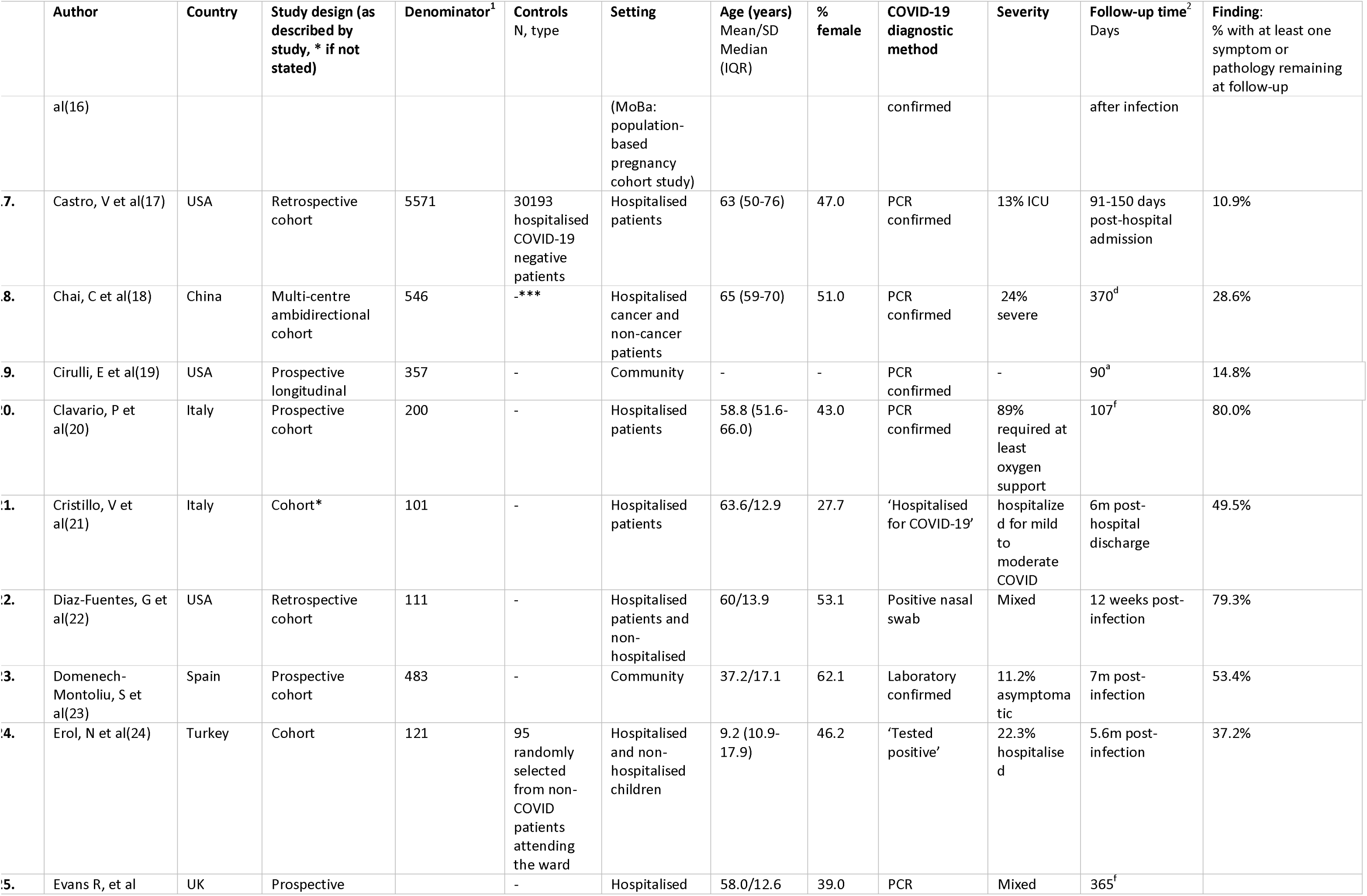
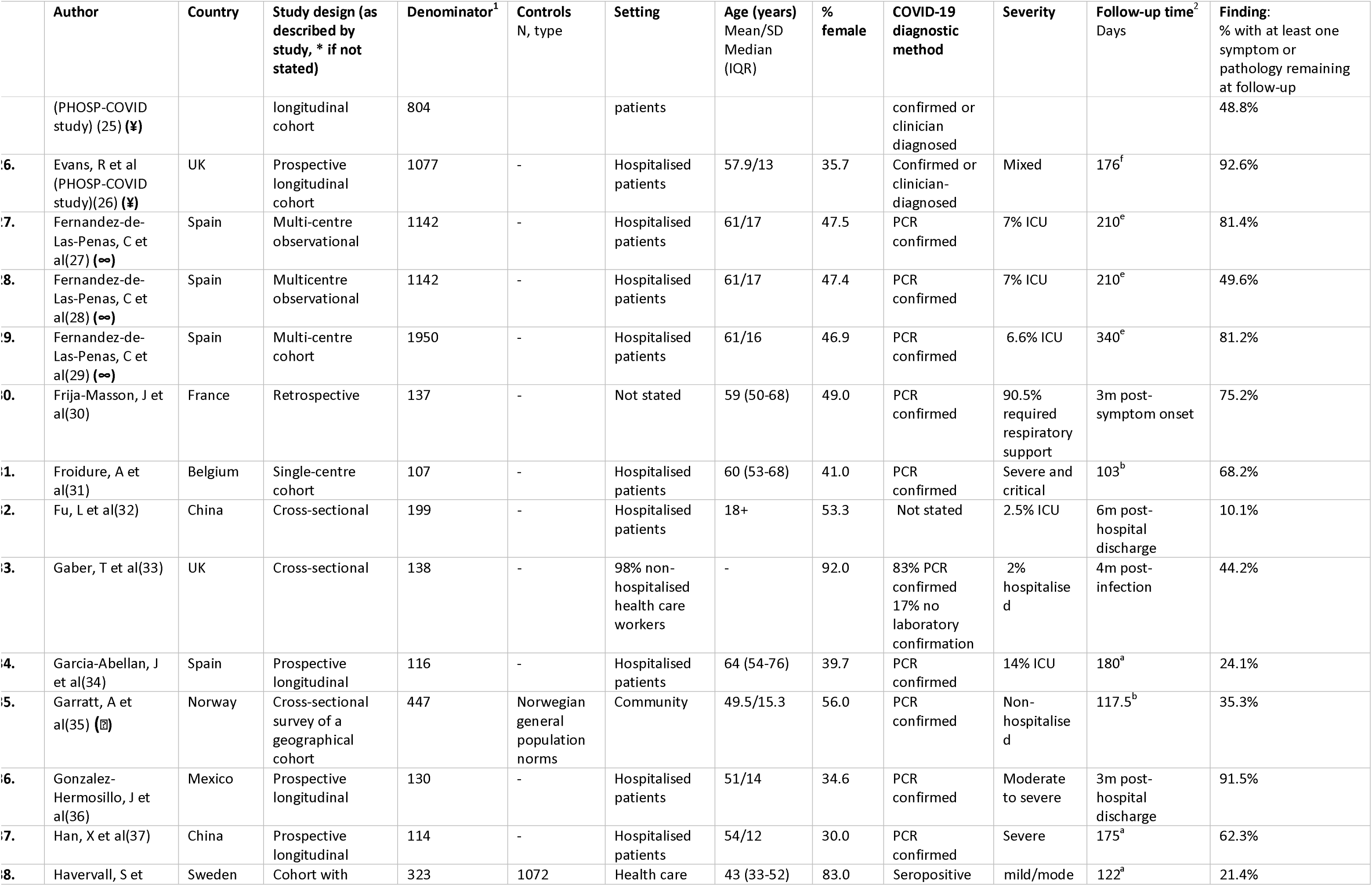
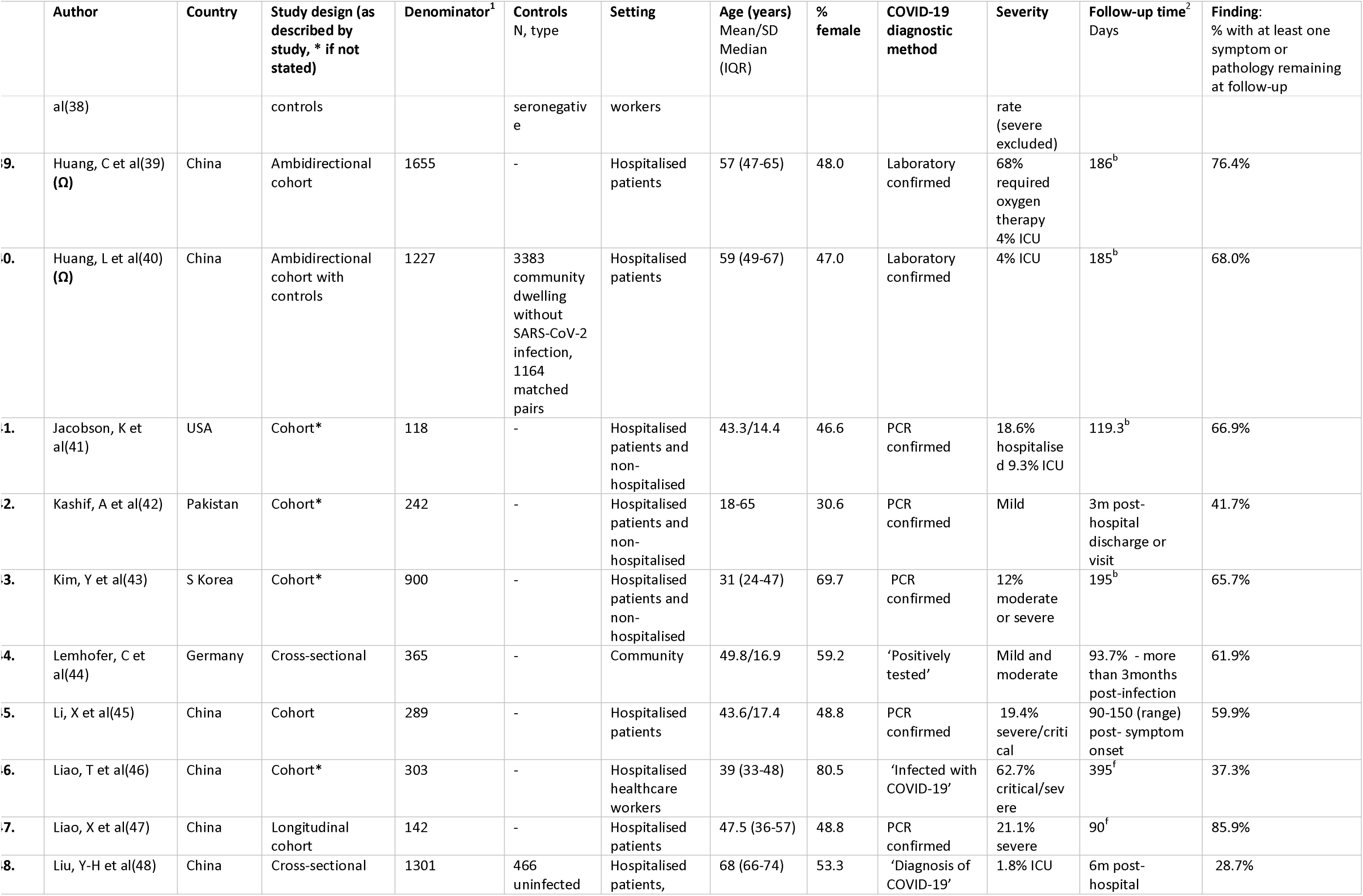
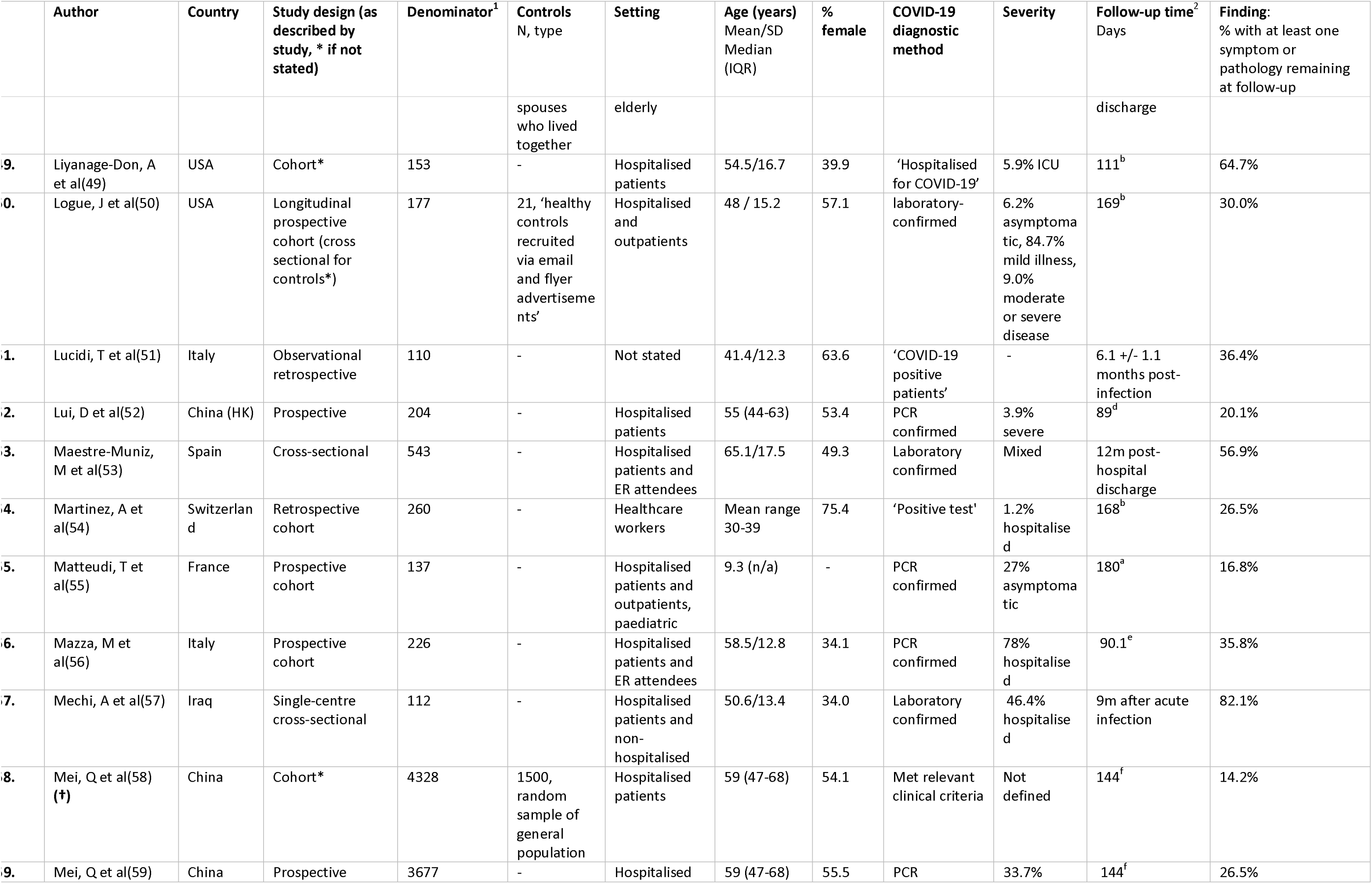
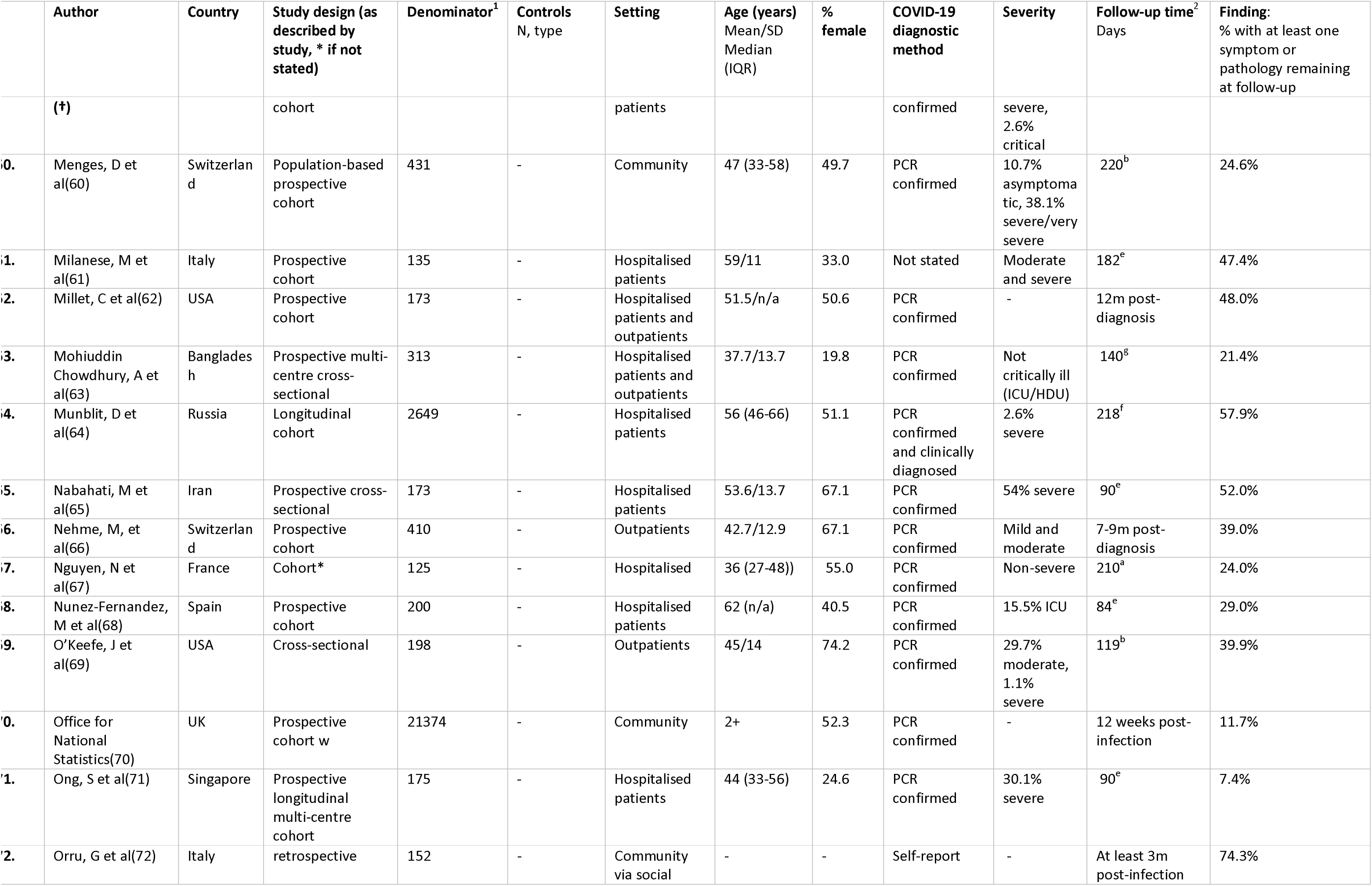
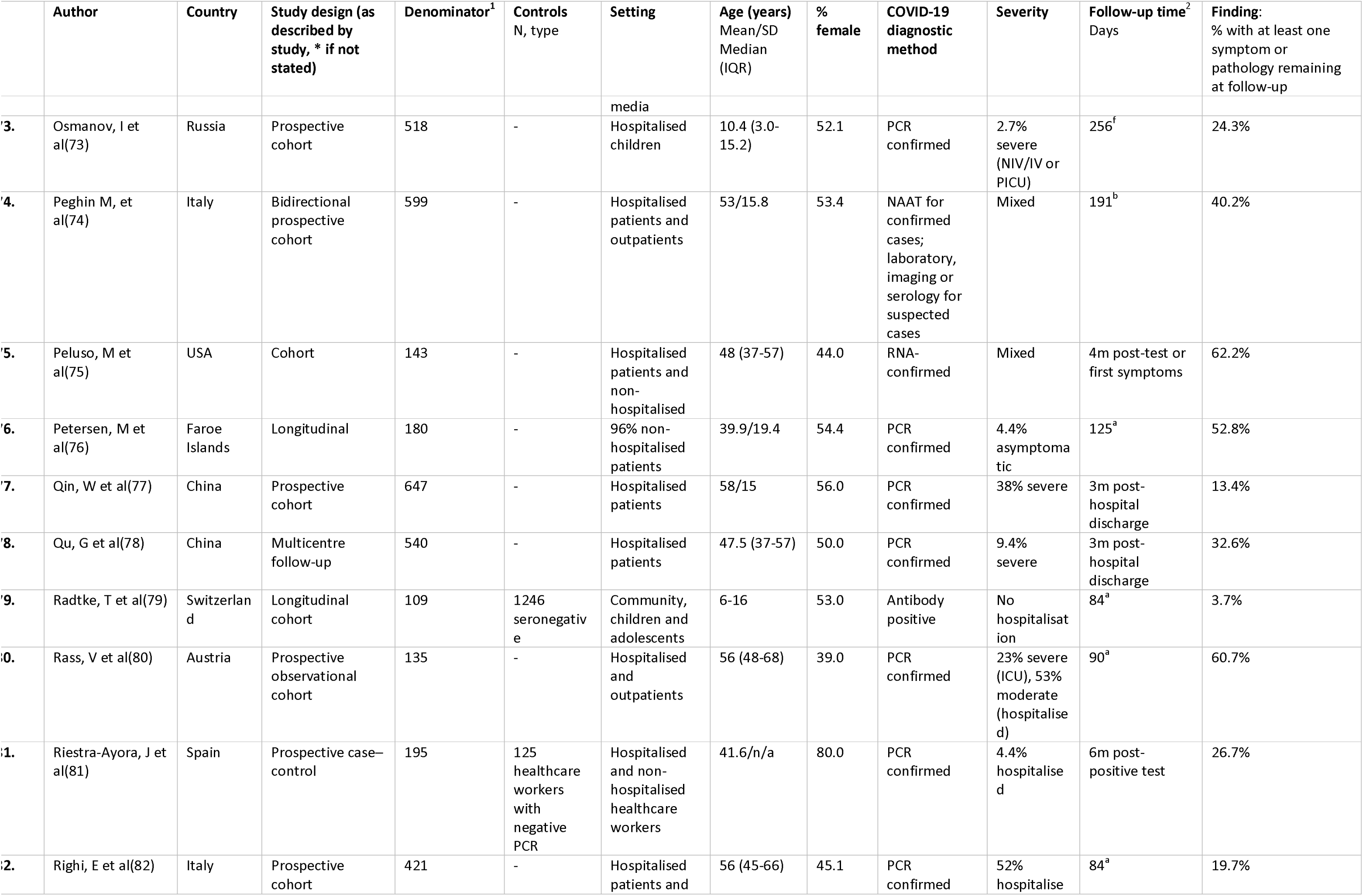
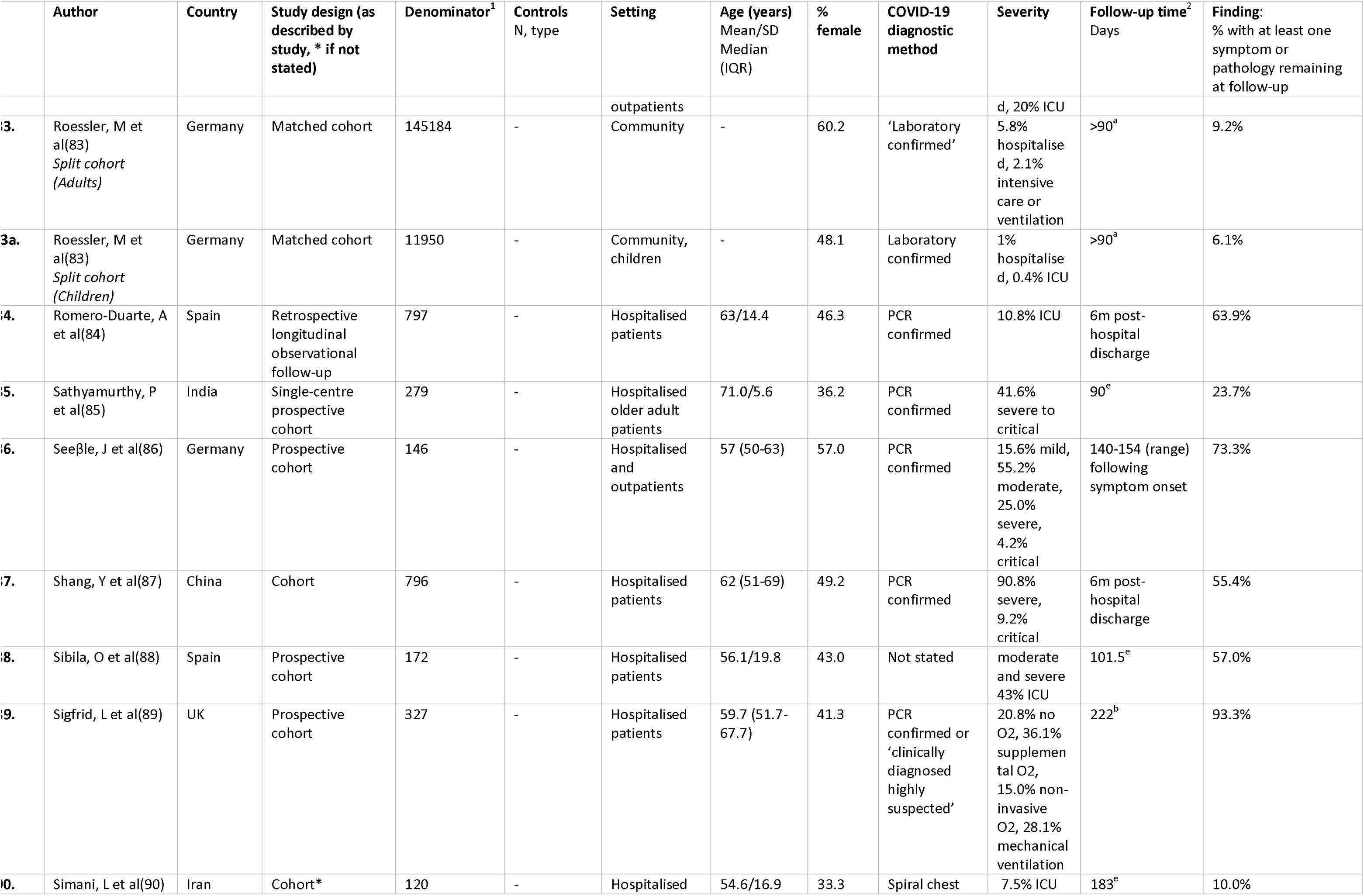
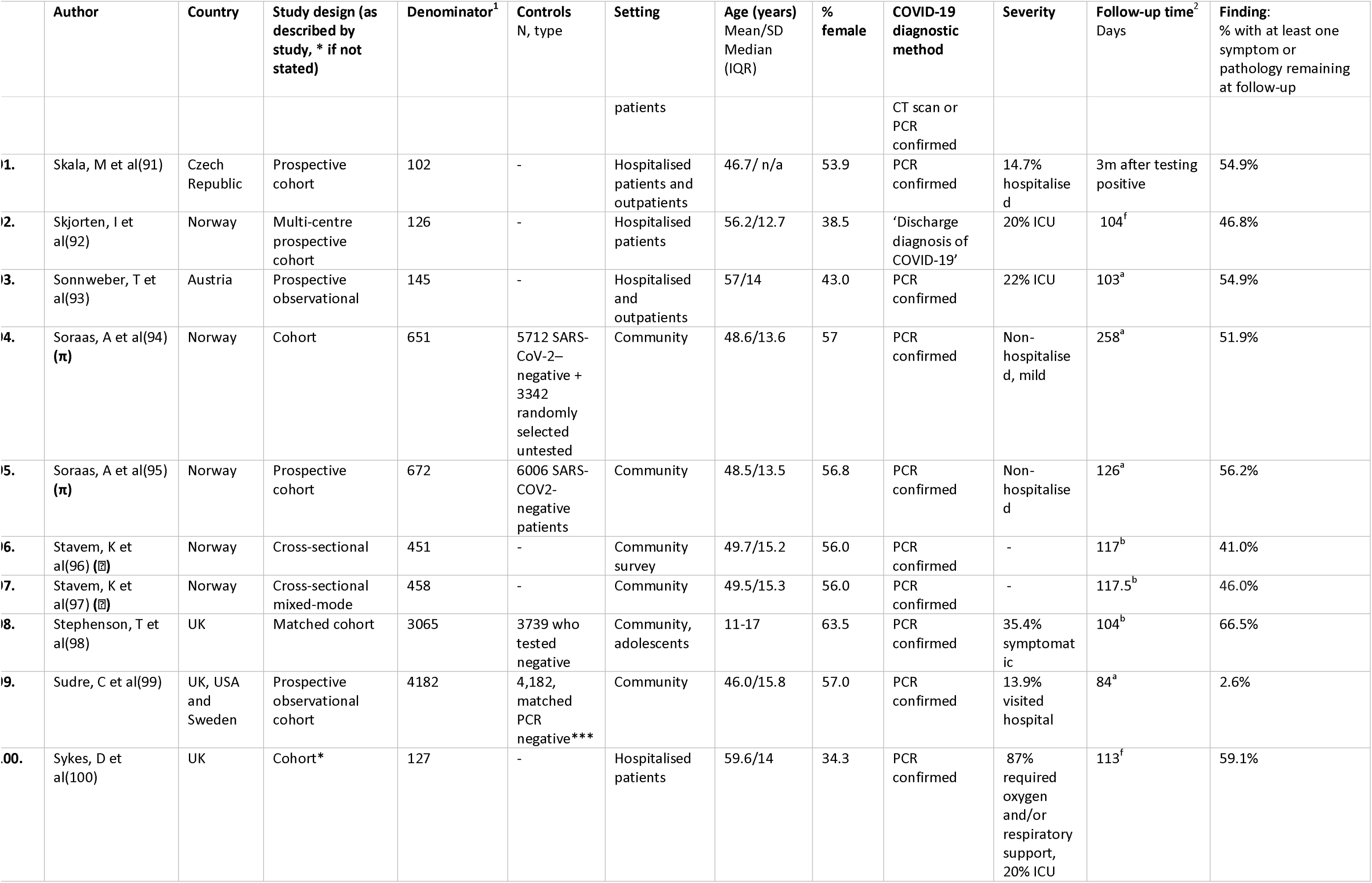
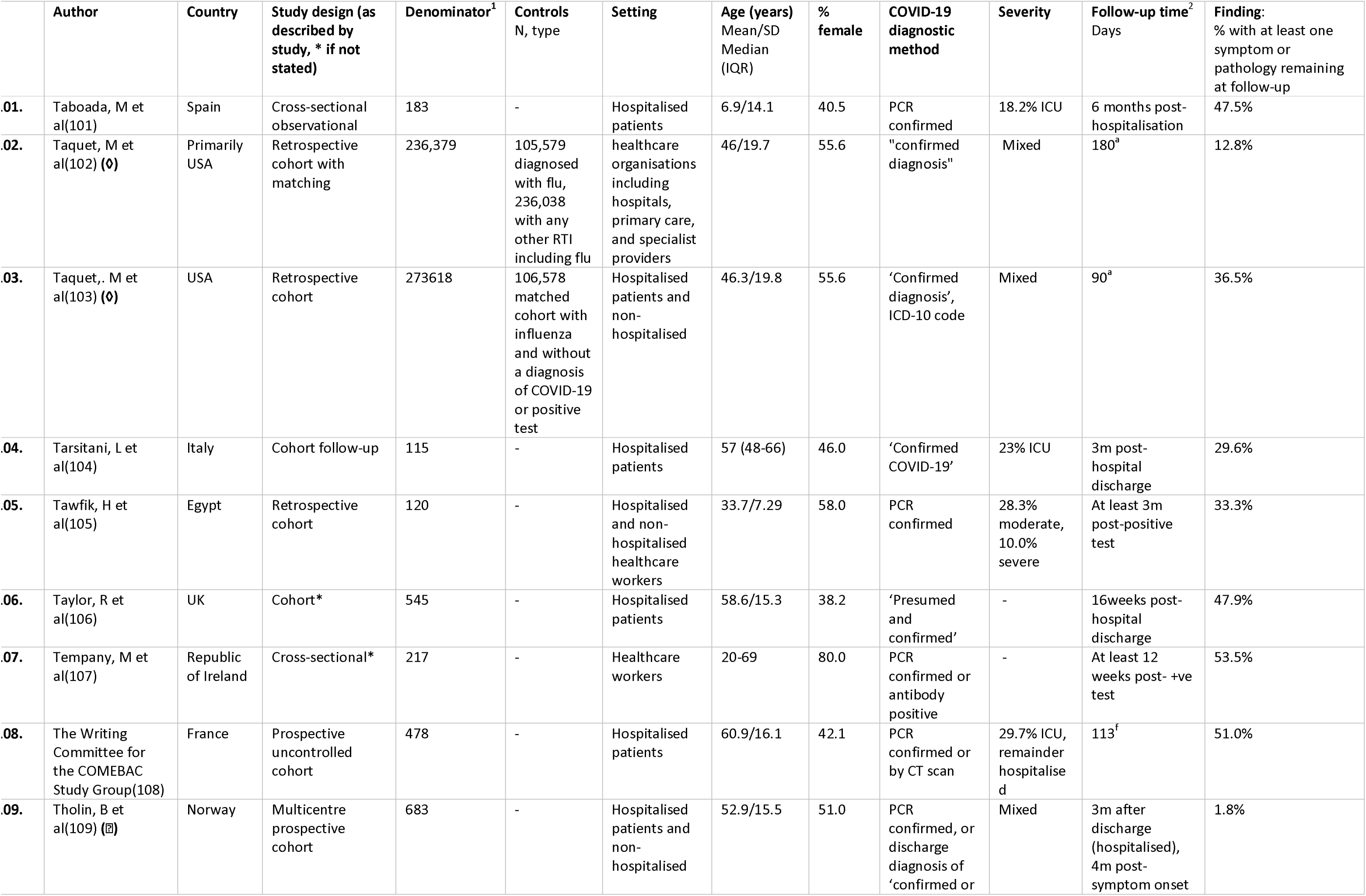
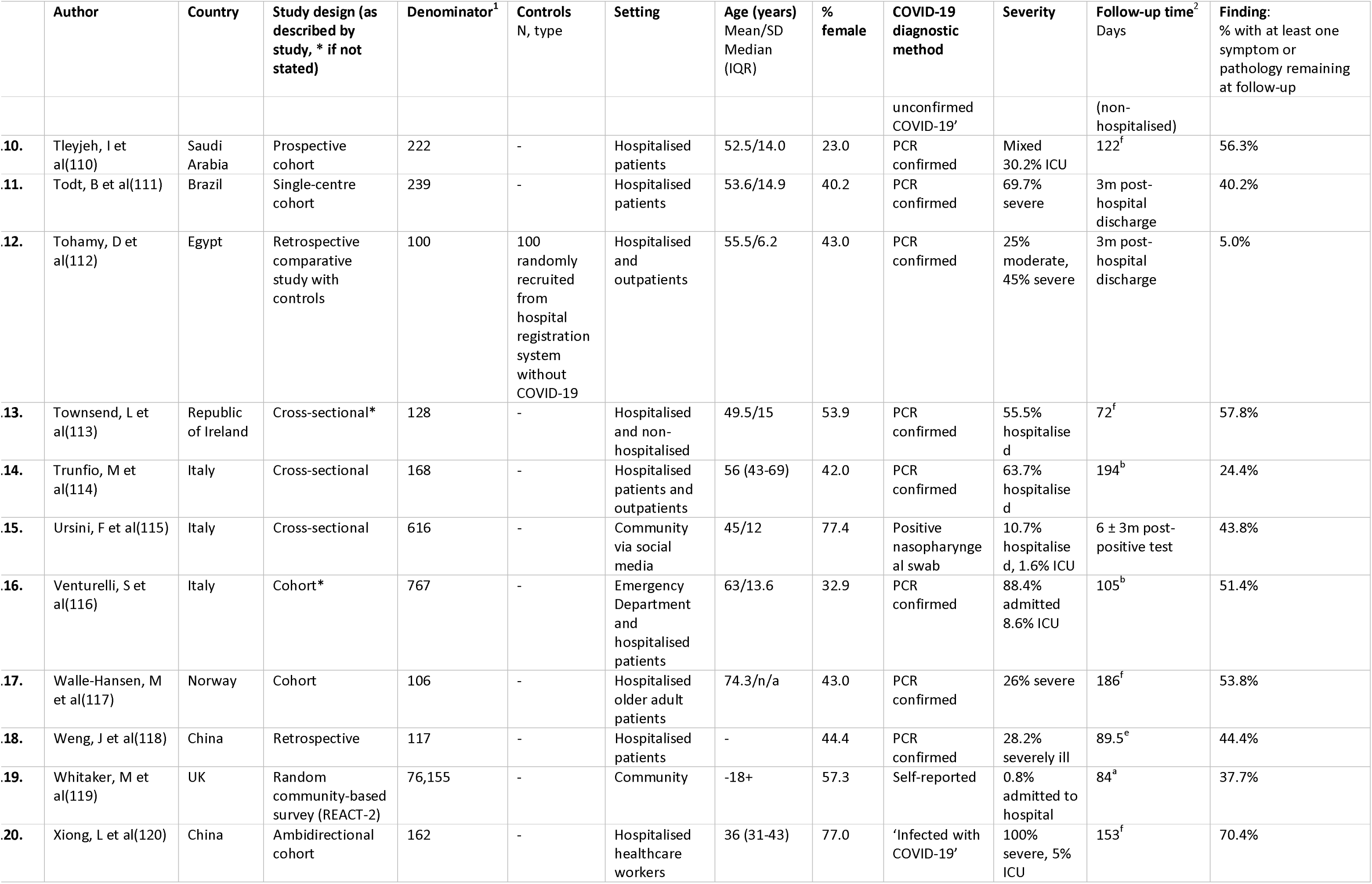
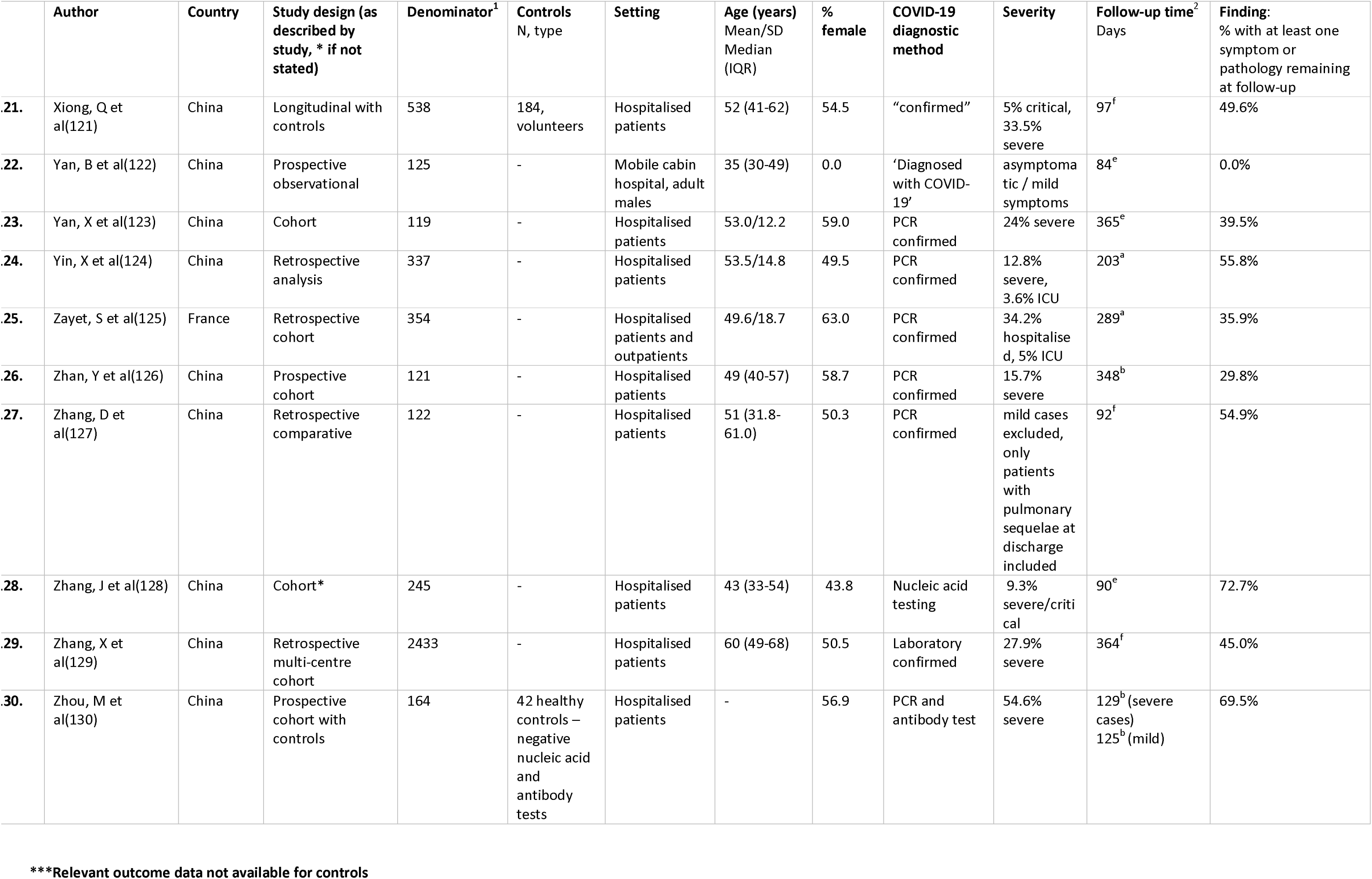
– Study characteristics and findings of first follow-up

However, hospitalisation did not always correspond with disease severity, probably due to local diagnostic, treatment, and containment policies. Most studies used PCR testing to identify COVID-19 cases at baseline. However most did not perform COVID-19 diagnostic tests at follow-up and therefore did not consider the impact of reinfection on their results. Out of the included studies, 21 were community-based studies, 17 outpatient settings, 3 social media and 8 healthcare worker-based studies.

### Prevalence estimates

The prevalence of Long Covid for studies with more than 12 weeks from infection ranged between 0% to 93% (pooled estimate (PE) 42.1%, 95% prediction interval (PI): 6.8% to 87.9%) (Figure 2). 73 included studies had a follow up of 12 weeks to 5 months, 49 had a follow-up of 6-11 months and 12 had a follow-up of 12 months or more. The range of prevalence in studies with follow-up of 12 months or more was 17% to 81% . Recognising most are not within-study comparisons, longer follow-up times showed higher pooled estimates (Supplementary Figure 1). For all complete and subgroup analyses except one, I^2^ was >75%. All subgroup analysis results including pooled estimates and prediction intervals can be found in Supplementary Table 4.

**Figure 2:**
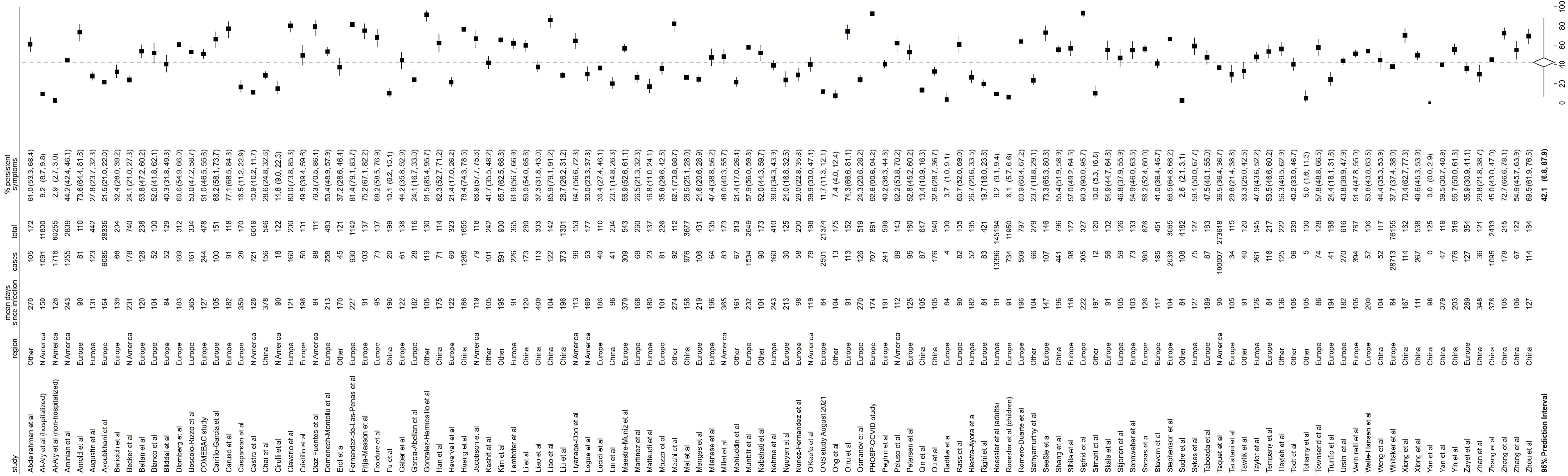
Forest plot of prevalence of Long Covid in the included studies, with 95% prediction intervals

The prevalence range in analyses where less than 10% of the participants were hospitalised was 0% to 67% (n=24). In studies where all participants were hospitalised for acute COVID-19 (n=65), the prevalence range was 5% to 93% (Supplementary Figure 2). 31 analyses had 10% or more of their sample admitted to intensive care unit (ICU) during their acute COVID-19 illness (Supplementary Figure 3). Studies including more hospitalised participants or more patients in ICU tended to report higher prevalence estimates (Supplementary Table 4). Likewise using the WHO CPS, studies including those with ambulatory mild disease (n=38) generally reported lower prevalence estimates than those with hospitalised severe disease who needed oxygen by NIV or high flow (n=27) (Supplementary Figure 4).

The prevalence of not returning to full health/fitness after at least 12 weeks from infection ranged between 8% to 70% (pooled estimate 34.5%, 95% PI: 4.3% to 85.9%, n=10) (Supplementary Figure 5). The prevalence of lower quality of life was 31% (n=2) (Supplementary Figure 6). With regards to individual symptoms, common symptoms reported included fatigue followed by breathing problems, sleep problems, tingling or itching, and joint/muscle aches and pains. With regards to pathology, lung pathology was the most common followed by heart or neurological pathology (Supplementary Figures 7-40).

There were very few studies with a low risk of bias (Supplementary Table 2). Few studies used a sample that was representative of all COVID-19 cases in the population. Approximately half of the studies indicated that symptoms had not been present prior to infection, while the rest did not report ascertaining this. When stratifying by risk of bias, generally lower prevalence estimates were seen in studies with COVID-19 diagnoses confirmed for all participants, studies scored as having a representative sample, studies with an internal or external non-COVID-19 comparator, studies that assessed all participants in the same way, and studies based on community participants (Supplementary Figure 41-42).

Comorbidities, ethnicity and other demographic data were not reported in all studies. Higher prevalence of Long Covid was observed in studies where populations had higher proportions of older people, males, people of non-white ethnicity, diabetes, hypertension, cardiovascular disease, and related comorbidities (Supplementary Figure 43).

**Figure 3:**
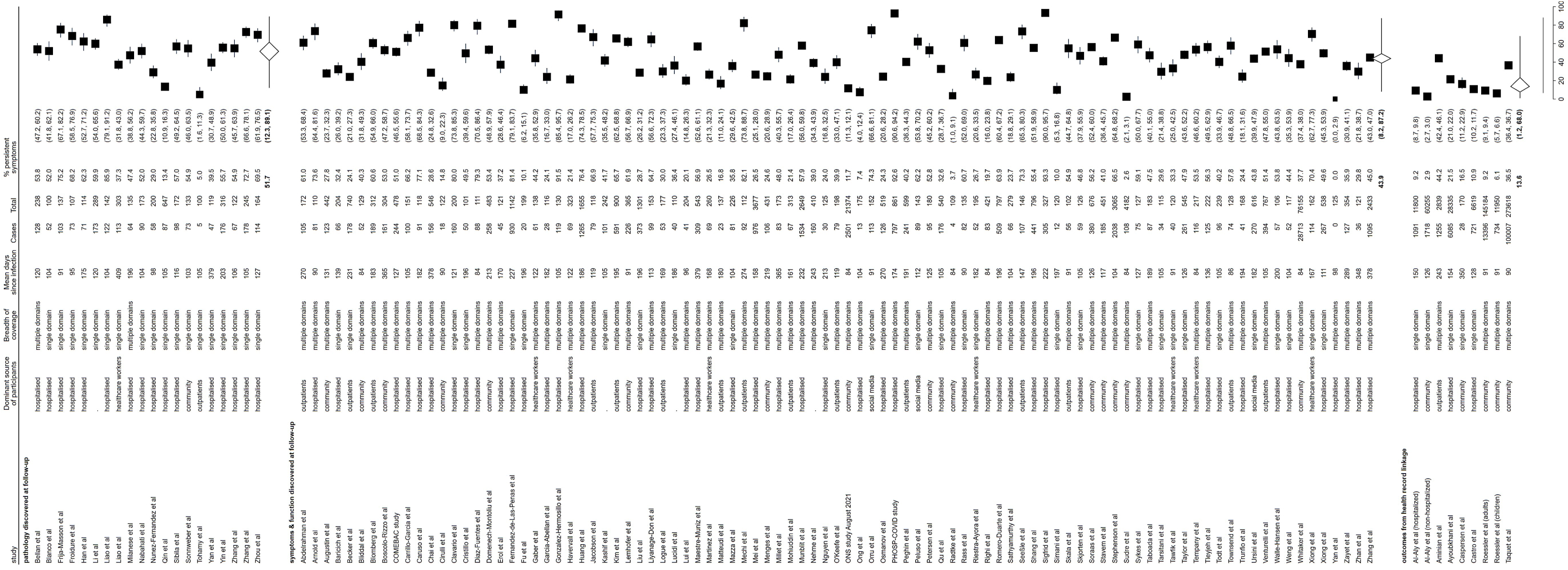
Forest plot of prevalence of Long Covid in the included studies by method of outcome assessment, with 95% prediction intervals

Prevalence also varied by study design. In cohort studies the range was 0% to 93% (pooled estimate 41.3%, 95% PI: 6.0% to 88.6%) and in cross sectional studies, the prevalence of Long Covid ranged between 10% and 82% (pooled estimate 45.9%, 95% PI: 11.2% to 85.1%) (Supplementary Figure 50). Prevalence estimates derived from assessing Long Covid as self-reported symptoms and function (n=93) on the whole tended to report higher prevalence than those that used clinical coding in healthcare records (n=7). However, studies that had dedicated pathology follow-up of COVID-19 patients (for example pulmonary function tests or scans with pathology discovered at follow-up) tended to report the highest prevalence (n=20) (Figure 3). Studies that defined Long Covid as at least one of multiple symptom or pathology domains tended to report a slightly higher prevalence than those that assessed a single symptom/pathology domain (Supplementary Figure 44).

**Figure 4:**
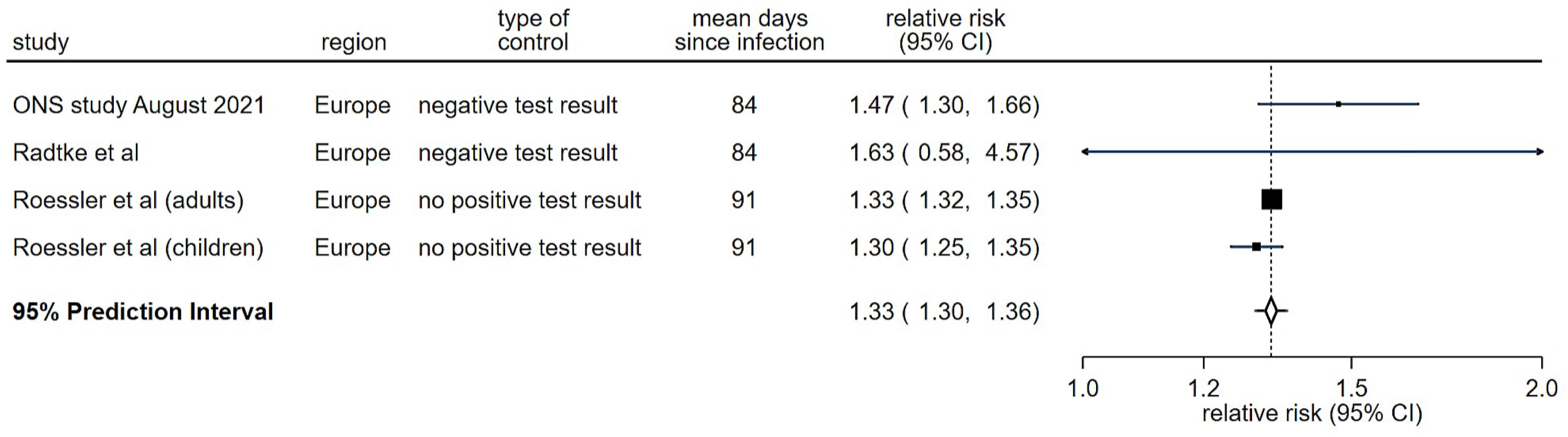
Forest plot of risk of Long Covid in included studies with community-based samples and controls assessed as having low risk of bias, with 95% prediction intervals

### Comparison to controls

Twenty-four of the 130 publications included comparison to at least one group of controls (Supplementary Figure 45). The majority of studies used test-negative controls (antigen and antibody, with some matching), but others used untested controls. In community-based studies with controls, the relative risk ranged between 1.0 to 51.4 (pooled relative risk 2.7, 95% PI: 0.2 to 39.4) and the absolute risk difference ranged between -1% to 35% (pooled risk difference 10.1%, 95% PI: - 12.7% to 32.8%) (Supplementary Figures 46-47). In community-based samples with controls and assessed as having a low risk of bias (n=4), the pooled relative risk of experiencing symptoms/ill health after COVID-19 was 1.33 compared to controls (95% PI: 1.30. 1.36, I^2^ =28.1%) (Figure 4) and the absolute risk difference between cases and controls ranged between 1% to 9% (Supplementary Figure 48).

There was no evidence of small-study effects such as publication bias (Supplementary Figure 49).

## Discussion

This systematic review which included 120 studies assessing Long Covid symptoms, functional status, or pathology published up to November 2021 demonstrates substantial between-study heterogeneity and wide variation in prevalence estimates. This is due to differences in study designs (cross sectional or longitudinal), sources of study samples (community, outpatient clinic, occupational, hospitalised) and number of assessed symptoms and method of assessment (self-reported individual or collective symptoms, healthcare records, clinical investigations at follow up). The only pooled estimate with low between-study heterogeneity was a 33% (95% PI: 30% to 36%) excess risk of experiencing prolonged symptoms in COVID-19 cases compared to controls in community-based studies with low risk of bias. Although studies that included controls showed, on the whole, lower net prevalence of Long Covid than studies that did not, the evidence from most of these studies is that COVID-19 is associated with a substantially higher risk of being ill 12 weeks after infection than those not infected.

The UK’s Office for National Statistics (ONS) produces population-level Long Covid prevalence estimates where the denominator is the whole population in the specific reported population group, for example, by age, sex, or occupation(144). These fall out of our inclusion criteria. The ONS also produced prevalence estimates based on following up those with confirmed SARSCoV2 infection and we used the most recent estimate within the review’s search period(83). This study used multiple approaches including assessing individual symptoms compared to controls and asking participants if they believe they have Long Covid. The latter approach, in the absence of a standardised method of assessment, may realistically be the best way to assess the presence of Long Covid as most people will take the combination of their symptoms, duration, fluctuation, effect on functional ability and change from pre-COVID19 health to shape their responses.

The lack of consensus on the precise definition of Long Covid plays an important part in the wide differences in prevalence assessments, however we found that specifically the way the question is asked and the source of retrieved clinical information at follow-up are likely to play a crucial role. The ONS study is an example of how different methods of assessment at time of follow-up can produce substantially different Long Covid estimates(83). This was illustrated by our analysis where studies that asked about multiple symptoms/domains tended to report higher prevalence estimates than single domain studies. Our analysis indicated higher prevalence estimates with longer follow-up time, though we recognise these were mostly not within-study comparisons. However, in four of ten longitudinal studies, prevalence was higher at the time of the second follow-up. These results could be explained by several factors e.g. by the episodic nature of Long Covid, whereby in the early stages people may feel they have got over their illness, but with passing time and phases of relapse and remittance, people may be more cautious about reporting they have recovered. People may also be developing new symptoms over time, or perhaps there is more study drop-out by people who feel they have recovered. Overall however, the results indicate that, over time, prevalence does not substantially reduce.

Studies that used questionnaires/surveys to ask participants about their symptoms, health status or quality of life tend to report higher prevalence estimates than those that recorded symptoms from healthcare records’ clinical coding. This is manifested in the prevalence from Al-Aly et al(15) studies being on the lower side in our analysis as we only included those with symptoms rather than recorded post-COVID-19 pathology, and such symptoms are expected to be severe enough to prompt seeking medical help and being recorded in medical notes. Studies that had dedicated pathology follow-up and discovery of COVID-19 patients tended to report the highest prevalence. This is possibly because, in addition to pathology that leads to recognisable signs and symptoms, specific medical investigations as part of the research protocol can pick up latent pathology that may not be accompanied by clinical manifestations.

Studies such as Al-Aly et al investigating medical diagnoses in the period following COVID-19, report cardiovascular, neurological, and other system-specific clinical sequelae providing a substantial excess burden in those who survived the acute phase of COVID-19(13). However, there is no agreement yet whether these outcomes are classed as Long Covid. They are generally not recorded by symptom studies and the WHO does not yet specifically include such outcomes within its clinical case definition of Post-COVID-19 Condition (also known as Long Covid) (1). A specific pathology diagnosed after COVID-19 could have been triggered by the infection, but identification as such will depend on the extent of clinical investigations identifying and labelling specific pathology as opposed to differences in the disease manifestation themselves.

Other sources of heterogeneity between studies include study design as some were cross-sectional with assessment taking place at one point in time, whereas others were longitudinal where assessment of COVID-19 status was conducted prior to the development of Long Covid. This assessment itself varied in terms of using PCR or antigen testing or self-reporting of history of acute infection.

Ideally, excess absolute risk in comparison to controls is a good measure to estimate the burden of Long Covid. This is likely dependent on the approach to control selection, whether based on self-report of absence of infection history or lab results that are not accurate enough to ascertain the state of previous infection (antigen or antibody), and timing of assessment given the predominant episodic nature of Long Covid.

Few studies had a low risk of bias, which suggests there is a gap in the evidence base for strong studies of Long Covid prevalence. In terms of causal inference, many studies were liable to potential collider bias, which presented as selection bias caused by restricting analyses to people who were hospitalised, self-selected for PCR or lateral flow tests based on symptoms, or simply volunteered their study participation(145). Similarly, our exploration of potential sources of heterogeneity may be prone to table 2 fallacy in the original studies, where these subgroups do not derive from the focal research question, so should be interpreted descriptively rather than causally(146).

The strengths of our review include comprehensive electronic searching for relevant studies and comprehensive assessment of risk of bias, data extraction and checking with each of these processes being done independently by two authors. We also adapted the Newcastle-Ottawa scale (Supplementary Table 3) for this prevalence systematic review which can be used by other researchers for risk assessment and/or to build high quality study designs. The quality assessment criteria and process were discussed within the study team which includes two authors with lived experience of Long Covid.

Our review was limited by the substantial between study heterogeneity. We used the most common reported symptom estimate for studies and did not combine multiple individual symptoms into one overall estimate of prevalence of Long Covid. The symptom with the highest prevalence differed from study to study, so may not be entirely comparable. We did not include more recent studies that assessed the prevalence of Long Covid following infection with different variants of SARSCoV2 and/or in double or triple vaccinated populations. Recent estimates point to a prevalence of 4-5% of reporting Long Covid at 12 to 16 weeks after first confirmed SARSCoV2 infection depending on variant, with no evidence of difference between variants among those who are triple vaccinated when infected(147). In those double vaccinated, the prevalence of persistent symptoms was around 10% compared to 15% of unvaccinated controls(148).

We extracted estimates of “new-onset” Long Covid/symptoms where possible. Where the proportion is of a symptom like fatigue for example, we picked the one quoted as new-onset fatigue if available, or we downgraded quality because it was not possible to ascertain that the symptom is ‘new’ following infection. Because Long Covid is a novel condition, prevalence of the condition is considered equivalent to cumulative incidence. When comparing with controls, we estimated cumulative incidence from reported absolute risk, when appropriate. When reporting risk ratio, we included incidence rate ratio and hazard ratios, but did not consider the odds ratio an adequate approximation because of the high potential prevalence in some populations.

We know that significant numbers of people experience ill health following SARSCoV2 infection. Long Covid impacts on society, particularly in places with continuing waves of infection. Through reviewing how different research approaches attempted to quantify the population burden of Long Covid, our findings provide insight into how to get more accurate estimates of prevalence and severity. With quantification of prevalence, we can understand the investment needed for prevention, diagnosis, and treatment as well as the policy decisions needed to resource healthcare and social care services and to mitigate the wider social and economic impact of Long Covid.

## Data Availability

All data used in this review is available in the published included studies.  Data extractions and analytic code is available from the authors on reasonable request.

## Contributors

NAA, DCG, RT, AA, VL, MW conceptualised and designed the study. MW drafted the protocol and search strategy with input from all co-authors. VL conducted the search. All authors contributed to screening the articles. MW, DCG, NZ, RT, CC extracted and quality-assessed the data. NAA, MW, DCG, NZ, CC contributed to the process of checking and verifying the extracted data. DCG planned and conducted the statistical analyses and produced the forest plots. MW, DCG, NZ, NAA interpreted the data and drafted the manuscript. All authors reviewed the final manuscript. All authors had full access to all the data in the study and had final responsibility for the decision to submit for publication.

## Potential conflicts of interest

The authors declare no competing interests.

## Acknowledgements

We thank Hannah Davies for her input in conceptualising this review. MW was supported by an NIHR Pre-doctoral Local Authority Fellowship (Ref: 302098). RT and VL are supported by the Research, Evidence and Development Initiative (READ-It: project number 300342-104) which is funded by UK aid from the UK government. NZ is supported by NIHR Applied Research Collaboration Wessex. DCG is a co-investigator on the NIHR-funded LOCOMOTION study. NAA has lived experience of Long Covid, is a co-investigator on the NIHR-funded STIMULATE-ICP and HI-COVE studies, has contributed in an advisory capacity to WHO and the EU Commission’s Expert Panel on effective ways of investing in health meetings in relation to post-COVID-19 condition, and has acted as a collaborator on some of the UK’s Office for National Statistics outputs on the prevalence of Long Covid. AA has lived experience of Long Covid, is a co-founder of the Patient-Led Research Collaborative and has contributed in an advisory capacity to NIH, CDC and WHO. The views and opinions expressed in this review are those of the authors and do not necessarily reflect those of the NIHR, the Department of Health and Social Care or the UK government’s official policies.

For the purpose of open access, the author has applied a CC BY public copyright licence to any Author Accepted Manuscript version arising from this submission.

## Supplementary material

Supplementary appendix

## Notes

### Competing Interest Statement

The authors have declared no competing interest.

### Clinical Protocols

https://www.crd.york.ac.uk/prospero/display_record.php?RecordID=218351

### Funding Statement

There was no specific funding source for this study.

### Summary of Updates

Inclusion of pooled estimates with prediction intervals

