## Supplementary material for "Systematic review of the prevalence of Long Covid"

### 1. Full search terms for one database (Medline)

The search strategies applied to Medline is shown; it was adapted for other electronic databases as needed.

Ovid MEDLINE(R) and In-Process, In-Data-Review & Other Non-Indexed Citations <1946 to October 29, 2021>

| | |
|---|---|
| 1 | ((COVID* or SARS-CoV-2 or SARSCoV2).tw. or Coronavirus/ or Coronavirus Infections/) adj2 (long-term or persistent symptoms or protracted or post-discharge or symptom* duration or persistent fatigue).tw. |
| 2 | ((COVID* or SARS-CoV-2 or SARSCoV2).tw. or Coronavirus/ or Coronavirus Infections/) adj3 (follow-up or longitudinal or consequenc* or sequela* or brain fog or months or after or survivor* or convalescent or convalescence).ti. |
| 3 | ((COVID* or SARS-CoV-2 or SARSCoV2).tw. or Coronavirus/ or Coronavirus Infections/) adj2 (long or long-haul*) |
| 4 | post-Covid*.tw. |
| 5 | Post-acute COVID 19 syndrome.mp. |
| 6 | Post-acute sequelae of COVID 19.mp. |
| 7 | PASC.mp. |
| 8 | Post-acute coronavirus syndrome.mp. |
| 9 | Chronic covid syndrome.mp. |
| 10 | Long-term covid syndrome.mp. |
| 11 | Post covid-19 condition.mp. |
| 12 | 1-11/or |

### 2. Supplementary Tables

Supplementary Table 1: List of studies excluded at full text screening stage with brief reason

| Title | Authors | Published Year | Journal, Volume, Issue, Pages, DOI |
|---|---|---|---|
| **Fewer than 100 participants (n=33)** | | | |
| 1. Do post-COVID-19 symptoms exist? A longitudinal study of COVID-19 sequelae in Wenzhou, China | Zhou, M.; Cai, J.; Sun, W.; Wu, J.; Wang, Y.; Gamber, M.; Fan, L.; He, G. | 2021 | Annales Medico Psychologiques. http://dx.doi.org/10.1016/j.amp.2021.03.003 |
| 2. Post-acute sequelae of COVID-19: Evidence of mood & cognitive impairment | Lamontagne, S. J.; Winters, M. F.; Pizzagalli, D. A.; Olmstead, M. C. | 2021 | Brain, Behavior, & Immunity Health 100347 https://dx.doi.org/10.1016/j.bbih.2021.100347 |
| 3. Investigation of Long COVID Prevalence and Its Relationship to Epstein-Barr Virus Reactivation | Gold, Jeffrey E.; Okyay, Ramazan A.; Licht, Warren E.; Hurley, David J. | 2021 | Pathogens (Basel, Switzerland) 10, 6 https://dx.doi.org/10.3390/pathogens10060763 |
| 4. Chronic fatigue syndrome: an emerging sequela in COVID-19 survivors? | Mantovani, Elisa; Mariotto, Sara; Gabbiani, Daniele; Dorelli, Gianluigi; Bozzetti, Silvia; Federico, Angela; Zanzoni, Serena; Girelli, Domenico; Crisafulli, Ernesto; Ferrari, Sergio; Tamburin, Stefano | 2021 | Journal of neurovirology 27, 4 631-637 https://dx.doi.org/10.1007/s13365-021-01002-x |
| 5. Long-term follow-up of patients with venous thromboembolism and COVID-19: Analysis of risk factors for death and major bleeding | Demelo-Rodriguez, Pablo; Ordieres-Ortega, Lucia; Ji, Zichen; Del Toro-Cervera, Jorge; de Miguel-Diez, Javier; Alvarez-Sala-Walther, Luis A.; Galeano-Valle, Francisco | 2021 | European journal of haematology 106, 5 716-723 https://dx.doi.org/10.1111/ejh.13603 |
| 6. Cerebral Micro-Structural Changes in COVID-19 Patients - An MRI-based 3-month Follow-up Study: A brief title: Cerebral Changes in COVID-19 | Lu, Y.; Li, X.; Geng, D.; Mei, N.; Wu, P. Y.; Huang, C. C.; Jia, T.; Zhao, Y.; Wang, D.; Xiao, A.; Yin, B. | 2020 | 25 100484 http://dx.doi.org/10.1016/j.eclinm.2020.100484 |
| 7. Chest CT and Clinical Follow-up of Discharged Patients with COVID-19 in Wenzhou City, Zhejiang, China | Liu C, Ye L. Xia R. Zheng X. Yuan C. Wang Z. Lin R. Shi D. Gao Y. Yao J. Sun Q. Wang X. Jin M. | 2020 | Annals of the american thoracic society 10.1513/AnnalsATS.202004-324OC |
| 8. Chest CT in COVID-19 pneumonia: what are the findings in mid-term follow-up? | Tabatabaei Smh, Rajebi H. Moghaddas F. Ghasemiadl M. Talari H. | 2020 | Emergency radiology 10.1007/s10140-020-01869-z |
| 9. Do COVID-19 Antibodies Provide Long-Term Protection? | Ansari, Sheeba; Memon, Mubeen; Kumar, Ratan; Memon, Sidra; Memon, Muhammad Khizar | 2021 | Cureus 13, 1 e12441 https://dx.doi.org/10.7759/cureus.12441 |
| 10. Follow-up study of the pulmonary function and related physiological characteristics of COVID-19 survivors three months after recovery | Zhao, Yu-Miao; Shang, Yao-Min; Song, Wen-Bin; Li, Qing-Quan; Xie, Hua; Xu, Qin-Fu; Jia, Jun-Li; Li, Li-Ming; Mao, Hong-Li; Zhou, Xiu-Man; Luo, Hong; Gao, Yan-Feng; Xu, Ai-Guo | 2020 | EClinicalMedicine 25 100463 https://dx.doi.org/10.1016/j.eclinm.2020.100463 |
| 11. Impact of coronavirus disease 2019 on pulmonary function in early convalescence phase | Huang, Yiying; Tan, Cuiyan; Wu, Jian; Chen, Meizhu; Wang, Zhenguo; Luo, Liyun; Zhou, Xiaorong; Liu, Xinran; Huang, Xiaoling; Yuan, Shican; Chen, Chaolin; Gao, Fen; Huang, Jin; Shan, Hong; Liu, Jing | 2020 | Respiratory research 21, 1 163 https://dx.doi.org/10.1186/s12931-020-01429-6 |
| 12. Medium-term effects of SARS-CoV-2 infection on multiple vital organs, exercise capacity, cognition, quality of life and mental health, post-hospital discharge | Raman, Betty; Cassar, Mark Philip; Tunnicliffe, Elizabeth M.; Filippini, Nicola; Griffanti, Ludovica; Alfaro-Almagro, Fidel; Okell, Thomas; Sheerin, Fintan; Xie, Cheng; Mahmod, Masliza; Mozes, Ferenc E.; Lewandowski, Adam J.; Ohuma, Eric O.; Holdsworth, David; Lamlum, Hanan; Woodman, Myles J.; Krasopoulos, Catherine; Mills, Rebecca; McConnell, Flora A. Kennedy; Wang, Chaoyue; Arthofer, Christoph; Lange, Frederik J.; Andersson, Jesper; Jenkinson, Mark; Antoniades, Charalambos; Channon, Keith M.; Shanmuganathan, Mayooran; Ferreira, Vanessa M.; Piechnik, Stefan K.; Klenerman, Paul; Brightling, Christopher; Talbot, Nick P.; Petousi, Nayia; Rahman, Najib M.; Ho, Ling-Pei; Saunders, Kate; Geddes, John R.; Harrison, | 2021 | EClinicalMedicine 31 100683 https://dx.doi.org/10.1016/j.eclinm.2020.100683 |

| | | | | |
|---|---|---|---|---|
| | | Paul J.; Pattinson, Kyle; Rowland, Matthew J.; Angus, Brian J.; Gleeson, Fergus; Pavlides, Michael; Koychev, Ivan; Miller, Karla L.; Mackay, Clare; Jezzard, Peter; Smith, Stephen M.; Neubauer, Stefan | | |
| 13. | Modified rehabilitation exercises for mild cases of COVID-19 | Zha L, Xu X. Wang D. Qiao G. Zhuang W. Huang S. | 2020 | Annals of palliative medicine 10.21037/apm-20-753 |
| 14. | Persistence of symptoms in patients after coronavirus disease (COVID-19) in a third level hospital of Puebla, Mexico. [Spanish] | Garcia, J. C. H.; Montellano, E. I. A.; Gonzalez, L. I. J.; Andrade, R. C. | 2020 | Medicina Interna de Mexico 36, 6 789-793 http://dx.doi.org/10.24245/mim.v36i6.4581 |
| 15. | Persisting olfactory dysfunction in patients after recovering from COVID-19 | Otte, M. S.; Klussmann, J. P.; Luers, J. C. | 2020 | The Journal of infection 81, 3 e58 https://dx.doi.org/10.1016/j.jinf.2020.06.054 |
| 16. | Post-acute COVID-19 syndrome negatively impacts health and wellbeing despite less severe acute infection (preprint) | Tabacof L, Tosto-Mancuso J. Wood J. Cortes M. Kontorovich A. McCarthy D. Rizk D. Nasr L. Breyman E. Mohammadi N. Kellner C. Putrino D. | 2020 | Medrxiv 2020.11.04.20226126 10.1101/2020.11.04.20226126 |
| 17. | Post-discharge persistent symptoms and health-related quality of life after hospitalization for COVID-19 | Garrigues E, Janvier P. Kherabi Y. Bot A. L. Hamon A. Gouze H. Doucet L. Berkani S. Oliosi E. Mallard E. Corre F. Zarrouk V. Moyer J. D. Galy A. Honsel V. Fantin B. Nguyen Y. | 2020 | Journal of infection 10.1016/j.jinf.2020.08.029 |
| 18. | Psychological Distress After Covid-19 Recovery: Reciprocal Effects With Temperament and Emotional Dysregulation. An Exploratory Study of Patients Over 60 Years of Age Assessed in a Post-acute Care Service | Janiri, Delfina; Kotzalidis, Georgios D.; Giuseppin, Giulia; Molinaro, Marzia; Modica, Marco; Montanari, Silvia; Terenzi, Beatrice; Carfi, Angelo; Landi, Francesco; Sani, Gabriele; Gemelli Against, Covid-Post-acute Care Study Group | 2020 | Frontiers in psychiatry 11 590135 https://dx.doi.org/10.3389/fpsyt.2020.590135 |
| 19. | Short-Term Follow-Up of Self-Isolated COVID-19 Patients with Smell and Taste Dysfunction in Greece: two Phenotypes of Recovery | Konstantinidis I, Delides A. Tsakiropoulou E. Maragoudakis P. Sapounas S. Tsiodras S. | 2020 | ORL; journal for oto-rhino-laryngology and its related specialties 10.1159/000511436 |
| 20. | Sofosbuvir and daclatasvir for the treatment of COVID-19 outpatients: a double-blind, randomized controlled trial | Roozbeh, Fatemeh; Saeedi, Majid; Alizadeh-Navaei, Reza; Hedayatizadeh-Omran, Akbar; Merat, Shahin; Wentzel, Hannah; Levi, Jacob; Hill, Andrew; Shamshirian, Amir | 2021 | The Journal of antimicrobial chemotherapy 76, 3 753-757 https://dx.doi.org/10.1093/jac/dkaa501 |
| 21. | Three-month Follow-up Study of Survivors of Coronavirus Disease 2019 after Discharge | Liang, Limei; Yang, Bohan; Jiang, Nanchuan; Fu, Wei; He, Xinliang; Zhou, Yaya; Ma, Wan Li; Wang, Xiaorong | 2020 | Journal of Korean medical science 35, 47 e418 https://dx.doi.org/10.3346/jkms.2020.35.e418 |
| 22. | Preliminary Evidence on Long COVID in children (preprint) | buonsenso d, Munblit D. De Rose C. Sinatti D. Ricchiuto A. Carfi A. Valentini P. | 2021 | Medrxiv 2021.01.23.21250375 10.1101/2021.01.23.21250375 |
| 23. | Preliminary evidence on long COVID in children | Buonsenso, D.; Munblit, D.; De Rose, C.; Sinatti, D.; Ricchiuto, A.; Carfi, A.; Valentini, P. | 2021 | Acta Paediatrica 110, 7 2208-2211 https://dx.doi.org/10.1111/apa.15870 |
| 24. | Pulmonary Sequelae at 4 Months After COVID-19 Infection: A Single-Centre Experience of a COVID Follow-Up Service | Robey, R. C.; Kemp, K.; Hayton, P.; Mudawi, D.; Wang, R.; Greaves, M.; Yioe, V.; Rivera-Ortega, P.; Avram, C.; Chaudhuri, N. | 2021 | Advances in Therapy 38, 8 4505-4519 https://dx.doi.org/10.1007/s12325-021-01833-4 |
| 25. | Pulmonary function and radiological features 4 months after COVID-19: first results from the national prospective observational Swiss COVID-19 lung study | Guler, Sabina A.; Ebner, Lukas; Aubry-Beigelman, Catherine; Bridevaux, Pierre-Olivier; Brutsche, Martin; Clarenbach, Christian; Garzoni, Christian; Geiser, Thomas K.; Lenoir, Alexandra; Mancinetti, Marco; Naccini, Bruno; Ott, Sebastian R.; Piquilloud, Lise; Prella, Maura; Que, Yok-Ai; Soccal, Paula M.; von Garnier, Christophe; Funke-Chambour, Manuela | 2021 | The European respiratory journal 57, 4 https://dx.doi.org/10.1183/13993003.03690-2020 |
| 26. | [Thrombus in the right atrium after COVID-19 pneumonia] | Alcoberro Torres, Lidia; Claver Garrido, Eduard; Moliner Borja, Pedro | 2020 | Trombo en la auricula derecha tras neumonia por COVID-19 73, 10 845 https://dx.doi.org/10.1016/j.recesp.2020.06.007 |
| 27. | Anormal pulmonary function and residual CT abnormalities in rehabilitating COVID-19 patients after discharge | You, Jingjing; Zhang, Lu; Ni-Jia-Ti, Ma-Yi-di-Li; Zhang, Jue; Hu, Fuyin; Chen, Luyan; Dong, Yuhao; Yang, Ke; Zhang, Bin; Zhang, Shuixing | 2020 | The Journal of infection 81, 2 e150-e152 https://dx.doi.org/10.1016/j.jinf.2020.06.003 |

| | | | |
|---|---|---|---|
| 28. Anormal pulmonary function and residual CT abnormalities in rehabilitating COVID-19 patients after discharge: a prospective cohort study | You J, Zhang L. Ni-Jia-Ti M. Y. Zhang J. Hu F. Chen L. Dong Y. Yang K. Zhang B. Zhang S. | 2020 | Journal of infection 10.1016/j.jinf.2020.06.003 |
| 29. COVID-19 in Children: Clinical Characteristics and Follow-Up Study | Ruan, P. S.; Xu, H. Q.; Wu, J. H.; Song, Q. F.; Qiu, H. Y. | 2020 | SN Comprehensive Clinical Medicine 2, 10 1713-1716 http://dx.doi.org/10.1007/s42399-020-00502-x |
| 30. Are there pulmonary sequelae in patients recovering from COVID-19? | Rogliani P, Calzetta L. Coppola A. Puxeddu E. Sergiacomi G. D'Amato D. Orlacchio A. | 2020 | Respiratory research 21, 1 286 10.1186/s12931-020-01550-6 |
| 31. Post SARS-CoV-2 Guillain-Barre syndrome | Arnaud S, Budowski C. Ng Wing Tin S. Degos B. | 2020 | Clinical neurophysiology 131, 7 1652-1654 10.1016/j.clinph.2020.05.003 |
| 32. Follow up of patients with severe coronavirus disease 2019 (COVID-19): pulmonary and extrapulmonary disease sequelae | Daher A, Balfanz P. Cornelissen C. Muller A. Bergs I. Marx N. Muller-Wieland D. Hartmann B. Dreher M. Muller T. | 2020 | Respiratory medicine 174 106197 10.1016/j.rmed.2020.106197 |
| 33. 'The long tail of Covid-19' - The detection of a prolonged inflammatory response after a SARS-CoV-2 infection in asymptomatic and mildly affected patients | Doykov, Ivan; Hallqvist, Jenny; Gilmour, Kimberly C.; Grandjean, Louis; Mills, Kevin; Heywood, Wendy E. | 2020 | F1000Research 9 1349 https://dx.doi.org/10.12688/f1000research.27287.1 |
| **No primary data (n=69)** | | | |
| 34. Reduced Diffusion Capacity in COVID-19 Survivors | Mendez, R.; Latorre, A.; Gonzalez-Jimenez, P.; Feced, L.; Bouzas, L.; Yepez, K.; Ferrando, A.; Zaldivar-Olmeda, E.; Reyes, S.; Menendez, R. | 2021 | Annals of the American Thoracic Society 18, 7 1253-1255 https://dx.doi.org/10.1513/AnnalsATS.202011-1452RL |
| 35. Urgent need of a management plan for survivors of COVID-19 | Celli, Bartolome; Fabbri, Leonardo M. | 2020 | The European respiratory journal 55, 4 https://dx.doi.org/10.1183/13993003.00764-2020 |
| 36. Direct SARS-CoV-2 infection of the heart potentiates the cardiovascular sequelae of COVID-19 | Bose, Rajendran J. C.; McCarthy, Jason R. | 2020 | Drug discovery today 25, 9 1559-1560 https://dx.doi.org/10.1016/j.drudis.2020.06.021 |
| 37. [COVID-19: A Pneumological Point of View - Long-Term Sequelae of COVID-19 - Implications For Follow-up In Respiratory Medicine] | Leo, Fabian; Wormanns, Dag; Grohe, Christian | 2020 | COVID-19 aus Sicht der Pneumologie - Langzeitfolgen und Implikationen fur die pneumologische Nachsorge. 145, 15 1086-1092 https://dx.doi.org/10.1055/a-1164-4040 |
| 38. Hydroxychloroquine and azithromycin as a treatment of COVID-19 | Fanin A, Calegari J. Beverina A. Tiraboschi S. | 2020 | Internal and emergency medicine 10.1007/s11739-020-02388-y |
| 39. The Post-COVID-19 Functional Status scale: a tool to measure functional status over time after COVID-19 | Klok, Frederikus A.; Boon, Gudula J. A. M.; Barco, Stefano; Endres, Matthias; Geelhoed, J. J. Miranda; Knauss, Samuel; Rezek, Spencer A.; Spruit, Martijn A.; Vehreschild, Jorg; Siegerink, Bob | 2020 | The European respiratory journal 56, 1 https://dx.doi.org/10.1183/13993003.01494-2020 |
| 40. Existing Data Sources in Clinical Epidemiology: the Danish COVID-19 Cohort | Pottegard A, Kristensen K. B. Reilev M. Lund L. C. Ernst M. T. Hallas J. Thomsen R. W. Christiansen C. F. SÃ¸rensen H. T. Johansen N. B. StÃ¸vring H. Christensen S. Kragh Thomsen M. Husby A. Voldstedlund M. KjÃ¦r J. Brun N. C. | 2020 | Clinical epidemiology 12 875-881 10.2147/CLEP.S257519 |
| 41. Covid-19: symptoms are common after acute phase of disease, Italian study shows | Wise, J. | 2020 | BMJ (Clinical research ed.) 370 m2804 10.1136/bmj.m2804 |
| 42. Post-COVID-19 follow-up clinic: depicting chronicity of a new disease | Rovere Querini, Patrizia; De Lorenzo, Rebecca; Conte, Caterina; Brioni, Elena; Lanzani, Chiara; Yacoub, Mona Rita; Chionna, Raffaella; Martinenghi, Sabina; Vitali, Giordano; Tresoldi, Moreno; Ciceri, Fabio | 2020 | Acta bio-medica : Atenei Parmensis 91, 9-S 22-28 https://dx.doi.org/10.23750/abm.v91i9-S.10146 |
| 43. Coordinated and sustained immune memory responses after mild COVID-19 | Alrubayyi, Aljawharah | 2020 | Nature reviews. Immunology 20, 11 648 https://dx.doi.org/10.1038/s41577-020-00450-6 |

| | | | | |
|---|---|---|---|---|
| 44. | Long COVID - What doesn't kill you may not make you stronger | Ashton, John | 2020 | Journal of the Royal Society of Medicine 113, 11 466-467 https://dx.doi.org/10.1177/0141076820971225 |
| 45. | Long COVID: let patients help define long-lasting COVID symptoms | Anonymous | 2020 | Nature 586, 7828 170 https://dx.doi.org/10.1038/d41586-020-02796-2 |
| 46. | Consequences of infections: SARS-CoV-2 can trigger the dreaded Guillain-Barre syndrome. [German] | Anonymous | 2020 | Klinikarzt 49, 5 188 http://dx.doi.org/10.1055/a-1166-1809 |
| 47. | Rehabilitation of post-COVID-19 patients | Asly, Mouna; Hazim, Asmaa | 2020 | The Pan African medical journal 36 168 https://dx.doi.org/10.11604/pamj.2020.36.168.23823 |
| 48. | Post-Acute COVID-19: An Overview and Approach to Classification | Amenta, Eva M.; Spallone, Amy Rodriguez-Barradas Maria C.; El Sahly, Hana M.; Atmar, Robert L.; Kulkarni, Prathit A. | 2020 | Open Forum Infectious Diseases |
| 49. | Detailed Organ System Analysis of Asymptomatic COVID19 patients using noninvasive mobile tool | Acculi Labs PvtLimited - Bengaluru, India | 2020 | ICTRP |
| 50. | Study for clinical effect of rehabilitation nursing program for patients with novel coronavirus pneumonia (COVID-19) | Affiliated Hospital of Zunyi Medical, University | 2020 | ICTRP |
| 51. | A Randomized Trial of Convalescent Plasma for COVID-19-Potentially Hopeful Signals | Casadevall, Arturo; Joyner, Michael J.; Pirofski, Liise-Anne | 2020 | JAMA 324, 5 455-457 https://dx.doi.org/10.1001/jama.2020.10218 |
| 52. | Post COVID-19 pneumology | Casan Clara, Pere; Martinez Gonzalez, Cristina | 2020 | La neumologia pos-COVID-19 56 Suppl 2 https://dx.doi.org/10.1016/j.arbres.2020.05.009 |
| 53. | Opportunities in Telemedicine, Lessons Learned After COVID-19 and the Way Into the Future | Abdel-Wahab, May; Rosenblatt, Eduardo; Prajogi, Ben; Zubizarretta, Eduardo; Mikhail, Miriam | 2020 | International journal of radiation oncology, biology, physics 108, 2 438-443 https://dx.doi.org/10.1016/j.ijrobp.2020.07.006 |
| 54. | Psychiatric Morbidities among COVID-19 Survivors | Doppalapudi, Lalasa; Lippmann, Steven | 2020 | Southern medical journal 113, 9 466-467 https://dx.doi.org/10.14423/SMJ.0000000000001146 |
| 55. | "Long covid": the Dutch response | Burgers, Jako | 2020 | BMJ (Clinical research ed.) 370 m3202 https://dx.doi.org/10.1136/bmj.m3202 |
| 56. | Covid-19 and chronic fatigue | Williams, Frances M. K.; Muirhead, Nina; Pariante, Carmine | 2020 | BMJ (Clinical research ed.) 370 m2922 https://dx.doi.org/10.1136/bmj.m2922 |
| 57. | COVID-19 Prevalence and Cognitive Deficits in Neurological Patients | Aarhus University, Hospital | 2020 | ClinicalTrials.gov |
| 58. | Coronavirus (COVID-19) antibody response in healthcare staff: what proportion of healthcare staff have COVID-19 antibodies? How long do the antibodies last? Do the antibodies protect against recurring infection? | Abbott | 2020 | ICTRP |
| 59. | Antioxidants as Adjuvant Therapy to Standard Therapy in Patients With COVID-19 | Adrian Palacios-Chavarria, M. D. Principal investigator Unidad Temporal Covid-en Centro Citibanamex | 2020 | ClinicalTrials.gov |
| 60. | HRCT chest imaging in pediartic, adult and geriatric patients with COVID-19 infection | adult, Hrct chest imaging in pediartic //; geriatric patients with, Covid-infection | 2020 | ICTRP |
| 61. | Pilot Study Into LDN and NAD+ for Treatment of Patients With Post-COVID-19 Syndrome | AgelessRx | 2020 | ClinicalTrials.gov |
| 62. | A Study of LAM-002A for the Prevention of Progression of COVID-19 | Ai Therapeutics, Inc | 2020 | ClinicalTrials.gov |
| 63. | Plasma Rich Antibodies From Recovered Patients From COVID19 | Ain Shams, University | 2020 | ClinicalTrials.gov |
| 64. | Measles Vaccine: Is There a Protective Role in COVID 19 Pandemic? | Ain Shams, University | 2020 | ClinicalTrials.gov |

| | | | | |
|---|---|---|---|---|
| 65. | Exchange Transfusion Versus Plasma From Convalescent Patients With Methylene Blue in Patients With COVID-19 | Ain Shams, University | 2020 | ClinicalTrials.gov |
| 66. | Efficacy and Safety of Favipiravir in Management of COVID-19 | Ain Shams, University | 2020 | ClinicalTrials.gov |
| 67. | COVID-19 in Patients With Anosmia in Egypt | Ain Shams, University | 2020 | ClinicalTrials.gov |
| 68. | Application of BCG Vaccine for Immune-prophylaxis Among Egyptian Healthcare Workers During the Pandemic of COVID-19 | Ain Shams, University | 2020 | ClinicalTrials.gov |
| 69. | Ayurveda strength and immunity enhancing protocol and immunity in health care workers : clinical trial of an Ayurveda Intervention | All India Institute Of, Ayurveda | 2020 | ICTRP |
| 70. | Efficacy of Sofosbuvir Plus Ledipasvir in Egyptian Patients With COVID-19 Compared to Standard Treatment | Almaza Military Fever, Hospital | 2020 | ClinicalTrials.gov |
| 71. | An open label physician initiated 28 day phase 2a study of ifenprodil on lung function in confirmed covid-19 infected patients with severe pneumonia | An Open Label Physician Initiated 28 Day Phase 2A Study Of Ifenprodil On Lung Function In Confirmed Covid-19 Infected Patients With Severe, Pneumonia | 2020 | ICTRP |
| 72. | Effect of a Nss to Reduce Complications in Patients With Covid-19 and Comorbidities in Stage III | Anahuac, University | 2020 | ClinicalTrials.gov |
| 73. | Cardiovascular Consequences After COVID-19 | Assistance Publique Hopitaux De, Marseille | 2020 | ClinicalTrials.gov |
| 74. | Protective Role of Inhaled Steroids for Covid-19 Infection | Assistance Publique - Hopitaux de, Paris | 2020 | ClinicalTrials.gov |
| 75. | Epidemiological Characteristics of Coronavirus Infection (SARS-CoV-2) in Patients With MS or NMO | Assistance Publique - Hopitaux de, Paris | 2020 | ClinicalTrials.gov |
| 76. | COVID19-FOIE National Observatory | Assistance Publique - Hopitaux de, Paris | 2020 | ClinicalTrials.gov |
| 77. | Cohort of Patients With Covid-19 Presenting Neurological or Psychiatric Disorders (CoCo-Neurosciences) | Assistance Publique - Hopitaux de, Paris | 2020 | ClinicalTrials.gov |
| 78. | A Multicenter Randomized Trial to Evaluate the Efficacy and Safety of Camostat Mesylate for the Treatment of SARS-CoV-2 Infection - COVID-19 in Ambulatory Adult Patients | Assistance Publique - Hopitaux de, Paris | 2020 | ClinicalTrials.gov |
| 79. | Post Covid Syndrome: Clinical Pattern and Functional Assessment | Assiut, University | 2020 | ClinicalTrials.gov |
| 80. | Post COVID-19 Functional Status in Egypt | Assiut, University | 2020 | ClinicalTrials.gov |
| 81. | Novel COVID-19, A National Analysis | Assiut, University | 2020 | ClinicalTrials.gov |
| 82. | Characteristics and Outcomes of Gastrointestinal Manifestations of COVID-19 | Assiut, University | 2020 | ClinicalTrials.gov |
| 83. | An Open Non-comparative Study of the Efficacy and Safety of Aprotinin in Patients Hospitalized With COVID-19 | Aviron, L. L. C. | 2020 | ClinicalTrials.gov |
| 84. | Long-term Evolution of Pulmonary Involvement of Novel SARS-COV-2 Infection (COVID-19): Follow the Covid Study | Azienda Ospedaliera Universitaria di Bologna Policlinico, S. Orsola Malpighi | 2020 | ClinicalTrials.gov |
| 85. | Rehabilitation Needs After COVID-19 Hospital Treatment | Azienda Unita Sanitaria Locale Reggio, Emilia | 2020 | ClinicalTrials.gov |
| 86. | Treatment With Mesenchymal Stem Cells for Severe Corona Virus Disease 2019(COVID-19) | Beijing, Hospital | 2020 | ClinicalTrials.gov |

| | | | |
|---|---|---|---|
| 87. Study for the exercise rehabilitation therapy for the dysfunction of cured discharged novel coronavirus pneumonia (COVID-19) patients | Beijing Sports, University | 2020 | ICTRP |
| 88. Novel coronavirus pneumonia (COVID-19) loaded autologous dendritic cell trial | Beijing, T. I. L. Therapeutics | 2020 | ICTRP |
| 89. Development and application of TCM body regulating protection scheme for the convalescent population of novel coronavirus pneumonia (COVID-19) | Beijing University of Chinese, Medicine | 2020 | ICTRP |
| 90. The Clinical Study of Carrimycin on Treatment Patients With COVID-19 | Beijing YouAn, Hospital | 2020 | ClinicalTrials.gov |
| 91. A Study to Assess Pulsed Inhaled Nitric Oxide vs Placebo in Subjects With Mild or Moderate COVID-19 | Bellerophon Pulse, Technologies | 2020 | ClinicalTrials.gov |
| 92. Corticosteroid Nasal Spray in COVID-19 Anosmia | Benha, University | 2020 | ClinicalTrials.gov |
| 93. Ulinastatin for COVID-19 in patients with breathlessness | Bharat, Serums; Vaccines ltd - Mimbai, India | 2020 | ICTRP |
| 94. Quality of life in COVID-19 survivors | Board of directors, Ciro | 2020 | ICTRP |
| 95. Functional status and health status in COVID-19 patients pre and post rehabilitation | Board of Directors, Ciro | 2020 | ICTRP |
| 96. Home Management of Adult Egyptian Mild COVID-19 Cases | Cairo, University | 2020 | ClinicalTrials.gov |
| 97. Dysphagia and Dysphonia Outcomes in SARS CoV-2 (COVID-19) Infection (DYADS Study) | Cambridge University Hospitals, N. H. S. Foundation Trust | 2020 | ClinicalTrials.gov |
| 98. CAP-1002 in Severe COVID-19 Disease | Capricor, Inc | 2020 | ClinicalTrials.gov |
| 99. Factors Associated With Clinical Outcomes in Patients Hospitalized for Covid-19 in GHT-93 Est | Centre Hospitalier Intercommunal Robert, Ballanger | 2020 | ClinicalTrials.gov |
| 100. Chronic Fatigue Etiology and Recovery in Covid-19 Patients: the Role of Fatigability | Centre Hospitalier Universitaire de Saint, Etienne | 2020 | ClinicalTrials.gov |
| 101. Impact of a Minimal Psychoeducational Intervention on Anxiety Among Hospitalized COVID-19 Patients in Denmark | Copenhagen University Hospital, Hvidovre | 2020 | ClinicalTrials.gov |
| 102. Long COVID and chronic COVID syndromes | Halpin, Stephen; Connor, Rory; Sivan, Manoj | 2020 | J. med. virol |
| **Other irrelevant (n=11)** | | | |
| 103. Association between Long COVID and Overweight/Obesity | Vimercati, L.; De Maria, L.; Quarato, M.; Caputi, A.; Gesualdo, L.; Migliore, G.; Cavone, D.; Sponselli, S.; Pipoli, A.; Inchingolo, F.; Scarano, A.; Lorusso, F.; Stefanizzi, P.; Tafuri, S. | 2021 | Journal of Clinical Medicine 10, 18 14 https://dx.doi.org/10.3390/jcm10184143 |
| 104. Anxiety and depression symptoms after COVID-19 infection: results from the COVID Symptom Study app | Klaser, K.; Thompson, E. J.; Nguyen, L. H.; Sudre, C. H.; Antonelli, M.; Murray, B.; Canas, L. S.; Molteni, E.; Graham, M. S.; Kerfoot, E.; Chen, L.; Deng, J.; May, A.; Hu, C.; Guest, A.; Selvachandran, S.; Drew, D. A.; Modat, M.; Chan, A. T.; Wolf, J.; Spector, T. D.; Hammers, A.; Duncan, E. L.; Ourselin, S.; Steves, C. J. | 2021 | MedRxiv : the Preprint Server for Health Sciences 8 https://dx.doi.org/10.1101/2021.07.07.21260137 |
| 105. Assessment and characterisation of post-COVID-19 manifestations | Kamal, Marwa; Abo Omirah, Marwa; Hussein, Amal; Saeed, Haitham | 2021 | International journal of clinical practice 75, 3 e13746 https://dx.doi.org/10.1111/ijcp.13746 |
| 106. Comparing the impact of Hydroxychloroquine based regimens and standard treatment on COVID-19 patient outcomes: a retrospective cohort study | Almazrou Sh, Almalki Z. S. Alanazi A. S. Alqahtani A. M. AlGhamd S. M. | 2020 | Saudi pharmaceutical journal 10.1016/j.jsps.2020.09.019 |

| | | | |
|---|---|---|---|
| 107. Cerebral Microbleeds and Leukoencephalopathy in Critically Ill Patients With COVID-19 | Agarwal S, Jain R. Dogra S. Krieger P. Lewis A. Nguyen V. Melmed K. Galetta S. | 2020 | Stroke; a journal of cerebral circulation STROKEAHA120030940 10.1161/STROKEAHA.120.030940 |
| 108. Pulmonary Function in Early Follow-up of patients with COVID-19 Pneumonia | Tabernero Huguet E, Urrutia Gajarte A. Ruiz Iturriaga L. A. Serrano Fernandez L. Marina Malanda N. Iriberri Pascual M. Zalacain Jorge R. | 2020 | Archivos de bronconeumologia 10.1016/j.arbres.2020.07.017 |
| 109. Post-traumatic stress disorder symptoms in COVID-19 survivors: online population survey | Chamberlain, Samuel R.; Grant, Jon E.; Trender, William; Hellyer, Peter; Hampshire, Adam | 2021 | BJPsych open 7, 2 e47 https://dx.doi.org/10.1192/bjo.2021.3 |
| 110. Acute and persistent symptoms in non-hospitalized PCR-confirmed COVID-19 patients (preprint) | Bliddal S, Banasik K. Pedersen O. B. Nissen I. Cantwell L. Schwinn M. Tulstrup M. Westergaard D. Ullum H. Brunak S. Tommerup N. Feenstra B. Geller F. Ostrowski S. R. Groenbaek K. Nielsen C. H. Nielsen S. D. Feldt-Rasmussen U. | 2021 | Medrxiv 2021.01.22.21249945 10.1101/2021.01.22.21249945 |
| 111. Cardiovascular involvement and its relationship with the severity of the acute phase and persistent symptoms during recovery from COVID-19 infection. [Spanish] | Parodi, J. B.; Jacob, P. B.; Toledo, G. C.; Micali, R. G.; Iacino, M. P.; Sotelo, B.; Bruno, C.; Pelletier, M.; Juarez, W. M.; Huerin, M. S. | 2021 | Revista Argentina de Cardiologia 89(4) 332-339 http://dx.doi.org/10.7775/rac.es.v89.i4.20426 |
| 112. Clinical and epidemiological characteristics and outcomes of Coronavirus disease-19 patients in a large longitudinal study | Merza, Muayad Aghali; Aswad, Serwan Mohamamed; Sulaiman, Hushyar Musa; Abdulah, Deldar Morad; Rasheed, Waleed Salih; Taib, Nezar Ismet | 2021 | International journal of health sciences 15, 4 29-41 |
| 113. SARS-CoV-2 pneumonia follow-up and long COVID in primary care: A retrospective observational study in Madrid city | Ares-Blanco, Sara; Perez Alvarez, Marta; Gefaell Larrondo, Ileana; Munoz, Cristina; Aguilar Ruiz, Vanesa; Castelo Jurado, Marta; Guisado-Clavero, Marina | 2021 | PloS one 16, 6 9 e0257604 https://dx.doi.org/10.1371/journal.pone.0257604 |
| **Publication reports a subset of results already included (n=2)** | | | |
| 114. The number of symptoms at the acute COVID-19 phase is associated with anxiety and depressive long-term post-COVID symptoms: A multicenter study | Fernandez-de-Las-Penas, Cesar; Pellicer-Valero, Oscar J.; Navarro-Pardo, Esperanza; Rodriguez-Jimenez, Jorge; Martin-Guerrero, Jose D.; Cigaran-Mendez, Margarita | 2021 | Journal of psychosomatic research 150 110625 https://dx.doi.org/10.1016/j.jpsychores.2021.110625 |
| 115. Similar prevalence of long-term post-COVID symptoms in patients with asthma: A case-control study | Fernandez-de-Las-Penas, C.; Torres-Macho, J.; Velasco-Arribas, M.; Arias-Navalon, J. A.; Guijarro, C.; Hernandez-Barrera, V.; Canto-Diez, M. | 2021 | Journal of Infection 83, 2 237-279 https://dx.doi.org/10.1016/j.jinf.2021.04.034 |
| **Too short follow-up (n=115)** | | | |
| 116. Persisting symptoms in patients following hospital admission with COVID-19: An observational cohort study | Pheasant, C.; Dixon, N.; Day, P.; Roberts, A.; Petruso-Osborn, N.; Lawley, J.; Wilson, E.; McWilliams, D. | 2021 | Intensive Care Medicine Experimental. Conference: European Society of Intensive Care Medicine Annual Congress, ESICM 9, SUPPL 1 http://dx.doi.org/10.1186/s40635-021-00415-6 |
| 117. Post-acute effects of SARS-CoV-2 infection in individuals not requiring hospital admission: a Danish population-based cohort study | Lund, Lars Christian; Hallas, Jesper; Nielsen, Henrik; Koch, Anders; Mogensen, Stine Hasling; Brun, Nikolai Constantin; Christiansen, Christian Fynbo; Thomsen, Reimar Wernich; Pottegard, Anton | 2021 | The Lancet. Infectious diseases 21, 10 1373-1382 https://dx.doi.org/10.1016/S1473-3099(21)00211-5 |
| 118. Post-COVID-19 functional status: Relation to age, smoking, hospitalization, and previous comorbidities | Mohamed Hussein, A. A.; Saad, M.; Zayan, H. E.; Abdelsayed, M.; Moustafa, M.; Ezzat, A. R.; Helmy, R.; Abd-Elaal, H.; Aly, K.; Abdelrheem, S.; Sayed, I. | 2021 | Annals of Thoracic Medicine 16, 3 260-265 https://dx.doi.org/10.4103/atm.atm_606_20 |
| 119. Duration and Risk Factors of Post-COVID Symptoms Following Recovery Among the Medical Doctors in Bangladesh | Sultana, Sarmin; Islam, Mohammad Tanvir; Salwa, Marium; Zakir Hossain, Shah M.; Hasan, Md Nazmul; Masum, Abdullah A.; Khan, Abed H.; Khan, Md Maruf Haque; Haque, M. Atiqul | 2021 | Cureus 13, 5 e15351 https://dx.doi.org/10.7759/cureus.15351 |
| 120. Longitudinal Radiological Findings in Patients With COVID-19 With Different Severities: From Onset to Long-Term Follow-Up After Discharge | Zhao, Y.; Wang, D.; Mei, N.; Yin, B.; Li, X.; Zheng, Y.; Xiao, A.; Yu, X.; Qiu, X.; Lu, Y.; Liu, L. | 2021 | Frontiers in Medicine 8 711435 https://dx.doi.org/10.3389/fmed.2021.711435 |

| | | | |
|---|---|---|---|
| 121. Post-acute COVID-19 syndrome. Incidence and risk factors: A Mediterranean cohort study | Moreno-Perez, Oscar; Merino, Esperanza; Leon-Ramirez, Jose-Manuel; Andres, Mariano; Ramos, Jose Manuel; Arenas-Jimenez, Juan; Asensio, Santos; Sanchez, Rosa; Ruiz-Torregrosa, Paloma; Galan, Irene; Scholz, Alexander; Amo, Antonio; Gonzalez-delaAleja, Pilar; Boix, Vicente; Gil, Joan; Covid Alc research group | 2021 | The Journal of infection 82, 3 378-383 https://dx.doi.org/10.1016/j.jinf.2021.01.004 |
| 122. Prevalence and Predictors of Persistence of COVID-19 Symptoms in Older Adults: A Single-Center Study | Tosato, M.; Carfi, A.; Martis, I.; Pais, C.; Ciciarello, F.; Rota, E.; Tritto, M.; Salerno, A.; Zazzara, M. B.; Martone, A. M.; Paglionico, A.; Petricca, L.; Brandi, V.; Capalbo, G.; Picca, A.; Calvani, R.; Marzetti, E.; Landi, F.; Gemelli Against, Covid-Post-Acute Care Team | 2021 | Journal of the American Medical Directors Association 22, 9 1840-1844 https://dx.doi.org/10.1016/j.jamda.2021.07.003 |
| 123. Assessment of Prolonged Physiological and Behavioral Changes Associated With COVID-19 Infection | Radin, Jennifer M.; Quer, Giorgio; Ramos, Edward; Baca-Motes, Katie; Gadaleta, Matteo; Topol, Eric J.; Steinhubl, Steven R. | 2021 | JAMA network open 4, 7 e2115959 https://dx.doi.org/10.1001/jamanetworkopen.2021.15959 |
| 124. Posttraumatic Stress Disorder in Patients After Severe COVID-19 Infection | Janiri, Delfina; Carfi, Angelo; Kotzalidis, Georgios D.; Bernabei, Roberto; Landi, Francesco; Sani, Gabriele; Gemelli Against, Covid-Post-Acute Care Study Group | 2021 | JAMA psychiatry 78, 5 567-569 https://dx.doi.org/10.1001/jamapsychiatry.2021.0109 |
| 125. Late Conditions Diagnosed 1-4 Months Following an Initial Coronavirus Disease 2019 (COVID-19) Encounter: A Matched-Cohort Study Using Inpatient and Outpatient Administrative Data-United States, 1 March-30 June 2020 | Chevinsky, J. R.; Tao, G.; Lavery, A. M.; Kukielka, E. A.; Click, E. S.; Malec, D.; Kompaniyets, L.; Bruce, B. B.; Yusuf, H.; Goodman, A. B.; Dixon, M. G.; Nakao, J. H.; Datta, S. D.; MacKenzie, W. R.; Kadri, S. S.; Saydah, S.; Giovanni, J. E.; Gundlapalli, A. V. | 2021 | Clinical Infectious Diseases 73, Suppl 1 S5-S16 https://dx.doi.org/10.1093/cid/ciab338 |
| 126. Persistent Poor Health after COVID-19 Is Not Associated with Respiratory Complications or Initial Disease Severity | Townsend, Liam; Dowds, Joanne; O'Brien, Kate; Sheill, Grainne; Dyer, Adam H.; O'Kelly, Brendan; Hynes, John P.; Mooney, Aoife; Dunne, Jean; Ni Cheallaigh, Cliona; O'Farrelly, Cliona; Bourke, Nollaig M.; Conlon, Niall; Martin-Loeches, Ignacio; Bergin, Colm; Nadarajan, Parthiban; Bannan, Ciaran | 2021 | Annals of the American Thoracic Society 18, 6 997-1003 https://dx.doi.org/10.1513/AnnalsATS.202009-1175OC |
| 127. Long-Term Symptoms Among Adults Tested for SARS-CoV-2 - United States, January 2020-April 2021 | Wanga, Valentine; Chevinsky, Jennifer R.; Dimitrov, Lina V.; Gerdes, Megan E.; Whitfield, Geoffrey P.; Bonacci, Robert A.; Nji, Miriam A. M.; Hernandez-Romieu, Alfonso C.; Rogers-Brown, Jessica S.; McLeod, Tim; Rushmore, Julie; Lutfy, Caitlyn; Bushman, Dena; Koumans, Emilia; Saydah, Sharon; Goodman, Alyson B.; Coleman King, Sallyann M.; Jackson, Brendan R.; Cope, Jennifer R. | 2021 | MMWR. Morbidity and mortality weekly report 70, 36 1235-1241 https://dx.doi.org/10.15585/mmwr.mm7036a1 |
| 128. Impact of severe SARS-CoV-2 infection on nutritional status and subjective functional loss in a prospective cohort of COVID-19 survivors | Quilliot, D.; Gerard, M.; Bonsack, O.; Malgras, A.; Vaillant, M. F.; Di Patrizio, P.; Jaussaud, R.; Ziegler, O.; Nguyen-Thi, P. L. | 2021 | BMJ Open 11, 7 e048948 https://dx.doi.org/10.1136/bmjopen-2021-048948 |
| 129. Coronavirus disease-19 and headache; impact on pre-existing and characteristics of de novo: a cross-sectional study | Al-Hashel, J. Y.; Abokalawa, F.; Alenzi, M.; Alroughani, R.; Ahmed, S. F. | 2021 | Journal of Headache & Pain 22, 1 97 https://dx.doi.org/10.1186/s10194-021-01314-7 |
| 130. Study of Post-COVID-19 Syndrome in Saudi Arabia | Mahmoud, Manal H.; Alghamdi, Fahad A.; Alghamdi, Ghaida A.; Alkhotani, Loai A.; Alrehaili, Mohammad A.; El-Deeb, Dalia K. | 2021 | Cureus 13, 9 e17787 https://dx.doi.org/10.7759/cureus.17787 |
| 131. Prolonged Symptoms After COVID-19 Infection in Outpatients | Ramakrishnan, Aditi; Zreloff, Jennifer; Moore, Miranda A.; Bergquist, Sharon H.; Cellai, Michele; Higdon, Jason; O'Keefe, James B.; Roberts, David; Wu, Henry M. | 2021 | Open forum infectious diseases 8, 3 ofab060 https://dx.doi.org/10.1093/ofid/ofab060 |
| 132. Clinical and Laboratory Follow-up After Hospitalization for COVID-19 at an Italian Tertiary Care Center | Spinicci, Michele; Vellere, Iacopo; Graziani, Lucia; Tilli, Marta; Borchi, Beatrice; Mencarini, Jessica; Campolmi, Irene; Gori, Leonardo; Rasero, Laura; Fattirolli, Francesco; Olivotto, Iacopo; Lavorini, Federico; Marchionni, Niccolo; Zammarchi, Lorenzo; Bartoloni, Alessandro; Careggi Post-acute, Covid-Study Group; | 2021 | Open forum infectious diseases 8, 3 3 ofab049 https://dx.doi.org/10.1093/ofid/ofab049 |

| | | | |
|---|---|---|---|
| | Fumagalli C, Silverii M. V. Ciani L. Zocchi C. Tassetti L. Marcucci R. Giusti B. Livi L. Giovannoni L. Parronchi P. Almerigogna F. Annunziato F. Mazzoni A. Maggi L. Liotta F. Cosmi L. Vultaggio A. Matucci A. Sticci S. Donati M. Defraia C. Giansanti F. Bacherini D. | | |
| 133. A mediating role for mental health in associations between COVID-19-related self-stigma, PTSD, quality of life, and insomnia among patients recovered from COVID-19 | Mahmoudi, Hosein; Saffari, Mohsen; Movahedi, Mahmoud; Sanaeinasab, Hormoz; Rashidi-Jahan, Hojat; Pourgholami, Morteza; Poorebrahim, Ali; Barshan, Jalal; Ghiami, Milad; Khoshmanesh, Saman; Potenza, Marc N.; Lin, Chung-Ying; Pakpour, Amir H. | 2021 | Brain and behavior 11, 5 e02138 https://dx.doi.org/10.1002/brb3.2138 |
| 134. Mapping each pre-existing condition's association to short-term and long-term COVID-19 complications | Venkatakrishnan, A. J.; Pawlowski, Colin; Zemmour, David; Hughes, Travis; Anand, Akash; Berner, Gabriela; Kayal, Nikhil; Puranik, Arjun; Conrad, Ian; Bade, Sairam; Barve, Rakesh; Sinha, Purushottam; O'Horo, John C.; Badley, Andrew D.; Halamka, John; Soundararajan, Venky | 2021 | NPJ digital medicine 4, 1 117 https://dx.doi.org/10.1038/s41746-021-00484-7 |
| 135. The COVID-19 Sequelae: A Cross-Sectional Evaluation of Post-recovery Symptoms and the Need for Rehabilitation of COVID-19 Survivors | Iqbal, Ayman; Iqbal, Kinza; Arshad Ali, Shajeea; Azim, Dua; Farid, Eisha; Baig, Mirza D.; Bin Arif, Taha; Raza, Mohammad | 2021 | Cureus 13, 2 e13080 https://dx.doi.org/10.7759/cureus.13080 |
| 136. Central Sensitization Phenotypes in Post Acute Sequelae of SARS-CoV-2 Infection (PASC): Defining the Post COVID Syndrome | Bierle, D. M.; Aakre, C. A.; Grach, S. L.; Salonen, B. R.; Croghan, I. T.; Hurt, R. T.; Ganesh, R. | 2021 | Journal of Primary Care & Community Health 12 https://dx.doi.org/10.1177/21501327211030826 |
| 137. Hypertension as a sequela in patients of SARS-CoV-2 infection | Chen, Ganxiao; Li, Xun; Gong, Zuojiong; Xia, Hao; Wang, Yao; Wang, Xuefen; Huang, Yan; Barajas-Martinez, Hector; Hu, Dan | 2021 | PloS one 16, 4 e0250815 https://dx.doi.org/10.1371/journal.pone.0250815 |
| 138. Pulmonary function and functional capacity in COVID-19 survivors with persistent dyspnoea | Cortes-Telles, Arturo; Lopez-Romero, Stephanie; Figueroa-Hurtado, Esperanza; Pou-Aguilar, Yuri Noemi; Wong, Alyson W.; Milne, Kathryn M.; Ryerson, Christopher J.; Guenette, Jordan A. | 2021 | Respiratory physiology & neurobiology 288 103644 https://dx.doi.org/10.1016/j.resp.2021.103644 |
| 139. COVID-19 Symptoms: Longitudinal Evolution and Persistence in Outpatient Settings | Nehme, M.; Braillard, O.; Alcoba, G.; Aebischer Perone, S.; Courvoisier, D.; Chappuis, F.; Guessous, I.; Covicare, Team | 2021 | Annals of Internal Medicine 174, 5 723-725 https://dx.doi.org/10.7326/M20-5926 |
| 140. Risk Predictors and Symptom Features of Long COVID Within a Broad Primary Care Patient Population Including Both Tested and Untested Patients | Jones, Rupert; Davis, Andrew; Stanley, Brooklyn; Julious, Steven; Ryan, Dermot; Jackson, David J.; Halpin, David M. G.; Hickman, Katherine; Pinnock, Hilary; Quint, Jennifer K.; Khunti, Kamlesh; Heaney, Liam G.; Oliver, Phillip; Siddiqui, Salman; Pavord, Ian; Jones, David H. M.; Hyland, Michael; Ritchie, Lewis; Young, Pam; Megaw, Tony; Davis, Steve; Walker, Samantha; Holgate, Stephen; Beecroft, Sue; Kemppinen, Anu; Appiagyei, Francis; Roberts, Emma-Jane; Preston, Megan; Hardjojo, Antony; Carter, Victoria; van Melle, Marije; Price, David | 2021 | Pragmatic and observational research 12 93-104 https://dx.doi.org/10.2147/POR.S316186 |
| 141. Recognition of Long-COVID-19 Patients in a Canadian Tertiary Hospital Setting: A Retrospective Analysis of Their Clinical and Laboratory Characteristics | Kozak, Robert; Armstrong, Susan M.; Salvant, Elsa; Ritzker, Claudia; Feld, Jordan; Biondi, Mia J.; Tsui, Hubert | 2021 | Pathogens (Basel, Switzerland) 10, 10 https://dx.doi.org/10.3390/pathogens10101246 |
| 142. Fatigue, perceived stigma, self-reported cognitive deficits and psychological morbidity in patients recovered from COVID-19 infection | Grover, Sandeep; Sahoo, Swapnajeet; Mishra, Eepsita; Gill, Kanwarbir Singh; Mehra, Aseem; Nehra, Ritu; Suman, Aarzoo; Bhalla, Ashish; Puri, Goverdhan Dutt | 2021 | Asian journal of psychiatry 64 102815 https://dx.doi.org/10.1016/j.ajp.2021.102815 |
| 143. Acute and longer-term psychological distress associated with testing positive for COVID-19: longitudinal evidence from a population-based study of US adults | Daly, Michael; Robinson, Eric | 2021 | medRxiv : the preprint server for health sciences https://dx.doi.org/10.1101/2021.03.25.2125432 6 |

| | | | |
|---|---|---|---|
| 144. Outcomes of Cardiovascular Magnetic Resonance Imaging in Patients Recently Recovered From Coronavirus Disease 2019 (COVID-19) | Puntmann, Valentina O.; Carerj, M. Ludovica; Wieters, Imke; Fahim, Masia; Arendt, Christophe; Hoffmann, Jedrzej; Shchendrygina, Anastasia; Escher, Felicitas; Vasa-Nicotera, Mariuca; Zeiher, Andreas M.; Vehreschild, Maria; Nagel, Eike | 2020 | JAMA cardiology 5, 11 1265-1273 https://dx.doi.org/10.1001/jamacardio.2020.3557 |
| 145. COVIDApp: a Health Application as an Innovative Strategy for the Management and Follow-Up of the COVID-19 Pandemic in Long-Term Care Facilities in Catalonia | Echeverria P, Mas-Bergas M. A. Puig J. Isnard M. Massot M. Vedia M. C. Peiro R. Ordorica Y. Pablo S. Ulldemolins M. Iruela M. Balart D. Ruiz-Serrats J. M. Herms J. Clotet B. Negredo E. | 2020 | JMIR public health and surveillance 10.2196/21163 |
| 146. Predictors of functional dependence after COVID-19: a retrospective examination among veterans | Leigh Ae, McCall J. Burke R. V. Rome R. Raines A. M. | 2020 | American journal of physical medicine & rehabilitation 10.1097/PHM.0000000000001614 |
| 147. Robust, reproducible clinical patterns in hospitalised patients with COVID-19 (preprint) | Millar Je, Neyton L. Seth S. Dunning J. Merson L. Murthy S. Russell C. D. Keating S. Swets M. Sudre C. H. Spector T. D. Ourselin S. Steves C. J. Wolf J. Docherty A. B. Harrison E. M. Openshaw P. J. M. Semple M. G. Baillie J. K. | 2020 | Medrxiv 2020.08.14.20168088 10.1101/2020.08.14.20168088 |
| 148. Clinico-demographic profile & hospital outcomes of COVID-19 patients admitted at a tertiary care centre in north India | Mohan A, Tiwari P. Bhatnagar S. Patel A. Maurya A. Dar L. Pahuja S. Garg R. Gupta N. Sahoo B. Gupta R. Meena V. P. Vig S. Pandit A. Mittal S. Madan K. Hadda V. Dwivedi T. Choudhary A. Brijwal M. Soneja M. Guleria R. Ratre B. Kumar B. Bhopale S. Panda S. Singh A. R. Singh S. Wundavalli L. | 2020 | Indian journal of medical research 10.4103/ijmr.IJMR_1788_20 |
| 149. Anesthesiologists' and intensive care providers' exposure to COVID-19 infection in a New York City academic center: a prospective cohort study assessing symptoms and COVID-19 antibody testing | Morcuende M, Guglielminotti J. Landau R. | 2020 | Anesthesia and analgesia 10.1213/ANE.0000000000005056 |
| 150. The Course of Mild and Moderate COVID-19 Infections-The Unexpected Long-Lasting Challenge | Xia L, Chen J. Friedemann T. Yang Z. Ling Y. Liu X. Lu S. Li T. Song Z. Huang W. Lu Y. Schroder S. Lu H. | 2020 | Open forum infectious diseases 7, 9 ofaa286 10.1093/ofid/ofaa286 |
| 151. Smell and taste in CoViD-19 patients: the forgotten sense | Gamba P, Zaniboni A. | 2020 | Recenti progressi in medicina 111, 10 614-618 10.1701/3453.34422 |
| 152. Population-scale longitudinal mapping of COVID-19 symptoms, behaviour and testing | Allen, William E.; Altae-Tran, Han; Briggs, James; Jin, Xin; McGee, Glen; Shi, Andy; Raghavan, Rumya; Kamariza, Mireille; Nova, Nicole; Pereta, Albert; Danford, Chris; Kamel, Amine; Gothe, Patrik; Milam, Evrhet; Aurambault, Jean; Primke, Thorben; Li, Weijie; Inkenbrandt, Josh; Huynh, Tuan; Chen, Evan; Lee, Christina; Croatto, Michael; Bentley, Helen; Lu, Wendy; Murray, Robert; Travassos, Mark; Coull, Brent A.; Openshaw, John; Greene, Casey S.; Shalem, Ophir; King, Gary; Probasco, Ryan; Cheng, David R.; Silbermann, Ben; Zhang, Feng; Lin, Xihong | 2020 | Nature human behaviour 4, 9 972-982 https://dx.doi.org/10.1038/s41562-020-00944-2 |
| 153. Population-scale Longitudinal Mapping of COVID-19 Symptoms, Behavior, and Testing Identifies Contributors to Continued Disease Spread in the United States | Allen, William E.; Altae-Tran, Han; Briggs, James; Jin, Xin; McGee, Glen; Raghavan, Rumya; Shi, Andy; Kamariza, Mireille; Nova, Nicole; Pereta, Albert; Danford, Chris; Kamel, Amine; Gothe, Patrik; Milam, Evrhet; Aurambault, Jean; Primke, Thorben; Li, Claire; Inkenbrandt, Josh; Huynh, Tuan; Chen, Evan; Lee, Christina; Croatto, Michael; Bentley, Helen; Lu, Wendy; Murray, Robert; Travassos, Mark; Openshaw, John; Coull, Brent; Greene, Casey; Shalem, Ophir; King, Gary; Probasco, Ryan; Cheng, David; Silbermann, Ben; Zhang, Feng; Lin, Xihong | 2020 | medRxiv : the preprint server for health sciences https://dx.doi.org/10.1101/2020.06.09.20126813 |

| | | | |
|---|---|---|---|
| 154. Symptom Characterization and Outcomes of Sailors in Isolation After a COVID-19 Outbreak on a US Aircraft Carrier | Alvarado, Gadiel R.; Pierson, Benjamin C.; Teemer, Eric S.; Gama, Hector J.; Cole, Ronald D.; Jang, Samuel S. | 2020 | JAMA network open 3, 10 e2020981 https://dx.doi.org/10.1001/jamanetworkopen.2020.20981 |
| 155. Xerostomia, gustatory and olfactory dysfunctions in patients with COVID-19 | Fantozzi Pj, Pampena E. Di Vanna D. Pellegrino E. Corbi D. Mammucari S. Alessi F. Pampena R. Bertazzoni G. Minisola S. Mastroianni C. M. Polimeni A. Romeo U. Villa A. | 2020 | American journal of otolaryngology 41, 6 102721 10.1016/j.amjoto.2020.102721 |
| 156. Correlation between immune response and self-reported depression during convalescence from COVID-19 | Yuan, Bo; Li, Weixin; Liu, Hanqing; Cai, Xin; Song, Shuo; Zhao, Jia; Hu, Xiaopeng; Li, Zhiwen; Chen, Yongxin; Zhang, Kai; Liu, Zhiyong; Peng, Jing; Wang, Cheng; Wang, Jianchun; An, Yawen | 2020 | Brain, behavior, and immunity 88 39-43 https://dx.doi.org/10.1016/j.bbi.2020.05.062 |
| 157. Smell and taste dysfunctions in COVID-19 are associated with younger age in ambulatory settings - a multicenter cross-sectional study | Izquierdo-Dominguez A, Rojas-Lechuga M. J. Chiesa-Estomba C. Calvo-Henriquez C. Ninchritz-Becerra E. Soriano-Reixach M. Poletti-Serafini D. Villarreal I. M. Maza-Solano J. M. Moreno-Luna R. Villarroel P. P. Mateos-Serrano B. Agudelo D. Valcarcel F. Del Cuvillo A. Santamaria A. Marino-Sanchez F. Aguilar J. Verges P. Inciarte A. Soriano A. Mullol J. Alobid I. | 2020 | Journal of investigational allergology & clinical immunology 10.18176/jiaci.0595 |
| 158. A Prospective Study of Neurologic Disorders in Hospitalized COVID-19 Patients in New York City | Frontera Ja, Sabadia S. Lalchan R. Fang T. Flusty B. Millar-Vernetti P. Snyder T. Berger S. Yang D. Granger A. Morgan N. Patel P. Gutman J. Melmed K. Agarwal S. Bokhari M. Andino A. Valdes E. Omari M. Kvernland A. Lillemoe K. Chou S. H. McNett M. Helbok R. Mainali S. Fink E. L. Robertson C. Schober M. Suarez J. I. Ziai W. Menon D. Friedman D. Friedman D. Holmes M. Huang J. Thawani S. Howard J. Abou-Fayssal N. Krieger P. Lewis A. Lord A. S. Zhou T. Kahn D. E. Czeisler B. M. Torres J. Yaghi S. Ishida K. Scher E. de Havenon A. Placantonakis D. Liu M. Wisniewski T. Troxel A. B. Balcer L. Galetta S. | 2020 | Neurology 10.1212/WNL.0000000000010979 |
| 159. Response System for and Epidemiological Features of COVID-19 in Gyeongsangnam-do Province in South Korea | Wi Ym, Lim S. J. Kim S. H. Lim S. Lee S. J. Ryu B. H. Hong S. I. Cho O. H. Moon K. Hong K. W. Kim S. Bae I. G. | 2020 | Clinical infectious diseases 10.1093/cid/ciaa967 |
| 160. Characteristics of Hospitalized COVID-19 Patients Discharged and Experiencing Same-Hospital Readmission - United States, March-August 2020 | Lavery, Amy M.; Preston, Leigh Ellyn; Ko, Jean Y.; Chevinsky, Jennifer R.; DeSisto, Carla L.; Pennington, Audrey F.; Kompaniyets, Lyudmyla; Datta, S. Deblina; Click, Eleanor S.; Golden, Thomas; Goodman, Alyson B.; Mac Kenzie, William R.; Boehmer, Tegan K.; Gundlapalli, Adi V. | 2020 | MMWR. Morbidity and mortality weekly report 69, 45 1695-1699 https://dx.doi.org/10.15585/mmwr.mm6945e2 |
| 161. Clinical characteristics and outcomes of adult patients admitted with COVID-19 in East London: a retrospective cohort analysis (preprint) | Cheng D, Calderwood C. Skyllberg E. Ainley A. | 2020 | Medrxiv 2020.10.08.20193623 10.1101/2020.10.08.20193623 |
| 162. Persistent symptoms 3â€…months after a SARS-CoV-2 infection: the post-COVID-19 syndrome? | GoÃ«rtz, Yvonne M. J.; Van Herck, Maarten; Delbressine, Jeannet M.; Vaes, Anouk W.; Meys, Roy; Machado, Felipe V. C.; Houben-Wilke, Sarah; Burtin, Chris; Posthuma, Rein; Franssen, Frits M. E.; van Loon, Nicole; Hajian, Bita; Spies, Yvonne; Vijlbrief, Herman; van â€™t Hul, Alex J.; Janssen, Daisy J. A.; Spruit, Martijn A. | 2020 | ERJ Open Research 00542-2020 |
| 163. The clinical course of COVID-19 in the outpatient setting: a prospective cohort study (preprint) | Blair Pw, Brown D. M. Jang M. Antar A. A. R. Keruly J. C. Bachu V. Townsend J. L. Tornheim J. A. Keller S. C. Sauer L. Thomas D. L. Manabe Y. C. | 2020 | Medrxiv 2020.09.01.20184937 10.1101/2020.09.01.20184937 |
| 164. Chest radiography is a poor predictor of respiratory symptoms and functional impairment in survivors of severe COVID-19 pneumonia | D'Cruz, Rebecca F.; Waller, Michael D.; Perrin, Felicity; Periselneris, Jimstan; Norton, Sam; Smith, Laura-Jane; Patrick, Tanya; Walder, David; Heitmann, Amadea; Lee, Kai; Madula, Rajiv; McNulty, William; Macedo, Patricia; Lyall, Rebecca; Warwick, Geoffrey; | 2021 | ERJ open research 7, 1 https://dx.doi.org/10.1183/23120541.00655-2020 |

| | | | |
|---|---|---|---|
| | Galloway, James B.; Birring, Surinder S.; Patel, Amit; Patel, Irem; Jolley, Caroline J. | | |
| 165. A Follow-up Study of Recovered Patients with COVID-19 in Wuhan, China | Luo S, Guo Y. Zhang X. Xu H. | 2020 | International journal of infectious diseases 10.1016/j.ijid.2020.05.119 |
| 166. A one-year hospital-based prospective COVID-19 open-cohort in the Eastern Mediterranean region: the Khorshid COVID Cohort (KCC) study | Sami R, Soltaninejad F. Amra B. Naderi Z. Haghjooy Javanmard S. Iraj B. Haji Ahmadi S. Shayganfar A. Dehghan M. Khademi N. Sadat Hosseini N. Mortazavi M. Mansourian M. Mananas M. A. Marateb H. R. Adibi P. | 2020 | PloS one 15, 11 e0241537 10.1371/journal.pone.0241537 |
| 167. Anxiety and depression in COVID-19 survivors: Role of inflammatory and clinical predictors | Mazza, Mario Gennaro; De Lorenzo, Rebecca; Conte, Caterina; Poletti, Sara; Vai, Benedetta; Bollettini, Irene; Melloni, Elisa Maria Teresa; Furlan, Roberto; Ciceri, Fabio; Rovere-Querini, Patrizia; group, Covid- BioB Outpatient Clinic Study; Benedetti, Francesco | 2020 | Brain, behavior, and immunity 89 594-600 https://dx.doi.org/10.1016/j.bbi.2020.07.037 |
| 168. Anxiety and depression symptoms after virological clearance of COVID-19: a cross-sectional study in Milan, Italy | Tomasoni D, Bai F. Castoldi R. Barbanotti D. Falcinella C. Mule G. Mondatore D. Tavelli A. Vegni E. Marchetti G. d'Arminio Monforte A. | 2020 | Journal of medical virology 10.1002/jmv.26459 |
| 169. Assessment and Characterization of Post-COVID-19 manifestations | Tolba M, Abo Omirah M. Hussein A. Saeed H. | 2020 | International journal of clinical practice e13746 10.1111/ijcp.13746 |
| 170. Bidirectional associations between COVID-19 and psychiatric disorder: retrospective cohort studies of 62 354 COVID-19 cases in the USA | Taquet, Maxime; Luciano, Sierra; Geddes, John R.; Harrison, Paul J. | 2021 | The lancet. Psychiatry 8, 2 130-140 https://dx.doi.org/10.1016/S2215-0366(20)30462-4 |
| 171. Characteristics, comorbidities, 30-day outcome and in-hospital mortality of patients hospitalised with COVID-19 in a Swiss area - a retrospective cohort study | Pellaud C, Grandmaison G. Pham Huu Thien H. P. Baumberger M. Carrel G. Ksouri H. Erard V. Chuard C. Hayoz D. Sridharan G. | 2020 | Swiss medical weekly 150 w20314 10.4414/smw.2020.20314 |
| 172. Characterization of Prolonged COVID-19 Symptoms in an Outpatient Telemedicine Clinic | Cellai M, O'Keefe J. B. | 2020 | Open forum infectious diseases 7, 10 ofaa420 10.1093/ofid/ofaa420 |
| 173. Clinical characteristics and outcomes of adult patients admitted with COVID-19 in East London: a retrospective cohort analysisc | Daryl, Cheng; Claire, Calderwood; Erik, Skyllberg; Adam, Ainley | | |
| 174. Clinical characteristics and short term outcomes after recovery from COVID-19 in patients with and without diabetes in Bangladesh | Akter F, Mannan A. Mehedi H. M. H. Rob M. A. Ahmed S. Salauddin A. Hossain M. S. Hasan M. M. | 2020 | Diabetes & metabolic syndrome 14, 6 2031-2038 10.1016/j.dsx.2020.10.016 |
| 175. Clinical Presentation of Coronavirus Disease 2019 (COVID-19) in Pregnant and Recently Pregnant People | Afshar Y, Gaw S. L. Flaherman V. J. Chambers B. D. Krakow D. Berghella V. Shamshirsaz A. A. Boatin A. A. Aldrovandi G. Greiner A. Riley L. Boscardin W. J. Jamieson D. J. Jacoby V. L. | 2020 | Obstetrics and gynecology 10.1097/AOG.0000000000004178 |
| 176. Epidemiological and Clinical Findings of Short-Term Recurrence of Severe Acute Respiratory Syndrome Coronavirus 2 Ribonucleic Acid Polymerase Chain Reaction Positivity in 1282 Discharged Coronavirus Disease 2019 Cases: a Multicenter, Retrospective, Observ | Chen Sl, Xu H. Feng H. Y. Sun J. F. Li X. Zhou L. Song W. L. Huang S. S. He J. L. Deng Y. Y. Wang R. J. Fang M. | 2020 | Open forum infectious diseases 7, 10 ofaa432 10.1093/ofid/ofaa432 |
| 177. Epidemiology and clinical outcome of COVID-19: a multi-centre cross sectional study from Bangladesh (preprint) | Mannan A, Mehedi H. M. H. Chy N. H. Qayum M. O. Akter F. Rob A. Biswas P. Hossain S. Ayub M. I. | 2020 | Medrxiv 2020.09.09.20191114 10.1101/2020.09.09.20191114 |
| 178. Evolution of Altered Sense of Smell or Taste in Patients With Mildly Symptomatic COVID-19 | Boscolo-Rizzo P, Borsetto D. Fabbris C. Spinato G. Frezza D. Menegaldo A. Mularoni F. Gaudioso P. Cazzador D. Marciani S. Frasconi S. Ferraro M. Berro C. Varago C. Nicolai P. Tirelli G. Da Mosto M. C. Obholzer R. Rigoli R. Polesel J. Hopkins C. | 2020 | JAMA otolaryngology-- head & neck surgery 10.1001/jamaoto.2020.1379 |

| | | | |
|---|---|---|---|
| 179. Fibrotic Changes Depicted by Thin-Section CT in Patients With COVID-19 at the Early Recovery Stage: Preliminary Experience | Yang, Zhen Lu; Chen, Chong; Huang, Lu; Zhou, Shu Chang; Hu, Yu Na; Xia, Li Ming; Li, Yan | 2020 | Frontiers in medicine 7 605088 https://dx.doi.org/10.3389/fmed.2020.605088 |
| 180. Follow-up of adults with non-critical COVID-19 two months after symptoms' onset | Carvalho-Schneider C, Laurent E. Lemaignen A. Beaufils E. Bourbao-Tournois C. Laribi S. Flament T. Ferreira-Maldent N. Bruyere F. Stefic K. Gaudy-Graffin C. Grammatico-Guillon L. Bernard L. | 2020 | Clinical microbiology and infection 10.1016/j.cmi.2020.09.052 |
| 181. Functional and cognitive outcomes after COVID-19 delirium | McLoughlin, Benjamin C.; Miles, Amy; Webb, Thomas E.; Knopp, Paul; Eyres, Clodagh; Fabbri, Ambra; Humphries, Fiona; Davis, Daniel | 2020 | European geriatric medicine 11, 5 857-862 https://dx.doi.org/10.1007/s41999-020-00353-8 |
| 182. Functional characteristics of patients with SARS-CoV-2 pneumonia at 30 days post infection | Frija-Masson J, Debray M. P. Gilbert M. Lescure F. X. Travert F. Borie R. Khalil A. Crestani B. d'Ortho M. P. Bancal C. | 2020 | The european respiratory journal 10.1183/13993003.01754-2020 |
| 183. Headache: a striking prodromal and persistent symptom, predictive of COVID-19 clinical evolution | Caronna E, Ballve A. Llaurado A. Gallardo V. J. Maria Ariton D. Lallana S. Maza S. L. Gadea M. O. Quibus L. Restrepo J. L. Rodrigo-Gisbert M. Vilaseca A. Gonzalez M. H. Gallo M. M. Alpuente A. Torres-Ferrus M. Borrell R. P. Alvarez-Sabin J. Pozo-Rosich P. | 2020 | Cephalalgia 40, 13 1410-1421 10.1177/0333102420965157 |
| 184. Is COVID-19 Associated With Posttraumatic Stress Disorder? | Horn, Mathilde; Wathelet, Marielle; Fovet, Thomas; Amad, Ali; Vuotto, Fanny; Faure, Karine; Astier, Thibault; Noel, Helene; Duhem, Margot Henry Stephane; Vaiva, Guillaume; D'Hondt, Fabien | 2020 | The Journal of clinical psychiatry 82, 1 https://dx.doi.org/10.4088/JCP.20m13641 |
| 185. Long COVID in the skin: a registry analysis of COVID-19 dermatological duration | McMahon De, Gallman A. E. Hruza G. J. Rosenbach M. Lipoff J. B. Desai S. R. French L. E. Lim H. Cyster J. G. Fox L. P. Fassett M. S. Freeman E. E. | 2021 | The lancet. Infectious diseases 10.1016/S1473-3099(20)30986-5 |
| 186. 'Long-COVID': a cross-sectional study of persisting symptoms, biomarker and imaging abnormalities following hospitalisation for COVID-19 | Mandal S, Barnett J. Brill S. E. Brown J. S. Denneny E. K. Hare S. S. Heightman M. Hillman T. E. Jacob J. Jarvis H. C. Lipman M. C. I. Naidu S. B. Nair A. Porter J. C. Tomlinson G. S. Hurst J. R. A. R. C. Study Group | 2020 | Thorax 10.1136/thoraxjnl-2020-215818 |
| 187. Longitudinal symptom dynamics of COVID-19 infection | Mizrahi, Barak; Shilo, Smadar; Rossman, Hagai; Kalkstein, Nir; Marcus, Karni; Barer, Yael; Keshet, Ayya; Shamir-Stein, Na'ama; Shalev, Varda; Zohar, Anat Ekka; Chodick, Gabriel; Segal, Eran | 2020 | Nature communications 11, 1 6208 https://dx.doi.org/10.1038/s41467-020-20053-y |
| 188. Medium Term Follow-Up of 337 Patients With Coronavirus Disease 2019 (COVID-19) in a Fangcang Shelter Hospital in Wuhan, China | Yan N, Wang W. Gao Y. Zhou J. Ye J. Xu Z. Cao J. Zhang J. | 2020 | Frontiers in medicine 7 373 10.3389/fmed.2020.00373 |
| 189. Olfactory and gustatory dysfunctions in 100 patients hospitalized for COVID-19: sex differences and recovery time in real-life | Meini S, Suardi L. R. Busoni M. Roberts A. T. Fortini A. | 2020 | European archives of oto-rhino-laryngology 10.1007/s00405-020-06102-8 |
| 190. Olfactory and Gustatory Outcomes in COVID-19: a Prospective Evaluation in Nonhospitalized Subjects | Paderno A, Mattavelli D. Rampinelli V. Grammatica A. Raffetti E. Tomasoni M. Gualtieri T. Taboni S. Zorzi S. Del Bon F. Lombardi D. Deganello A. Redaelli De Zinis L. O. Schreiber A. | 2020 | Otolaryngology--head and neck surgery 10.1177/0194599820939538 |
| 191. Organizing pneumonia of COVID-19: Time-dependent evolution and outcome in CT findings | Wang, Yan; Jin, Chao; Wu, Carol C.; Zhao, Huifang; Liang, Ting; Liu, Zhe; Jian, Zhijie; Li, Runqing; Wang, Zekun; Li, Fen; Zhou, Jie; Cai, Shubo; Liu, Yang; Li, Hao; Liang, Yukun; Tian, Cong; Yang, Jian | 2020 | PloS one 15, 11 e0240347 https://dx.doi.org/10.1371/journal.pone.0240347 |
| 192. Patterns of smell recovery in 751 patients affected by the COVID-19 outbreak | Chiesa-Estomba Cm, Lechien J. R. Radulesco T. Michel J. Sowerby L. J. Hopkins C. Saussez S. | 2020 | European journal of neurology 10.1111/ene.14440 |
| 193. Persistence of COVID-19 Symptoms after Recovery in Mexican Population | Galvan-Tejada, Carlos E.; Herrera-Garcia, Cintya Fabiola; Godina-Gonzalez, Susana; Villagrana-Banuelos, Karen E.; Amaro, Juan Daniel De Luna; Herrera-Garcia, Karla; Rodriguez-Quinones, Carolina; Zanella-Calzada, Laura A.; Ramirez-Barranco, Julio; Avila, Jocelyn L. Ruiz de; Reyes-Escobedo, Fuensanta; Celaya-Padilla, Jose M.; | 2020 | International journal of environmental research and public health 17, 24 https://dx.doi.org/10.3390/ijerph17249367 |

| | | | | |
|---|---|---|---|---|
| | | Galvan-Tejada, Jorge I.; Gamboa-Rosales, Hamurabi; Martinez-Acuna, Monica; Cervantes-Villagrana, Alberto; Rivas-Santiago, Bruno; Gonzalez-Curiel, Irma E. | | |
| 194. | Persistence of symptoms and quality of life at 35 days after hospitalization for COVID-19 infection | Jacobs, Laurie G.; Gourna Paleoudis, Elli; Lesky-Di Bari, Dineen; Nyirenda, Themba; Friedman, Tamara; Gupta, Anjali; Rasouli, Lily; Zetkulic, Marygrace; Balani, Bindu; Ogedegbe, Chinwe; Bawa, Harinder; Berrol, Lauren; Qureshi, Nabiha; Aschner, Judy L. | 2020 | PloS one 15, 12 e0243882 https://dx.doi.org/10.1371/journal.pone.0243882 |
| 195. | Persistent Post-COVID-19 Inflammatory Interstitial Lung Disease: an Observational Study of Corticosteroid Treatment | Myall Kj, Mukherjee B. Castanheira A. M. Lam J. L. Benedetti G. Mak S. M. Preston R. Thillai M. Dewar A. Molyneaux P. L. West A. G. | 2021 | Annals of the American Thoracic Society 10.1513/AnnalsATS.202008-1002OC |
| 196. | Persistent Symptoms in Patients After Acute COVID-19 | Carfi, Angelo; Bernabei, Roberto; Landi, Francesco; Gemelli Against, Covid-Post-Acute Care Study Group | 2020 | JAMA 324, 6 603-605 https://dx.doi.org/10.1001/jama.2020.12603 |
| 197. | Post-COVID-19 Functional Status: relation to age, smoking, hospitalization and comorbidities (preprint) | Mohamed-Hussein A, Galal I. Saad M. Zayan H. E. Abdelsayed M. Moustafa M. Ezzat A. R. Helmy R. Abd Elaal H. Aly K. Abderheem S. | 2020 | Medrxiv 2020.08.26.20182618 10.1101/2020.08.26.20182618 |
| 198. | Post-discharge health status and symptoms in patients with severe COVID-19 | Weerahandi, Himali; Hochman, Katherine A.; Simon, Emma; Blaum, Caroline; Chodosh, Joshua; Duan, Emily; Garry, Kira; Kahan, Tamara; Karmen-Tuohy, Savannah; Karpel, Hannah; Mendoza, Felicia; Prete, Alexander M.; Quintana, Lindsey; Rutishauser, Jennifer; Santos Martinez, Leticia; Shah, Kanan; Sharma, Sneha; Simon, Elias; Stirniman, Ana; Horwitz, Leora | 2020 | medRxiv : the preprint server for health sciences https://dx.doi.org/10.1101/2020.08.11.2017274 2 |
| 199. | Post-Discharge Health Status and Symptoms in Patients with Severe COVID-19 | Weerahandi H, Hochman K. A. Simon E. Blaum C. Chodosh J. Duan E. Garry K. Kahan T. Karmen-Tuohy S. L. Karpel H. C. Mendoza F. Prete A. M. Quintana L. Rutishauser J. Santos Martinez L. Shah K. Sharma S. Simon E. Stirniman A. Z. Horwitz L. I. | 2021 | Journal of general internal medicine 10.1007/s11606-020-06338-4 |
| 200. | Post-discharge symptoms and rehabilitation needs in survivors of COVID-19 infection: a cross-sectional evaluation | Halpin Sj, McIvor C. Whyatt G. Adams A. Harvey O. McLean L. Walshaw C. Kemp S. Corrado J. Singh R. Collins T. O'Connor R. J. Sivan M. | 2020 | Journal of medical virology 10.1002/jmv.26368 |
| 201. | Postdischarge symptoms and rehabilitation needs in survivors of COVID-19 infection: A cross-sectional evaluation | Halpin, Stephen J.; McIvor, Claire; Whyatt, Gemma; Adams, Anastasia; Harvey, Olivia; McLean, Lyndsay; Walshaw, Christopher; Kemp, Steven; Corrado, Joanna; Singh, Rajinder; Collins, Tamsin; O'Connor, Rory J.; Sivan, Manoj | 2021 | Journal of medical virology 93, 2 1013-1022 https://dx.doi.org/10.1002/jmv.26368 |
| 202. | Predictors of Health-Related Quality of Life and Influencing Factors for COVID-19 Patients, a Follow-Up at One Month | Chen, Ke-Yang; Li, Ting; Gong, Fang-Hua; Zhang, Jin-San; Li, Xiao-Kun | 2020 | Frontiers in psychiatry 11 668 https://dx.doi.org/10.3389/fpsyt.2020.00668 |
| 203. | Prevalence and recovery time of olfactory and gustatory dysfunction in hospitalized patients with COVID-19 in Wuhan, China | Lv, Hao; Zhang, Wei; Zhu, Zhanyong; Xiong, Qiutang; Xiang, Rong; Wang, Yingying; Shi, Wendan; Deng, Zhifeng; Xu, Yu | 2020 | International journal of infectious diseases : 100 507-512 https://dx.doi.org/10.1016/j.ijid.2020.09.039 |
| 204. | Prevalence and Reversibility of Smell Dysfunction Measured Psychophysically in a Cohort of COVID-19 patients | Moein St, Hashemian S. M. R. Tabarsi P. Doty R. L. | 2020 | International forum of allergy & rhinology 10.1002/alr.22680 |
| 205. | Prolonged and Late-Onset Symptoms of Coronavirus Disease 2019 | Miyazato, Yusuke; Morioka, Shinichiro; Tsuzuki, Shinya; Akashi, Masako; Osanai, Yasuyo; Tanaka, Keiko; Terada, Mari; Suzuki, Michiyo; Kutsuna, Satoshi; Saito, Sho; Hayakawa, Kayoko; Ohmagari, Norio | 2020 | Open forum infectious diseases 7, 11 ofaa507 https://dx.doi.org/10.1093/ofid/ofaa507 |
| 206. | Prolonged complaints of chemosensory loss after COVID-19 | Fjaeldstad, Alexander Wieck | 2020 | Danish medical journal 67, 8 |

| | | | |
|---|---|---|---|
| 207. Psychiatric morbidity and protracted symptoms after COVID-19 | Poyraz, Burc Cagri; Poyraz, Cana Aksoy; Olgun, Yesim; Gurel, Ozge; Alkan, Sena; Ozdemir, Yusuf Emre; Balkan, Ilker Inanc; Karaali, Ridvan | 2021 | Psychiatry research 295 113604 https://dx.doi.org/10.1016/j.psychres.2020.113604 |
| 208. Pulmonary function and health-related quality of life after COVID-19 pneumonia | van der Sar-van der Brugge S, Talman S. Boonman-de Winter L. de Mol M. Hoefman E. van Etten R. W. De Backer I. C. | 2020 | Respiratory medicine 176 106272 10.1016/j.rmed.2020.106272 |
| 209. Reduced maximal aerobic capacity after COVID-19 in young adult recruits, Switzerland, May 2020 | Crameri, Giovanni Andrea Gerardo; Bielecki, Michel; Zust, Roland; Buehrer, Thomas Werner; Stanga, Zeno; Deuel, Jeremy Werner | 2020 | Euro surveillance : bulletin Europeen sur les maladies transmissibles = European communicable disease bulletin 25, 36 https://dx.doi.org/10.2807/1560-7917.ES.2020.25.36.2001542 |
| 210. Remdesivir for the Treatment of Covid-19 - Preliminary Report | Beigel Jh, Tomashek K. M. Dodd L. E. Mehta A. K. Zingman B. S. Kalil A. C. Hohmann E. Chu H. Y. Luetkemeyer A. Kline S. Lopez de Castilla D. Finberg R. W. Dierberg K. Tapson V. Hsieh L. Patterson T. F. Paredes R. Sweeney D. A. Short W. R. Touloumi G. Lye D. C. Ohmagari N. Oh M. D. Ruiz-Palacios G. M. Benfield T. Fatkenheuer G. Kortepeter M. G. Atmar R. L. Creech C. B. Lundgren J. Babiker A. G. Pett S. Neaton J. D. Burgess T. H. Bonnett T. Green M. Makowski M. Osinusi A. Nayak S. Lane H. C. | 2020 | New England journal of medicine 10.1056/NEJMoa2007764 |
| 211. Residual clinical damage after COVID-19: a retrospective and prospective observational cohort study | De Lorenzo R, Conte C. Lanzani C. Benedetti F. Roveri L. Mazza M. G. Brioni E. Giacalone G. Canti V. Sofia V. D'Amico M. Di Napoli D. Ambrosio A. Scarpellini P. Castagna A. Landoni G. Zangrillo A. Bosi E. Tresoldi M. Ciceri F. Rovere-Querini P. | 2020 | PloS one 15, 10 e0239570 10.1371/journal.pone.0239570 |
| 212. Risk factors associated with mental illness in hospital discharged patients infected with COVID-19 in Wuhan, China | Liu D, Baumeister R. F. Veilleux J. C. Chen C. Liu W. Yue Y. Zhang S. | 2020 | Psychiatry research 292 113297 10.1016/j.psychres.2020.113297 |
| 213. Self-reported taste and smell disorders in patients with COVID-19: distinct features in China | Liu Js, Yi-Ke D. Hai W. Zhi-Chao W. Bo L. Jin M. Chao H. Li P. Yang L. Isam A. De-Yun W. Ming Z. Joaquim M. Zheng | 2020 | Medrxiv 10.1101/2020.06.12.20128298 |
| 214. Short-term outpatient follow-up of COVID-19 patients: A multidisciplinary approach | de Graaf, M. A.; Antoni, M. L.; ter Kuile, M. M.; Arbous, M. S.; Duinisveld, A. J. F.; Feltkamp, M. C. W.; Groeneveld, G. H.; Hinnen, S. C. H.; Janssen, V. R.; Lijfering, W. M.; Omara, S.; Postmus, P. E.; Ramai, S. R. S.; Rius-Ottenheim, N.; Schalij, M. J.; Schiemanck, S. K.; Smid, L.; Stoger, J. L.; Visser, L. G.; de Vries, J. J. C.; Wijngaarden, M. A.; Geelhoed, J. J. M.; Roukens, A. H. E. | 2021 | EClinicalMedicine 100731 http://dx.doi.org/10.1016/j.eclinm.2021.100731 |
| 215. Smell and taste recovery in coronavirus disease 2019 patients: a 60-day objective and prospective study | Vaira, L. A.; Hopkins, C.; Petrocelli, M.; Lechien, J. R.; Chiesa-Estomba, C. M.; Salzano, G.; Cucurullo, M.; Salzano, F. A.; Saussez, S.; Boscolo-Rizzo, P.; Biglioli, F.; De Riu, G. | 2020 | The Journal of laryngology and otology 134, 8 703-709 https://dx.doi.org/10.1017/S0022215120001826 |
| 216. Subjective smell and taste changes during the COVID-19 pandemic: short term recovery | Reiter Er, Coelho D. H. Kons Z. A. Costanzo R. M. | 2020 | American journal of otolaryngology 41, 6 102639 10.1016/j.amjoto.2020.102639 |
| 217. Symptom Course in COVID-19 Outpatients | Keefe Jb, Tong D. C. Datoo O. Keefe G. A. | 2020 | Medrxiv 2020.06.05.20123471 10.1101/2020.06.05.20123471 |
| 218. Symptom Duration and Risk Factors for Delayed Return to Usual Health Among Outpatients with COVID-19 in a Multistate Health Care Systems Network - United States, March-June 2020 | Tenforde, Mark W.; Kim, Sara S.; Lindsell, Christopher J.; Billig Rose, Erica; Shapiro, Nathan I.; Files, D. Clark; Gibbs, Kevin W.; Erickson, Heidi L.; Steingrub, Jay S.; Smithline, Howard A.; Gong, Michelle N.; Aboodi, Michael S.; Exline, Matthew C.; Henning, Daniel J.; Wilson, Jennifer G.; Khan, Akram; Qadir, Nida; Brown, Samuel M.; Peltan, Ithan D.; Rice, Todd W.; Hager, David N.; Ginde, Adit A.; Stubblefield, | 2020 | MMWR. Morbidity and mortality weekly report 69, 30 993-998 https://dx.doi.org/10.15585/mmwr.mm6930e1 |

| | | | |
|---|---|---|---|
| | William B.; Patel, Manish M.; Self, Wesley H.; Feldstein, Leora R.; Investigators, I. V. Y. Network; Team, Cdc Covid- Response | | |
| 219. The Clinical Features and Outcomes of Discharged Coronavirus Disease 2019 Patients:A Prospective Cohort Study | Wang X, Xu H. Jiang H. Wang L. Lu C. Wei X. Liu J. Xu S. | 2020 | QJM : monthly journal of the Association of Physicians 10.1093/qjmed/hcaa178 |
| 220. The correlation between BMI and COVID-19 outcomes | Khasawneh, L.; Al-Omar, K.; Tarifi, A. A.; Shaout, D. M. K.; Abu-Ghazal, S. Y. A.; Alfazza, A. T. S. M. | 2020 | Systematic Reviews in Pharmacy 11, 6 1236-1239 http://dx.doi.org/10.31838/srp.2020.6.180 |
| 221. The Kids Are Not Alright: A Preliminary Report of Post-COVID Syndrome in University Students | Walsh-Messinger, Julie; Manis, Hannah; Vrabec, Alison; Sizemore, Jenna; Bishof, Karyn; Debidda, Marcella; Malaspina, Dolores; Greenspan, Noah | 2020 | medRxiv : the preprint server for health sciences https://dx.doi.org/10.1101/2020.11.24.20238261 |
| 222. The safety of home discharge for low-risk emergency department patients presenting with coronavirus-like symptoms during the COVID-19 pandemic: a retrospective cohort study | Berdahl Ct, Glennon N. C. Henreid A. J. Torbati S. S. | 2020 | Journal of the american college of emergency physicians open 10.1002/emp2.12230 |
| 223. Treatment, Persistent Symptoms, and Depression in People Infected with COVID-19 in Bangladesh | Islam, Md Saiful; Ferdous, Most Zannatul; Islam, Ummay Soumayia; Mosaddek, Abu Syed Md; Potenza, Marc N.; Pardhan, Shahina | 2021 | International journal of environmental research and public health 18, 4 https://dx.doi.org/10.3390/ijerph18041453 |
| 224. Comparison of clinical, haematological, biochemical findings and significance of co-morbidities amongst COVID-19 positive survivors and nonsurvivors | Bharat, V.; Singhal, M.; Varma, A.; Jindal, S. | 2021 | Journal of Clinical and Diagnostic Research 15(4) EC22-EC26 http://dx.doi.org/10.7860/JCDR/2021/47148.14811 |
| 225. Evolving Phenotypes of non-hospitalized Patients that Indicate Long Covid | Estiri, Hossein; Strasser, Zachary H.; Brat, Gabriel A.; Semenov, Yevgeniy R.; Consortium for Characterization of, Covid-by E. H. R.; Patel, Chirag J.; Murphy, Shawn N. | 2021 | medRxiv : the preprint server for health sciences https://dx.doi.org/10.1101/2021.04.25.21255923 |
| 226. Cognitive deficits in people who have recovered from COVID-19 relative to controls: an N=84,285 online study (preprint) | Hampshire A, Trender W. Chamberlain S. Jolly A. Grant J. E. Patrick F. Mazibuko N. Williams S. Barnby J. M. Hellyer P. Mehta M. A. | 2020 | Medrxiv 2020.10.20.20215863 10.1101/2020.10.20.20215863 |
| 227. Cognitive deficits in people who have recovered from COVID-19 | Hampshire, A.; Trender, W.; Chamberlain, S. R.; Jolly, A. E.; Grant, J. E.; Patrick, F.; Mazibuko, N.; Williams, S. C.; Barnby, J. M.; Hellyer, P.; Mehta, M. A. | 2021 | EClinicalMedicine 39 101044 https://dx.doi.org/10.1016/j.eclinm.2021.101044 |
| 228. Post-acute sequelae of COVID-19 in a non-hospitalized cohort: results from the Arizona CoVHORT (preprint) | Bell Ml, Catalfamo C. J. Farland L. V. Ernst K. C. Jacobs E. T. Klimentidis Y. C. Jehn M. Pogreba-Brown K. | 2021 | Medrxiv 2021.03.29.21254588 10.1101/2021.03.29.21254588 |
| 229. Clinical characteristics and outcomes of inpatients with neurologic disease and COVID-19 in Brescia, Lombardy, Italy | Benussi A, Pilotto A. Premi E. Libri I. Giunta M. Agosti C. Alberici A. Baldelli E. Benini M. Bonacina S. Brambilla L. Caratozzolo S. Cortinovis M. Costa A. Piccinelli S. C. Cottini E. Cristillo V. Delrio I. Filosto M. Gamba M. Gazzina S. Gilberti N. Gipponi S. Imarisio A. Invernizzi P. Leggio U. Leonardi M. Liberini P. Locatelli M. Masciocchi S. Poli L. Rao R. Risi B. Rozzini L. Scalvini A. Schiano di Cola F. Spezi R. Vergani V. Volonghi I. Zoppi N. Borroni B. Magoni M. Pezzini A. Padovani A. | 2020 | Neurology 10.1212/WNL.0000000000009848 |

| | | | | |
|---|---|---|---|---|
| 230. | COVID-19 outpatient management: shorter time to recovery in Healthcare workers according to an electronic daily symptoms assessment | Breugnon E, Thollot H. Fraissenon A. Saunier F. Labetoulle R. Pillet S. Lucht F. Berthelot P. Botelho-Nevers E. Gagneux-Brunon A. | 2020 | Medecine et maladies infectieuses 10.1016/j.medmal.2020.10.001 |
| **Wrong outcomes (n=77)** | | | | |
| 231. | Mortality after surgery with SARS-CoV-2 infection in England: a population-wide epidemiological study | Abbott, T. E. F.; Fowler, A. J.; Dobbs, T. D.; Gibson, J.; Shahid, T.; Dias, P.; Akbari, A.; Whitaker, I. S.; Pearse, R. M. | 2021 | British Journal of Anaesthesia 127, 2 205-214 https://dx.doi.org/10.1016/j.bja.2021.05.018 |
| 232. | Characterizing "long-COVID" using real world data: post-discharge clinical course among patients initially hospitalized for COVID-19 | Eldridge E, Corbett E. Jones J. Mahmood S. Lin N. D. | 2021 | Pharmacoepidemiology and drug safety 30, SUPPL 1 165- 10.1002/pds.5305 |
| 233. | 30-Day Readmission Rate Of Covid-19 Patients Discharged From A Tertiary Care University Hospital In Turkey; An Observational, Single-Center Study | Uyaroglu Oa, Basaran N. C. Ozisik L. Dizman G. T. Eroglu I. Sahin T. K. Tas Z. Inkaya A. C. Tanriover M. D. Metan G. Guven G. S. Unal S. | 2020 | International journal for quality in health care : journal of the international society for quality in health care 10.1093/intqhc/mzaa144 |
| 234. | Thyroid function abnormalities in COVID-19 patients | Wang W, Su X. Ding Y. Fan W. Su J. Chen Z. Zhao H. Xu K. Ni Q. Xu X. Qiu Y. Teng L. | 2020 | Medrxiv 2020.06.15.20130807 10.1101/2020.06.15.20130807 |
| 235. | Characteristics and outcomes of asthmatic patients with COVID-19 pneumonia who require hospitalisation | Beurnier A, Jutant E. M. Jevnikar M. Boucly A. Pichon J. Preda M. Frank M. Laurent J. Richard C. Monnet X. Duranteau J. Harrois A. Chaumais M. C. Bellin M. F. Noel N. Bulifon S. Jais X. Parent F. Seferian A. Savale L. Sitbon O. Montani D. Humbert M. | 2020 | The european respiratory journal 10.1183/13993003.01875-2020 |
| 236. | Risk Factors Associated With Long-Term Hospitalization in Patients With COVID-19: A Single-Centered, Retrospective Study | Wu, Yiqun; Hou, Bingbo; Liu, Jielan; Chen, Yingying; Zhong, Ping | 2020 | Frontiers in medicine 7 315 https://dx.doi.org/10.3389/fmed.2020.00315 |
| 237. | Compare the epidemiological and clinical features of imported and local COVID-19 cases in Hainan, China | Wu B, Lei Z. Y. Wu K. L. He J. R. Cao H. J. Fu J. Chen F. Chen Y. Chen B. Zhou X. L. Huang T. Wu T. Du Y. G. Chen S. X. Xiao F. R. Gao Z. L. He J. Lin F. Lin B. L. | 2020 | Infectious diseases of poverty 9, 1 143 10.1186/s40249-020-00755-7 |
| 238. | Tocilizumab in patients with severe COVID-19: a single-center observational analysis | Knorr Jp, Colomy V. Mauriello C. M. Ha S. | 2020 | Journal of medical virology 10.1002/jmv.26191 |
| 239. | Clinical characteristics of coronavirus disease 2019 in Gansu province, China | Yue H, Bai X. Wang J. Yu Q. Liu W. Pu J. Wang X. Hu J. Xu D. Li X. Kang N. Li L. Lu W. Feng T. Ding L. Li X. Qi X. Gansu Provincial Medical Treatment Expert Group of Covid | 2020 | Annals of palliative medicine 10.21037/apm-20-887 |
| 240. | Presenting characteristics, smoking versus diabetes and outcome among patients hospitalized with COVID-19 | Abbas Hm, Nassir K. F. Al Khames Aga Q. A. Al-Gharawi A. A. Rasheed J. I. Al-Obaidy M. W. Al Jubouri A. M. Jaber A. S. Al Khames Aga L. A. | 2020 | Journal of medical virology 10.1002/jmv.26487 |
| 241. | Association between Initial Chest CT or Clinical Features and Clinical Course in Patients with Coronavirus Disease 2019 Pneumonia | Liu Z, Jin C. Wu C. C. Liang T. Zhao H. Wang Y. Wang Z. Li F. Zhou J. Cai S. Zeng L. Yang J. | 2020 | Korean journal of radiology 21, 6 736-745 10.3348/kjr.2020.0171 |
| 242. | Efficacy of corticosteroids in non-intensive care unit patients with COVID-19 pneumonia from the New York Metropolitan region | Majmundar M, Kansara T. Lenik J. M. Park H. Ghosh K. Doshi R. Shah P. Kumar A. Amin H. Chaudhari S. Habtes I. | 2020 | PloS one 15, 9 e0238827 10.1371/journal.pone.0238827 |
| 243. | Impact of comorbidities on clinical prognosis in 1280 patients with different types of COVID-19 | Fang H, Liu Q. Xi M. Xiong D. He J. Luo P. Li Z. | 2020 | Journal of investigative medicine 10.1136/jim-2020-001555 |
| 244. | Death, discharge and arrhythmias among patients with COVID-19 and cardiac injury | Si D, Du B. Ni L. Yang B. Sun H. Jiang N. Liu G. Masse S. Jin L. Nanthakumar J. Bhaskaran A. Yang P. Nanthakumar K. | 2020 | CMAJ : Canadian Medical Association journal 10.1503/cmaj.200879 |
| 245. | An integrated multidisciplinary model of COVID-19 recovery care | O'Brien H, Tracey M. J. Ottewill C. O'Brien M. E. Morgan R. K. Costello R. W. Gunaratnam C. Ryan D. McElvaney N. G. McConkey S. | 2020 | Irish journal of medical science 10.1007/s11845-020-02354-9 |

| | | | |
|---|---|---|---|
| | J. McNally C. Curley G. F. MacHale S. Gillan D. Pender N. Barry H. de Barra E. Kiernan F. M. Sulaiman I. Hurley K. | | |
| 246. Clinical outcomes of COVID-19 in Wuhan, China: a large cohort study | Liu J, Zhang S. Wu Z. Shang Y. Dong X. Li G. Zhang L. Chen Y. Ye X. Du H. Liu Y. Wang T. Huang S. Chen L. Wen Z. Qu J. Chen D. | 2020 | Annals of intensive care 10, 1 99 10.1186/s13613-020-00706-3 |
| 247. Associations of medications used during hospitalization and immunological changes in patients with COVID-19 during 3-month follow-up | Liu C, Dun Y. Liu P. You B. Shu K. Luo H. Ripley-Gonzalez J. W. Liu S. Liu J. Li B. | 2020 | International immunopharmacology 107121 10.1016/j.intimp.2020.107121 |
| 248. Temporal radiographic changes in COVID-19 patients: relationship to disease severity and viral clearance | Liu X, Zhou H. Zhou Y. Wu X. Zhao Y. Lu Y. Tan W. Yuan M. Ding X. Zou J. Li R. Liu H. Ewing R. M. Hu Y. Nie H. Wang Y. | 2020 | Scientific reports 10, 1 10263 10.1038/s41598-020-66895-w |
| 249. Nervous System Involvement in COVID-19: results from a Retrospective Consecutive Neuroimaging Cohort | Klironomos S, Tzortzakakis A. Kits A. Ohberg C. Kollia E. Ahoromazdae A. Almqvist H. Aspelin A. Martin H. Ouellette R. Al-Saadi J. Hasselberg M. Haghgou M. Pedersen M. Petersson S. Finnsson J. Lundberg J. Falk Delgado A. Granberg T. | 2020 | Radiology 202791 10.1148/radiol.2020202791 |
| 250. Age, Frailty, and Comorbidity as Prognostic Factors for Short-Term Outcomes in Patients With Coronavirus Disease 2019 in Geriatric Care | Hagg S, Jylhava J. Wang Y. Xu H. Metzner C. Annetorp M. Garcia-Ptacek S. Khedri M. Bostrom A. M. Kadir A. Johansson A. Kivipelto M. Eriksdotter M. Cederholm T. Religa D. | 2020 | Journal of the American Medical Directors Association 10.1016/j.jamda.2020.08.014 |
| 251. Impact of COVID-19 on liver function: results from an internal medicine unit in Northern Italy | Lenti Mv, Borrelli de Andreis F. Pellegrino I. Klersy C. Merli S. Miceli E. Aronico N. Mengoli C. Di Stefano M. Cococcia S. Santacroce G. Soriano S. Melazzini F. Delliponti M. Baldanti F. Triarico A. Corazza G. R. Pinzani M. Di Sabatino A. Internal Medicine Covid-Team | 2020 | Internal and emergency medicine 10.1007/s11739-020-02425-w |
| 252. Is there a correlation between pulmonary inflammation index with COVID-19 disease severity and outcome? (preprint) | Mohamed-Hussein A, Galal I. Mohamed Mmar Abd Elaal H. Aly K. M. E. | 2020 | Medrxiv 2020.09.09.20182592 10.1101/2020.09.09.20182592 |
| 253. Factors Affecting COVID-19 Outcomes in Cancer Patients: a First Report From Guy's Cancer Center in London | Russell B, Moss C. Papa S. Irshad S. Ross P. Spicer J. Kordasti S. Crawley D. Wylie H. Cahill F. Haire A. Zaki K. Rahman F. Sita-Lumsden A. Josephs D. Enting D. Lei M. Ghosh S. Harrison C. Swampillai A. Sawyer E. D'Souza A. Gomberg S. Fields P. Wrench D. Raj K. Gleeson M. Bailey K. Dillon R. Streetly M. Rigg A. Sullivan R. Dolly S. Van Hemelrijck M. | 2020 | Frontiers in oncology 10 10.3389/fonc.2020.01279 |
| 254. Lopinavir-ritonavir versus hydroxychloroquine for viral clearance and clinical improvement in patients with mild to moderate coronavirus disease 2019 | Kim Jw, Kim E. J. Kwon H. H. Jung C. Y. Kim K. C. Choe J. Y. Hong H. L. | 2020 | The Korean journal of internal medicine 10.3904/kjim.2020.224 |
| 255. Clinical outcomes and adverse events in patients hospitalised with COVID -19, treated with off- label hydroxychloroquine and azithromycin | Kelly M, O'Connor R. Townsend L. Coghlan M. Relihan E. Moriarty M. Carr B. Melanophy G. Doyle C. Bannan C. O'Riordan R. Merry C. Clarke S. Bergin C. | 2020 | British journal of clinical pharmacology 10.1111/bcp.14482 |
| 256. Predicting individual risk for COVID19 complications using EMR data | Kinar Y, Lanyado A. Shoshan A. Yesharim R. Domany T. Shalev V. Chodcik G. | 2020 | Medrxiv 2020.06.03.20121574 10.1101/2020.06.03.20121574 |
| 257. App-based tracking of self-reported COVID-19 symptoms | Zens M, Brammertz A. Herpich J. Sudkamp N. Hinterseer M. | 2020 | Journal of medical Internet research 10.2196/21956 |
| 258. Post-discharge venous thromboembolism following hospital admission with COVID-19 | Roberts Ln, Whyte M. B. Georgiou L. Giron G. Czuprynska J. Rea C. Vadher B. Patel R. Gee E. Arya R. | 2020 | Blood 10.1182/blood.2020008086 |
| 259. Temporal lung changes in high-resolution chest computed tomography for coronavirus disease 2019 | Chen C, Wang X. Dong J. Nie D. Chen Q. Yang F. Chen W. Hu Q. | 2020 | Journal of international medical research 48, 9 10.1177/0300060520950990 |
| 260. The spectrum of biochemical alterations associated with organ dysfunction and inflammatory status and their | Oussalah, Abderrahim; Gleye, Stanislas; Urmes, Isabelle Clerc; Laugel, Elodie; Barbe, Francoise; Orlowski, Sophie; Malaplate, Catherine; Aimone-Gastin, Isabelle; Caillierez, Beatrice Maatem; | 2020 | EClinicalMedicine 27 100554 https://dx.doi.org/10.1016/j.eclinm.2020.100554 |

| | | | | |
|---|---|---|---|---|
| | association with disease outcomes in severe COVID-19: A longitudinal cohort and time-series design study | Merten, Marc; Jeannesson, Elise; Kormann, Raphael; Olivier, Jean-Luc; Rodriguez-Gueant, Rosa-Maria; Namour, Fares; Bevilacqua, Sybille; Thilly, Nathalie; Losser, Marie-Reine; Kimmoun, Antoine; Frimat, Luc; Levy, Bruno; Gibot, Sebastien; Schvoerer, Evelyne; Gueant, Jean-Louis | | |
| 261. | Disease Course and Outcomes of COVID-19 Among Hospitalized Patients with Gastrointestinal Manifestations | Laszkowska M, Faye A. S. Kim J. Truong H. Silver E. R. Ingram M. May B. Ascherman B. Bartram L. Zucker J. Sobieszczyk M. E. Abrams J. A. Lebwohl B. Freedberg D. E. Hur C. | 2020 | Clinical gastroenterology and hepatology 10.1016/j.cgh.2020.09.037 |
| 262. | A Controllable Inflammatory Response and Temporary Abnormal Coagulation in Moderate Disease of COVID-19 in Wuhan, China | Liu Y, Zhang X. Qiao J. Gong R. You Q. Sun J. Liu W. Sun B. | 2020 | Journal of clinical medicine research 12, 9 590-597 10.14740/jocmr4293 |
| 263. | Molecular and serological characterization of SARS-CoV-2 infection among COVID-19 patients | Li L, Liang Y. Hu F. Yan H. Li Y. Xie Z. Huang L. Zhao J. Wan Z. Wang H. Shui J. Cai W. Tang S. | 2020 | Virology 551 26-35 10.1016/j.virol.2020.09.008 |
| 264. | Coronavirus disease 2019 in elderly patients: Characteristics and prognostic factors based on 4-week follow-up | Wang, Lang; He, Wenbo; Yu, Xiaomei; Hu, Dalong; Bao, Mingwei; Liu, Huafen; Zhou, Jiali; Jiang, Hong | 2020 | The Journal of infection 80, 6 639-645 https://dx.doi.org/10.1016/j.jinf.2020.03.019 |
| 265. | Factors associated with the duration of hospitalisation among COVID-19 patients in Vietnam: A survival analysis | Thai, Pham Quang; Toan, Do Thi Thanh; Son, Dinh Thai; Van, Hoang Thi Hai; Minh, Luu Ngoc; Hung, Le Xuan; Toan, Ngo Van; Hoat, Luu Ngoc; Luong, Duong Huy; Khue, Luong Ngoc; Khoa, Nguyen Trong; Huong, Le Thi | 2020 | Epidemiology and infection 148 e114 https://dx.doi.org/10.1017/S0950268820001259 |
| 266. | Analysis of thin-section CT in patients with coronavirus disease (COVID-19) after hospital discharge | Wei, Jiangping; Yang, Hong; Lei, Pinggui; Fan, Bing; Qiu, Yingying; Zeng, Bingliang; Yu, Peng; Lv, Jian; Jian, Yinchao; Wan, Chengfeng | 2020 | Journal of X-ray science and technology 28, 3 383-389 https://dx.doi.org/10.3233/XST-200685 |
| 267. | The first consecutive 5000 patients with Coronavirus Disease 2019 from Qatar; a nation-wide cohort study | Omrani As, Almaslamani M. A. Daghfal J. Alattar R. A. Elgara M. Shaar S. H. Ibrahim T. B. H. Zaqout A. Bakdach D. Akkari A. M. Baiou A. Alhariri B. Elajez R. Husain A. A. M. Badawi M. N. Abid F. B. Abu Jarir S. H. Abdalla S. Kaleeckal A. Choda K. Chinta V. R. Sherbash M. A. Al-Ismail K. Abukhattab M. Ait Hssain A. Coyle P. V. Bertollini R. Frenneaux M. P. Alkhal A. Al-Kuwari H. M. | 2020 | BMC infectious diseases 20, 1 777 10.1186/s12879-020-05511-8 |
| 268. | Comparison of clinical characteristics of COVID-19 between elderly patients and young patients: a study based on a 28-day follow-up | Zhang L, Fan T. Yang S. Feng H. Hao B. Lu Z. Xiong R. Shen X. Jiang W. Wang W. Geng Q. | 2020 | Aging 12 10.18632/aging.104077 |
| 269. | Rate of venous thromboembolism in a prospective all-comers cohort with COVID-19 | Rieder M, Goller I. Jeserich M. Baldus N. Pollmeier L. Wirth L. Supady A. Bode C. Busch H. J. Schmid B. Duerschmied D. Gauchel N. Lother A. | 2020 | Journal of thrombosis and thrombolysis 10.1007/s11239-020-02202-8 |
| 270. | Clinical characteristics and prognosis of hospitalized COVID-19 patients with incident sustained tachyarrhythmias: a multicenter observational study | Russo V, Di Maio M. Mottola F. Pagnano G. Attena E. Verde N. Di Micco P. Silverio A. Scudiero F. Nunziata L. Fele N. D'Andrea A. Parodi G. Albani S. Scacciatella P. Nigro G. Severino S. | 2020 | European journal of clinical investigation e13387 10.1111/eci.13387 |
| 271. | Characteristics and Outcomes in Patients With COVID-19 and Acute Ischemic Stroke: the Global COVID-19 Stroke Registry | Ntaios G, Michel P. Georgiopoulos G. Guo Y. Li W. Xiong J. Calleja P. Ostos F. Gonzalez-Ortega G. Fuentes B. Alonso de Lecinana M. Diez-Tejedor E. Garcia-Madrona S. Masjuan J. DeFelipe A. Turc G. Goncalves B. Domigo V. Dan G. A. Vezeteu R. Christensen H. Christensen L. M. Meden P. Hajdarevic L. Rodriguez-Lopez A. Diaz-Otero F. Garcia-Pastor A. Gil-Nunez A. Maslias E. Strambo D. Werring D. J. Chandratheva A. Benjamin L. Simister R. Perry R. Beyrouti R. Jabbour P. Sweid A. Tjoumakaris S. Cuadrado-Godia E. Campello A. R. Roquer J. Moreira T. Mazya M. V. Bandini F. Matz K. | 2020 | Stroke; a journal of cerebral circulation STROKEAHA120031208 10.1161/STROKEAHA.120.031208 |

| | | | |
|---|---|---|---|
| | Iversen H. K. Gonzalez-Duarte A. Tiu C. Ferrari J. Vosko M. R. Salzer H. J. F. Lamprecht B. Dunser M. W. Cereda C. W. Quintero A. B. C. Korompoki E. Soriano-Navarro E. Soto-Ramirez L. E. Castaneda-Mendez P. F. Bay-Sansores D. Arauz A. Cano-Nigenda V. Kristoffersen E. S. Tiainen M. Strbian D. Putaala J. Lip G. Y. H. | | |
| 272. Covid-19 Demographics from a Tertiary Care Center: does It Depreciate Quality-of-Life? | Revathishree K, Shyam Sudhakar S. Indu R. Srinivasan K. | 2020 | Indian journal of otolaryngology and head and neck surgery 10.1007/s12070-020-02144-w |
| 273. Characteristics and Outcomes of 599 Patients Infected with COVID-19 in Wuhan, China: based on an Online Reported Sample | Liu D, Wang Y. Wang J. Liu J. Yue Y. Liu W. Wang Z. | 2020 | Journal of medical Internet research 10.2196/20108 |
| 274. Clinical features of COVID-19 convalescent patients with re-positive nucleic acid detection | Zhu, Hui; Fu, Liyun; Jin, Yinhua; Shao, Jiale; Zhang, Shun; Zheng, Nanhong; Fan, Lingyan; Yu, Zhe; Ying, Jun; Hu, Yaoren; Chen, Tongen; Chen, Yanglingzi; Chen, Min; Chen, Mingjue; Xiong, Zi; Kang, Junfei; Jin, Jiachang; Cai, Ting; Ye, Honghua | 2020 | Journal of clinical laboratory analysis 34, 7 e23392 https://dx.doi.org/10.1002/jcla.23392 |
| 275. COVID-19 Infection Among Healthcare Workers in a National Healthcare System: the Qatar Experience | Alajmi J, Jeremijenko A. M. Abraham J. C. Alishaq M. Concepcion E. G. Butt A. A. Abou-Samra A. B. | 2020 | International journal of infectious diseases 10.1016/j.ijid.2020.09.027 |
| 276. Clinical characteristics of recovered COVID-19 patients with re-detectable positive RNA test | An J, Liao X. Xiao T. Qian S. Yuan J. Ye H. Qi F. Shen C. Wang L. Liu Y. Cheng X. Li N. Cai Q. Wang F. Chen J. Li G. Cai Q. Liu Y. Wang Y. Zhang F. Fu Y. He Q. Tan X. Liu L. Zhang Z. | 2020 | Annals of translational medicine 8, 17 1084 10.21037/atm-20-5602 |
| 277. Health-care workers with COVID-19 living in Mexico City: clinical characterization and related outcomes | Antonio-Villa Ne, Bello-Chavolla O. Y. Vargas-Vazquez A. Fermin-Martinez C. A. Marquez-Salinas A. Bahena-Lopez J. P. | 2020 | Clinical infectious diseases 10.1093/cid/ciaa1487 |
| 278. COVID-19 and venous thromboembolism in intensive care or medical ward | Avruscio G, Camporese G. Campello E. Bernardi E. Persona P. Passarella C. Noventa F. Cola M. Navalesi P. Cattelan A. Tiberio I. Boscolo A. Spiezia L. Simioni P. Covid- V. T. E. study group | 2020 | Clinical and translational science 10.1111/cts.12907 |
| 279. Early Anti-SARS-CoV-2 Convalescent Plasma in Patients Admitted for COVID-19: a Randomized Phase II Clinical Trial (preprint) | Balcells Me, Rojas L. Le Corre N. Martinez-Valdebenito C. Ceballos M. E. Ferres M. Chang M. Vizcaya C. Mondaca S. Huete A. Castro R. Sarmiento M. Villarroel L. Pizarro A. Ross P. Santander J. Lara B. Ferrada M. Vargas-Salas S. Beltran-Pavez C. Soto-Rifo R. Valiente-Echeverria F. Caglevic C. Mahave M. Selman C. Gazitua R. Briones J. L. Villarroel-Espindola F. Balmaceda C. Espinoza M. A. Pereira J. Nervi B. | 2020 | Medrxiv 2020.09.17.20196212 10.1101/2020.09.17.20196212 |
| 280. Vitamin D and survival in COVID-19 patients: a quasi-experimental study | Annweiler C, Hanotte B. de l'Eprevier C. G. Sabatier J. M. Lafaie L. Celarier T. | 2020 | Journal of steroid biochemistry and molecular biology 105771 10.1016/j.jsbmb.2020.105771 |
| 281. Management and outcomes of post-acute COVID-19 patients in Northern Italy | Vitacca M, Migliori G. B. Spanevello A. Melazzini M. G. Ambrosino N. | 2020 | European journal of internal medicine 10.1016/j.ejim.2020.06.005 |
| 282. Risk factors for disease severity, unimprovement, and mortality of COVID-19 patients in Wuhan, China | Zhang J, Wang X. Jia X. Li J. Hu K. Chen G. Wei J. Gong Z. Zhou C. Yu H. Yu M. Lei H. Cheng F. Zhang B. Xu Y. Wang G. Dong W. | 2020 | Clinical microbiology and infection 10.1016/j.cmi.2020.04.012 |
| 283. Experience with tocilizumab in severe COVID-19 pneumonia after 80 days of follow-up: A retrospective cohort study | Moreno-Perez, Oscar; Andres, Mariano; Leon-Ramirez, Jose-Manuel; Sanchez-Paya, Jose; Rodriguez, Juan Carlos; Sanchez, Rosario; Garcia-Sevila, Raquel; Boix, Vicente; Gil, Joan; Merino, Esperanza | 2020 | Journal of autoimmunity 114 102523 https://dx.doi.org/10.1016/j.jaut.2020.102523 |
| 284. Clinical Characteristics of Coronavirus Disease 2019 in Hainan, China | Yan S, Song X. Lin F. Zhu H. Wang X. Li M. Ruan J. Lin C. Liu X. Wu Q. Luo Z. Fu W. Chen S. Yuan Y. Liu S. Yao J. Lv C. | 2020 | Medrxiv 2020.03.19.20038539 10.1101/2020.03.19.20038539 |
| 285. Outcomes Among Patients Hospitalized With COVID-19 and Acute Kidney Injury | Ng Jh, Hirsch J. S. Hazzan A. Wanchoo R. Shah H. H. Malieckal D. A. Ross D. W. Sharma P. Sakhiya V. Fishbane S. Jhaveri K. D. Northwell Nephrology Covid-Research Consortium | 2020 | American journal of kidney diseases 10.1053/j.ajkd.2020.09.002 |

| | | | |
|---|---|---|---|
| 286. Clinical characteristics of 60 discharged cases of 2019 novel coronavirus-infected pneumonia in Taizhou, China | Jiang Y, He S. Zhang C. Wang X. Chen X. Jin Y. He Z. Cai M. Lin Z. Ying L. Mou J. Zhao H. Lin R. Zhang S. Wu X. Chen H. Lv D. | 2020 | Annals of translational medicine 8, 8 547 10.21037/atm.2020.04.20 |
| 287. Characteristics of recovered COVID-19 patients with recurrent positive RT-PCR findings in Wuhan, China: a retrospective study | Shui Tj, Li C. Liu H. B. Chen X. Zhang B. K. | 2020 | BMC infectious diseases 20, 1 749 10.1186/s12879-020-05463-z |
| 288. A compromised specific humoral immune response against the SARS-CoV-2 receptor-binding domain is related to viral persistence and periodic shedding in the gastrointestinal tract | Hu F, Chen F. Ou Z. Fan Q. Tan X. Wang Y. Pan Y. Ke B. Li L. Guan Y. Mo X. Wang J. Wang J. Luo C. Wen X. Li M. Ren P. Ke C. Li J. Lei C. Tang X. Li F. | 2020 | Cellular & molecular immunology 10.1038/s41423-020-00550-2 |
| 289. Clinical characteristics and risk factors of patients with severe COVID-19 in Jiangsu province, China: a retrospective multicentre cohort study | Liu S, Luo H. Wang Y. Cuevas L. E. Wang D. Ju S. Yang Y. | 2020 | BMC infectious diseases 20, 1 584 10.1186/s12879-020-05314-x |
| 290. Patient Characteristics and Outcomes of 11,721 Patients with COVID19 Hospitalized Across the United States | Fried Mw, Crawford J. M. Mospan A. R. Watkins S. E. Munoz Hernandez B. Zink R. C. Elliott S. Burleson K. Landis C. Reddy K. R. Brown R. S. | 2020 | Clinical infectious diseases 10.1093/cid/ciaa1268 |
| 291. Analysis of Risk Factors on Readmission Cases of COVID-19 in the Republic of Korea: using Nationwide Health Claims Data | Jeon Wh, Seon J. Y. Park S. Y. Oh I. H. | 2020 | International journal of environmental research and public health 17, 16 10.3390/ijerph17165844 |
| 292. Quantifying the prevalence of SARS-CoV-2 long-term shedding among non-hospitalized COVID-19 patients | Agarwal, Vineet; Venkatakrishnan, Aiveliagaram J.; Puranik, Arjun; Lopez-Marquez, Agustin; Challener, Douglas W.; O Horo, John C.; Badley, Andrew D.; Halamka, John D.; Morice, William G.; Soundararajan, Venky | 2020 | medRxiv : the preprint server for health sciences https://dx.doi.org/10.1101/2020.06.02.20120774 |
| 293. A cohort of patients with COVID-19 in a major teaching hospital in Europe | Borobia Am, Carcas A. J. Arnalich F. Alvarez-Sala R. Montserrat J. Quintana M. Figueira J. C. Torres Santos-Olmo R. M. Garcia-Rodriguez J. Martin-Vega A. Ramirez E. Buno A. Martinez-Ales G. Garcia-Arenzana N. Marti de Gracia M. Moreno F. Reinoso-Barbero F. Martin-Quiros A. Rivera A. Mingorance J. Carpio C. C. Prieto Arribas D. Rey Cuevas E. Prados M. C. Rios J. J. Hernan M. Frias J. Arribas J. R. | 2020 | Medrxiv 2020.04.29.20080853 10.1101/2020.04.29.20080853 |
| 294. Clinical characteristics and outcome of hospitalized COVID-19 patients with diabetes: a single-center, retrospective study in Iran | Akbariqomi M, Sadat Hosseini M. Rashidiani J. Sedighian H. Biganeh H. Heidari R. Moosazadeh Moghaddam M. Farnoosh G. Kooshki H. | 2020 | Diabetes research and clinical practice 108467 10.1016/j.diabres.2020.108467 |
| 295. Clinical progression of patients with COVID-19 in Shanghai, China | Chen J, Qi T. Liu L. Ling Y. Qian Z. Li T. Li F. Xu Q. Zhang Y. Xu S. Song Z. Zeng Y. Shen Y. Shi Y. Zhu T. Lu H. | 2020 | Journal of infection 10.1016/j.jinf.2020.03.004 |
| 296. Repeat Chest Ct Scans In Moderate-to-Severe Patients' Management During the Covid-19 Pandemic: observations From a Single Centre in Wuhan, China | Chen L, Wang Q. Wu H. Hu J. Zhang J. | 2020 | Radiation protection dosimetry 10.1093/rpd/ncaa106 |
| 297. Post discharge positive re-tests in COVID-19: common but clinically non-significant | Abdullah Ms, Chong P. L. Asli R. Momin R. N. Mani B. I. Metussin D. Chong V. H. | 2020 | Infectious diseases (london, england) 10.1080/23744235.2020.1780309 |
| 298. Clinical Characteristics of Asymptomatic Patients with SARS-CoV-2 in Zhejiang: an Imperceptible Source of Infection | Dai W, Chen X. Xu X. Leng Z. Yu W. Lin H. Li H. Lin J. Qiu Z. Dai Y. | 2020 | Canadian respiratory journal 2020 2045341 10.1155/2020/2045341 |
| 299. SARS-CoV-2 RT-PCR profile in 298 Indian COVID-19 patients : a retrospective observational study | bhattacharya b, Kumar R. Meena DVp Soneja M. Vig S. Rastogi V. Bhatnagar S. Mohan A. Wig N. | 2020 | Medrxiv 2020.06.19.20135905 10.1101/2020.06.19.20135905 |
| 300. Discharge Clinical Characteristics and Post-Discharge Events in Patients with Severe COVID-19: a Descriptive Case Series | Saab Fg, Chiang J. N. Brook R. Adamson P. C. Fulcher J. A. Halperin E. Manuel V. Goodman-Meza D. | 2021 | Journal of general internal medicine 10.1007/s11606-020-06494-7 |

| | | | |
|---|---|---|---|
| 301. Characteristics, Treatment Outcomes and Role of Hydroxychloroquine among 522 COVID-19 hospitalized patients in Jaipur City: an Epidemio-Clinical Study | Bhandari S, Singh A. Sharma R. Rankawat G. Banerjee S. Gupta V. Dube A. Kakkar S. Sharma S. Keswani P. Agrawal A. Tak A. Nawal C. L. | 2020 | Journal of the Association of Physicians of India 68, 6 13-19 |
| 302. Baseline Characteristics and Associated Factors of Mortality in COVID-19 Patients; an Analysis of 16000 Cases in Tehran, Iran | Zali A, Gholamzadeh S. Mohammadi G. Azizmohammad Looha M. Akrami F. Zarean E. Vafaee R. Maher A. Khodadoost M. | 2020 | Archives of academic emergency medicine 8, 1 e70 |
| 303. Tocilizumab improves significantly clinical outcomes of patients with moderate or severe COVID-19 pneumonia | Assistance Publique - Hopitaux de, Paris | 2020 | |
| 304. Prevalence and Outcomes of D-Dimer Elevation in Hospitalized Patients With COVID-19 | Berger Js, Kunichoff D. Adhikari S. Ahuja T. Amoroso N. Aphinyanaphongs Y. Cao M. Goldenberg R. Hindenburg A. Horowitz J. Parnia S. Petrilli C. Reynolds H. Simon E. Slater J. Yaghi S. Yuriditsky E. Hochman J. Horwitz L. I. | 2020 | Arteriosclerosis, thrombosis, and vascular biology ATVBAHA120314872 10.1161/ATVBAHA.120.314872 |
| 305. Frequency of serological non-responders and false-negative RT-PCR results in SARS-CoV-2 testing: a population-based study | Baron Rc, Risch L. Weber M. Thiel S. Grossmann K. Wohlwend N. Lung T. Hillmann D. Ritzler M. Bigler S. Egli K. Ferrara F. Bodmer T. Imperiali M. Heer S. Renz H. Flatz L. Kohler P. Vernazza P. Kahlert C. R. Paprotny M. Risch M. | 2020 | Clinical chemistry and laboratory medicine 10.1515/cclm-2020-0978 |
| 306. Characteristics and predictors of death among 4,035 consecutively hospitalized patients with COVID-19 in Spain | Berenguer J, Ryan P. Rodriguez-Bano J. Jarrin I. Carratala J. Pachon J. Yllescas M. Arribas J. R. Covid-Spain Study Group | 2020 | Clinical microbiology and infection 10.1016/j.cmi.2020.07.024 |
| 307. COVIDApp as an Innovative Strategy for the Management and Follow-Up of COVID-19 Cases in Long-Term Care Facilities in Catalonia: Implementation Study | Echeverria, Patricia; Mas Bergas, Miquel Angel; Puig, Jordi; Isnard, Mar; Massot, Mireia; Vedia, Cristina; Peiro, Ricardo; Ordorica, Yolanda; Pablo, Sara; Ulldemolins, Maria; Iruela, Merce; Balart, Dolors; Ruiz, Jose Maria; Herms, Jordi; Clotet Sala, Bonaventura; Negredo, Eugenia | 2020 | JMIR public health and surveillance 6, 3 e21163 https://dx.doi.org/10.2196/21163 |
| **Wrong population (n=14)** | | | |
| 308. Psychological distress among people with probable COVID-19 infection: analysis of the UK Household Longitudinal Study | Niedzwiedz, Claire L.; Benzeval, Michaela; Hainey, Kirsten; Leyland, Alastair H.; Katikireddi, Srinivasa Vittal | 2021 | BJPsych open 7, 3 e104 https://dx.doi.org/10.1192/bjo.2021.63 |
| 309. Recovery from COVID-19: a sprint or marathon? 6-month follow-up data from online long COVID-19 support group members | Vaes, Anouk W.; Goertz, Yvonne M. J.; Van Herck, Maarten; Machado, Felipe V. C.; Meys, Roy; Delbressine, Jeannet M.; Houben-Wilke, Sarah; Gaffron, Swetlana; Maier, Dieter; Burtin, Chris; Posthuma, Rein; van Loon, Nicole P. H.; Franssen, Frits M. E.; Hajian, Bita; Simons, Sami O.; van Boven, Job F. M.; Klok, Frederikus A.; Spaetgens, Bart; Pinxt, Claire M. H.; Liu, Limmie Y. L.; Wesseling, Geertjan; Spies, Yvonne; Vijlbrief, Herman; van 't Hul, Alex J.; Janssen, Daisy J. A.; Spruit, Martijn A. | 2021 | ERJ open research 7, 2 https://dx.doi.org/10.1183/23120541.00141-2021 |
| 310. Epidemiological pattern, incidence and outcomes of COVID-19 in liver transplant patients | Colmenero J, Rodriguez-Peralvarez M. Salcedo M. Arias-Milla A. Munoz-Serrano A. Graus J. Nuno J. Gastaca M. Bustamante-Schneider J. Cachero A. Llado L. Caballero A. Fernandez-Yunquera A. Loinaz C. Fernandez I. Fondevilla C. Navasa M. Inarrairaegui M. Castells L. Pascual S. Ramirez P. Vinaixa C. Gonzalez-Dieguez M. L. Gonzalez-Grande R. Hierro L. Nogueras F. Otero A. Alamo J. M. Blanco-Fernandez G. Fabrega E. Garcia-Pajares F. Montero J. L. Tome S. De la Rosa G. Pons J. A. | 2020 | Journal of hepatology 10.1016/j.jhep.2020.07.040 |

| | | | |
|---|---|---|---|
| 311. What Is the Impact of the Coronavirus-19 Pandemic on Quality of Life and Other Patient-reported Outcomes? An Analysis of the Hand-Wrist Study Cohort | Cohen A, Selles R. W. De Ridder W. A. Stege Mhpt Souer J. S. Wouters R. M. | 2020 | Clinical orthopaedics and related research 10.1097/CORR.0000000000001514 |
| 312. Outcomes of novel coronavirus disease 2019 (COVID-19) infection in 107 patients with cancer from Wuhan, China | Zhang H, Wang L. Chen Y. Wu Q. Chen G. Shen X. Wang Q. Yan Y. Yu Y. Zhong Y. Wang X. Chua M. L. K. Xie C. | 2020 | Cancer 10.1002/cncr.33042 |
| 313. Long-term patient-reported symptoms of COVID-19: an analysis of social media data | Juan, M. Banda; Gurdas Viguruji, Singh; Osaid, Alser; Daniel, Prieto-Alhambra | 2020 | Medrxiv 2020. 10.1101/2020.07.29.20164418 |
| 314. Changes in Physical Activity and Sedentary Behavior in Response to COVID-19 and Their Associations with Mental Health in 3052 US Adults | Meyer, Jacob; McDowell, Cillian; Lansing, Jeni; Brower, Cassandra; Smith, Lee; Tully, Mark; Herring, Matthew | 2020 | International journal of environmental research and public health 17, 18 https://dx.doi.org/10.3390/ijerph17186469 |
| 315. Care Dependency in Non-Hospitalized Patients with COVID-19 | Vaes, Anouk W.; Machado, Felipe V. C.; Meys, Roy; Delbressine, Jeannet M.; Goertz, Yvonne M. J.; Van Herck, Maarten; Houben-Wilke, Sarah; Franssen, Frits M. E.; Vijlbrief, Herman; Spies, Yvonne; Van â€™t Hul, Alex J.; Burtin, Chris; Janssen, Daisy J. A.; Spruit, Martijn A. | 2020 | Journal of Clinical Medicine 9, 9 2946-2946 |
| 316. Persistent symptoms after Covid-19: qualitative study of 114 long Covid patients and draft quality criteria for services (preprint) | Ladds E, Rushforth A. Wieringa S. Taylor S. Rayner C. Husain L. Greenhalgh T. | 2020 | Medrxiv 2020.10.13.20211854 10.1101/2020.10.13.20211854 |
| 317. What Is the Impact of the COVID-19 Pandemic on Quality of Life and Other Patient-reported Outcomes? An Analysis of the Hand-Wrist Study Cohort | Cohen A, Selles R. W. De Ridder W. A. Ter Stege M. H. P. Souer J. S. Wouters R. M. Handâ€“Wrist Study Group Collaborators | 2020 | Clinical orthopaedics and related research 10.1097/CORR.0000000000001514 |
| 318. Construct validity of the Post-COVID-19 Functional Status Scale in adult subjects with COVID-19 | Machado Fvc, Meys R. Delbressine J. M. Vaes A. W. Goertz Y. M. J. van Herck M. Houben-Wilke S. Boon Gjam Barco S. Burtin C. van 't Hul A. Posthuma R. Franssen F. M. E. Spies Y. Vijlbrief H. Pitta F. Rezek S. A. Janssen D. J. A. Siegerink B. Klok F. A. Spruit M. A. | 2021 | Health and quality of life outcomes 19, 1 40 10.1186/s12955-021-01691-2 |
| 319. Characterizing Long COVID in an International Cohort: 7 Months of Symptoms and Their Impact (preprint) | Davis He, Assaf G. S. McCorkell L. Wei H. Low R. J. Reâ€™em Y. Redfield S. Austin J. P. Akrami A. | 2020 | Medrxiv 2020.12.24.20248802 10.1101/2020.12.24.20248802 |
| 320. Multi-organ impairment in low-risk individuals with long COVID | Andrea, Dennis; Malgorzata, Wamil; Sandeep, Kapur; Johann, Alberts; Andrew, Badley; Gustav Anton, Decker; Stacey, A. Rizza; Rajarshi, Banerjee; Amitava, Banerjee | 2020 | 10.1101/2020.10.14.20212555 |
| 321. Post-acute COVID-19 syndrome negatively impacts health and wellbeing despite less severe acute infection | Laura, Tabacof; Jenna, Tosto-Mancuso; Jamie, Wood; Mar, Cortes; Amy, Kontorovich; Dayna, McCarthy; Dahlia, Rizk; Leila, Nasr; Erica, Breyman; Nicki, Mohammadi; Christopher, Kellner; David, Putrino | 2020 | Medrxiv 2020. 10.1101/2020.11.04.20226126 |
| **Wrong setting (n=2)** | | | |
| 322. Residual Lung Injury in Patients Recovering From COVID-19 Critical Illness: A Prospective Longitudinal Point-of-Care Lung Ultrasound Study | Alharthy, A.; Abuhamdah, M.; Balhamar, A.; Faqihi, F.; Nasim, N.; Ahmad, S.; Noor, A.; Tamim, H.; Alqahtani, S. A.; Abdulaziz Al Saud, Aaasb; Kutsogiannis, D. J.; Brindley, P. G.; Memish, Z. A.; Karakitsos, D.; Blaivas, M. | 2020 | Journal of ultrasound in medicine : official journal of the American Institute of Ultrasound in Medicine 10.1002/jum.15563 |
| 323. CT Quantification and Machine-learning Models for Assessment of Disease Severity and Prognosis of COVID-19 Patients | Cai W, Liu T. Xue X. Luo G. Wang X. Shen Y. Fang Q. Sheng J. Chen F. Liang T. | 2020 | Academic radiology 10.1016/j.acra.2020.09.004 |
| **Wrong study design (n=13)** | | | |
| 324. Prevalence of Functional Limitation in COVID-19 Recovered Patients Using the Post COVID-19 Functional Status Scale | Pant, Pankaj; Joshi, Aishana; Basnet, Babin; Shrestha, Bibek Man; Bista, Navindra Raj; Bam, Niraj; Das, Santa Kumar | 2021 | JNMA; journal of the Nepal Medical Association 59, 233 https://dx.doi.org/10.31729/jnma.5980 |

| | | | | |
|---|---|---|---|---|
| 325. | Long term morbidity and mortality in covid patients discharged from hospital with or without steroid as discharge medication | Routray, P.; Samal, S.; Mishra, D. | 2021 | Intensive Care Medicine Experimental. Conference: European Society of Intensive Care Medicine Annual Congress, 9 SUPPL 1 http://dx.doi.org/10.1186/s40635-021-00415-6 |
| 326. | Prevalence of mental illness among COVID-19 survivors in South Korea: nationwide cohort | Park, Hye Yoon; Song, In-Ae; Lee, So Hee; Sim, Min Young; Oh, Hong Sang; Song, Kyoung-Ho; Yu, Eun-Seung; Park, Hye Youn; Oh, Tak Kyu | 2021 | BJPsych open 7, 6 e183 https://dx.doi.org/10.1192/bjo.2021.1001 |
| 327. | Outcomes Among Patients Referred to Outpatient Rehabilitation Clinics After COVID-19 diagnosis - United States, January 2020-March 2021 | Rogers-Brown, Jessica S.; Wanga, Valentine; Okoro, Catherine; Brozowsky, Diane; Evans, Alan; Hopwood, David; Cope, Jennifer R.; Jackson, Brendan R.; Bushman, Dena; Hernandez-Romieu, Alfonso C.; Bonacci, Robert A.; McLeod, Tim; Chevinsky, Jennifer R.; Goodman, Alyson B.; Dixon, Meredith G.; Lutfy, Caitlyn; Rushmore, Julie; Koumans, Emily; Morris, Sapna Bamrah; Thompson, William | 2021 | MMWR. Morbidity and mortality weekly report 70, 27 967-971 https://dx.doi.org/10.15585/mmwr.mm7027a2 |
| 328. | Evaluating the depression, anxiety, stress, and predictors of psychological morbidity among COVID-19 Survivors in Mashhad, Iran | Salimi, Z.; Najafi, R.; Khalesi, A.; Oskoei, R.; Moharreri, F.; Khaniki, S. H.; Shahini, N.; Soltanifar, A.; Ardabili, H. M. | 2021 | Iranian Journal of Psychiatry and Behavioral Sciences 15(2) e108972 http://dx.doi.org/10.5812/ijpbs.108972 |
| 329. | "Post-COVID-19 syndrome:" The New Pandemic Affecting Healthcare Workers and How the Frontline Warriors Are Battling it | Rao, S.; Amara, V.; Chaudhuri, S.; Rao, B. K.; Todur, P. | 2021 | Indian Journal of Palliative Care 27, 2 313-318 https://dx.doi.org/10.25259/IJPC_160_21 |
| 330. | Severe Fatigue and Memory Impairment Are Associated with Lower Serum Level of Anti-SARS-CoV-2 Antibodies in Patients with Post-COVID Symptoms | Molnar, T.; Varnai, R.; Schranz, D.; Zavori, L.; Peterfi, Z.; Sipos, D.; Tokes-Fuzesi, M.; Illes, Z.; Buki, A.; Csecsei, P. | 2021 | Journal of Clinical Medicine 10, 19 23 https://dx.doi.org/10.3390/jcm10194337 |
| 331. | Prevalence of Post-traumatic Stress Symptoms and Its Associations With Quality of Life, Demographic and Clinical Characteristics in COVID-19 Survivors During the Post-COVID-19 Era | Yuan, Yuan; Liu, Zi-Han; Zhao, Yan-Jie; Zhang, Qinge; Zhang, Ling; Cheung, Teris; Jackson, Todd; Jiang, Guo-Qing; Xiang, Yu-Tao | 2021 | Frontiers in psychiatry 12 665507 https://dx.doi.org/10.3389/fpsyt.2021.665507 |
| 332. | Brain imaging before and after COVID-19 in UK Biobank | Douaud, Gwenaelle; Lee, Soojin; Alfaro-Almagro, Fidel; Arthofer, Christoph; Wang, Chaoyue; Lange, Frederik; Andersson, Jesper L. R.; Griffanti, Ludovica; Duff, Eugene; Jbabdi, Saad; Taschler, Bernd; Winkler, Anderson; Nichols, Thomas E.; Collins, Rory; Matthews, Paul M.; Allen, Naomi; Miller, Karla L.; Smith, Stephen M. | 2021 | medRxiv : the preprint server for health sciences https://dx.doi.org/10.1101/2021.06.1 1.21258690 |
| 333. | Efficacy and safety of tocilizumab in severe COVID-19 patients: a single-centre retrospective cohort study | Campochiaro C, Della-Torre E. Cavalli G. De Luca G. Ripa M. Boffini N. Tomelleri A. Baldissera E. Rovere-Querini P. Ruggeri A. Monti G. De Cobelli F. Zangrillo A. Tresoldi M. Castagna A. Dagna L. | 2020 | European journal of internal medicine 76 43-49 10.1016/j.ejim.2020.05.021 |
| 334. | Early COVID-19 Therapy with Azithromycin Plus Nitazoxanide, Ivermectin or Hydroxychloroquine in Outpatient Settings Significantly Reduced Symptoms Compared to Known Outcomes in Untreated Patients | Flavio, A. Cadegiani; Andy, Goren; Carlos Gustavo, Wambier; John, McCoy | 2021 | New Microbes New Infect. 2021 Sep;43:100915. doi: 10.1016/j.nmni.2021.100915. Epub 2021 Jul 7 |
| 335. | Pre-existing conditions are associated with long-COVID patientsâ€™ hospitalization, despite confirmed clearance of SARS-CoV-2 virus (preprint) | Pawlowski C, Venkatakrishnan A. J. Ramudu E. Kirkup C. Puranik A. Kayal N. Berner G. Anand A. Barve R. Oâ€™Horo J. C. Badley A. D. Soundararajan V. | 2020 | Medrxiv 2020.10.28.20221655 10.1101/2020.10.28.20221655 |
| 336. | Identifying patients at risk of post-discharge complications related to COVID-19 infection | Hall J, Myall K. Lam J. L. Mason T. Mukherjee B. West A. Dewar A. | 2021 | Thorax 10.1136/thoraxjnl-2020-215861 |

Supplementary Table 2: Risk of bias

Adapted Newcastle Ottawa Scale: ○ (0.5) ● (1) or zero

| | Author | Selection 1 | Selection 2 | Selection 3 | Compara-bility | Outcome 1 | Outcome 2 | Outcome 3 | Outcome 4 | Outcome 5 | Outcome 6 |
|---|---|---|---|---|---|---|---|---|---|---|---|
| 1. | Abdelrahman, M et al(1) | ● | | | ● | ● | | ● | | ● | |
| 2. | Al-Aly, Z et al *(Split cohort: non-hospitalised)*(2) | ● | ● | ● | ● | ● | ● | ● | ● | ● | |
| *2a.* | Al-Aly, Z et al *(Split cohort: hospitalised)*(2) | ● | | ● | ● | ● | ● | ● | ● | ● | |
| 3. | Aminian, A et al(3) | ● | ● | | ● | ● | ● | | | ● | |
| 4. | Arnold, D et al(4) | ○ | | | ● | ● | ● | ● | ● | | |
| 5. | Augustin, M et al(5) | ● | ● | | ● | ● | ● | ● | ● | | |
| 6. | Ayoubkhani, D et al(6) | ○ | | ● | ● | ● | ● | | | ● | |
| 7. | Baricich, A et al(7) | ○ | | ● | ● | | ● | | | | |
| 8. | Becker, J et al(8) | ● | ● | | ● | ● | ● | | | ● | |
| 9. | Bellan, M et al(9) | ● | | ● | ● | ● | ● | ● | | ● | |
| 10. | Blanco, J et al(10) | ● | | | ● | ● | ● | | ● | ● | |
| 11. | Bliddal, S et al(11) | ● | | | ● | ● | ● | ● | ● | ● | |
| 12. | Blomberg, B et al(12) | ● | | ● | ● | ● | ● | ● | ● | ● | |
| 13. | Boscolo-Rizzo, P et al(13) | ● | ● | | ● | ● | ● | ● | ● | ● | |
| 14. | Carrillo-Garcia, P et al(14) | ○ | | ● | ● | ● | ● | ● | ● | ● | |
| 15. | Caruso, D et al(15) | ● | | | ● | ● | ● | ● | | ● | |
| 16. | Caspersen, I et al(16) | ● | ● | ● | ● | ● | ● | ● | ● | ● | |
| 17. | Castro, V et al(17) | ● | | ● | | ● | ● | ● | ● | ● | |
| 18. | Chai, C et al(18) | ● | | | | ● | ● | ● | ● | | |
| 19. | Cirulli et al(19) | ● | ● | ● | ● | ● | ● | ● | ● | | |
| 20. | Clavario, P et al(20) | ● | | | ● | ● | ● | ● | ● | ● | |
| 21. | Cristillo, V et al(21) | | | | ● | | ● | | ● | | |
| 22. | Diaz-Fuentes, G et al(22) | ● | | | ● | ● | ● | ● | | ● | |
| 23. | Domenech-Montoliu, S et al(23) | ● | ● | | ● | ● | ● | ● | ● | ● | |
| 24. | Erol, N et al(24) | ● | | ● | | ● | ● | ● | | ● | |
| 25. | Evans, R et al (PHOSP-COVID study) (25) | ○ | | ● | ● | ● | ● | ● | | | ● |
| 26. | Evans, R et al (PHOSP-COVID study) (26) | ○ | | ● | ● | | ● | ● | | ● | |
| 27. | Fernandez-de-Las-Penas, C et al (27) | ● | | | ● | ● | ● | ● | ● | ● | |
| 28. | Fernandez-de-Las-Penas, C et al(28) | ● | | | ● | ● | ● | | ● | ● | |
| 29. | Fernandez-de-Las-Penas, C et al(29) | ● | | ● | ● | ● | ● | | ● | ● | |

| | | | | | | | | | | | |
|---|---|---|---|---|---|---|---|---|---|---|---|
| 30. | Frija-Masson, J et al(30) | ● | | | ● | ● | ● | | ● | ● | ● |
| 31. | Froidure, A et al(31) | ● | | | ● | | ● | | ● | ● | |
| 32. | Fu, L et al(32) | ○ | | ● | ● | ● | ● | | ● | ● | |
| 33. | Gaber, T et al(33) | ○ | | | ● | ● | | ● | | | |
| 34. | Garcia-Abellan, J et al(34) | ● | | | ● | | ● | ● | ● | ● | |
| 35. | Garratt, A et al (35) | ● | | ● | ● | ● | ● | ● | | ● | |
| 36. | Gonzalez-Hermosillo, J et al(36) | ● | | ● | ● | ● | ● | ● | ● | ● | |
| 37. | Han, X et al(37) | ● | | | ● | ● | ● | | ● | ● | |
| 38. | Havervall, S et al(38) | ● | | ○ | ● | ● | ● | ● | ● | ● | |
| 39. | Huang, C et al(39) | ● | | | ● | | ● | ● | ● | ● | ● |
| 40. | Huang, L et al(40) | ● | | ● | ● | ● | ● | ● | ● | ● | |
| 41. | Jacobson, K et al(41) | ● | | | | ● | ● | ● | ● | ● | |
| 42. | Kashif, A et al(42) | ● | ● | | ● | ● | ● | ● | | ● | |
| 43. | Kim, Y et al(43) | ● | | | ● | ● | ● | ● | ● | | |
| 44. | Lemhofer, C et al(44) | ● | ● | | ● | ● | ● | ● | ● | | |
| 45. | Li, X et al(45) | ● | | | ● | | ● | | ● | | |
| 46. | Liao, T et al(46) | ● | | | ● | | ● | | | | |
| 47. | Liao, X et al(47) | ● | | | ● | | ● | | | ● | |
| 48. | Liu, Y-H et al(48) | ○ | | ● | ● | ● | ● | | | | |
| 49. | Liyanage-Don, A et al(49) | | | ● | ● | ● | ● | ● | | | |
| 50. | Logue, J et al(50) | ● | | ● | ● | | ● | ● | | ● | |
| 51. | Lucidi, T et al(51) | | | | ● | ● | ● | | ● | ● | |
| 52. | Lui, D et al(52) | ● | ● | ● | ● | ● | ● | ● | ● | ● | |
| 53. | Maestre-Muniz, M et al(53) | ● | | | ● | ● | ● | ● | | ● | |
| 54. | Martinez, A et al(54) | ● | | ● | ● | ● | ● | ● | ● | ● | |
| 55. | Matteudi, T et al(55) | ● | | | ● | ● | ● | ● | | | |
| 56. | Mazza, M et al(56) | ● | | ● | ● | ● | ● | | ● | ● | |
| 57. | Mechi, A et al(57) | ● | | | ● | ● | ● | ● | ● | ● | |
| 58. | Mei, Q et al(58) | ○ | | ● | ● | ● | ● | | | ● | |
| 59. | Mei, Q et al(59) | ● | | | ● | ● | ● | ● | ● | ● | |
| 60. | Menges, D et al(60) | ● | | ● | ● | ● | ● | ● | | ● | |
| 61. | Milanese, M et al(61) | ○ | | | ● | ● | ● | | | ● | ● |
| 62. | Millet, C et al(62) | ● | ● | | ● | ● | ● | ● | ● | | |
| 63. | Mohiuddin Chowdhury, A et al(63) | ● | | | ● | ● | | ● | ● | | |
| 64. | Munblit, D et al(64) | ○ | | | ● | ● | ● | ● | ● | | |

| | | | | | | | | | | | |
|---|---|---|---|---|---|---|---|---|---|---|---|
| 65. | Nabahati, M et al(65) | ● | | | ● | | ● | | | | |
| 66. | Nehme, M, et al(66) | ● | ● | ● | ● | ● | ● | ● | ● | | |
| 67. | Nguyen, N et al(67) | ● | | ● | ● | ● | ● | | ● | | |
| 68. | Nunez-Fernandez, M et al(68) | ● | | ● | ● | ● | ● | | ● | ● | |
| 69. | O'Keefe, J et al(69) | ● | | ● | ● | ● | ● | ● | | ● | |
| 70. | Office for National Statistics(70) | ● | ● | ● | ● | ● | ● | ● | ● | ● | |
| 71. | Ong, S et al(71) | ● | | | ● | ● | ● | ● | ● | | |
| 72. | Orru, G et al(72) | | | | | | | ● | | | |
| 73. | Osmanov, I et al(73) | ● | | ● | ● | ● | ● | ● | | | |
| 74. | Peghin M, et al(74) | ○ | | | ● | ● | ● | ● | ● | | ● |
| 75. | Peluso, M et al(75) | ● | | ● | ● | ● | ● | ● | ● | ● | |
| 76. | Petersen, M et al(76) | ● | ● | | ● | ● | ● | ● | | ● | |
| 77. | Qin, W et al(77) | ● | | | ● | | ● | ● | ● | ● | |
| 78. | Qu, G et al(78) | ● | | | ● | ● | ● | ● | | ● | |
| 79. | Radtke, T et al(79) | ● | ● | ● | ● | ● | ● | ● | ● | ● | |
| 80. | Rass, V et al(80) | ● | | ● | ● | | ● | ● | ● | ● | |
| 81. | Riestra-Ayora, J et al(81) | ● | | ● | ● | ● | ● | | | ● | |
| 82. | Righi, E et al(82) | ● | ● | ● | ● | ● | ● | ● | ● | | |
| 83. | Roessler, M et al(83) *(Split cohort: adults)* | ● | ● | ● | ● | ● | ● | ● | ● | ● | |
| 83a. | Roessler, M et al(83) *(Split cohort: children)* | ● | ● | ● | ● | ● | ● | ● | ● | ● | |
| 84. | Romero-Duarte, A et al(84) | ● | | | ● | ● | ● | ● | ● | ● | |
| 85. | Sathyamurthy, P et al(85) | ● | | ● | ● | ● | ● | ● | | ● | |
| 86. | Seeβle, J et al(86) | ● | | ● | ● | | ● | ● | ● | | |
| 87. | Shang, Y et al(87) | ● | | | ● | ● | ● | ● | ● | | |
| 88. | Sibila, O et al(88) | ○ | | | ● | ● | ● | ● | ● | ● | |
| 89. | Sigfrid, L et al(89) | ○ | | ● | ● | ● | ● | ● | | | |
| 90. | Simani, L et al(90) | ○ | | ● | ● | ● | ● | | ● | | ● |
| 91. | Skala, M et al(91) | ● | | ● | ● | ● | ● | ● | ● | ● | |
| 92. | Skjorten, I et al(92) | ○ | | | ● | ● | ● | | ● | ● | |
| 93. | Sonnweber, T et al(93) | ● | | ● | ● | ● | ● | ● | ● | ● | |
| 94. | Soraas, A et al(94) | ● | | ● | | ● | ● | ● | ● | ● | |
| 95. | Soraas, A et al(95) | ● | | ● | ● | ● | ● | ● | ● | ● | |
| 96. | Stavem, K et al(96) | ● | | | ● | ● | ● | ● | | | |
| 97. | Stavem, K et al(97) | ● | ● | | ● | ● | ● | | | | |

| | | | | | | | | | | | |
|---|---|---|---|---|---|---|---|---|---|---|---|
| 98. | Stephenson, T et al(98) | ● | | ● | ● | ● | ● | ● | ● | ● | |
| 99. | Sudre, C et al(99) | ● | ● | ● | ● | ● | ● | ● | ● | ● | |
| 100. | Sykes, D et al(100) | ● | | | ● | ● | ● | ● | | | |
| 101. | Taboada, M et al(101) | ● | | ● | ● | ● | ● | | ● | ● | |
| 102. | Taquet, M et al(102) | ○ | | ● | ● | | ● | | ● | ● | |
| 103. | Taquet, M et al(103) | ● | ● | ● | ● | ● | ● | ● | ● | ● | |
| 104. | Tarsitani, L et al(104) | ○ | | ● | ● | ● | ● | | ● | | |
| 105. | Tawfik, H et al(105) | ● | | | ● | ● | ● | ● | | | |
| 106. | Taylor, R et al(106) | | | ● | ● | ● | ● | ● | ● | ● | |
| 107. | Tempany, M et al(107) | ● | | | ● | ● | ● | ● | | ● | |
| 108. | The Writing Committee for the COMEBAC Study Group(108) | ○ | | ● | | | ● | ● | ● | | |
| 109. | Tholin, B et al(109) | ● | | | ● | ● | | | ● | ● | |
| 110. | Tleyjeh, I et al(110) | ● | | ● | ● | ● | ● | ● | ● | ● | ● |
| 111. | Todt, B et al(111) | ● | | ● | ● | ● | ● | ● | ● | | |
| 112. | Tohamy, D et al(112) | ● | | ● | | ● | ● | | ● | ● | |
| 113. | Townsend, L et al(113) | ● | | ● | ● | ● | ● | | | | ● |
| 114. | Trunfio, M et al(114) | ● | ● | | ● | ● | ● | ● | ● | ● | |
| 115. | Ursini, F et al(115) | ● | ● | | ● | ● | ● | ● | | ● | |
| 116. | Venturelli, S et al(116) | ● | | ● | ● | | ● | ● | ● | ● | |
| 117. | Walle-Hansen, M et al(117) | ● | | ● | ● | | ● | | ● | | |
| 118. | Weng, J et al(118) | ● | | ● | ● | ● | | ● | ● | ● | |
| 119. | Whitaker, M et al(119) | | | | ● | ● | ● | ● | ● | ● | |
| 120. | Xiong, L et al(120) | ○ | | | ● | | ● | | | | |
| 121. | Xiong, Q et al(121) | ● | | ● | | ● | ● | ● | | | |
| 122. | Yan, B et al(122) | ● | | | ● | ● | | | | ● | |
| 123. | Yan, X et al(123) | ● | | | ● | ● | ● | | | ● | |
| 124. | Yin, X et al(124) | ● | | | ● | | ● | | | ● | |
| 125. | Zayet, S et al(125) | ● | | ● | ● | ● | ● | ● | ● | ● | |
| 126. | Zhan, Y et al(126) | ● | | | ● | ● | ● | | ● | ● | |
| 127. | Zhang, D et al(127) | ● | | | | | ● | | ● | | |
| 128. | Zhang, J et al(128) | ● | | | ● | | ● | | ● | | |
| 129. | Zhang, X et al(129) | ● | | | ● | ● | ● | ● | ● | ● | |
| 130. | Zhou, M et al(130) | ● | | ● | ● | ● | ● | ● | ● | ● | |

Supplementary Table 3: Adapted Newcastle-Ottawa Scale Risk of Bias Tool

| | | |
|---|---|---|
| Selection/denominator 1: Definition/diagnostic criteria | ● | Confirmed by laboratory (any - Ag/Ab/PCR) |
| | ○ | clinical diagnosis – not lab confirmed |
| | | Self-diagnosed – not lab or clinically confirmed |
| Selection/denominator 2: Representativeness of all COVID-19 cases in the population (adult or children) | ● | Representative of community-based COVID19 cases |
| | | hospitalised only |
| | | <70% response rate or response rate not stated |
| | | Intensive care patients included but proportion not stated and/or ICU patient outcome data not separated/stratified |
| | | Healthcare workers only or other demographic-specific e.g. age, gender |
| | | Pre-existing condition patient group e.g. transplant patients |
| | | Specific groups e.g. support groups |
| Selection/denominator 3: Outcome of interest (LC symptoms) was not present prior to infection | ● | Health status assessment pre-coronavirus |
| | | No health status assessment |
| | ● | Comparison to test-negative controls |
| Comparability: Source of participants without LC symptoms | ● | Drawn from same source as those with LC symptoms |
| | | drawn from a different source |
| | | no description |
| Outcome/numerator 1: Systematic assessment | ● | All participants assessed |
| | | Targeted/unstructured |
| | | Different methods of data collection used for different participants |
| Outcome/numerator 2: Quality of assessment | ● | Clinician diagnosis of LC symptoms/pathology using a structured approach |
| | ● | Systematic symptoms/pathology assessment |
| | ● | Standardised rating scales |
| | ● | High-quality qualitative methods |
| | | Unstructured reporting of diagnosis, symptoms or severity |
| Outcome/numerator 3: Comprehensiveness of LC symptoms/pathology assessed | ● | Good range of LC symptoms assessed with severity captured |
| | | Few symptoms assessed |
| | | single pathology assessed |
| Outcome/numerator 4: Follow-up period | ● | Same time-point for all, follow-up time adequate |
| | | Follow-up time-point based on hospital admission or length of hospitalisation, with no indication of symptom-onset or test date |
| | | Unclear length of illness |
| | | Not all patients discharged, so actual length of illness unknown |
| Outcome/numerator 5: Loss to follow-up | ● | complete follow-up – all subjects accounted for |
| | ● | subjects lost to follow-up unlikely to introduce bias – small number lost [> 70% followed-up] or description provided of those lost |
| | | no statement |
| | | no follow-up (cross-sectional) |
| Outcome/numerator 6: Effect of interventions | ● | Differential effect of intervention considered e.g. treatment |
| | | No consideration of interventions |

Supplementary Table 4: Subgroup analysis

| **Subgroup** | **Number of studies** | **Range of Long Covid prevalence** | **Pooled estimate** | **Prediction intervals** | **Figure** |
|---|---|---|---|---|---|
| Length of follow-up | | | | | Supplementary Figure 1 |
| 12wks – 5m | 73 | 0% - 92% | 39.8% | 5.1% - 89.1% | |
| 6 – 11m | 49 | 10% - 93% | 44.9% | 8.0% - 88.4% | |
| 12+m | 12 | 17% - 81% | 48.5% | 12.7% - 86% | |
| Percentage hospitalised | | | | | Supplementary Figure 2 |
| <10% | 24 | 0% - 67% | 26.4% | 2.6% - 82.8% | |
| 10-99% | 29 | 3% - 79% | 41.1% | 8.4% - 84.2% | |
| 100% | 65 | 5% - 93% | 47.5% | 8.3% - 90.0% | |
| Percentage admitted to intensive care | | | | | Supplementary Figure 3 |
| <5% | 48 | 0% - 86% | 34.9% | 5.2% - 84.0% | |
| 5% – 9.9% | 16 | 3% - 81% | 41.8% | 3.5% - 93.5% | |
| 10+% | 31 | 5% - 93% | 48.8% | 5.7% - 93.7% | |
| Severity of acute infection | | | | | Supplementary Figure 4 |
| Ambulatory mild disease: symptomatic, independent | 17 | 0% - 67% | 23.5% | 1.6% - 85.7% | |
| Ambulatory mild disease: symptomatic, assistance sought | 21 | 3% - 62% | 35.1% | 5.8% - 82.5% | |
| Hospitalised moderate disease: no oxygen therapy | 17 | 10% - 86% | 46.1% | 7.0% - 90.6% | |
| Hospitalised moderate disease: oxygen by mask or nasal prongs | 37 | 5% - 81% | 44.0% | 10.7% - 83.7% | |
| Hospitalised severe disease: oxygen by NIV or high flow | 27 | 9% - 93% | 54.8% | 7.7% - 94.7% | |
| Sample recruitment source | | | | | Supplementary Figure 42 |
| Community | 21 | 3% - 67% | 25.6% | 1.8% - 86.3% | |
| Healthcare workers | 8 | 21% - 70% | 38.4% | 9.3% - 79.0% | |
| Outpatients | 17 | 5% - 82% | 45.5% | 8.4% - 88.4% | |
| Social media | 3 | 44% - 74% | 60.4% | 0.0% - 100.0% | |

| Subgroup | Number of studies | Range of Long Covid prevalence | Pooled estimate | Prediction intervals | Figure |
|---|---|---|---|---|---|
| Hospitalised | 69 | 0% - 93% | 46.5% | 8.2% - 89.4% | |
| Method of outcome assessment | | | | | Figure 3 |
| Pathology discovered at follow-up | 20 | 5% - 86% | 51.7% | 12.3% - 89.1% | |
| Symptoms & function discovered at follow-up | 93 | 0% - 93% | 43.9% | 8.2% - 87.2% | |
| Outcomes from health record linkage | 9 | 3% - 44% | 13.6% | 1.2% - 68.0% | |
| Risk factor or comorbidities | | | | | Supplementary Figure 43 |
| <50yrs | 41 | - | 38.5% | 7.9% - 82.1% | |
| 50+yrs | 66 | - | 47.7% | 7.9% - 90.6% | |
| <50% female | 59 | - | 45.6% | 5.5% - 92.4% | |
| 50+% female | 58 | - | 38.7% | 8.5% - 81.2% | |
| <50% white ethnicity | 8 | - | 56.3% | 22.3% - 85.2% | |
| 50+% white ethnicity | 16 | - | 37.6% | 1.7% - 95.3% | |
| <10% current smoker | 21 | - | 55.7% | 11.4% - 92.4% | |
| 10+% current smoker | 32 | - | 50.5% | 12.4% - 88.0% | |
| <20% pre-existing obesity | 18 | - | 37.5% | 3.7% -90.3% | |
| 20+% pre-existing obesity | 19 | - | 48.7% | 5.5% - 94.0% | |
| <10% pre-existing diabetes | 34 | - | 35.4% | 5.7% - 83.2% | |
| 10+% pre-existing diabetes | 41 | - | 51.9% | 8.3% - 92.8% | |
| <30% pre-existing hypertension | 42 | - | 37.3% | 7.0% - 82.5% | |
| 30+% pre-existing hypertension | 30 | - | 58.5% | 16.9% - 90.7% | |
| 5+% pre-existing cholesterol disorder | 13 | - | 52.6% | 18.3% - 84.6% | |
| <10% pre-existing CVD | 39 | - | 38.2% | 5.9% - 85.9% | |
| 10+% pre-existing CVD | 30 | - | 54.7% | 9.4% - 93.4% | |
| <5% pre-existing cancer | 34 | - | 44.9% | 6.0% - 91.2% | |
| 5+% pre-existing cancer | 13 | - | 43.9% | 11.1% - 83.1% | |
| <10% pre-existing respiratory disease | 40 | - | 40.1% | 6.4% - 86.6% | |
| 10+% pre-existing respiratory disease | 29 | - | 49.3% | 7.4% - 92.2% | |
| <5% pre-existing liver disease | 17 | - | 39.3% | 3.4% - 92.2% | |

| Subgroup | Number of studies | Range of Long Covid prevalence | Pooled estimate | Prediction intervals | Figure |
|---|---|---|---|---|---|
| 5+% pre-existing liver disease | 5 | - | 64.3% | 12.3% - 95.9% | |
| <5% pre-existing kidney disease | 23 | - | 42.3% | 7.8% - 86.4% | |
| 5+% pre-existing kidney disease | 13 | - | 61.2% | 5.6% - 97.7% | |
| 5+% pre-existing thyroid disease | 4 | - | 36.2% | 2.3% - 93.1% | |
| <5% pre-existing neurological disorder | 5 | - | 31.2% | 1.4% - 93.7% | |
| 5+% pre-existing neurological disorder | 7 | - | 34.5% | 2.2% - 92.4% | |
| <5% pre-existing immunological disorder or allergy | 9 | - | 27.8% | 2.9% - 83.1 % | |
| 5+% pre-existing immunological disorder or allergy | 15 | - | 52.0% | 3.8% - 96.7% | |
| <10% pre-existing anxiety or depression | 5 | - | 43.2% | 0.1% - 99.8% | |
| Domains of outcome assessment | | | | | Supplementary Figure 44 |
| Single domain | 51 | 3% - 86% | 41.4% | 6.9% - 87.1% | |
| Multiple domains | 71 | 0% - 93% | 42.6% | 6.3% - 89.2% | |
| Risk difference in studies with controls | 26 | -1% - 55% (RD) | 13.9% (RD) | -16.2% - 43.9% (RD) | Supplementary Figure 45 |
| Relative risk in community-based samples with controls | 13 | 1.0 – 51.4 (RR) | 2.7 (RR) | 0.2 – 39.4 (RR) | Supplementary Figure 46 |
| Risk difference in community-based samples with controls | 14 | -1% – 35% (RD) | 10.1% (RD) | -12.7% - 32.8% (RD) | Supplementary Figure 47 |
| Relative risk in community-based samples with controls with low risk of bias | 4 | 1.30 – 1.63 (RR) | 1.33 (RR) | 1.30 – 1.36 (RR) | Figure 4 |
| Risk difference in community-based samples with controls with low risk of bias | 4 | 1% - 9% (RD) | 4.8% (RD) | -13.2% - 22.7% (RD) | Supplementary Figure 48 |
| Study design: | | | | | Supplementary Figure 50 |
| Cohort | 102 | 0% - 93% | 41.3% | 6.0% - 88.6% | |
| Cross-sectional | 20 | 10% - 82% | 45.9% | 11.2% - 85.1% | |

### 3. Supplementary methods

An average WHO Clinical Progression Scale (WHO CPS) value was assigned to each study based on the reported proportion of recruited participants in the acute phase described as being asymptomatic, symptomatic but independent, symptomatic seeking assistance, hospitalised, receiving different types of supplemental oxygen, or admitted to intensive care units. The average WHO CPS score was allocated to each study was based on the WHO Clinical Progression Scale for individuals(131) and published estimates of severity(132) . Where exact WHO CPS categories were not reported, an average was taken over potential categories consistent with the information reported. If type of hospital treatment was not reported, we assumed hospitalisation implied half the patients received low flow oxygen and half received high flow oxygen or non-invasive ventilation. Unless otherwise indicated, we assumed patients admitted to ICU were allocated half to low flow oxygen or non-invasive ventilation and half to mechanical ventilation. If ICU use was not reported in community-based settings and occupational cohorts, we assumed this was half of the proportion of participants who were hospitalised.

### 4. Supplementary figures

Supplementary Figure 1: Forest plot of prevalence of Long Covid in the included studies by length of follow-up with 95% prediction intervals

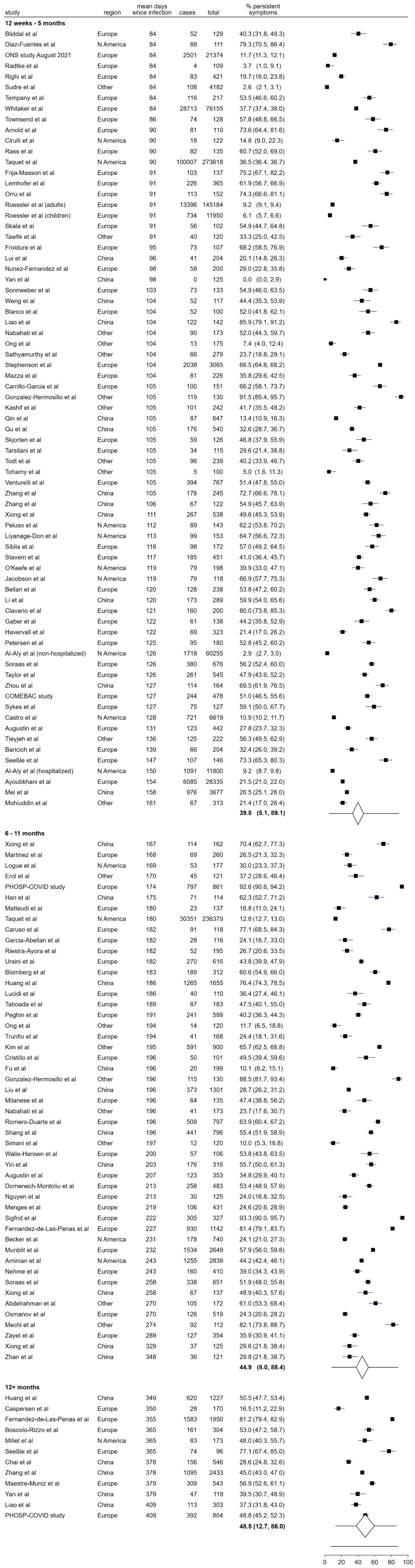

Supplementary Figure 2: Forest plot of prevalence of Long Covid in the included studies by percentage hospitalised with 95% prediction intervals

| study | region | mean days since infection | cases | total | % persistent symptoms |
|---|---|---|---|---|---|
| **<10% hospitalised** | | | | | |
| Al-Aly et al (non-hospitalized) | N America | 126 | 1718 | 60255 | 2.9 (2.7, 3.0) |
| Augustin et al | Europe | 131 | 123 | 442 | 27.8 (23.7, 32.3) |
| Bliddal et al | Europe | 84 | 52 | 129 | 40.3 (31.8, 49.3) |
| Boscolo-Rizzo et al | Europe | 365 | 161 | 304 | 53.0 (47.2, 58.7) |
| Caspersen et al | Europe | 350 | 28 | 170 | 16.5 (11.2, 22.9) |
| Cirulli et al | N America | 90 | 18 | 122 | 14.8 (9.0, 22.3) |
| Domenech-Montoliu et al | Europe | 213 | 258 | 483 | 53.4 (48.9, 57.9) |
| Gaber et al | Europe | 122 | 61 | 138 | 44.2 (35.8, 52.9) |
| Havervall et al | Europe | 122 | 69 | 323 | 21.4 (17.0, 26.2) |
| Kashif et al | Other | 105 | 101 | 242 | 41.7 (35.5, 48.2) |
| Lemhofer et al | Europe | 91 | 226 | 365 | 61.9 (56.7, 66.9) |
| Martinez et al | Europe | 168 | 69 | 260 | 26.5 (21.3, 32.3) |
| Nehme et al | Europe | 243 | 160 | 410 | 39.0 (34.3, 43.9) |
| ONS study August 2021 | Europe | 84 | 2501 | 21374 | 11.7 (11.3, 12.1) |
| Petersen et al | Europe | 125 | 95 | 180 | 52.8 (45.2, 60.2) |
| Radtke et al | Europe | 84 | 4 | 109 | 3.7 (1.0, 9.1) |
| Riestra-Ayora et al | Europe | 182 | 52 | 195 | 26.7 (20.6, 33.5) |
| Roessler et al (adults) | Europe | 91 | 13396 | 145184 | 9.2 (9.1, 9.4) |
| Roessler et al (children) | Europe | 91 | 734 | 11950 | 6.1 (5.7, 6.6) |
| Soraas et al | Europe | 126 | 380 | 676 | 56.2 (52.4, 60.0) |
| Stavem et al | Europe | 117 | 185 | 451 | 41.0 (36.4, 45.7) |
| Stephenson et al | Europe | 104 | 2038 | 3065 | 66.5 (64.8, 68.2) |
| Whitaker et al | Europe | 84 | 28713 | 76155 | 37.7 (37.4, 38.0) |
| Yan et al | China | 98 | 0 | 125 | 0.0 (0.0, 2.9) |
| | | | | | **26.4 (2.6, 82.8)** |
| **10-99% hospitalised** | | | | | |
| Abdelrahman et al | Other | 270 | 105 | 172 | 61.0 (53.3, 68.4) |
| Aminian et al | N America | 243 | 1255 | 2839 | 44.2 (42.4, 46.1) |
| Becker et al | N America | 231 | 178 | 740 | 24.1 (21.0, 27.3) |
| Blomberg et al | Europe | 183 | 189 | 312 | 60.6 (54.9, 66.0) |
| Diaz-Fuentes et al | N America | 84 | 88 | 111 | 79.3 (70.5, 86.4) |
| Erol et al | Other | 170 | 45 | 121 | 37.2 (28.6, 46.4) |
| Jacobson et al | N America | 119 | 79 | 118 | 66.9 (57.7, 75.3) |
| Kim et al | Other | 195 | 591 | 900 | 65.7 (62.5, 68.8) |
| Logue et al | N America | 169 | 53 | 177 | 30.0 (23.3, 37.3) |
| Matteudi et al | Europe | 180 | 23 | 137 | 16.8 (11.0, 24.1) |
| Mazza et al | Europe | 104 | 81 | 226 | 35.8 (29.6, 42.5) |
| Menges et al | Europe | 219 | 106 | 431 | 24.6 (20.6, 28.9) |
| Millet et al | N America | 365 | 83 | 173 | 48.0 (40.3, 55.7) |
| Mohiuddin et al | Other | 161 | 67 | 313 | 21.4 (17.0, 26.4) |
| O'Keefe et al | N America | 119 | 79 | 198 | 39.9 (33.0, 47.1) |
| Peghin et al | Europe | 191 | 241 | 599 | 40.2 (36.3, 44.3) |
| Peluso et al | N America | 112 | 89 | 143 | 62.2 (53.8, 70.2) |
| Rass et al | Europe | 90 | 82 | 135 | 60.7 (52.0, 69.0) |
| Righi et al | Europe | 84 | 83 | 421 | 19.7 (16.0, 23.8) |
| Seeßle et al | Europe | 147 | 107 | 146 | 73.3 (65.3, 80.3) |
| Skala et al | Europe | 91 | 56 | 102 | 54.9 (44.7, 64.8) |
| Sonnweber et al | Europe | 103 | 73 | 133 | 54.9 (46.0, 63.5) |
| Sudre et al | Other | 84 | 108 | 4182 | 2.6 (2.1, 3.1) |
| Taquet et al | N America | 90 | 100007 | 273618 | 36.5 (36.4, 36.7) |
| Tawfik et al | Other | 91 | 40 | 120 | 33.3 (25.0, 42.5) |
| Townsend et al | Europe | 86 | 74 | 128 | 57.8 (48.8, 66.5) |
| Trunfio et al | Europe | 194 | 41 | 168 | 24.4 (18.1, 31.6) |
| Ursini et al | Europe | 182 | 270 | 616 | 43.8 (39.9, 47.9) |
| Zayet et al | Europe | 289 | 127 | 354 | 35.9 (30.9, 41.1) |
| | | | | | **41.1 (8.4, 84.2)** |
| **100% hospitalised** | | | | | |
| Al-Aly et al (hospitalized) | N America | 150 | 1091 | 11800 | 9.2 (8.7, 9.8) |
| Arnold et al | Europe | 90 | 81 | 110 | 73.6 (64.4, 81.6) |
| Ayoubkhani et al | Europe | 154 | 6085 | 28335 | 21.5 (21.0, 22.0) |
| Baricich et al | Europe | 139 | 66 | 204 | 32.4 (26.0, 39.2) |
| Bellan et al | Europe | 120 | 128 | 238 | 53.8 (47.2, 60.2) |
| Blanco et al | Europe | 104 | 52 | 100 | 52.0 (41.8, 62.1) |
| COMEBAC study | Europe | 127 | 244 | 478 | 51.0 (46.5, 55.6) |
| Carrillo-Garcia et al | Europe | 105 | 100 | 151 | 66.2 (58.1, 73.7) |
| Caruso et al | Europe | 182 | 91 | 118 | 77.1 (68.5, 84.3) |
| Castro et al | N America | 128 | 721 | 6619 | 10.9 (10.2, 11.7) |
| Chai et al | China | 378 | 156 | 546 | 28.6 (24.8, 32.6) |
| Clavario et al | Europe | 121 | 160 | 200 | 80.0 (73.8, 85.3) |
| Cristillo et al | Europe | 196 | 50 | 101 | 49.5 (39.4, 59.6) |
| Fernandez-de-Las-Penas et al | Europe | 227 | 930 | 1142 | 81.4 (79.1, 83.7) |
| Frija-Masson et al | Europe | 91 | 103 | 137 | 75.2 (67.1, 82.2) |
| Froidure et al | Europe | 95 | 73 | 107 | 68.2 (58.5, 76.9) |
| Fu et al | China | 196 | 20 | 199 | 10.1 (6.2, 15.1) |
| Garcia-Abellan et al | Europe | 182 | 28 | 116 | 24.1 (16.7, 33.0) |
| Gonzalez-Hermosillo et al | Other | 105 | 119 | 130 | 91.5 (85.4, 95.7) |
| Han et al | China | 175 | 71 | 114 | 62.3 (52.7, 71.2) |
| Huang et al | China | 186 | 1265 | 1655 | 76.4 (74.3, 78.5) |
| Li et al | China | 120 | 173 | 289 | 59.9 (54.0, 65.6) |
| Liao et al | China | 409 | 113 | 303 | 37.3 (31.8, 43.0) |
| Liao et al | China | 104 | 122 | 142 | 85.9 (79.1, 91.2) |
| Liu et al | China | 196 | 373 | 1301 | 28.7 (26.2, 31.2) |
| Liyanage-Don et al | N America | 113 | 99 | 153 | 64.7 (56.6, 72.3) |
| Lui et al | China | 96 | 41 | 204 | 20.1 (14.8, 26.3) |
| Maestre-Muniz et al | Europe | 379 | 309 | 543 | 56.9 (52.6, 61.1) |
| Mei et al | China | 158 | 976 | 3677 | 26.5 (25.1, 28.0) |
| Milanese et al | Europe | 196 | 64 | 135 | 47.4 (38.8, 56.2) |
| Munblit et al | Europe | 232 | 1534 | 2649 | 57.9 (56.0, 59.8) |
| Nabahati et al | Other | 104 | 90 | 173 | 52.0 (44.3, 59.7) |
| Nguyen et al | Europe | 213 | 30 | 125 | 24.0 (16.8, 32.5) |
| Nunez-Fernandez et al | Europe | 98 | 58 | 200 | 29.0 (22.8, 35.8) |
| Ong et al | Other | 104 | 13 | 175 | 7.4 (4.0, 12.4) |
| Osmanov et al | Europe | 270 | 126 | 519 | 24.3 (20.6, 28.2) |
| PHOSP-COVID study | Europe | 174 | 797 | 861 | 92.6 (90.6, 94.2) |
| Qin et al | China | 105 | 87 | 647 | 13.4 (10.9, 16.3) |
| Qu et al | China | 105 | 176 | 540 | 32.6 (28.7, 36.7) |
| Romero-Duarte et al | Europe | 196 | 509 | 797 | 63.9 (60.4, 67.2) |
| Sathyamurthy et al | Other | 104 | 66 | 279 | 23.7 (18.8, 29.1) |
| Shang et al | China | 196 | 441 | 796 | 55.4 (51.9, 58.9) |
| Sibila et al | Europe | 116 | 98 | 172 | 57.0 (49.2, 64.5) |
| Sigfrid et al | Europe | 222 | 305 | 327 | 93.3 (90.0, 95.7) |
| Simani et al | Other | 197 | 12 | 120 | 10.0 (5.3, 16.8) |
| Skjorten et al | Europe | 105 | 59 | 126 | 46.8 (37.9, 55.9) |
| Sykes et al | Europe | 127 | 75 | 127 | 59.1 (50.0, 67.7) |
| Taboada et al | Europe | 189 | 87 | 183 | 47.5 (40.1, 55.0) |
| Tarsitani et al | Europe | 105 | 34 | 115 | 29.6 (21.4, 38.8) |
| Taylor et al | Europe | 126 | 261 | 545 | 47.9 (43.6, 52.2) |
| Tleyjeh et al | Other | 136 | 125 | 222 | 56.3 (49.5, 62.9) |
| Todt et al | Other | 105 | 96 | 239 | 40.2 (33.9, 46.7) |
| Tohamy et al | Other | 105 | 5 | 100 | 5.0 (1.6, 11.3) |
| Venturelli et al | Europe | 105 | 394 | 767 | 51.4 (47.8, 55.0) |
| Walle-Hansen et al | Europe | 200 | 57 | 106 | 53.8 (43.8, 63.5) |
| Weng et al | China | 104 | 52 | 117 | 44.4 (35.3, 53.9) |
| Xiong et al | China | 111 | 267 | 538 | 49.6 (45.3, 53.9) |
| Xiong et al | China | 167 | 114 | 162 | 70.4 (62.7, 77.3) |
| Yan et al | China | 379 | 47 | 119 | 39.5 (30.7, 48.9) |
| Yin et al | China | 203 | 176 | 316 | 55.7 (50.0, 61.3) |
| Zhan et al | China | 348 | 36 | 121 | 29.8 (21.8, 38.7) |
| Zhang et al | China | 378 | 1095 | 2433 | 45.0 (43.0, 47.0) |
| Zhang et al | China | 106 | 67 | 122 | 54.9 (45.7, 63.9) |
| Zhang et al | China | 105 | 178 | 245 | 72.7 (66.6, 78.1) |
| Zhou et al | China | 127 | 114 | 164 | 69.5 (61.9, 76.5) |
| | | | | | **47.5 (8.3, 90.0)** |

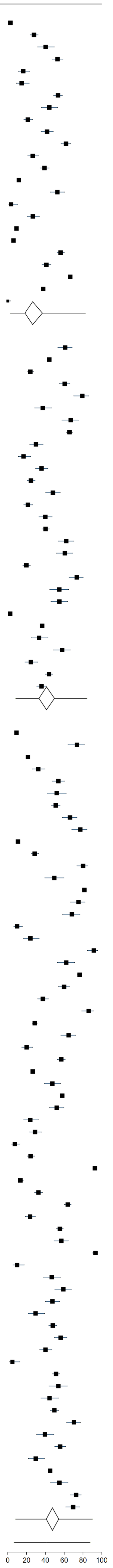

Supplementary Figure 3: Forest plot of prevalence of Long Covid in the included studies by percentage admitted to intensive care with 95% prediction intervals

| study | region | mean days since infection | cases | total | % persistent symptoms |
|---|---|---|---|---|---|
| **<5% ICU** | | | | | |
| Abdelrahman et al | Other | 270 | 105 | 172 | 61.0 (53.3, 68.4) |
| Al-Aly et al (non-hospitalized) | N America | 126 | 1718 | 60255 | 2.9 (2.7, 3.0) |
| Aminian et al | N America | 243 | 1255 | 2839 | 44.2 (42.4, 46.1) |
| Augustin et al | Europe | 131 | 123 | 442 | 27.8 (23.7, 32.3) |
| Bliddal et al | Europe | 84 | 52 | 129 | 40.3 (31.8, 49.3) |
| Blomberg et al | Europe | 183 | 189 | 312 | 60.6 (54.9, 66.0) |
| Boscolo-Rizzo et al | Europe | 365 | 161 | 304 | 53.0 (47.2, 58.7) |
| Caspersen et al | Europe | 350 | 28 | 170 | 16.5 (11.2, 22.9) |
| Chai et al | China | 378 | 156 | 546 | 28.6 (24.8, 32.6) |
| Cirulli et al | N America | 90 | 18 | 122 | 14.8 (9.0, 22.3) |
| Cristillo et al | Europe | 196 | 50 | 101 | 49.5 (39.4, 59.6) |
| Domenech-Montoliu et al | Europe | 213 | 258 | 483 | 53.4 (48.9, 57.9) |
| Fu et al | China | 196 | 20 | 199 | 10.1 (6.2, 15.1) |
| Gaber et al | Europe | 122 | 61 | 138 | 44.2 (35.8, 52.9) |
| Havervall et al | Europe | 122 | 69 | 323 | 21.4 (17.0, 26.2) |
| Huang et al | China | 186 | 1265 | 1655 | 76.4 (74.3, 78.5) |
| Kashif et al | Other | 105 | 101 | 242 | 41.7 (35.5, 48.2) |
| Lemhofer et al | Europe | 91 | 226 | 365 | 61.9 (56.7, 66.9) |
| Liao et al | China | 104 | 122 | 142 | 85.9 (79.1, 91.2) |
| Lui et al | China | 96 | 41 | 204 | 20.1 (14.8, 26.3) |
| Martinez et al | Europe | 168 | 69 | 260 | 26.5 (21.3, 32.3) |
| Matteudi et al | Europe | 180 | 23 | 137 | 16.8 (11.0, 24.1) |
| Menges et al | Europe | 219 | 106 | 431 | 24.6 (20.6, 28.9) |
| Mohiuddin et al | Other | 161 | 67 | 313 | 21.4 (17.0, 26.4) |
| Munblit et al | Europe | 232 | 1534 | 2649 | 57.9 (56.0, 59.8) |
| Nehme et al | Europe | 243 | 160 | 410 | 39.0 (34.3, 43.9) |
| Nguyen et al | Europe | 213 | 30 | 125 | 24.0 (16.8, 32.5) |
| ONS study August 2021 | Europe | 84 | 2501 | 21374 | 11.7 (11.3, 12.1) |
| Osmanov et al | Europe | 270 | 126 | 519 | 24.3 (20.6, 28.2) |
| Peghin et al | Europe | 191 | 241 | 599 | 40.2 (36.3, 44.3) |
| Peluso et al | N America | 112 | 89 | 143 | 62.2 (53.8, 70.2) |
| Petersen et al | Europe | 125 | 95 | 180 | 52.8 (45.2, 60.2) |
| Radtke et al | Europe | 84 | 4 | 109 | 3.7 (1.0, 9.1) |
| Riestra-Ayora et al | Europe | 182 | 52 | 195 | 26.7 (20.6, 33.5) |
| Roessler et al (adults) | Europe | 91 | 13396 | 145184 | 9.2 (9.1, 9.4) |
| Roessler et al (children) | Europe | 91 | 734 | 11950 | 6.1 (5.7, 6.6) |
| Seeßle et al | Europe | 147 | 107 | 146 | 73.3 (65.3, 80.3) |
| Shang et al | China | 196 | 441 | 796 | 55.4 (51.9, 58.9) |
| Sibila et al | Europe | 116 | 98 | 172 | 57.0 (49.2, 64.5) |
| Soraas et al | Europe | 126 | 380 | 676 | 56.2 (52.4, 60.0) |
| Stavem et al | Europe | 117 | 185 | 451 | 41.0 (36.4, 45.7) |
| Stephenson et al | Europe | 104 | 2038 | 3065 | 66.5 (64.8, 68.2) |
| Taquet et al | N America | 90 | 100007 | 273618 | 36.5 (36.4, 36.7) |
| Ursini et al | Europe | 182 | 270 | 616 | 43.8 (39.9, 47.9) |
| Whitaker et al | Europe | 84 | 28713 | 76155 | 37.7 (37.4, 38.0) |
| Yan et al | China | 98 | 0 | 125 | 0.0 (0.0, 2.9) |
| Yin et al | China | 203 | 176 | 316 | 55.7 (50.0, 61.3) |
| Zhang et al | China | 378 | 1095 | 2433 | 45.0 (43.0, 47.0) |
| | | | | | **34.9 (5.2, 84.0)** |
| **5-9.9% ICU** | | | | | |
| Ayoubkhani et al | Europe | 154 | 6085 | 28335 | 21.5 (21.0, 22.0) |
| Diaz-Fuentes et al | N America | 84 | 88 | 111 | 79.3 (70.5, 86.4) |
| Fernandez-de-Las-Penas et al | Europe | 227 | 930 | 1142 | 81.4 (79.1, 83.7) |
| Jacobson et al | N America | 119 | 79 | 118 | 66.9 (57.7, 75.3) |
| Liu et al | China | 196 | 373 | 1301 | 28.7 (26.2, 31.2) |
| Liyanage-Don et al | N America | 113 | 99 | 153 | 64.7 (56.6, 72.3) |
| Logue et al | N America | 169 | 53 | 177 | 30.0 (23.3, 37.3) |
| Mei et al | China | 158 | 976 | 3677 | 26.5 (25.1, 28.0) |
| Simani et al | Other | 197 | 12 | 120 | 10.0 (5.3, 16.8) |
| Skala et al | Europe | 91 | 56 | 102 | 54.9 (44.7, 64.8) |
| Sudre et al | Other | 84 | 108 | 4182 | 2.6 (2.1, 3.1) |
| Venturelli et al | Europe | 105 | 394 | 767 | 51.4 (47.8, 55.0) |
| Xiong et al | China | 167 | 114 | 162 | 70.4 (62.7, 77.3) |
| Zayet et al | Europe | 289 | 127 | 354 | 35.9 (30.9, 41.1) |
| Zhan et al | China | 348 | 36 | 121 | 29.8 (21.8, 38.7) |
| Zhou et al | China | 127 | 114 | 164 | 69.5 (61.9, 76.5) |
| | | | | | **41.8 (3.5, 93.5)** |
| **10+% ICU** | | | | | |
| Al-Aly et al (hospitalized) | N America | 150 | 1091 | 11800 | 9.2 (8.7, 9.8) |
| Baricich et al | Europe | 139 | 66 | 204 | 32.4 (26.0, 39.2) |
| Bellan et al | Europe | 120 | 128 | 238 | 53.8 (47.2, 60.2) |
| COMEBAC study | Europe | 127 | 244 | 478 | 51.0 (46.5, 55.6) |
| Caruso et al | Europe | 182 | 91 | 118 | 77.1 (68.5, 84.3) |
| Castro et al | N America | 128 | 721 | 6619 | 10.9 (10.2, 11.7) |
| Clavario et al | Europe | 121 | 160 | 200 | 80.0 (73.8, 85.3) |
| Frija-Masson et al | Europe | 91 | 103 | 137 | 75.2 (67.1, 82.2) |
| Froidure et al | Europe | 95 | 73 | 107 | 68.2 (58.5, 76.9) |
| Garcia-Abellan et al | Europe | 182 | 28 | 116 | 24.1 (16.7, 33.0) |
| Gonzalez-Hermosillo et al | Other | 105 | 119 | 130 | 91.5 (85.4, 95.7) |
| Nunez-Fernandez et al | Europe | 98 | 58 | 200 | 29.0 (22.8, 35.8) |
| Ong et al | Other | 104 | 13 | 175 | 7.4 (4.0, 12.4) |
| PHOSP-COVID study | Europe | 174 | 797 | 861 | 92.6 (90.6, 94.2) |
| Rass et al | Europe | 90 | 82 | 135 | 60.7 (52.0, 69.0) |
| Righi et al | Europe | 84 | 83 | 421 | 19.7 (16.0, 23.8) |
| Romero-Duarte et al | Europe | 196 | 509 | 797 | 63.9 (60.4, 67.2) |
| Sigfrid et al | Europe | 222 | 305 | 327 | 93.3 (90.0, 95.7) |
| Skjorten et al | Europe | 105 | 59 | 126 | 46.8 (37.9, 55.9) |
| Sonnweber et al | Europe | 103 | 73 | 133 | 54.9 (46.0, 63.5) |
| Sykes et al | Europe | 127 | 75 | 127 | 59.1 (50.0, 67.7) |
| Taboada et al | Europe | 189 | 87 | 183 | 47.5 (40.1, 55.0) |
| Tarsitani et al | Europe | 105 | 34 | 115 | 29.6 (21.4, 38.8) |
| Tawfik et al | Other | 91 | 40 | 120 | 33.3 (25.0, 42.5) |
| Tleyjeh et al | Other | 136 | 125 | 222 | 56.3 (49.5, 62.9) |
| Todt et al | Other | 105 | 96 | 239 | 40.2 (33.9, 46.7) |
| Tohamy et al | Other | 105 | 5 | 100 | 5.0 (1.6, 11.3) |
| Townsend et al | Europe | 86 | 74 | 128 | 57.8 (48.8, 66.5) |
| Walle-Hansen et al | Europe | 200 | 57 | 106 | 53.8 (43.8, 63.5) |
| Weng et al | China | 104 | 52 | 117 | 44.4 (35.3, 53.9) |
| Zhang et al | China | 106 | 67 | 122 | 54.9 (45.7, 63.9) |
| | | | | | **48.8 (5.7, 93.7)** |

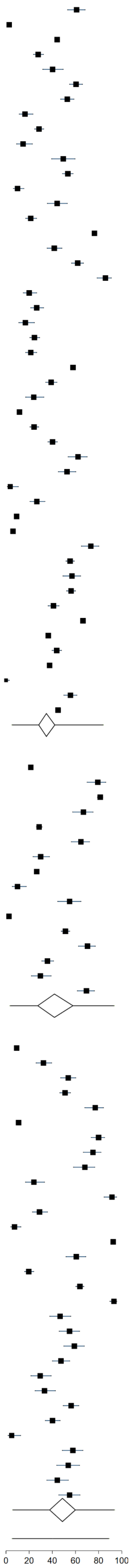

Supplementary Figure 4: Forest plot of prevalence of Long Covid in the included studies by severity of acute infection using the WHO Clinical Progression Scale with 95% prediction intervals

| study | region | mean days since infection | cases | total | % persistent symptoms |
|---|---|---|---|---|---|
| **Ambulatory mild disease: symptomatic, independent** | | | | | |
| Bliddal et al | Europe | 84 | 52 | 129 | 40.3 (31.8, 49.3) |
| Caspersen et al | Europe | 350 | 28 | 170 | 16.5 (11.2, 22.9) |
| Cirulli et al | N America | 90 | 18 | 122 | 14.8 (9.0, 22.3) |
| Domenech-Montoliu et al | Europe | 213 | 258 | 483 | 53.4 (48.9, 57.9) |
| Gaber et al | Europe | 122 | 61 | 138 | 44.2 (35.8, 52.9) |
| Lemhofer et al | Europe | 91 | 226 | 365 | 61.9 (56.7, 66.9) |
| Martinez et al | Europe | 168 | 69 | 260 | 26.5 (21.3, 32.3) |
| Matteudi et al | Europe | 180 | 23 | 137 | 16.8 (11.0, 24.1) |
| ONS study August 2021 | Europe | 84 | 2501 | 21374 | 11.7 (11.3, 12.1) |
| Roessler et al (adults) | Europe | 91 | 13396 | 145184 | 9.2 (9.1, 9.4) |
| Roessler et al (children) | Europe | 91 | 734 | 11950 | 6.1 (5.7, 6.6) |
| Stavem et al | Europe | 117 | 185 | 451 | 41.0 (36.4, 45.7) |
| Stephenson et al | Europe | 104 | 2038 | 3065 | 66.5 (64.8, 68.2) |
| Sudre et al | Other | 84 | 108 | 4182 | 2.6 (2.1, 3.1) |
| Tempany et al | Europe | 84 | 116 | 217 | 53.5 (46.6, 60.2) |
| Whitaker et al | Europe | 84 | 28713 | 76155 | 37.7 (37.4, 38.0) |
| Yan et al | China | 98 | 0 | 125 | 0.0 (0.0, 2.9) |
| | | | | | **23.5 (1.6, 85.7)** |
| **Ambulatory mild disease: symptomatic, assistance sought** | | | | | |
| Abdelrahman et al | Other | 270 | 105 | 172 | 61.0 (53.3, 68.4) |
| Al-Aly et al (non-hospitalized) | N America | 126 | 1718 | 60255 | 2.9 (2.7, 3.0) |
| Augustin et al | Europe | 131 | 123 | 442 | 27.8 (23.7, 32.3) |
| Blomberg et al | Europe | 183 | 189 | 312 | 60.6 (54.9, 66.0) |
| Boscolo-Rizzo et al | Europe | 365 | 161 | 304 | 53.0 (47.2, 58.7) |
| Havervall et al | Europe | 122 | 69 | 323 | 21.4 (17.0, 26.2) |
| Kashif et al | Other | 105 | 101 | 242 | 41.7 (35.5, 48.2) |
| Menges et al | Europe | 219 | 106 | 431 | 24.6 (20.6, 28.9) |
| Mohiuddin et al | Other | 161 | 67 | 313 | 21.4 (17.0, 26.4) |
| Nehme et al | Europe | 243 | 160 | 410 | 39.0 (34.3, 43.9) |
| O'Keefe et al | N America | 119 | 79 | 198 | 39.9 (33.0, 47.1) |
| Peghin et al | Europe | 191 | 241 | 599 | 40.2 (36.3, 44.3) |
| Peluso et al | N America | 112 | 89 | 143 | 62.2 (53.8, 70.2) |
| Petersen et al | Europe | 125 | 95 | 180 | 52.8 (45.2, 60.2) |
| Radtke et al | Europe | 84 | 4 | 109 | 3.7 (1.0, 9.1) |
| Riestra-Ayora et al | Europe | 182 | 52 | 195 | 26.7 (20.6, 33.5) |
| Skala et al | Europe | 91 | 56 | 102 | 54.9 (44.7, 64.8) |
| Soraas et al | Europe | 126 | 380 | 676 | 56.2 (52.4, 60.0) |
| Taquet et al | N America | 90 | 100007 | 273618 | 36.5 (36.4, 36.7) |
| Tawfik et al | Other | 91 | 40 | 120 | 33.3 (25.0, 42.5) |
| Ursini et al | Europe | 182 | 270 | 616 | 43.8 (39.9, 47.9) |
| | | | | | **35.1 (5.8, 82.5)** |
| **Hospitalised moderate disease: no oxygen therapy** | | | | | |
| Aminian et al | N America | 243 | 1255 | 2839 | 44.2 (42.4, 46.1) |
| Becker et al | N America | 231 | 178 | 740 | 24.1 (21.0, 27.3) |
| Diaz-Fuentes et al | N America | 84 | 88 | 111 | 79.3 (70.5, 86.4) |
| Fu et al | China | 196 | 20 | 199 | 10.1 (6.2, 15.1) |
| Garcia-Abellan et al | Europe | 182 | 28 | 116 | 24.1 (16.7, 33.0) |
| Jacobson et al | N America | 119 | 79 | 118 | 66.9 (57.7, 75.3) |
| Kim et al | Other | 195 | 591 | 900 | 65.7 (62.5, 68.8) |
| Liao et al | China | 104 | 122 | 142 | 85.9 (79.1, 91.2) |
| Logue et al | N America | 169 | 53 | 177 | 30.0 (23.3, 37.3) |
| Lui et al | China | 96 | 41 | 204 | 20.1 (14.8, 26.3) |
| Maestre-Muniz et al | Europe | 379 | 309 | 543 | 56.9 (52.6, 61.1) |
| Mechi et al | Other | 274 | 92 | 112 | 82.1 (73.8, 88.7) |
| Munblit et al | Europe | 232 | 1534 | 2649 | 57.9 (56.0, 59.8) |
| Townsend et al | Europe | 86 | 74 | 128 | 57.8 (48.8, 66.5) |
| Trunfio et al | Europe | 194 | 41 | 168 | 24.4 (18.1, 31.6) |
| Zayet et al | Europe | 289 | 127 | 354 | 35.9 (30.9, 41.1) |
| Zhan et al | China | 348 | 36 | 121 | 29.8 (21.8, 38.7) |
| | | | | | **46.1 (7.0, 90.6)** |
| **Hospitalised moderate disease: oxygen by mask or nasal prongs** | | | | | |
| Arnold et al | Europe | 90 | 81 | 110 | 73.6 (64.4, 81.6) |
| Baricich et al | Europe | 139 | 66 | 204 | 32.4 (26.0, 39.2) |
| Bellan et al | Europe | 120 | 128 | 238 | 53.8 (47.2, 60.2) |
| Castro et al | N America | 128 | 721 | 6619 | 10.9 (10.2, 11.7) |
| Chai et al | China | 378 | 156 | 546 | 28.6 (24.8, 32.6) |
| Cristillo et al | Europe | 196 | 50 | 101 | 49.5 (39.4, 59.6) |
| Fernandez-de-Las-Penas et al | Europe | 227 | 930 | 1142 | 81.4 (79.1, 83.7) |
| Han et al | China | 175 | 71 | 114 | 62.3 (52.7, 71.2) |
| Huang et al | China | 186 | 1265 | 1655 | 76.4 (74.3, 78.5) |
| Li et al | China | 120 | 173 | 289 | 59.9 (54.0, 65.6) |
| Liao et al | China | 409 | 113 | 303 | 37.3 (31.8, 43.0) |
| Liu et al | China | 196 | 373 | 1301 | 28.7 (26.2, 31.2) |
| Mazza et al | Europe | 104 | 81 | 226 | 35.8 (29.6, 42.5) |
| Mei et al | China | 158 | 976 | 3677 | 26.5 (25.1, 28.0) |
| Millet et al | N America | 365 | 83 | 173 | 48.0 (40.3, 55.7) |
| Nguyen et al | Europe | 213 | 30 | 125 | 24.0 (16.8, 32.5) |
| Nunez-Fernandez et al | Europe | 98 | 58 | 200 | 29.0 (22.8, 35.8) |
| Ong et al | Other | 104 | 13 | 175 | 7.4 (4.0, 12.4) |
| Osmanov et al | Europe | 270 | 126 | 519 | 24.3 (20.6, 28.2) |
| Qu et al | China | 105 | 176 | 540 | 32.6 (28.7, 36.7) |
| Rass et al | Europe | 90 | 82 | 135 | 60.7 (52.0, 69.0) |
| Seeßle et al | Europe | 147 | 107 | 146 | 73.3 (65.3, 80.3) |
| Sonnweber et al | Europe | 103 | 73 | 133 | 54.9 (46.0, 63.5) |
| Sykes et al | Europe | 127 | 75 | 127 | 59.1 (50.0, 67.7) |
| Taboada et al | Europe | 189 | 87 | 183 | 47.5 (40.1, 55.0) |
| Tarsitani et al | Europe | 105 | 34 | 115 | 29.6 (21.4, 38.8) |
| Taylor et al | Europe | 126 | 261 | 545 | 47.9 (43.6, 52.2) |
| Todt et al | Other | 105 | 96 | 239 | 40.2 (33.9, 46.7) |
| Tohamy et al | Other | 105 | 5 | 100 | 5.0 (1.6, 11.3) |
| Walle-Hansen et al | Europe | 200 | 57 | 106 | 53.8 (43.8, 63.5) |
| Weng et al | China | 104 | 52 | 117 | 44.4 (35.3, 53.9) |
| Xiong et al | China | 167 | 114 | 162 | 70.4 (62.7, 77.3) |
| Yan et al | China | 379 | 47 | 119 | 39.5 (30.7, 48.9) |
| Yin et al | China | 203 | 176 | 316 | 55.7 (50.0, 61.3) |
| Zhang et al | China | 106 | 67 | 122 | 54.9 (45.7, 63.9) |
| Zhang et al | China | 378 | 1095 | 2433 | 45.0 (43.0, 47.0) |
| Zhou et al | China | 127 | 114 | 164 | 69.5 (61.9, 76.5) |
| | | | | | **44.0 (10.7, 83.7)** |
| **Hospitalised severe disease: oxygen by NIV or high flow** | | | | | |
| Al-Aly et al (hospitalized) | N America | 150 | 1091 | 11800 | 9.2 (8.7, 9.8) |
| Ayoubkhani et al | Europe | 154 | 6085 | 28335 | 21.5 (21.0, 22.0) |
| Blanco et al | Europe | 104 | 52 | 100 | 52.0 (41.8, 62.1) |
| COMEBAC study | Europe | 127 | 244 | 478 | 51.0 (46.5, 55.6) |
| Carrillo-Garcia et al | Europe | 105 | 100 | 151 | 66.2 (58.1, 73.7) |
| Caruso et al | Europe | 182 | 91 | 118 | 77.1 (68.5, 84.3) |
| Clavario et al | Europe | 121 | 160 | 200 | 80.0 (73.8, 85.3) |
| Frija-Masson et al | Europe | 91 | 103 | 137 | 75.2 (67.1, 82.2) |
| Froidure et al | Europe | 95 | 73 | 107 | 68.2 (58.5, 76.9) |
| Gonzalez-Hermosillo et al | Other | 105 | 119 | 130 | 91.5 (85.4, 95.7) |
| Liyanage-Don et al | N America | 113 | 99 | 153 | 64.7 (56.6, 72.3) |
| Milanese et al | Europe | 196 | 64 | 135 | 47.4 (38.8, 56.2) |
| Nabahati et al | Other | 104 | 90 | 173 | 52.0 (44.3, 59.7) |
| PHOSP-COVID study | Europe | 174 | 797 | 861 | 92.6 (90.6, 94.2) |
| Qin et al | China | 105 | 87 | 647 | 13.4 (10.9, 16.3) |
| Righi et al | Europe | 84 | 83 | 421 | 19.7 (16.0, 23.8) |
| Romero-Duarte et al | Europe | 196 | 509 | 797 | 63.9 (60.4, 67.2) |
| Sathyamurthy et al | Other | 104 | 66 | 279 | 23.7 (18.8, 29.1) |
| Shang et al | China | 196 | 441 | 796 | 55.4 (51.9, 58.9) |
| Sibila et al | Europe | 116 | 98 | 172 | 57.0 (49.2, 64.5) |
| Sigfrid et al | Europe | 222 | 305 | 327 | 93.3 (90.0, 95.7) |
| Simani et al | Other | 197 | 12 | 120 | 10.0 (5.3, 16.8) |
| Skjorten et al | Europe | 105 | 59 | 126 | 46.8 (37.9, 55.9) |
| Tleyjeh et al | Other | 136 | 125 | 222 | 56.3 (49.5, 62.9) |
| Venturelli et al | Europe | 105 | 394 | 767 | 51.4 (47.8, 55.0) |
| Xiong et al | China | 111 | 267 | 538 | 49.6 (45.3, 53.9) |
| Zhang et al | China | 105 | 178 | 245 | 72.7 (66.6, 78.1) |
| | | | | | **54.8 (7.7, 94.7)** |

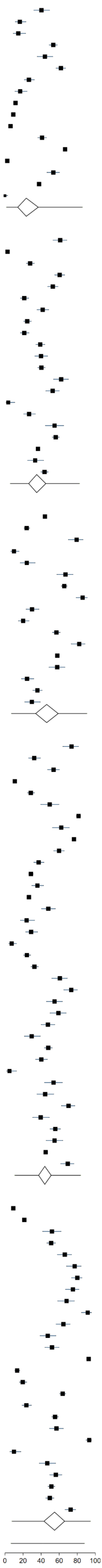

Supplementary Figure 5: Forest plot of prevalence of Long Covid in studies that reported outcome of not returned to full health/fitness with 95% prediction intervals

| study | region | mean days since infection | % Persistent Not Returned to Full Health/Fitness | |
|---|---|---|---|---|
| Domenech-Montoliu et al | Europe | 213 | 18.0 | (14.7, 21.7) |
| Martinez et al | Europe | 168 | 26.5 | (21.3, 32.3) |
| ONS study August 2021 | Europe | 84 | 7.5 | (7.2, 7.9) |
| PHOSP-COVID study | Europe | 166 | 54.9 | (52.7, 57.1) |
| PHOSP-COVID study | Europe | 174 | 51.7 | (48.2, 55.1) |
| Soraas et al | Europe | 258 | 20.0 | (17.0, 23.3) |
| Taboada et al | Europe | 189 | 55.7 | (48.2, 63.1) |
| Tleyjeh et al | Other | 136 | 28.8 | (23.0, 35.3) |
| Todt et al | Other | 105 | 40.1 | (33.9, 46.7) |
| Xiong et al | China | 167 | 70.4 | (62.7, 77.3) |
| **95% Prediction Interval** | | | | |

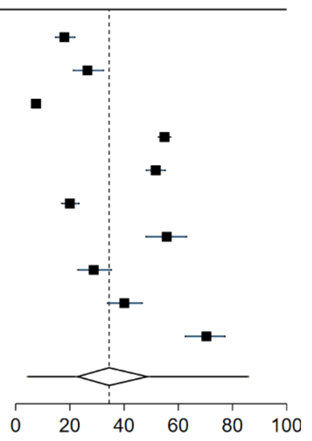

Supplementary Figure 6: Forest plot of prevalence of Long Covid in studies that reported lower quality of life with 95% prediction intervals

| study | region | mean days since infection | % Persistent Lower QoL |
|---|---|---|---|
| Logue et al | N America | 169 | 30.7 (23.8, 38.3) |
| Rass et al | Europe | 90 | 31.0 (21.8, 41.7) |
| **95% Prediction Interval** | | | |

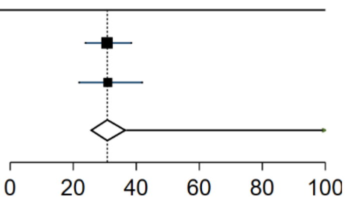

Supplementary Figure 7: Forest plot of prevalence of Long Covid in studies that reported abdominal pain as a persistent symptom with 95% prediction intervals

| study | region | mean days since infection | % Persistent Abdominal Pain | |
|---|---|---|---|---|
| Al-Aly et al (hospitalized) | N America | 150 | 0.1 | (0.0, 0.1) |
| Al-Aly et al (non-hospitalized) | N America | 126 | 0.6 | (0.5, 0.6) |
| Boscolo-Rizzo et al | Europe | 365 | 0.3 | (0.0, 1.7) |
| Domenech-Montoliu et al | Europe | 213 | 4.1 | (2.5, 6.3) |
| Gonzalez-Hermosillo et al | Other | 105 | 15.4 | (9.7, 22.8) |
| Munblit et al | Europe | 232 | 3.9 | (3.2, 4.7) |
| Orru et al | Europe | 91 | 23.0 | (16.6, 30.5) |
| Osmanov et al | Europe | 270 | 2.0 | (1.0, 3.6) |
| PHOSP-COVID study | Europe | 174 | 17.1 | (14.3, 20.1) |
| Radtke et al | Europe | 84 | 0.9 | (0.0, 5.0) |
| Sigfrid et al | Europe | 222 | 17.7 | (13.7, 22.3) |
| Stavem et al | Europe | 117 | 3.2 | (1.8, 5.3) |
| Ursini et al | Europe | 182 | 17.5 | (14.6, 20.7) |
| Weng et al | China | 104 | 6.8 | (3.0, 13.0) |
| Zayet et al | Europe | 289 | 2.3 | (1.0, 4.5) |
| **95% Prediction Interval** | | | | |

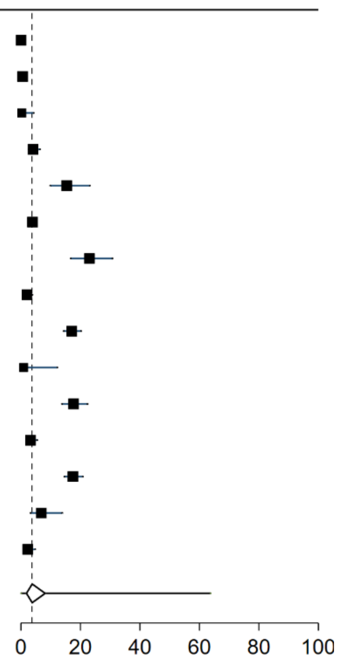

Supplementary Figure 8: Forest plot of prevalence of Long Covid in studies that reported muscle or joint aches/pains as a persistent symptom with 95% prediction intervals

| study | region | mean days since infection | % Persistent Aches or Pains In Joints or Muscles | |
|---|---|---|---|---|
| Abdelrahman et al | Other | 270 | 12.2 | (7.7, 18.1) |
| Al-Aly et al (hospitalized) | N America | 150 | 2.3 | (2.0, 2.6) |
| Al-Aly et al (non-hospitalized) | N America | 126 | 1.4 | (1.3, 1.5) |
| Bellan et al | Europe | 120 | 5.9 | (3.3, 9.7) |
| Bliddal et al | Europe | 84 | 5.5 | (2.2, 11.0) |
| Boscolo-Rizzo et al | Europe | 365 | 9.2 | (6.2, 13.0) |
| Caspersen et al | Europe | 350 | 2.3 | (1.4, 3.6) |
| Cirulli et al | N America | 90 | 13.0 | (7.5, 20.6) |
| Cristillo et al | Europe | 196 | 25.7 | (17.6, 35.4) |
| Diaz-Fuentes et al | N America | 84 | 4.5 | (1.5, 10.2) |
| Domenech-Montoliu et al | Europe | 213 | 11.6 | (8.9, 14.8) |
| Erol et al | Other | 170 | 19.0 | (12.4, 27.1) |
| Garcia-Abellan et al | Europe | 182 | 6.9 | (3.0, 13.1) |
| Gonzalez-Hermosillo et al | Other | 105 | 40.8 | (32.2, 49.7) |
| Havervall et al | Europe | 122 | 1.5 | (0.5, 3.5) |
| Huang et al | China | 186 | 9.0 | (7.7, 10.5) |
| Jacobson et al | N America | 119 | 17.9 | (11.5, 26.1) |
| Kashif et al | Other | 105 | 35.1 | (29.1, 41.5) |
| Lemhofer et al | Europe | 91 | 26.0 | (21.6, 30.8) |
| Liyanage-Don et al | N America | 113 | 23.5 | (17.1, 31.1) |
| Logue et al | N America | 169 | 5.6 | (2.7, 10.0) |
| Maestre-Muniz et al | Europe | 379 | 11.4 | (8.9, 14.4) |
| Matteudi et al | Europe | 180 | 9.5 | (5.1, 15.7) |
| Mechi et al | Other | 274 | 27.7 | (19.6, 36.9) |
| Mei et al | China | 158 | 1.9 | (1.5, 2.4) |
| Menges et al | Europe | 219 | 35.3 | (30.8, 40.0) |
| Millet et al | N America | 365 | 18.5 | (13.0, 25.1) |
| Mohiuddin et al | Other | 161 | 0.6 | (0.1, 2.2) |
| Munblit et al | Europe | 232 | 6.7 | (5.8, 7.7) |
| Nehme et al | Europe | 243 | 6.3 | (4.2, 9.2) |
| O'Keefe et al | N America | 119 | 8.6 | (5.1, 13.4) |
| Ong et al | Other | 104 | 1.1 | (0.1, 3.9) |
| Orru et al | Europe | 91 | 61.1 | (53.0, 69.0) |
| Osmanov et al | Europe | 270 | 1.2 | (0.4, 2.6) |
| PHOSP-COVID study | Europe | 174 | 57.2 | (53.4, 61.0) |
| Peluso et al | N America | 112 | 35.0 | (26.5, 44.4) |
| Petersen et al | Europe | 125 | 11.0 | (6.8, 16.5) |
| Rass et al | Europe | 90 | 11.0 | (6.2, 17.8) |
| Roessler et al (adults) | Europe | 91 | 3.4 | (3.3, 3.5) |
| Roessler et al (children) | Europe | 91 | 1.9 | (1.6, 2.1) |
| Romero-Duarte et al | Europe | 196 | 15.3 | (12.9, 18.0) |
| Sathyamurthy et al | Other | 104 | 1.1 | (0.2, 3.2) |
| Seeßle et al | Europe | 147 | 16.4 | (10.8, 23.5) |
| Shang et al | China | 196 | 13.8 | (11.5, 16.4) |
| Sibila et al | Europe | 116 | 4.7 | (2.1, 9.1) |
| Sigfrid et al | Europe | 222 | 37.0 | (31.8, 42.5) |
| Skala et al | Europe | 91 | 3.9 | (1.1, 9.6) |
| Sonnweber et al | Europe | 103 | 24.0 | (17.1, 32.2) |
| Soraas et al | Europe | 258 | 13.0 | (10.5, 15.8) |
| Stavem et al | Europe | 117 | 8.6 | (6.2, 11.6) |
| Stephenson et al | Europe | 104 | 5.4 | (4.6, 6.3) |
| Sykes et al | Europe | 127 | 49.6 | (40.6, 58.6) |
| Taquet et al | N America | 90 | 7.2 | (7.1, 7.3) |
| Tawfik et al | Other | 91 | 18.3 | (11.9, 26.4) |
| Tleyjeh et al | Other | 136 | 9.0 | (5.6, 13.6) |
| Todt et al | Other | 105 | 39.5 | (33.3, 46.0) |
| Ursini et al | Europe | 182 | 30.7 | (27.1, 34.5) |
| Venturelli et al | Europe | 105 | 3.8 | (2.5, 5.4) |
| Walle-Hansen et al | Europe | 200 | 33.0 | (24.2, 42.8) |
| Zayet et al | Europe | 289 | 9.3 | (6.5, 12.8) |
| Zhang et al | China | 378 | 7.9 | (6.9, 9.0) |
| **95% Prediction Interval** | | | | |

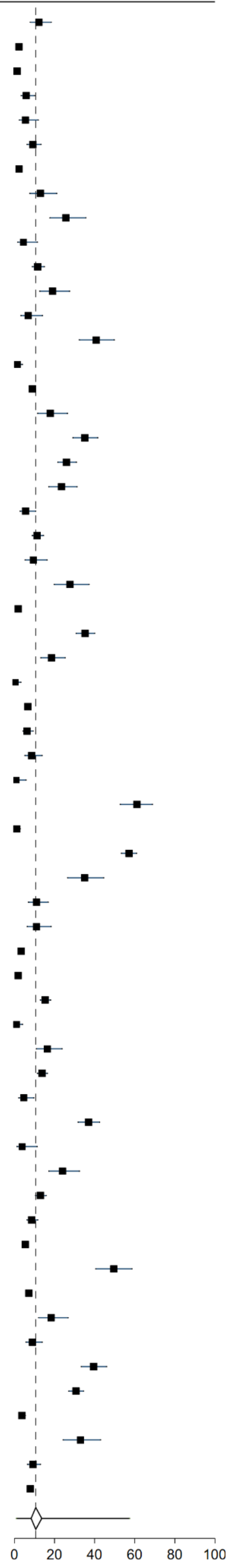

Supplementary Figure 9: Forest plot of prevalence of Long Covid in studies that reported alopecia as a persistent symptom with 95% prediction intervals

| study | region | mean days since infection | % Persistent Alopecia |
|---|---|---|---|
| Abdelrahman et al | Other | 270 | 10.5 (6.4, 16.1) |
| Caruso et al | Europe | 182 | 15.3 (9.3, 23.0) |
| Domenech-Montoliu et al | Europe | 213 | 24.6 (20.8, 28.7) |
| Fernandez-de-Las-Penas et al | Europe | 227 | 26.3 (23.8, 28.9) |
| Huang et al | China | 186 | 22.0 (20.0, 24.1) |
| Jacobson et al | N America | 119 | 12.0 (6.7, 19.3) |
| Maestre-Muniz et al | Europe | 379 | 1.7 (0.8, 3.2) |
| Mei et al | China | 158 | 1.8 (1.4, 2.3) |
| Munblit et al | Europe | 232 | 10.0 (8.9, 11.2) |
| Nehme et al | Europe | 243 | 3.2 (1.7, 5.4) |
| Osmanov et al | Europe | 270 | 1.8 (0.8, 3.4) |
| Peghin et al | Europe | 191 | 3.7 (2.3, 5.5) |
| Romero-Duarte et al | Europe | 196 | 3.0 (1.9, 4.4) |
| Shang et al | China | 196 | 0.5 (0.1, 1.3) |
| Tawfik et al | Other | 91 | 3.3 (0.9, 8.2) |
| Ursini et al | Europe | 182 | 25.3 (21.9, 28.9) |
| Xiong et al | China | 111 | 28.6 (24.8, 32.6) |
| **95% Prediction Interval** | | | |

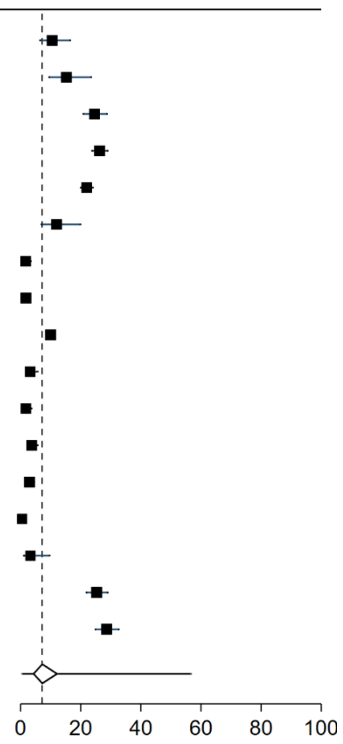

Supplementary Figure 10: Forest plot of prevalence of Long Covid in studies that reported anxiety, depression or mood change as a persistent symptom with 95% prediction intervals

| study | region | mean days since infection | % Persistent Anxiety, Depression or Mood Change | |
|---|---|---|---|---|
| Caspersen et al | Europe | 350 | 1.2 | (0.6, 2.3) |
| Munblit et al | Europe | 232 | 4.0 | (3.3, 4.8) |
| PHOSP-COVID study | Europe | 174 | 20.8 | (17.8, 24.1) |
| Romero-Duarte et al | Europe | 196 | 4.4 | (3.1, 6.1) |
| Sykes et al | Europe | 127 | 38.6 | (30.1, 47.6) |
| **95% Prediction Interval** | | | | |

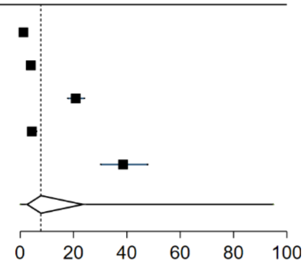

Supplementary Figure 11: Forest plot of prevalence of Long Covid in studies that reported breathing problems as a persistent symptom with 95% prediction intervals

| study | region | mean days since infection | % Persistent Breathing Problems | |
|---|---|---|---|---|
| Abdelrahman et al | Other | 270 | 22.0 | (16.0, 28.9) |
| Al-Aly et al (hospitalized) | N America | 150 | 1.2 | (1.1, 1.5) |
| Al-Aly et al (non-hospitalized) | N America | 126 | 2.8 | (2.7, 3.0) |
| Arnold et al | Europe | 90 | 39.0 | (29.9, 48.9) |
| Augustin et al | Europe | 131 | 8.6 | (6.2, 11.6) |
| Bellan et al | Europe | 120 | 5.5 | (3.0, 9.2) |
| Bliddal et al | Europe | 84 | 10.0 | (5.6, 16.2) |
| Blomberg et al | Europe | 183 | 15.0 | (10.9, 20.0) |
| Boscolo-Rizzo et al | Europe | 365 | 12.8 | (9.3, 17.1) |
| COMEBAC study | Europe | 127 | 16.3 | (13.1, 19.9) |
| Caruso et al | Europe | 182 | 32.2 | (23.9, 41.4) |
| Caspersen et al | Europe | 350 | 9.9 | (7.9, 12.2) |
| Chai et al | China | 378 | 6.6 | (4.7, 9.0) |
| Cirulli et al | N America | 90 | 16.1 | (10.0, 24.0) |
| Clavario et al | Europe | 121 | 59.0 | (51.8, 65.9) |
| Diaz-Fuentes et al | N America | 84 | 55.9 | (46.1, 65.3) |
| Domenech-Montoliu et al | Europe | 213 | 6.8 | (4.7, 9.4) |
| Erol et al | Other | 170 | 3.3 | (0.9, 8.2) |
| Fernandez-de-Las-Penas et al | Europe | 227 | 23.5 | (21.1, 26.1) |
| Froidure et al | Europe | 95 | 35.7 | (27.4, 44.7) |
| Gaber et al | Europe | 122 | 40.0 | (31.6, 48.5) |
| Garcia-Abellan et al | Europe | 182 | 4.3 | (1.4, 9.8) |
| Gonzalez-Hermosillo et al | Other | 105 | 50.8 | (41.9, 59.6) |
| Han et al | China | 175 | 14.0 | (8.2, 21.8) |
| Havervall et al | Europe | 122 | 3.4 | (1.7, 6.0) |
| Jacobson et al | N America | 119 | 26.5 | (18.8, 35.5) |
| Kim et al | Other | 195 | 5.8 | (4.4, 7.5) |
| Lemhofer et al | Europe | 91 | 26.0 | (21.6, 30.8) |
| Liao et al | China | 104 | 1.2 | (0.1, 4.1) |
| Liyanage-Don et al | N America | 113 | 19.0 | (13.1, 26.1) |
| Logue et al | N America | 169 | 6.7 | (3.5, 11.4) |
| Lui et al | China | 96 | 7.8 | (4.5, 12.4) |
| Maestre-Muniz et al | Europe | 379 | 19.3 | (16.1, 22.9) |
| Mechi et al | Other | 274 | 30.4 | (22.0, 39.8) |
| Mei et al | China | 158 | 3.7 | (3.1, 4.4) |
| Menges et al | Europe | 219 | 22.3 | (18.5, 26.6) |
| Millet et al | N America | 365 | 25.4 | (19.1, 32.6) |
| Mohiuddin et al | Other | 161 | 3.8 | (2.0, 6.5) |
| Munblit et al | Europe | 232 | 17.0 | (15.6, 18.5) |
| Nehme et al | Europe | 243 | 11.7 | (8.8, 15.2) |
| O'Keefe et al | N America | 119 | 12.6 | (8.3, 18.1) |
| Ong et al | Other | 104 | 1.7 | (0.4, 4.9) |
| Orru et al | Europe | 91 | 40.1 | (32.3, 48.4) |
| Osmanov et al | Europe | 270 | 1.4 | (0.6, 2.8) |
| PHOSP-COVID study | Europe | 174 | 48.1 | (44.5, 51.7) |
| Peghin et al | Europe | 191 | 6.0 | (4.2, 8.2) |
| Peluso et al | N America | 112 | 22.0 | (14.9, 30.6) |
| Petersen et al | Europe | 125 | 9.0 | (5.2, 14.2) |
| Qin et al | China | 105 | 8.7 | (6.6, 11.1) |
| Righi et al | Europe | 84 | 9.0 | (6.5, 12.2) |
| Roessler et al (adults) | Europe | 91 | 3.1 | (3.0, 3.2) |
| Roessler et al (children) | Europe | 91 | 1.0 | (0.9, 1.2) |
| Romero-Duarte et al | Europe | 196 | 28.0 | (24.9, 31.3) |
| Sathyamurthy et al | Other | 104 | 1.8 | (0.6, 4.1) |
| Seeßle et al | Europe | 147 | 25.3 | (18.5, 33.2) |
| Shang et al | China | 196 | 20.4 | (17.6, 23.3) |
| Sibila et al | Europe | 116 | 39.5 | (32.2, 47.3) |
| Sigfrid et al | Europe | 222 | 53.5 | (47.9, 59.0) |
| Skala et al | Europe | 91 | 23.0 | (15.2, 32.5) |
| Skjorten et al | Europe | 105 | 46.8 | (37.9, 55.9) |
| Sonnweber et al | Europe | 103 | 36.0 | (27.9, 44.9) |
| Soraas et al | Europe | 126 | 10.0 | (7.8, 12.5) |
| Stavem et al | Europe | 117 | 16.0 | (12.7, 19.7) |
| Stephenson et al | Europe | 104 | 23.4 | (21.9, 24.9) |
| Sykes et al | Europe | 127 | 59.1 | (50.0, 67.7) |
| Taboada et al | Europe | 189 | 10.4 | (6.4, 15.7) |
| Taquet et al | N America | 90 | 7.9 | (7.8, 8.0) |
| Tawfik et al | Other | 91 | 26.7 | (19.0, 35.5) |
| Tleyjeh et al | Other | 136 | 40.1 | (33.6, 46.9) |
| Todt et al | Other | 105 | 27.9 | (22.4, 33.9) |
| Trunfio et al | Europe | 194 | 12.5 | (7.9, 18.5) |
| Ursini et al | Europe | 182 | 43.8 | (39.9, 47.9) |
| Venturelli et al | Europe | 105 | 21.8 | (18.9, 24.9) |
| Xiong et al | China | 111 | 39.0 | (34.9, 43.3) |
| Yin et al | China | 203 | 27.0 | (22.3, 32.1) |
| Zayet et al | Europe | 289 | 11.0 | (7.9, 14.7) |
| Zhan et al | China | 348 | 18.2 | (11.8, 26.2) |
| Zhang et al | China | 378 | 4.1 | (3.4, 5.0) |
| **95% Prediction Interval** | | | | |

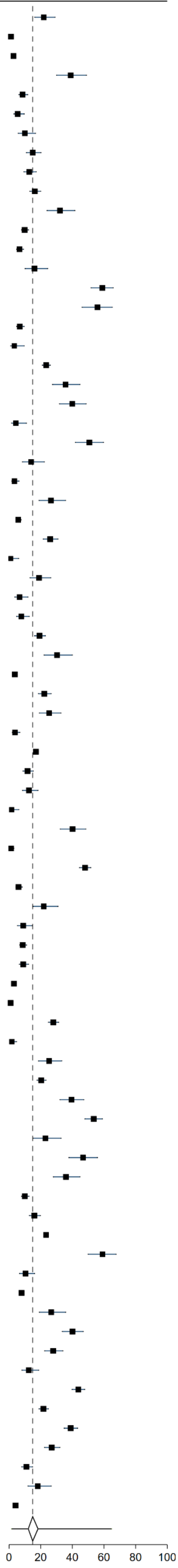

Supplementary Figure 12: Forest plot of prevalence of Long Covid in studies that reported chest pain as a persistent symptom **with 95% prediction intervals**

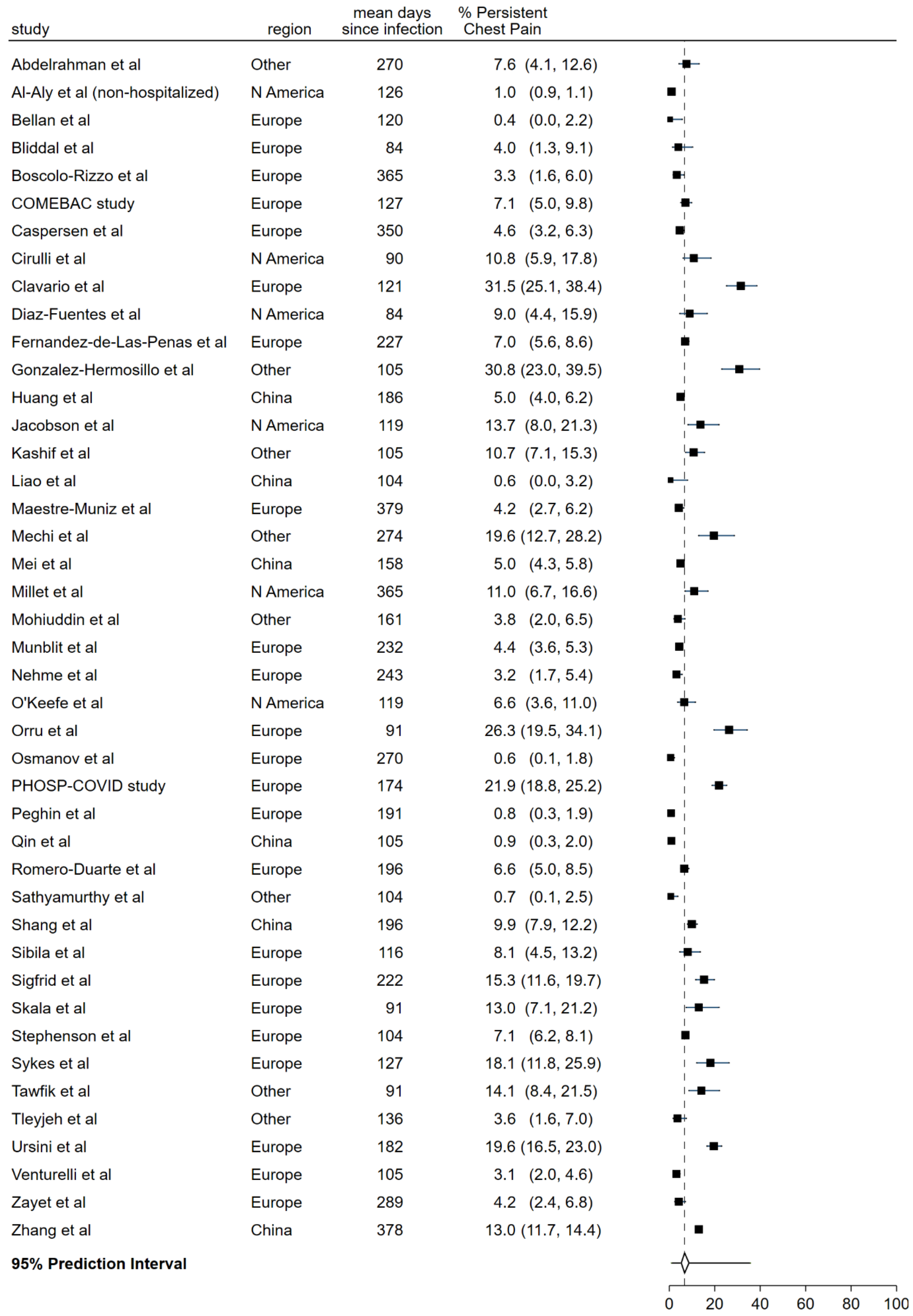

Supplementary Figure 13: Forest plot of prevalence of Long Covid in studies that reported chills as a persistent symptom with 95% prediction intervals

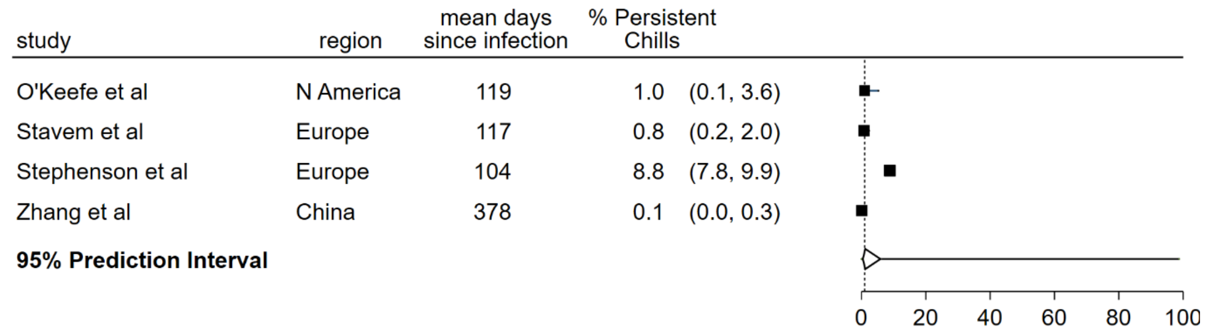

Supplementary Figure 14: Forest plot of prevalence of Long Covid in studies that reported cognition or memory problems as a persistent symptom with 95% prediction intervals

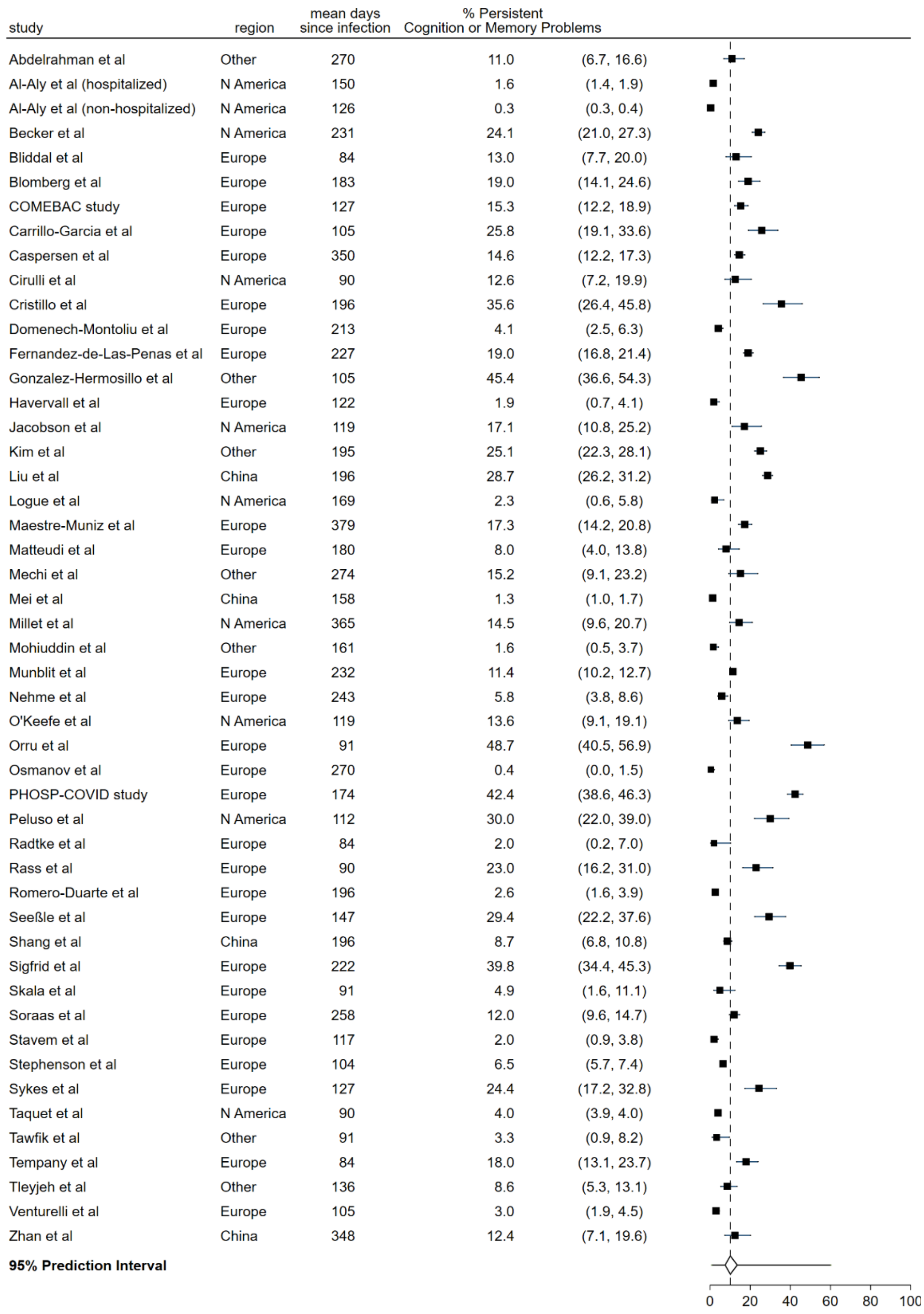

Supplementary Figure 15: Forest plot of prevalence of Long Covid in studies that reported cough as a persistent symptom with 95% prediction intervals

| study | region | mean days since infection | % Persistent Cough |
|---|---|---|---|
| Abdelrahman et al | Other | 270 | 14.0 (9.2, 20.2) |
| Bellan et al | Europe | 120 | 2.5 (0.9, 5.4) |
| Bliddal et al | Europe | 84 | 4.5 (1.7, 9.6) |
| Blomberg et al | Europe | 183 | 6.0 (3.4, 9.7) |
| Boscolo-Rizzo et al | Europe | 365 | 3.6 (1.8, 6.3) |
| COMEBAC study | Europe | 127 | 4.4 (2.7, 6.7) |
| Caruso et al | Europe | 182 | 18.6 (12.1, 26.9) |
| Caspersen et al | Europe | 350 | 2.6 (1.6, 4.0) |
| Chai et al | China | 378 | 6.4 (4.5, 8.8) |
| Cirulli et al | N America | 90 | 18.5 (12.0, 26.6) |
| Diaz-Fuentes et al | N America | 84 | 42.3 (33.2, 52.1) |
| Domenech-Montoliu et al | Europe | 213 | 5.4 (3.6, 7.8) |
| Erol et al | Other | 170 | 0.8 (0.0, 4.5) |
| Fernandez-de-Las-Penas et al | Europe | 227 | 2.1 (1.3, 3.1) |
| Froidure et al | Europe | 95 | 11.1 (6.2, 17.9) |
| Garcia-Abellan et al | Europe | 182 | 4.3 (1.4, 9.8) |
| Han et al | China | 175 | 6.1 (2.5, 12.1) |
| Jacobson et al | N America | 119 | 8.5 (4.1, 15.0) |
| Kashif et al | Other | 105 | 6.6 (3.8, 10.5) |
| Liao et al | China | 104 | 15.7 (10.6, 22.0) |
| Logue et al | N America | 169 | 2.8 (0.9, 6.4) |
| Lui et al | China | 96 | 7.8 (4.5, 12.4) |
| Mechi et al | Other | 274 | 17.9 (11.3, 26.2) |
| Mei et al | China | 158 | 2.4 (1.9, 3.0) |
| Millet et al | N America | 365 | 13.3 (8.6, 19.3) |
| Mohiuddin et al | Other | 161 | 5.1 (2.9, 8.1) |
| Munblit et al | Europe | 232 | 6.7 (5.8, 7.7) |
| Nehme et al | Europe | 243 | 3.7 (2.1, 6.0) |
| O'Keefe et al | N America | 119 | 9.6 (5.7, 14.9) |
| Ong et al | Other | 104 | 2.3 (0.6, 5.8) |
| Orru et al | Europe | 91 | 21.0 (14.9, 28.4) |
| Osmanov et al | Europe | 270 | 1.0 (0.3, 2.3) |
| PHOSP-COVID study | Europe | 174 | 26.4 (23.3, 29.7) |
| Peghin et al | Europe | 191 | 2.0 (1.0, 3.5) |
| Petersen et al | Europe | 125 | 9.0 (5.2, 14.2) |
| Qin et al | China | 105 | 6.0 (4.3, 8.1) |
| Romero-Duarte et al | Europe | 196 | 19.2 (16.5, 22.1) |
| Sathyamurthy et al | Other | 104 | 4.3 (2.2, 7.4) |
| Seeßle et al | Europe | 147 | 10.3 (5.9, 16.4) |
| Shang et al | China | 196 | 12.4 (10.2, 14.9) |
| Sibila et al | Europe | 116 | 18.0 (12.6, 24.6) |
| Sigfrid et al | Europe | 222 | 15.3 (11.6, 19.7) |
| Skala et al | Europe | 91 | 14.0 (7.9, 22.4) |
| Soraas et al | Europe | 126 | 8.0 (6.0, 10.3) |
| Stavem et al | Europe | 117 | 6.0 (4.0, 8.6) |
| Stephenson et al | Europe | 104 | 3.2 (2.6, 3.9) |
| Sykes et al | Europe | 127 | 35.4 (27.2, 44.4) |
| Tleyjeh et al | Other | 136 | 27.5 (21.7, 33.9) |
| Trunfio et al | Europe | 194 | 4.2 (1.7, 8.4) |
| Venturelli et al | Europe | 105 | 3.0 (1.9, 4.5) |
| Zayet et al | Europe | 289 | 2.5 (1.1, 4.7) |
| Zhang et al | China | 378 | 4.1 (3.4, 4.9) |
| **95% Prediction Interval** | | | |

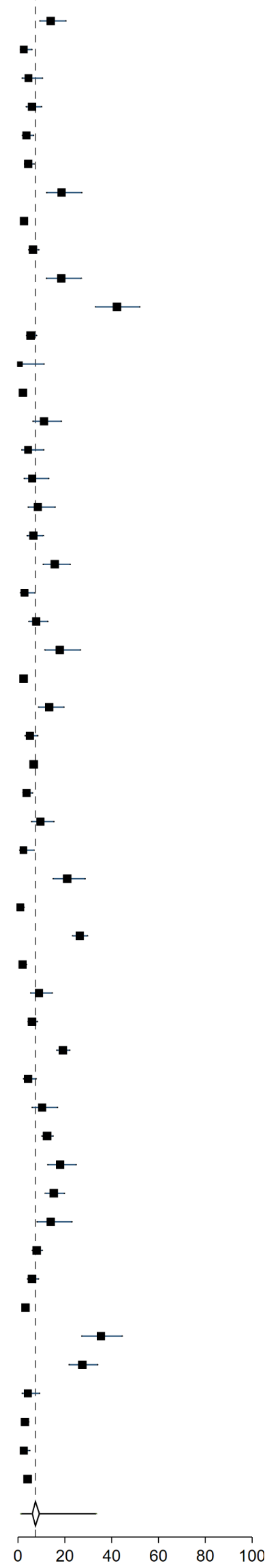

Supplementary Figure 16: Forest plot of prevalence of Long Covid in studies that reported dizziness as a persistent symptom with 95% prediction intervals

| study | region | mean days since infection | % Persistent Dizziness |
|---|---|---|---|
| Blomberg et al | Europe | 183 | 10.0 (6.5, 14.5) |
| Boscolo-Rizzo et al | Europe | 365 | 3.2 (1.5, 5.8) |
| Caspersen et al | Europe | 350 | 2.9 (1.8, 4.4) |
| Cirulli et al | N America | 90 | 7.4 (3.4, 13.5) |
| Cristillo et al | Europe | 196 | 16.8 (10.1, 25.6) |
| Domenech-Montoliu et al | Europe | 213 | 4.6 (2.9, 6.9) |
| Erol et al | Other | 170 | 5.8 (2.4, 11.6) |
| Gonzalez-Hermosillo et al | Other | 105 | 39.2 (30.8, 48.2) |
| Kashif et al | Other | 105 | 14.5 (10.3, 19.6) |
| Mechi et al | Other | 274 | 7.0 (3.4, 12.5) |
| Millet et al | N America | 365 | 8.7 (5.0, 14.0) |
| Munblit et al | Europe | 232 | 5.3 (4.5, 6.2) |
| Nehme et al | Europe | 243 | 3.2 (1.7, 5.4) |
| O'Keefe et al | N America | 119 | 4.0 (1.7, 7.7) |
| Orru et al | Europe | 91 | 22.4 (16.0, 29.8) |
| Osmanov et al | Europe | 270 | 1.0 (0.3, 2.4) |
| PHOSP-COVID study | Europe | 174 | 33.2 (29.6, 36.9) |
| Rass et al | Europe | 90 | 7.0 (3.2, 12.8) |
| Romero-Duarte et al | Europe | 196 | 1.3 (0.6, 2.4) |
| Sathyamurthy et al | Other | 104 | 1.4 (0.4, 3.5) |
| Seeßle et al | Europe | 147 | 16.4 (10.8, 23.5) |
| Sigfrid et al | Europe | 222 | 35.8 (30.6, 41.2) |
| Stephenson et al | Europe | 104 | 13.7 (12.5, 15.0) |
| Tawfik et al | Other | 91 | 12.5 (7.2, 19.8) |
| Tleyjeh et al | Other | 136 | 0.9 (0.1, 3.2) |
| Zhang et al | China | 378 | 3.3 (2.6, 4.1) |
| **95% Prediction Interval** | | | |

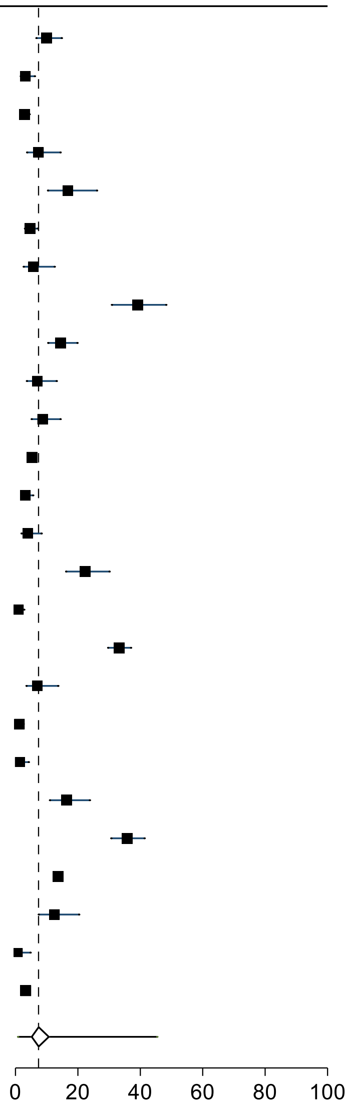

Supplementary Figure 17: Forest plot of prevalence of Long Covid in studies that reported ear problems as a persistent symptom with 95% prediction intervals

| study | region | mean days since infection | % Persistent Ear Problems |
|---|---|---|---|
| Cirulli et al | N America | 90 | 4.2 (1.4, 9.5) |
| Domenech-Montoliu et al | Europe | 213 | 4.8 (3.1, 7.1) |
| Mei et al | China | 158 | 0.1 (0.0, 0.4) |
| Munblit et al | Europe | 232 | 5.1 (4.3, 6.0) |
| O'Keefe et al | N America | 119 | 4.0 (1.7, 7.7) |
| Orru et al | Europe | 91 | 14.5 (9.3, 21.1) |
| Stavem et al | Europe | 117 | 1.1 (0.4, 2.5) |
| Stephenson et al | Europe | 104 | 6.2 (5.4, 7.1) |
| Tleyjeh et al | Other | 136 | 0.5 (0.0, 2.8) |
| Ursini et al | Europe | 182 | 20.0 (16.9, 23.4) |
| Zayet et al | Europe | 289 | 4.5 (2.6, 7.2) |
| **95% Prediction Interval** | | | |

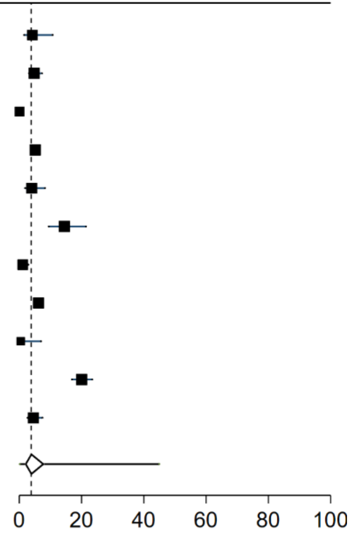

Supplementary Figure 18: Forest plot of prevalence of Long Covid in studies that reported eye problems as a persistent symptom with 95% prediction intervals

| study | region | mean days since infection | % Persistent Eye Problems |
|---|---|---|---|
| Domenech-Montoliu et al | Europe | 213 | 3.7 (2.2, 5.8) |
| Orru et al | Europe | 91 | 21.0 (14.9, 28.4) |
| Stavem et al | Europe | 117 | 4.0 (2.4, 6.2) |
| Ursini et al | Europe | 182 | 25.0 (21.6, 28.6) |
| **95% Prediction Interval** | | | |

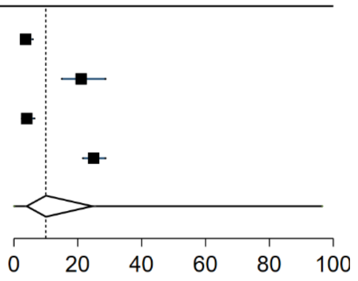

Supplementary Figure 19: Forest plot of prevalence of Long Covid in studies that reported fatigue as a persistent symptom with 95% prediction intervals

| study | region | mean days since infection | % Persistent Fatigue |
|---|---|---|---|
| Abdelrahman et al | Other | 270 | 37.3 (30.0, 44.9) |
| Al-Aly et al (hospitalized) | N America | 150 | 3.6 (3.3, 4.0) |
| Al-Aly et al (non-hospitalized) | N America | 126 | 1.3 (1.2, 1.3) |
| Arnold et al | Europe | 90 | 39.0 (29.9, 48.9) |
| Augustin et al | Europe | 131 | 9.7 (7.1, 12.9) |
| Bliddal et al | Europe | 84 | 16.0 (10.2, 23.5) |
| Blomberg et al | Europe | 183 | 30.0 (24.2, 36.4) |
| Boscolo-Rizzo et al | Europe | 365 | 27.3 (22.4, 32.7) |
| COMEBAC study | Europe | 127 | 28.0 (24.0, 32.2) |
| Caruso et al | Europe | 182 | 5.9 (2.4, 11.8) |
| Caspersen et al | Europe | 350 | 13.6 (11.3, 16.2) |
| Castro et al | N America | 128 | 10.9 (10.2, 11.7) |
| Chai et al | China | 378 | 10.6 (8.2, 13.5) |
| Cirulli et al | N America | 90 | 8.0 (3.7, 14.6) |
| Clavario et al | Europe | 121 | 57.5 (50.3, 64.4) |
| Diaz-Fuentes et al | N America | 84 | 5.4 (2.0, 11.4) |
| Domenech-Montoliu et al | Europe | 213 | 18.0 (14.7, 21.7) |
| Erol et al | Other | 170 | 0.8 (0.0, 4.5) |
| Fernandez-de-Las-Penas et al | Europe | 227 | 60.8 (57.9, 63.6) |
| Froidure et al | Europe | 95 | 25.0 (17.8, 33.4) |
| Gaber et al | Europe | 122 | 39.0 (30.9, 47.8) |
| Garcia-Abellan et al | Europe | 182 | 10.3 (5.4, 17.2) |
| Gonzalez-Hermosillo et al | Other | 105 | 53.0 (44.1, 61.9) |
| Havervall et al | Europe | 122 | 6.8 (4.3, 10.1) |
| Huang et al | China | 186 | 63.0 (60.6, 65.3) |
| Jacobson et al | N America | 119 | 30.8 (22.6, 40.0) |
| Kashif et al | Other | 105 | 41.7 (35.5, 48.2) |
| Kim et al | Other | 195 | 26.4 (23.5, 29.4) |
| Lemhofer et al | Europe | 91 | 37.5 (32.5, 42.7) |
| Liyanage-Don et al | N America | 113 | 20.3 (14.2, 27.5) |
| Logue et al | N America | 169 | 13.6 (8.9, 19.6) |
| Lui et al | China | 96 | 3.4 (1.4, 6.9) |
| Maestre-Muniz et al | Europe | 379 | 19.3 (16.1, 22.9) |
| Mechi et al | Other | 274 | 61.6 (51.9, 70.6) |
| Mei et al | China | 158 | 1.5 (1.1, 1.9) |
| Menges et al | Europe | 219 | 54.1 (49.2, 58.8) |
| Millet et al | N America | 365 | 21.0 (15.6, 27.2) |
| Mohiuddin et al | Other | 161 | 7.4 (4.7, 10.9) |
| Munblit et al | Europe | 232 | 25.1 (23.4, 26.8) |
| Nehme et al | Europe | 243 | 20.7 (16.9, 24.9) |
| O'Keefe et al | N America | 119 | 21.2 (15.7, 27.6) |
| Ong et al | Other | 104 | 0.6 (0.0, 3.3) |
| Orru et al | Europe | 91 | 74.3 (66.6, 81.1) |
| Osmanov et al | Europe | 270 | 10.7 (8.1, 13.8) |
| PHOSP-COVID study | Europe | 174 | 56.2 (52.6, 59.8) |
| Peghin et al | Europe | 191 | 13.1 (10.5, 16.1) |
| Peluso et al | N America | 112 | 28.0 (20.1, 37.0) |
| Petersen et al | Europe | 125 | 29.0 (22.5, 36.3) |
| Radtke et al | Europe | 84 | 2.8 (0.6, 7.8) |
| Rass et al | Europe | 90 | 27.0 (19.5, 35.4) |
| Righi et al | Europe | 84 | 9.0 (6.5, 12.2) |
| Romero-Duarte et al | Europe | 196 | 22.1 (19.3, 25.2) |
| Sathyamurthy et al | Other | 104 | 8.9 (5.8, 12.9) |
| Seeßle et al | Europe | 147 | 39.0 (31.1, 47.5) |
| Shang et al | China | 196 | 25.3 (22.3, 28.4) |
| Sigfrid et al | Europe | 222 | 82.8 (78.1, 86.8) |
| Simani et al | Other | 197 | 10.0 (5.3, 16.8) |
| Skala et al | Europe | 91 | 21.6 (14.0, 30.8) |
| Soraas et al | Europe | 258 | 52.0 (48.1, 55.9) |
| Stavem et al | Europe | 118 | 46.0 (41.4, 50.7) |
| Stephenson et al | Europe | 104 | 39.0 (37.3, 40.7) |
| Sykes et al | Europe | 127 | 37.8 (29.3, 46.8) |
| Taquet et al | N America | 90 | 5.9 (5.8, 6.0) |
| Tawfik et al | Other | 91 | 33.3 (25.0, 42.5) |
| Tempany et al | Europe | 84 | 41.0 (34.4, 47.9) |
| Tleyjeh et al | Other | 136 | 29.7 (23.8, 36.2) |
| Townsend et al | Europe | 86 | 52.3 (43.3, 61.2) |
| Venturelli et al | Europe | 105 | 24.3 (21.3, 27.4) |
| Xiong et al | China | 111 | 28.3 (24.5, 32.3) |
| Zayet et al | Europe | 289 | 19.2 (15.2, 23.7) |
| Zhan et al | China | 348 | 11.6 (6.5, 18.7) |
| Zhang et al | China | 378 | 27.7 (26.0, 29.5) |
| **95% Prediction Interval** | | | |

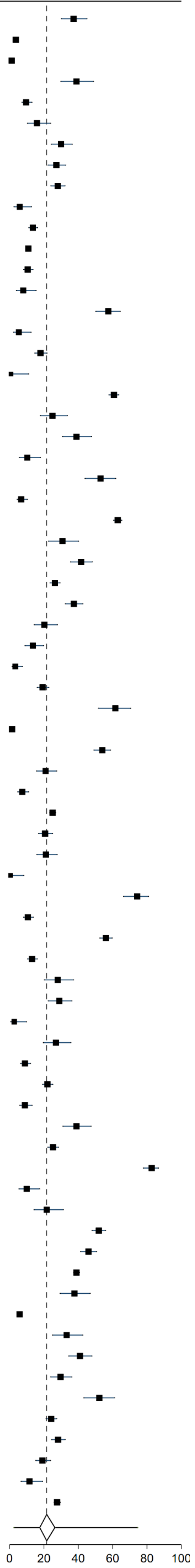

Supplementary Figure 20: Forest plot of prevalence of Long Covid in studies that reported fever as a persistent symptom with 95% prediction intervals

| study | region | mean days since infection | % Persistent Fever |
|---|---|---|---|
| Al-Aly et al (hospitalized) | N America | 150 | 0.5 (0.4, 0.6) |
| Al-Aly et al (non-hospitalized) | N America | 126 | 0.1 (0.1, 0.1) |
| Bliddal et al | Europe | 84 | 0.5 (0.0, 2.8) |
| Blomberg et al | Europe | 183 | 2.0 (0.5, 5.0) |
| Boscolo-Rizzo et al | Europe | 365 | 1.0 (0.2, 2.9) |
| Caspersen et al | Europe | 350 | 0.7 (0.2, 1.6) |
| Cirulli et al | N America | 90 | 1.7 (0.2, 6.0) |
| Domenech-Montoliu et al | Europe | 213 | 1.0 (0.3, 2.3) |
| Jacobson et al | N America | 119 | 0.9 (0.0, 4.9) |
| Logue et al | N America | 169 | 1.1 (0.1, 3.9) |
| Mohiuddin et al | Other | 161 | 0.6 (0.1, 2.2) |
| Orru et al | Europe | 91 | 19.1 (13.2, 26.2) |
| Romero-Duarte et al | Europe | 196 | 7.0 (5.3, 9.0) |
| Seeßle et al | Europe | 147 | 0.7 (0.0, 3.8) |
| Sigfrid et al | Europe | 222 | 11.0 (7.8, 14.9) |
| Soraas et al | Europe | 126 | 3.0 (1.8, 4.8) |
| Stavem et al | Europe | 117 | 0.4 (0.0, 1.4) |
| Stephenson et al | Europe | 104 | 1.6 (1.2, 2.1) |
| Sykes et al | Europe | 127 | 10.2 (5.6, 16.9) |
| Tleyjeh et al | Other | 136 | 0.9 (0.1, 3.2) |
| Ursini et al | Europe | 182 | 24.5 (21.2, 28.1) |
| Venturelli et al | Europe | 105 | 0.5 (0.1, 1.3) |
| Xiong et al | China | 111 | 23.6 (20.1, 27.4) |
| Zayet et al | Europe | 289 | 0.3 (0.0, 1.7) |
| **95% Prediction Interval** | | | |

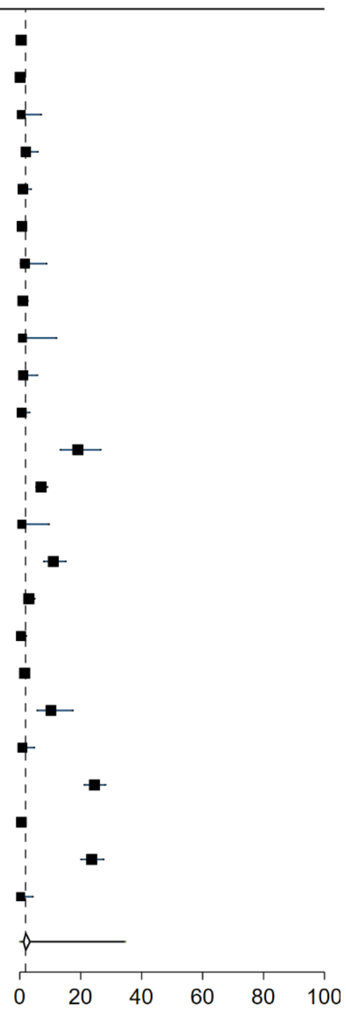

Supplementary Figure 21: Forest plot of prevalence of Long Covid in studies that reported nausea or vomiting as a persistent symptom with 95% prediction intervals

| study | region | mean days since infection | % Persistent Nausea or Vomitting |
|---|---|---|---|
| Abdelrahman et al | Other | 270 | 4.7 (2.1, 9.1) |
| Al-Aly et al (hospitalized) | N America | 150 | 1.3 (1.1, 1.5) |
| Al-Aly et al (non-hospitalized) | N America | 126 | 0.4 (0.3, 0.4) |
| Bellan et al | Europe | 120 | 1.3 (0.3, 3.7) |
| Bliddal et al | Europe | 84 | 2.0 (0.4, 5.7) |
| Blomberg et al | Europe | 183 | 6.0 (3.4, 9.7) |
| Boscolo-Rizzo et al | Europe | 365 | 1.6 (0.5, 3.7) |
| Caruso et al | Europe | 182 | 14.4 (8.3, 22.7) |
| Cirulli et al | N America | 90 | 5.0 (1.9, 10.6) |
| Domenech-Montoliu et al | Europe | 213 | 4.3 (2.7, 6.5) |
| Fernandez-de-Las-Penas et al | Europe | 227 | 7.2 (5.8, 8.9) |
| Gonzalez-Hermosillo et al | Other | 105 | 14.6 (9.0, 21.9) |
| Huang et al | China | 186 | 5.0 (4.0, 6.2) |
| Jacobson et al | N America | 119 | 6.8 (3.0, 12.9) |
| Kashif et al | Other | 105 | 12.0 (8.2, 16.8) |
| Lemhofer et al | Europe | 91 | 14.0 (10.6, 18.0) |
| Logue et al | N America | 169 | 2.8 (0.9, 6.4) |
| Lui et al | China | 96 | 0.5 (0.0, 2.8) |
| Maestre-Muniz et al | Europe | 379 | 1.1 (0.4, 2.4) |
| Mei et al | China | 158 | 1.1 (0.8, 1.5) |
| Munblit et al | Europe | 232 | 2.5 (1.9, 3.2) |
| Nehme et al | Europe | 243 | 2.2 (1.0, 4.1) |
| O'Keefe et al | N America | 119 | 0.7 (0.1, 2.5) |
| Ong et al | Other | 104 | 0.6 (0.0, 3.3) |
| Orru et al | Europe | 91 | 22.4 (16.0, 29.8) |
| Osmanov et al | Europe | 270 | 2.0 (1.0, 3.6) |
| PHOSP-COVID study | Europe | 174 | 17.0 (14.2, 20.0) |
| Peghin et al | Europe | 191 | 1.5 (0.7, 2.8) |
| Petersen et al | Europe | 125 | 3.0 (1.0, 6.8) |
| Roessler et al (adults) | Europe | 91 | 2.7 (2.6, 2.8) |
| Roessler et al (children) | Europe | 91 | 1.9 (1.6, 2.1) |
| Romero-Duarte et al | Europe | 196 | 10.3 (8.3, 12.6) |
| Seeßle et al | Europe | 147 | 4.8 (1.9, 9.6) |
| Shang et al | China | 196 | 10.9 (8.8, 13.3) |
| Sibila et al | Europe | 116 | 0.6 (0.0, 3.3) |
| Sigfrid et al | Europe | 222 | 17.7 (13.7, 22.3) |
| Skala et al | Europe | 91 | 2.0 (0.2, 7.0) |
| Sonnweber et al | Europe | 103 | 9.0 (4.7, 15.2) |
| Soraas et al | Europe | 126 | 6.0 (4.3, 8.1) |
| Stavem et al | Europe | 117 | 1.1 (0.4, 2.5) |
| Stephenson et al | Europe | 104 | 3.0 (2.4, 3.7) |
| Taquet et al | N America | 90 | 8.3 (8.2, 8.4) |
| Tawfik et al | Other | 91 | 5.0 (1.9, 10.6) |
| Tleyjeh et al | Other | 136 | 1.4 (0.3, 4.0) |
| Ursini et al | Europe | 182 | 11.2 (8.8, 14.0) |
| Venturelli et al | Europe | 105 | 0.9 (0.4, 1.9) |
| Weng et al | China | 104 | 44.4 (35.3, 53.9) |
| Zayet et al | Europe | 289 | 2.5 (1.1, 4.7) |
| Zhang et al | China | 378 | 0.7 (0.4, 1.1) |
| **95% Prediction Interval** | | | |

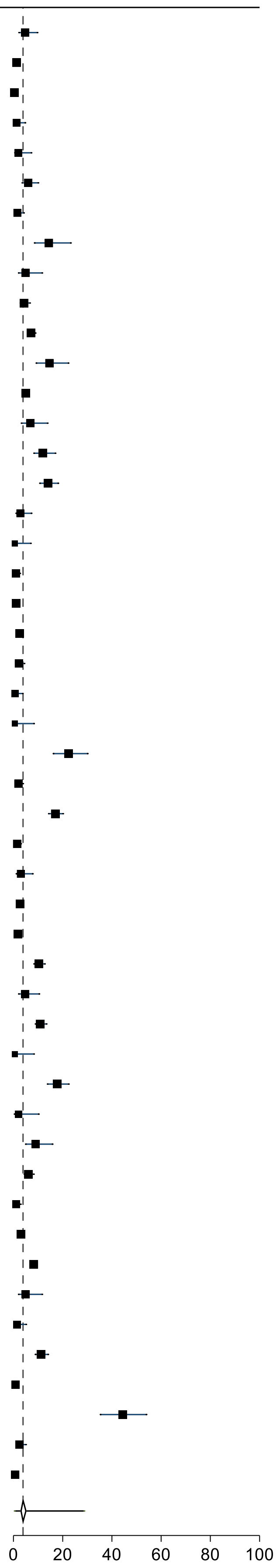

Supplementary Figure 22: Forest plot of prevalence of Long Covid in studies that reported headache as a persistent symptom with 95% prediction intervals

| study | region | mean days since infection | % Persistent Headache |
|---|---|---|---|
| Abdelrahman et al | Other | 270 | 10.5 (6.4, 16.1) |
| Al-Aly et al (hospitalized) | N America | 150 | 0.2 (0.1, 0.3) |
| Al-Aly et al (non-hospitalized) | N America | 126 | 0.4 (0.4, 0.5) |
| Bliddal et al | Europe | 84 | 6.5 (2.8, 12.4) |
| Blomberg et al | Europe | 183 | 11.0 (7.4, 15.5) |
| Boscolo-Rizzo et al | Europe | 365 | 3.6 (1.8, 6.3) |
| COMEBAC study | Europe | 127 | 4.8 (3.1, 7.1) |
| Caspersen et al | Europe | 350 | 4.9 (3.5, 6.7) |
| Cirulli et al | N America | 90 | 7.7 (3.6, 14.1) |
| Cristillo et al | Europe | 196 | 8.9 (4.2, 16.2) |
| Domenech-Montoliu et al | Europe | 213 | 14.9 (11.9, 18.4) |
| Erol et al | Other | 170 | 2.5 (0.5, 7.1) |
| Gonzalez-Hermosillo et al | Other | 105 | 29.2 (21.6, 37.8) |
| Havervall et al | Europe | 122 | 2.5 (1.1, 4.9) |
| Huang et al | China | 186 | 2.0 (1.4, 2.8) |
| Jacobson et al | N America | 119 | 6.0 (2.4, 11.9) |
| Kashif et al | Other | 105 | 19.0 (14.3, 24.5) |
| Liyanage-Don et al | N America | 113 | 13.1 (8.2, 19.5) |
| Logue et al | N America | 169 | 5.0 (2.3, 9.3) |
| Lui et al | China | 96 | 0.5 (0.0, 2.8) |
| Maestre-Muniz et al | Europe | 379 | 9.0 (6.7, 11.7) |
| Matteudi et al | Europe | 180 | 5.8 (2.5, 11.1) |
| Mechi et al | Other | 274 | 17.9 (11.3, 26.2) |
| Millet et al | N America | 365 | 16.8 (11.5, 23.2) |
| Mohiuddin et al | Other | 161 | 1.2 (0.3, 3.0) |
| Munblit et al | Europe | 232 | 7.4 (6.4, 8.5) |
| Nehme et al | Europe | 243 | 10.0 (7.3, 13.3) |
| O'Keefe et al | N America | 119 | 11.6 (7.5, 16.9) |
| Orru et al | Europe | 91 | 46.7 (38.6, 55.0) |
| Osmanov et al | Europe | 270 | 3.5 (2.1, 5.5) |
| PHOSP-COVID study | Europe | 174 | 33.4 (29.8, 37.1) |
| Peghin et al | Europe | 191 | 2.7 (1.5, 4.3) |
| Peluso et al | N America | 112 | 17.0 (10.7, 25.0) |
| Petersen et al | Europe | 125 | 7.0 (3.8, 11.7) |
| Rass et al | Europe | 90 | 5.0 (2.0, 10.0) |
| Romero-Duarte et al | Europe | 196 | 5.3 (3.8, 7.1) |
| Sathyamurthy et al | Other | 104 | 0.4 (0.0, 2.2) |
| Seeßle et al | Europe | 147 | 14.4 (9.1, 21.1) |
| Shang et al | China | 196 | 1.9 (1.1, 3.1) |
| Sibila et al | Europe | 116 | 1.7 (0.4, 4.9) |
| Sigfrid et al | Europe | 222 | 39.4 (34.1, 45.0) |
| Skala et al | Europe | 91 | 5.9 (2.2, 12.4) |
| Soraas et al | Europe | 126 | 13.0 (10.5, 15.8) |
| Stavem et al | Europe | 117 | 6.0 (4.0, 8.6) |
| Stephenson et al | Europe | 104 | 23.2 (21.7, 24.7) |
| Taquet et al | N America | 90 | 4.6 (4.6, 4.7) |
| Tawfik et al | Other | 91 | 19.2 (12.6, 27.4) |
| Tleyjeh et al | Other | 136 | 8.6 (5.3, 13.1) |
| Venturelli et al | Europe | 105 | 0.5 (0.1, 1.3) |
| Zayet et al | Europe | 289 | 9.6 (6.7, 13.2) |
| Zhang et al | China | 378 | 2.3 (1.7, 3.0) |
| **95% Prediction Interval** | | | |

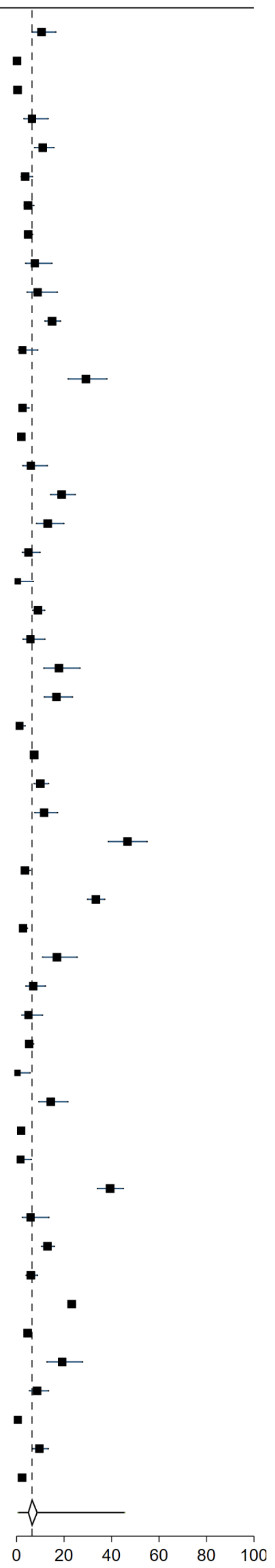

Supplementary Figure 23: Forest plot of prevalence of Long Covid in studies that reported persistent hypertension as a persistent symptom with 95% prediction intervals

| study | region | mean days since infection | % Persistent Hypertension |
|---|---|---|---|
| Al-Aly et al (hospitalized) | N America | 150 | 1.7 (1.4, 1.9) |
| Al-Aly et al (non-hospitalized) | N America | 126 | 1.5 (1.4, 1.6) |
| Mohiuddin et al | Other | 161 | 1.3 (0.3, 3.2) |
| Shang et al | China | 196 | 0.4 (0.1, 1.1) |
| **95% Prediction Interval** | | | |

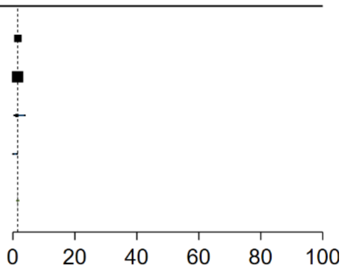

Supplementary Figure 24: Forest plot of prevalence of Long Covid in studies that reported palpitations as a persistent symptom with 95% prediction intervals

| study | region | mean days since infection | % Persistent Palpitations |
|---|---|---|---|
| Abdelrahman et al | Other | 270 | 0.6 (0.0, 3.3) |
| Blomberg et al | Europe | 183 | 6.0 (3.4, 9.7) |
| Caspersen et al | Europe | 350 | 5.6 (4.1, 7.5) |
| Chai et al | China | 378 | 2.0 (1.0, 3.6) |
| Cirulli et al | N America | 90 | 6.6 (2.9, 12.6) |
| Clavario et al | Europe | 121 | 25.5 (19.6, 32.1) |
| Erol et al | Other | 170 | 4.1 (1.4, 9.4) |
| Fernandez-de-Las-Penas et al | Europe | 227 | 6.7 (5.3, 8.3) |
| Havervall et al | Europe | 122 | 1.9 (0.8, 3.9) |
| Huang et al | China | 186 | 9.0 (7.7, 10.5) |
| Jacobson et al | N America | 119 | 6.0 (2.4, 11.9) |
| Kashif et al | Other | 105 | 12.4 (8.5, 17.2) |
| Lui et al | China | 96 | 0.5 (0.0, 2.8) |
| Maestre-Muniz et al | Europe | 379 | 4.6 (3.0, 6.7) |
| Mechi et al | Other | 274 | 16.1 (9.8, 24.2) |
| Mei et al | China | 158 | 1.7 (1.3, 2.2) |
| Millet et al | N America | 365 | 8.7 (5.0, 14.0) |
| Munblit et al | Europe | 232 | 7.1 (6.1, 8.2) |
| Nehme et al | Europe | 243 | 3.4 (1.9, 5.7) |
| O'Keefe et al | N America | 119 | 8.1 (4.7, 12.8) |
| Osmanov et al | Europe | 270 | 1.1 (0.3, 2.5) |
| Qin et al | China | 105 | 9.7 (7.6, 12.3) |
| Romero-Duarte et al | Europe | 196 | 3.1 (2.0, 4.5) |
| Seeßle et al | Europe | 147 | 15.1 (9.7, 21.9) |
| Tawfik et al | Other | 91 | 13.3 (7.8, 20.7) |
| Zhang et al | China | 378 | 4.2 (3.5, 5.1) |
| **95% Prediction Interval** | | | |

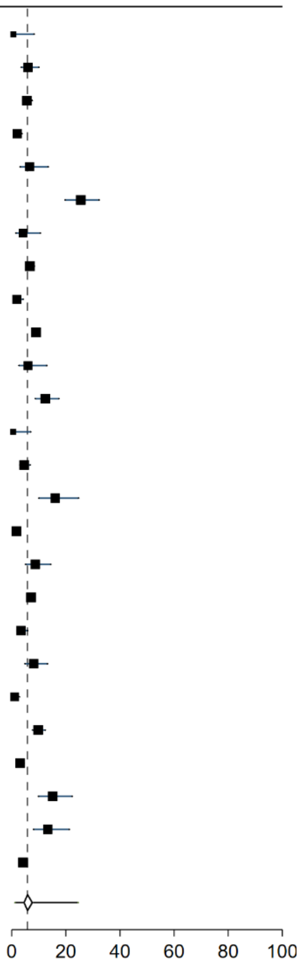

Supplementary Figure 25: Forest plot of prevalence of Long Covid in studies that reported psychological distress as a persistent symptom with 95% prediction intervals

| study | region | mean days since infection | % Persistent Psychological Distress | |
|---|---|---|---|---|
| Aminian et al | N America | 243 | 1.3 | (0.9, 1.8) |
| Shang et al | China | 196 | 1.0 | (0.4, 2.0) |
| Tempany et al | Europe | 84 | 15.7 | (11.1, 21.2) |
| **95% Prediction Interval** | | | | |

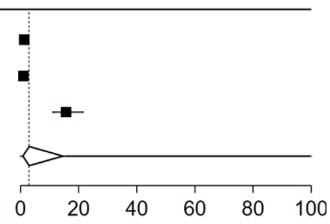

Supplementary Figure 26: Forest plot of prevalence of Long Covid in studies that reported PTSD as a persistent symptom with 95% prediction intervals

| study | region | mean days since infection | % Persistent PTSD | |
|---|---|---|---|---|
| Al-Aly et al (hospitalized) | N America | 150 | 1.4 | (1.2, 1.6) |
| Al-Aly et al (non-hospitalized) | N America | 126 | 0.9 | (0.8, 1.0) |
| Bellan et al | Europe | 120 | 42.9 | (36.5, 49.4) |
| COMEBAC study | Europe | 127 | 5.0 | (3.2, 7.3) |
| Liao et al | China | 409 | 21.9 | (16.5, 28.1) |
| Liyanage-Don et al | N America | 113 | 23.5 | (17.1, 31.1) |
| PHOSP-COVID study | Europe | 166 | 12.9 | (11.5, 14.5) |
| PHOSP-COVID study | Europe | 174 | 12.2 | (10.3, 14.3) |
| Rass et al | Europe | 90 | 11.0 | (5.6, 18.8) |
| Simani et al | Other | 197 | 5.8 | (2.4, 11.6) |
| Tarsitani et al | Europe | 105 | 10.4 | (5.5, 17.5) |
| Tawfik et al | Other | 91 | 14.2 | (8.5, 21.7) |
| **95% Prediction Interval** | | | | |

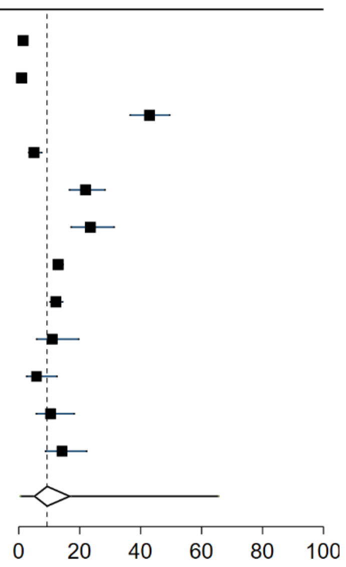

Supplementary Figure 27: Forest plot of prevalence of Long Covid in studies that reported skin problems as a persistent symptom with 95% prediction intervals

| study | region | mean days since infection | % Persistent Skin Problems | |
|---|---|---|---|---|
| Caspersen et al | Europe | 350 | 1.9 | (1.1, 3.1) |
| Domenech-Montoliu et al | Europe | 213 | 5.2 | (3.4, 7.6) |
| Nehme et al | Europe | 243 | 1.5 | (0.5, 3.2) |
| Orru et al | Europe | 91 | 18.4 | (12.6, 25.5) |
| Roessler et al (adults) | Europe | 91 | 0.6 | (0.5, 0.6) |
| Stavem et al | Europe | 117 | 1.1 | (0.4, 2.5) |
| **95% Prediction Interval** | | | | |

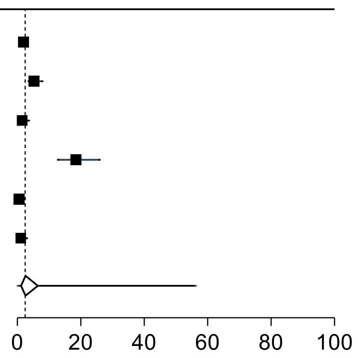

Supplementary Figure 28: Forest plot of prevalence of Long Covid in studies that reported sleep problems as a persistent symptom with 95% prediction intervals

| study | region | mean days since infection | % Persistent Sleep Problems | |
|---|---|---|---|---|
| Abdelrahman et al | Other | 270 | 13.4 | (8.7, 19.4) |
| Al-Aly et al (hospitalized) | N America | 150 | 1.5 | (1.3, 1.8) |
| Al-Aly et al (non-hospitalized) | N America | 126 | 1.5 | (1.3, 1.6) |
| Arnold et al | Europe | 90 | 24.0 | (16.4, 33.3) |
| Blomberg et al | Europe | 183 | 5.0 | (2.7, 8.4) |
| Boscolo-Rizzo et al | Europe | 365 | 7.2 | (4.6, 10.7) |
| COMEBAC study | Europe | 127 | 18.8 | (15.4, 22.6) |
| Caspersen et al | Europe | 350 | 2.9 | (1.8, 4.4) |
| Chai et al | China | 378 | 1.1 | (0.4, 2.4) |
| Cirulli et al | N America | 90 | 8.4 | (4.1, 14.9) |
| Cristillo et al | Europe | 196 | 36.6 | (27.3, 46.8) |
| Domenech-Montoliu et al | Europe | 213 | 11.6 | (8.9, 14.8) |
| Fernandez-de-Las-Penas et al | Europe | 227 | 34.5 | (31.7, 37.3) |
| Fu et al | China | 196 | 10.1 | (6.3, 15.2) |
| Gaber et al | Europe | 122 | 49.0 | (40.4, 57.5) |
| Gonzalez-Hermosillo et al | Other | 105 | 45.4 | (36.6, 54.3) |
| Havervall et al | Europe | 122 | 2.8 | (1.3, 5.3) |
| Huang et al | China | 186 | 26.0 | (23.9, 28.2) |
| Kashif et al | Other | 105 | 21.1 | (16.1, 26.8) |
| Kim et al | Other | 195 | 19.2 | (16.7, 21.9) |
| Lemhofer et al | Europe | 91 | 30.1 | (25.5, 35.1) |
| Maestre-Muniz et al | Europe | 379 | 11.0 | (8.5, 13.9) |
| Mazza et al | Europe | 104 | 3.1 | (1.3, 6.3) |
| Mei et al | China | 158 | 2.1 | (1.7, 2.6) |
| Munblit et al | Europe | 232 | 39.8 | (36.1, 43.7) |
| Nehme et al | Europe | 243 | 5.6 | (3.6, 8.3) |
| O'Keefe et al | N America | 119 | 11.1 | (7.1, 16.3) |
| Osmanov et al | Europe | 270 | 6.9 | (4.9, 9.4) |
| Peluso et al | N America | 112 | 21.0 | (14.1, 29.4) |
| Radtke et al | Europe | 84 | 1.8 | (0.2, 6.5) |
| Rass et al | Europe | 90 | 34.0 | (26.1, 42.7) |
| Romero-Duarte et al | Europe | 196 | 4.9 | (3.5, 6.6) |
| Seeßle et al | Europe | 147 | 29.4 | (22.2, 37.6) |
| Shang et al | China | 196 | 23.2 | (20.3, 26.3) |
| Sigfrid et al | Europe | 222 | 46.2 | (40.7, 51.7) |
| Sonnweber et al | Europe | 103 | 28.0 | (20.6, 36.5) |
| Sykes et al | Europe | 127 | 33.9 | (25.7, 42.8) |
| Tawfik et al | Other | 91 | 15.0 | (9.1, 22.7) |
| Tempany et al | Europe | 84 | 30.9 | (24.8, 37.5) |
| Tleyjeh et al | Other | 136 | 5.0 | (2.5, 8.8) |
| Ursini et al | Europe | 182 | 32.3 | (28.6, 36.2) |
| Xiong et al | China | 111 | 17.7 | (14.6, 21.2) |
| **95% Prediction Interval** | | | | |

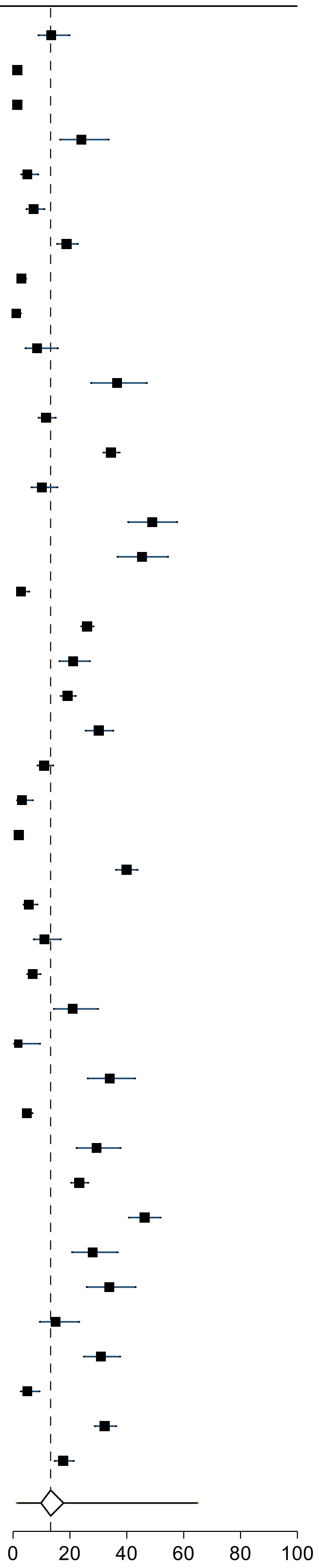

Supplementary Figure 29: Forest plot of prevalence of Long Covid in studies that reported speech or language problems as a persistent symptom with 95% prediction intervals

| study | region | mean days since infection | % Persistent Speech or Language Problems | |
|---|---|---|---|---|
| Becker et al | N America | 231 | 15.0 | (12.5, 17.8) |
| Munblit et al | Europe | 232 | 0.8 | (0.5, 1.2) |
| Osmanov et al | Europe | 270 | 0.2 | (0.0, 1.1) |
| PHOSP-COVID study | Europe | 174 | 16.7 | (13.9, 19.7) |
| Rass et al | Europe | 90 | 2.0 | (0.4, 5.7) |
| Sigfrid et al | Europe | 222 | 16.2 | (12.4, 20.7) |
| **95% Prediction Interval** | | | | |

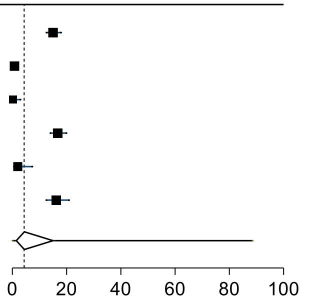

Supplementary Figure 30: Forest plot of prevalence of Long Covid in studies that reported problems with taste or smell as a persistent symptom with 95% prediction intervals

| study | region | mean days since infection | % Persistent Problems with Taste or Smell | |
|---|---|---|---|---|
| Abdelrahman et al | Other | 270 | 7.0 | (3.7, 11.9) |
| Augustin et al | Europe | 131 | 12.4 | (9.5, 15.8) |
| Bellan et al | Europe | 120 | 5.0 | (2.6, 8.6) |
| Bliddal et al | Europe | 84 | 7.5 | (3.7, 13.4) |
| Blomberg et al | Europe | 183 | 27.0 | (21.6, 33.0) |
| Boscolo-Rizzo et al | Europe | 365 | 22.0 | (17.4, 27.0) |
| COMEBAC study | Europe | 127 | 5.2 | (3.4, 7.6) |
| Caruso et al | Europe | 182 | 15.3 | (9.3, 23.0) |
| Caspersen et al | Europe | 350 | 16.6 | (14.0, 19.4) |
| Castro et al | N America | 128 | 2.1 | (1.8, 2.5) |
| Cirulli et al | N America | 90 | 14.3 | (8.5, 21.9) |
| Cristillo et al | Europe | 196 | 49.5 | (39.4, 59.6) |
| Diaz-Fuentes et al | N America | 84 | 1.8 | (0.2, 6.4) |
| Domenech-Montoliu et al | Europe | 213 | 16.1 | (13.0, 19.7) |
| Fernandez-de-Las-Penas et al | Europe | 227 | 3.3 | (2.3, 4.5) |
| Gonzalez-Hermosillo et al | Other | 105 | 12.3 | (7.2, 19.2) |
| Havervall et al | Europe | 122 | 10.8 | (7.6, 14.7) |
| Huang et al | China | 186 | 11.0 | (9.5, 12.6) |
| Jacobson et al | N America | 119 | 21.4 | (14.3, 29.9) |
| Lemhofer et al | Europe | 91 | 17.3 | (13.6, 21.6) |
| Logue et al | N America | 169 | 13.6 | (8.9, 19.6) |
| Lucidi et al | Europe | 186 | 36.4 | (27.4, 46.1) |
| Lui et al | China | 96 | 5.4 | (2.7, 9.4) |
| Maestre-Muniz et al | Europe | 379 | 7.2 | (5.2, 9.7) |
| Mechi et al | Other | 274 | 8.0 | (3.7, 14.7) |
| Mei et al | China | 158 | 0.3 | (0.1, 0.6) |
| Millet et al | N America | 365 | 6.4 | (3.2, 11.2) |
| Munblit et al | Europe | 232 | 3.0 | (2.4, 3.7) |
| Nehme et al | Europe | 243 | 16.8 | (13.3, 20.8) |
| Nguyen et al | Europe | 213 | 24.0 | (16.8, 32.5) |
| O'Keefe et al | N America | 119 | 10.6 | (6.7, 15.8) |
| Ong et al | Other | 104 | 0.6 | (0.0, 3.3) |
| Orru et al | Europe | 91 | 30.9 | (23.7, 38.9) |
| Osmanov et al | Europe | 270 | 5.6 | (3.8, 7.9) |
| PHOSP-COVID study | Europe | 174 | 10.4 | (8.2, 13.0) |
| Peghin et al | Europe | 191 | 10.4 | (8.1, 13.1) |
| Peluso et al | N America | 112 | 15.0 | (9.0, 23.0) |
| Petersen et al | Europe | 125 | 23.0 | (17.1, 29.9) |
| Rass et al | Europe | 90 | 16.0 | (10.2, 23.5) |
| Riestra-Ayora et al | Europe | 182 | 26.7 | (20.6, 33.5) |
| Roessler et al (adults) | Europe | 91 | 1.5 | (1.5, 1.6) |
| Roessler et al (children) | Europe | 91 | 0.7 | (0.5, 0.8) |
| Romero-Duarte et al | Europe | 196 | 7.2 | (5.5, 9.2) |
| Sathyamurthy et al | Other | 104 | 0.8 | (0.1, 2.9) |
| Seeßle et al | Europe | 147 | 17.1 | (11.4, 24.2) |
| Sigfrid et al | Europe | 222 | 12.8 | (9.4, 16.9) |
| Skala et al | Europe | 91 | 20.6 | (13.2, 29.7) |
| Sonnweber et al | Europe | 103 | 19.0 | (12.6, 26.7) |
| Soraas et al | Europe | 126 | 14.0 | (11.5, 16.8) |
| Stavem et al | Europe | 117 | 12.0 | (9.1, 15.4) |
| Stephenson et al | Europe | 104 | 13.5 | (12.3, 14.8) |
| Sykes et al | Europe | 127 | 10.2 | (5.6, 16.9) |
| Tawfik et al | Other | 91 | 10.0 | (4.7, 18.1) |
| Taylor et al | Europe | 126 | 16.3 | (13.3, 19.7) |
| Tleyjeh et al | Other | 136 | 0.5 | (0.0, 2.8) |
| Trunfio et al | Europe | 194 | 7.7 | (4.2, 12.8) |
| Ursini et al | Europe | 182 | 25.5 | (22.1, 29.1) |
| Venturelli et al | Europe | 105 | 3.0 | (1.9, 4.5) |
| Zayet et al | Europe | 289 | 26.6 | (22.1, 31.6) |
| Zhang et al | China | 378 | 1.4 | (1.0, 1.9) |
| **95% Prediction Interval** | | | | |

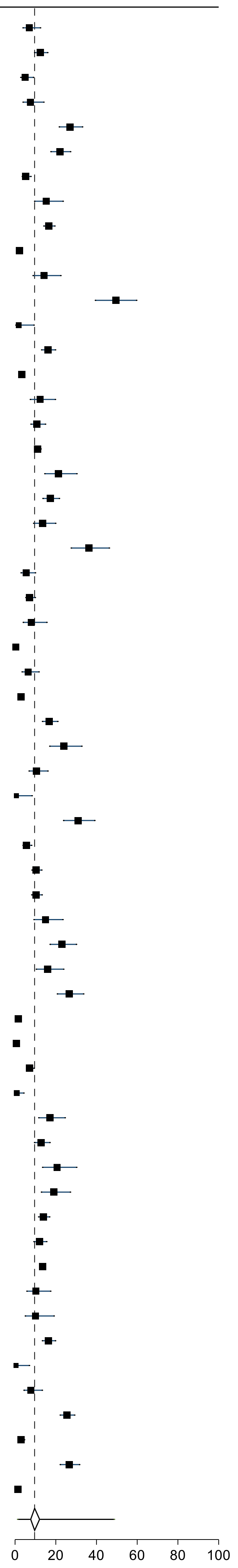

Supplementary Figure 31: Forest plot of prevalence of Long Covid in studies that reported sore throat as a persistent symptom with 95% prediction intervals

| study | region | mean days since infection | % Persistent Sore Throat | |
|---|---|---|---|---|
| Bliddal et al | Europe | 84 | 3.5 | (1.1, 8.0) |
| Boscolo-Rizzo et al | Europe | 365 | 2.0 | (0.7, 4.3) |
| Domenech-Montoliu et al | Europe | 213 | 5.8 | (3.9, 8.3) |
| Huang et al | China | 186 | 4.0 | (3.1, 5.0) |
| Jacobson et al | N America | 119 | 2.6 | (0.5, 7.4) |
| Logue et al | N America | 169 | 2.8 | (0.9, 6.4) |
| Maestre-Muniz et al | Europe | 379 | 2.2 | (1.1, 3.8) |
| Mei et al | China | 158 | 1.1 | (0.8, 1.5) |
| Nehme et al | Europe | 243 | 2.4 | (1.2, 4.4) |
| O'Keefe et al | N America | 119 | 0.5 | (0.0, 2.8) |
| Orru et al | Europe | 91 | 17.8 | (12.0, 24.8) |
| Petersen et al | Europe | 125 | 2.0 | (0.5, 5.0) |
| Seeßle et al | Europe | 147 | 4.8 | (1.9, 9.6) |
| Shang et al | China | 196 | 1.4 | (0.7, 2.5) |
| Soraas et al | Europe | 126 | 7.0 | (5.2, 9.2) |
| Stavem et al | Europe | 117 | 4.9 | (3.1, 7.3) |
| Stephenson et al | Europe | 104 | 9.5 | (8.5, 10.6) |
| Sykes et al | Europe | 127 | 11.8 | (6.8, 18.7) |
| Taquet et al | N America | 90 | 5.7 | (5.6, 5.8) |
| Tleyjeh et al | Other | 136 | 0.5 | (0.0, 2.8) |
| Zayet et al | Europe | 289 | 2.5 | (1.1, 4.7) |
| Zhang et al | China | 378 | 1.0 | (0.6, 1.5) |
| **95% Prediction Interval** | | | | |

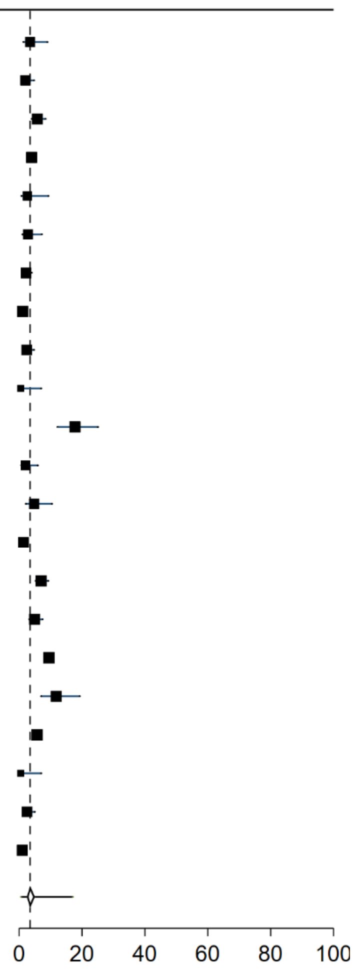

Supplementary Figure 32: Forest plot of prevalence of Long Covid in studies that reported tingling or itching as a persistent symptom with 95% prediction intervals

| study | region | mean days since infection | % Persistent Tingling or Itching | |
|---|---|---|---|---|
| Abdelrahman et al | Other | 270 | 7.6 | (4.1, 12.6) |
| Blomberg et al | Europe | 183 | 4.0 | (1.8, 7.5) |
| COMEBAC study | Europe | 127 | 10.7 | (8.1, 13.8) |
| Cirulli et al | N America | 90 | 7.5 | (3.5, 13.8) |
| Cristillo et al | Europe | 196 | 21.8 | (14.2, 31.1) |
| Gonzalez-Hermosillo et al | Other | 105 | 46.9 | (38.1, 55.9) |
| Munblit et al | Europe | 232 | 3.5 | (2.8, 4.3) |
| Orru et al | Europe | 91 | 34.9 | (27.3, 43.0) |
| Osmanov et al | Europe | 270 | 0.4 | (0.1, 1.5) |
| PHOSP-COVID study | Europe | 174 | 37.2 | (33.5, 41.0) |
| Rass et al | Europe | 90 | 25.0 | (17.4, 33.9) |
| Romero-Duarte et al | Europe | 196 | 3.4 | (2.3, 4.9) |
| Tawfik et al | Other | 91 | 7.5 | (3.5, 13.8) |
| Ursini et al | Europe | 182 | 20.1 | (17.0, 23.5) |
| **95% Prediction Interval** | | | | |

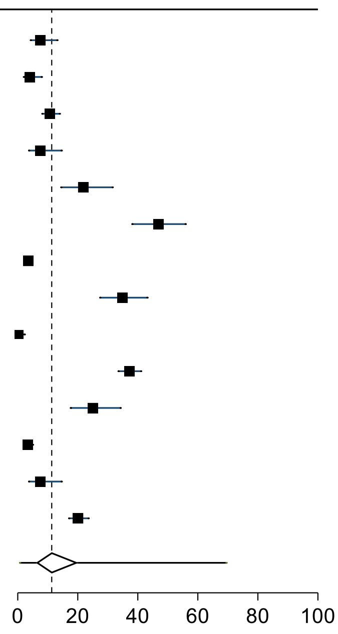

Supplementary Figure 33: Forest plot of prevalence of Long Covid in studies that reported vascular problems as a persistent symptom with 95% prediction intervals

| study | region | mean days since infection | % Persistent Vascular Problems | |
|---|---|---|---|---|
| Al-Aly et al (hospitalized) | N America | 150 | 1.3 | (1.1, 1.5) |
| Al-Aly et al (non-hospitalized) | N America | 126 | 0.3 | (0.2, 0.3) |
| Roessler et al (adults) | Europe | 91 | 0.9 | (0.8, 0.9) |
| Roessler et al (children) | Europe | 91 | 0.3 | (0.2, 0.4) |
| Romero-Duarte et al | Europe | 196 | 4.4 | (3.1, 6.1) |
| **95% Prediction Interval** | | | | |

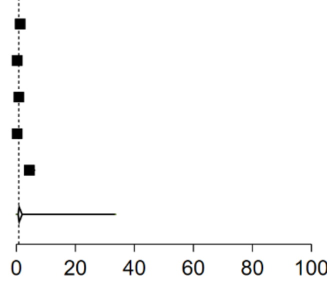

Supplementary Figure 34: Forest plot of prevalence of Long Covid in studies that reported weakness as a persistent symptom with 95% prediction intervals

| study | region | mean days since infection | % Persistent Weakness |
|---|---|---|---|
| Bellan et al | Europe | 120 | 53.8 (47.2, 60.2) |
| COMEBAC study | Europe | 127 | 8.4 (6.1, 11.3) |
| Cirulli et al | N America | 90 | 12.1 (6.8, 19.4) |
| Cristillo et al | Europe | 196 | 28.7 (20.1, 38.6) |
| Domenech-Montoliu et al | Europe | 213 | 8.3 (6.0, 11.1) |
| Lemhofer et al | Europe | 91 | 18.4 (14.6, 22.8) |
| Maestre-Muniz et al | Europe | 379 | 7.4 (5.3, 9.9) |
| Mei et al | China | 158 | 0.1 (0.0, 0.4) |
| Menges et al | Europe | 219 | 11.2 (8.4, 14.6) |
| Munblit et al | Europe | 232 | 10.3 (9.2, 11.5) |
| O'Keefe et al | N America | 119 | 6.1 (3.2, 10.4) |
| Ong et al | Other | 104 | 0.6 (0.0, 3.3) |
| Osmanov et al | Europe | 270 | 0.4 (0.0, 1.4) |
| PHOSP-COVID study | Europe | 174 | 46.3 (42.4, 50.1) |
| Peluso et al | N America | 112 | 15.0 (9.1, 22.7) |
| Qin et al | China | 105 | 13.4 (10.9, 16.3) |
| Rass et al | Europe | 90 | 5.0 (2.0, 10.0) |
| Romero-Duarte et al | Europe | 196 | 3.4 (2.3, 4.9) |
| Sigfrid et al | Europe | 222 | 37.6 (32.3, 43.1) |
| Todt et al | Other | 105 | 14.0 (9.9, 19.0) |
| Walle-Hansen et al | Europe | 200 | 33.0 (24.2, 42.8) |
| **95% Prediction Interval** | | | |

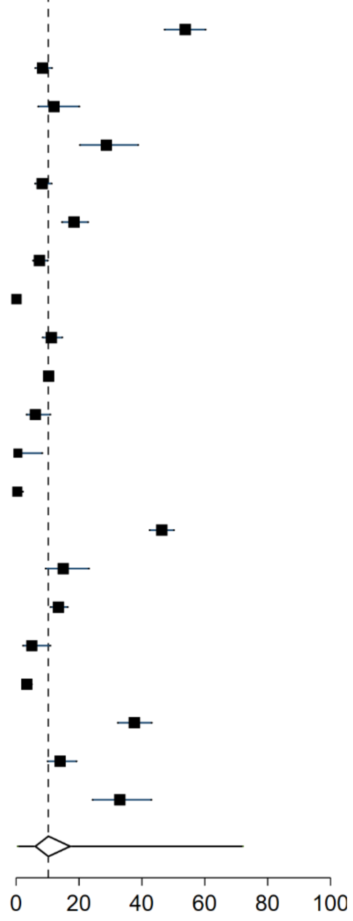

Supplementary Figure 35: Forest plot of prevalence of Long Covid in studies that reported on heart pathology with 95% prediction intervals

| study | region | mean days since infection | % Persistent Heart Pathology | |
|---|---|---|---|---|
| Al-Aly et al (hospitalized) | N America | 150 | 0.8 | (0.6, 0.9) |
| Al-Aly et al (non-hospitalized) | N America | 126 | 0.8 | (0.8, 0.9) |
| Aminian et al | N America | 243 | 22.0 | (20.5, 23.6) |
| Ayoubkhani et al | Europe | 154 | 2.6 | (2.4, 2.8) |
| Gonzalez-Hermosillo et al | Other | 105 | 40.8 | (32.2, 49.7) |
| Mei et al | China | 158 | 7.6 | (6.8, 8.5) |
| Roessler et al (adults) | Europe | 91 | 2.8 | (2.7, 2.8) |
| Roessler et al (children) | Europe | 91 | 0.7 | (0.6, 0.9) |
| Sonnweber et al | Europe | 103 | 55.0 | (46.0, 63.5) |
| Tohamy et al | Other | 105 | 2.0 | (0.2, 7.0) |
| Ursini et al | Europe | 182 | 14.6 | (11.9, 17.7) |
| Xiong et al | China | 111 | 13.0 | (10.3, 16.2) |
| **95% Prediction Interval** | | | | |

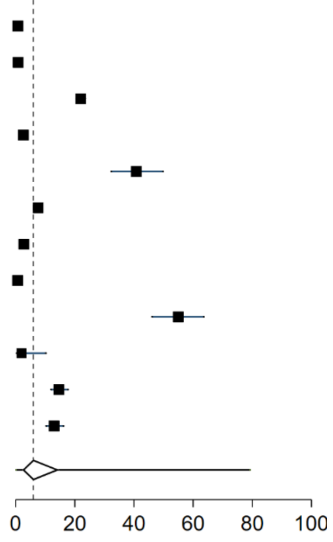

Supplementary Figure 36: Forest plot of prevalence of Long Covid in studies that reported on kidney pathology with 95% prediction intervals

| study | region | mean days since infection | % Persistent Kidney Pathology | |
|---|---|---|---|---|
| Aminian et al | N America | 243 | 6.3 | (5.4, 7.3) |
| Ayoubkhani et al | Europe | 154 | 0.6 | (0.5, 0.7) |
| Mei et al | China | 158 | 0.3 | (0.2, 0.5) |
| Roessler et al (adults) | Europe | 91 | 0.4 | (0.3, 0.4) |
| Roessler et al (children) | Europe | 91 | 0.1 | (0.0, 0.2) |
| Romero-Duarte et al | Europe | 196 | 4.5 | (3.2, 6.2) |
| **95% Prediction Interval** | | | | |

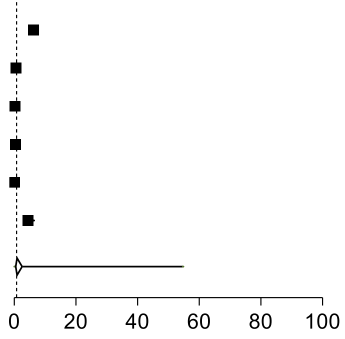

Supplementary Figure 37: Forest plot of prevalence of Long Covid in studies that reported on liver pathology with 95% prediction intervals

| study | region | mean days since infection | % Persistent Liver Pathology | |
|---|---|---|---|---|
| Aminian et al | N America | 243 | 2.7 | (2.1, 3.4) |
| Ayoubkhani et al | Europe | 154 | 0.2 | (0.2, 0.3) |
| Mei et al | China | 158 | 0.4 | (0.2, 0.6) |
| **95% Prediction Interval** | | | | |

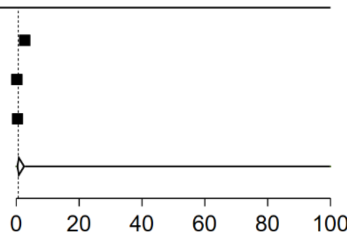

Supplementary Figure 38: Forest plot of prevalence of Long Covid in studies that reported on lung pathology with 95% prediction intervals

| study | region | mean days since infection | % Persistent Lung Pathology | |
|---|---|---|---|---|
| Al-Aly et al (hospitalized) | N America | 150 | 9.2 | (8.7, 9.8) |
| Al-Aly et al (non-hospitalized) | N America | 126 | 1.2 | (1.0, 1.3) |
| Aminian et al | N America | 243 | 22.9 | (21.4, 24.5) |
| Arnold et al | Europe | 90 | 14.0 | (8.1, 22.1) |
| Ayoubkhani et al | Europe | 154 | 21.5 | (21.0, 22.0) |
| Blanco et al | Europe | 104 | 52.0 | (41.8, 62.1) |
| Caruso et al | Europe | 182 | 72.0 | (63.0, 79.9) |
| Caspersen et al | Europe | 350 | 7.4 | (5.7, 9.5) |
| Diaz-Fuentes et al | N America | 84 | 46.8 | (35.3, 58.5) |
| Frija-Masson et al | Europe | 91 | 75.2 | (67.1, 82.2) |
| Froidure et al | Europe | 95 | 67.0 | (57.3, 75.7) |
| Han et al | China | 175 | 62.3 | (52.7, 71.2) |
| Li et al | China | 120 | 59.9 | (54.0, 65.6) |
| Liao et al | China | 409 | 37.3 | (31.8, 43.0) |
| Liao et al | China | 104 | 85.9 | (79.1, 91.2) |
| Mei et al | China | 158 | 9.2 | (8.3, 10.2) |
| Milanese et al | Europe | 196 | 47.0 | (38.4, 55.8) |
| Nabahati et al | Other | 104 | 52.0 | (44.3, 59.7) |
| Nunez-Fernandez et al | Europe | 98 | 29.0 | (22.8, 35.8) |
| Sibila et al | Europe | 116 | 57.0 | (49.2, 64.5) |
| Skala et al | Europe | 91 | 47.0 | (36.9, 57.2) |
| Skjorten et al | Europe | 105 | 31.0 | (23.9, 38.8) |
| Yan et al | China | 379 | 39.0 | (30.1, 48.1) |
| Yin et al | China | 203 | 55.8 | (50.2, 61.4) |
| Zhang et al | China | 105 | 72.7 | (66.6, 78.1) |
| Zhang et al | China | 106 | 54.9 | (45.7, 63.9) |
| **95% Prediction Interval** | | | | |

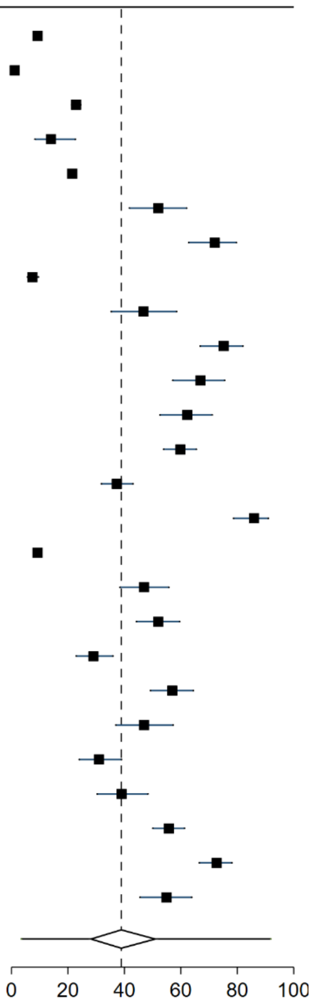

Supplementary Figure 39: Forest plot of prevalence of Long Covid in studies that reported on neurological pathology with 95% prediction intervals

| study | region | mean days since infection | % Persistent Neurological pathology | |
|---|---|---|---|---|
| Al-Aly et al (hospitalized) | N America | 150 | 4.0 | (3.6, 4.3) |
| Al-Aly et al (non-hospitalized) | N America | 126 | 1.9 | (1.8, 2.0) |
| Aminian et al | N America | 243 | 9.0 | (8.0, 10.1) |
| Cristillo et al | Europe | 196 | 26.7 | (18.4, 36.5) |
| Mei et al | China | 158 | 7.9 | (7.0, 8.8) |
| Nehme et al | Europe | 243 | 2.2 | (1.0, 4.1) |
| Peghin et al | Europe | 191 | 9.6 | (7.3, 12.3) |
| Rass et al | Europe | 90 | 15.0 | (9.4, 22.3) |
| Roessler et al (adults) | Europe | 91 | 3.2 | (3.2, 3.3) |
| Roessler et al (children) | Europe | 91 | 1.7 | (1.5, 2.0) |
| Tohamy et al | Other | 105 | 2.0 | (0.2, 7.0) |
| **95% Prediction Interval** | | | | |

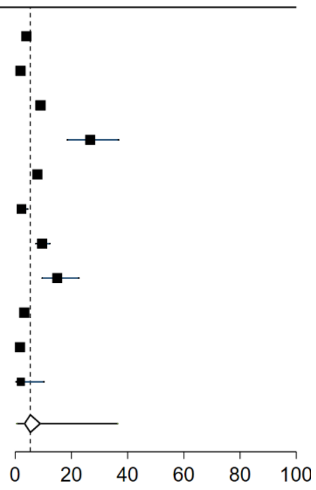

Supplementary Figure 40: Forest plot of prevalence of Long Covid in studies that reported on pancreatic pathology with 95% prediction intervals

| study | region | mean days since infection | % Persistent Pancreas Pathology | |
|---|---|---|---|---|
| Ayoubkhani et al | Europe | 154 | 1.1 | (1.0, 1.2) |
| Mei et al | China | 158 | 2.4 | (1.9, 2.9) |
| Romero-Duarte et al | Europe | 196 | 0.9 | (0.4, 1.8) |
| **95% Prediction Interval** | | | | |

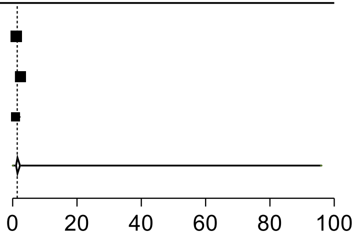

Supplementary Figure 41: Prevalence of Long Covid by risk of bias domains with 95% prediction intervals

| Risk of bias domain | % persistent symptoms | number of studies | $I^2$ |
|---|---|---|---|
| **Selection 1: All participants have confirmed diagnosis** | | | |
| 1 | 40.3 (6.3 to 87.0) | 98 | 99.8% |
| 0.5 | 49.1 (5.5 to 94.1) | 18 | 99.6% |
| 0 | 51.9 (14.7 to 87.1) | 6 | 97.2% |
| **Selection 2: Representative sample** | | | |
| 1 | 23.2 (2.3 to 79.7) | 23 | 100.0% |
| 0 | 47.3 (10.2 to 87.6) | 99 | 99.6% |
| **Selection 3: Internal or external non-Covid comparator** | | | |
| 1 | 34.7 (3.6 to 88.5) | 58 | 99.9% |
| 0 | 50.0 (14.9 to 85.1) | 63 | |
| **Comparability: All participants drawn from same source** | | | |
| 1 | 42.4 (6.9 to 88.0) | 113 | 99.9% |
| 0 | 38.5 (2.8 to 93.1) | 9 | 99.2% |
| **Outcome 1: All participants assessed in the same way** | | | |
| 1 | 38.8 (6.0 to 86.4) | 100 | 99.9% |
| 0 | 57.4 (14.6 to 91.4) | 22 | 98.2% |
| **Outcome 2: Standardised assessment of symptoms** | | | |
| 1 | 42.3 (6.9 to 87.9) | 116 | 99.9% |
| 0 | 32.3 (0.1 to 99.4) | 6 | 99.0% |
| **Outcome 3: Comprehensive range of symptoms** | | | |
| 1 | 42.7 (6.1 to 89.5) | 87 | 99.9% |
| 0 | 41.0 (8.8 to 83.4) | 35 | 98.9% |
| **Outcome 4: All participants have same length of follow-up** | | | |
| 1 | 39.5 (5.5 to 88.0) | 81 | 99.9% |
| 0 | 47.7 (10.6 to 87.5) | 41 | 99.1% |
| **Outcome 5: Losses to follow-up unlikely to introduce bias** | | | |
| 1 | 40.8 (5.3 to 89.4) | 81 | 99.9% |
| 0 | 44.6 (10.1 to 85.1) | 41 | 98.5% |
| **Outcome 6: Differential effect of interventions considered** | | | |
| 1 | 51.1 (5.0 to 95.4) | 7 | 98.7% |
| 0 | 41.6 (6.6 to 87.8) | 115 | 99.9% |

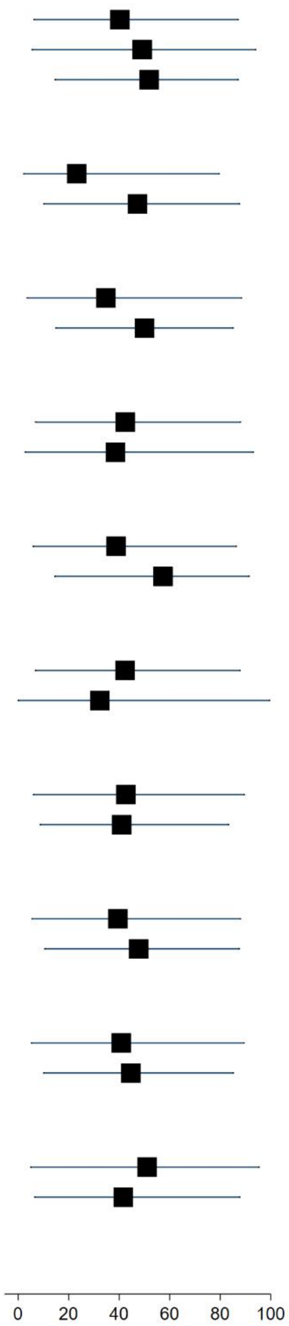

Supplementary Figure 42: Forest plot of prevalence of Long Covid in all included studies by sample recruitment source with 95% prediction intervals

| study | region | mean days since infection | cases | total | % persistent symptoms |
|---|---|---|---|---|---|
| **community** | | | | | |
| Al-Aly et al (non-hospitalized) | N America | 126 | 1718 | 60255 | 2.9 (2.7, 3.0) |
| Augustin et al | Europe | 131 | 123 | 442 | 27.8 (23.7, 32.3) |
| Bliddal et al | Europe | 84 | 52 | 129 | 40.3 (31.8, 49.3) |
| Boscolo-Rizzo et al | Europe | 365 | 161 | 304 | 53.0 (47.2, 58.7) |
| Caspersen et al | Europe | 350 | 28 | 170 | 16.5 (11.2, 22.9) |
| Cirulli et al | N America | 90 | 18 | 122 | 14.8 (9.0, 22.3) |
| Domenech-Montoliu et al | Europe | 213 | 258 | 483 | 53.4 (48.9, 57.9) |
| Lemhofer et al | Europe | 91 | 226 | 365 | 61.9 (56.7, 66.9) |
| Menges et al | Europe | 219 | 106 | 431 | 24.6 (20.6, 28.9) |
| ONS study August 2021 | Europe | 84 | 2501 | 21374 | 11.7 (11.3, 12.1) |
| Petersen et al | Europe | 125 | 95 | 180 | 52.8 (45.2, 60.2) |
| Radtke et al | Europe | 84 | 4 | 109 | 3.7 (1.0, 9.1) |
| Roessler et al (adults) | Europe | 91 | 13396 | 145184 | 9.2 (9.1, 9.4) |
| Roessler et al (children) | Europe | 91 | 734 | 11950 | 6.1 (5.7, 6.6) |
| Sonnweber et al | Europe | 103 | 73 | 133 | 54.9 (46.0, 63.5) |
| Soraas et al | Europe | 126 | 380 | 676 | 56.2 (52.4, 60.0) |
| Stavem et al | Europe | 117 | 185 | 451 | 41.0 (36.4, 45.7) |
| Stephenson et al | Europe | 104 | 2038 | 3065 | 66.5 (64.8, 68.2) |
| Sudre et al | Other | 84 | 108 | 4182 | 2.6 (2.1, 3.1) |
| Taquet et al | N America | 90 | 100007 | 273618 | 36.5 (36.4, 36.7) |
| Whitaker et al | Europe | 84 | 28713 | 76155 | 37.7 (37.4, 38.0) |
| | | | | | **25.6 (1.8, 86.3)** |
| **healthcare workers** | | | | | |
| Gaber et al | Europe | 122 | 61 | 138 | 44.2 (35.8, 52.9) |
| Havervall et al | Europe | 122 | 69 | 323 | 21.4 (17.0, 26.2) |
| Liao et al | China | 409 | 113 | 303 | 37.3 (31.8, 43.0) |
| Martinez et al | Europe | 168 | 69 | 260 | 26.5 (21.3, 32.3) |
| Riestra-Ayora et al | Europe | 182 | 52 | 195 | 26.7 (20.6, 33.5) |
| Tawfik et al | Other | 91 | 40 | 120 | 33.3 (25.0, 42.5) |
| Tempany et al | Europe | 84 | 116 | 217 | 53.5 (46.6, 60.2) |
| Xiong et al | China | 167 | 114 | 162 | 70.4 (62.7, 77.3) |
| | | | | | **38.4 (9.3, 79.0)** |
| **outpatients** | | | | | |
| Abdelrahman et al | Other | 270 | 105 | 172 | 61.0 (53.3, 68.4) |
| Aminian et al | N America | 243 | 1255 | 2839 | 44.2 (42.4, 46.1) |
| Becker et al | N America | 231 | 178 | 740 | 24.1 (21.0, 27.3) |
| Blomberg et al | Europe | 183 | 189 | 312 | 60.6 (54.9, 66.0) |
| Jacobson et al | N America | 119 | 79 | 118 | 66.9 (57.7, 75.3) |
| Kim et al | Other | 195 | 591 | 900 | 65.7 (62.5, 68.8) |
| Logue et al | N America | 169 | 53 | 177 | 30.0 (23.3, 37.3) |
| Matteudi et al | Europe | 180 | 23 | 137 | 16.8 (11.0, 24.1) |
| Mechi et al | Other | 274 | 92 | 112 | 82.1 (73.8, 88.7) |
| Mohiuddin et al | Other | 161 | 67 | 313 | 21.4 (17.0, 26.4) |
| O'Keefe et al | N America | 119 | 79 | 198 | 39.9 (33.0, 47.1) |
| Peghin et al | Europe | 191 | 241 | 599 | 40.2 (36.3, 44.3) |
| Seeßle et al | Europe | 147 | 107 | 146 | 73.3 (65.3, 80.3) |
| Skala et al | Europe | 91 | 56 | 102 | 54.9 (44.7, 64.8) |
| Tohamy et al | Other | 105 | 5 | 100 | 5.0 (1.6, 11.3) |
| Townsend et al | Europe | 86 | 74 | 128 | 57.8 (48.8, 66.5) |
| Venturelli et al | Europe | 105 | 394 | 767 | 51.4 (47.8, 55.0) |
| | | | | | **45.5 (8.4, 88.4)** |
| **social media** | | | | | |
| Orru et al | Europe | 91 | 113 | 152 | 74.3 (66.6, 81.1) |
| Peluso et al | N America | 112 | 89 | 143 | 62.2 (53.8, 70.2) |
| Ursini et al | Europe | 182 | 270 | 616 | 43.8 (39.9, 47.9) |
| | | | | | **60.4 (0.0, 100.0)** |
| **hospitalised** | | | | | |
| Al-Aly et al (hospitalized) | N America | 150 | 1091 | 11800 | 9.2 (8.7, 9.8) |
| Arnold et al | Europe | 90 | 81 | 110 | 73.6 (64.4, 81.6) |
| Ayoubkhani et al | Europe | 154 | 6085 | 28335 | 21.5 (21.0, 22.0) |
| Baricich et al | Europe | 139 | 66 | 204 | 32.4 (26.0, 39.2) |
| Bellan et al | Europe | 120 | 128 | 238 | 53.8 (47.2, 60.2) |
| Blanco et al | Europe | 104 | 52 | 100 | 52.0 (41.8, 62.1) |
| COMEBAC study | Europe | 127 | 244 | 478 | 51.0 (46.5, 55.6) |
| Carrillo-Garcia et al | Europe | 105 | 100 | 151 | 66.2 (58.1, 73.7) |
| Caruso et al | Europe | 182 | 91 | 118 | 77.1 (68.5, 84.3) |
| Castro et al | N America | 128 | 721 | 6619 | 10.9 (10.2, 11.7) |
| Chai et al | China | 378 | 156 | 546 | 28.6 (24.8, 32.6) |
| Clavario et al | Europe | 121 | 160 | 200 | 80.0 (73.8, 85.3) |
| Cristillo et al | Europe | 196 | 50 | 101 | 49.5 (39.4, 59.6) |
| Diaz-Fuentes et al | N America | 84 | 88 | 111 | 79.3 (70.5, 86.4) |
| Erol et al | Other | 170 | 45 | 121 | 37.2 (28.6, 46.4) |
| Fernandez-de-Las-Penas et al | Europe | 227 | 930 | 1142 | 81.4 (79.1, 83.7) |
| Frija-Masson et al | Europe | 91 | 103 | 137 | 75.2 (67.1, 82.2) |
| Froidure et al | Europe | 95 | 73 | 107 | 68.2 (58.5, 76.9) |
| Fu et al | China | 196 | 20 | 199 | 10.1 (6.2, 15.1) |
| Garcia-Abellan et al | Europe | 182 | 28 | 116 | 24.1 (16.7, 33.0) |
| Gonzalez-Hermosillo et al | Other | 105 | 119 | 130 | 91.5 (85.4, 95.7) |
| Han et al | China | 175 | 71 | 114 | 62.3 (52.7, 71.2) |
| Huang et al | China | 186 | 1265 | 1655 | 76.4 (74.3, 78.5) |
| Liao et al | China | 104 | 122 | 142 | 85.9 (79.1, 91.2) |
| Liu et al | China | 196 | 373 | 1301 | 28.7 (26.2, 31.2) |
| Liyanage-Don et al | N America | 113 | 99 | 153 | 64.7 (56.6, 72.3) |
| Lui et al | China | 96 | 41 | 204 | 20.1 (14.8, 26.3) |
| Maestre-Muniz et al | Europe | 379 | 309 | 543 | 56.9 (52.6, 61.1) |
| Mazza et al | Europe | 104 | 81 | 226 | 35.8 (29.6, 42.5) |
| Mei et al | China | 158 | 976 | 3677 | 26.5 (25.1, 28.0) |
| Milanese et al | Europe | 196 | 64 | 135 | 47.4 (38.8, 56.2) |
| Millet et al | N America | 365 | 83 | 173 | 48.0 (40.3, 55.7) |
| Munblit et al | Europe | 232 | 1534 | 2649 | 57.9 (56.0, 59.8) |
| Nabahati et al | Other | 104 | 90 | 173 | 52.0 (44.3, 59.7) |
| Nguyen et al | Europe | 213 | 30 | 125 | 24.0 (16.8, 32.5) |
| Nunez-Fernandez et al | Europe | 98 | 58 | 200 | 29.0 (22.8, 35.8) |
| Ong et al | Other | 104 | 13 | 175 | 7.4 (4.0, 12.4) |
| Osmanov et al | Europe | 270 | 126 | 519 | 24.3 (20.6, 28.2) |
| PHOSP-COVID study | Europe | 174 | 797 | 861 | 92.6 (90.6, 94.2) |
| Qin et al | China | 105 | 87 | 647 | 13.4 (10.9, 16.3) |
| Qu et al | China | 105 | 176 | 540 | 32.6 (28.7, 36.7) |
| Rass et al | Europe | 90 | 82 | 135 | 60.7 (52.0, 69.0) |
| Righi et al | Europe | 84 | 83 | 421 | 19.7 (16.0, 23.8) |
| Romero-Duarte et al | Europe | 196 | 509 | 797 | 63.9 (60.4, 67.2) |
| Sathyamurthy et al | Other | 104 | 66 | 279 | 23.7 (18.8, 29.1) |
| Shang et al | China | 196 | 441 | 796 | 55.4 (51.9, 58.9) |
| Sibila et al | Europe | 116 | 98 | 172 | 57.0 (49.2, 64.5) |
| Sigfrid et al | Europe | 222 | 305 | 327 | 93.3 (90.0, 95.7) |
| Simani et al | Other | 197 | 12 | 120 | 10.0 (5.3, 16.8) |
| Skjorten et al | Europe | 105 | 59 | 126 | 46.8 (37.9, 55.9) |
| Sykes et al | Europe | 127 | 75 | 127 | 59.1 (50.0, 67.7) |
| Taboada et al | Europe | 189 | 87 | 183 | 47.5 (40.1, 55.0) |
| Tarsitani et al | Europe | 105 | 34 | 115 | 29.6 (21.4, 38.8) |
| Taylor et al | Europe | 126 | 261 | 545 | 47.9 (43.6, 52.2) |
| Tleyjeh et al | Other | 136 | 125 | 222 | 56.3 (49.5, 62.9) |
| Todt et al | Other | 105 | 96 | 239 | 40.2 (33.9, 46.7) |
| Trunfio et al | Europe | 194 | 41 | 168 | 24.4 (18.1, 31.6) |
| Walle-Hansen et al | Europe | 200 | 57 | 106 | 53.8 (43.8, 63.5) |
| Weng et al | China | 104 | 52 | 117 | 44.4 (35.3, 53.9) |
| Xiong et al | China | 111 | 267 | 538 | 49.6 (45.3, 53.9) |
| Yan et al | China | 379 | 47 | 119 | 39.5 (30.7, 48.9) |
| Yan et al | China | 98 | 0 | 125 | 0.0 (0.0, 2.9) |
| Yin et al | China | 203 | 176 | 316 | 55.7 (50.0, 61.3) |
| Zayet et al | Europe | 289 | 127 | 354 | 35.9 (30.9, 41.1) |
| Zhan et al | China | 348 | 36 | 121 | 29.8 (21.8, 38.7) |
| Zhang et al | China | 378 | 1095 | 2433 | 45.0 (43.0, 47.0) |
| Zhang et al | China | 106 | 67 | 122 | 54.9 (45.7, 63.9) |
| Zhang et al | China | 105 | 178 | 245 | 72.7 (66.6, 78.1) |
| Zhou et al | China | 127 | 114 | 164 | 69.5 (61.9, 76.5) |
| | | | | | **46.5 (8.2, 89.4)** |

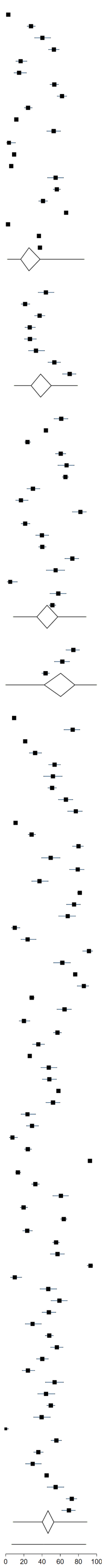

Supplementary Figure 43: Prevalence of Long Covid by risk factor or comorbidities with 95% prediction intervals

| Risk factor or comorbidity | % persistent symptoms | number of studies | $I^2$ |
|---|---|---|---|
| **Age** | | | |
| <50 years | 38.5 (7.9 to 82.1) | 41 | 99.1% |
| 50+ years | 47.7 (7.9 to 90.6) | 66 | 99.6% |
| **Gender** | | | |
| <50% female | 45.6 (5.5 to 92.4) | 59 | 99.6% |
| 50%+ female | 38.7 (8.5 to 81.2) | 58 | 99.9% |
| **Ethnicity** | | | |
| <50% white ethnicity | 56.3 (22.3 to 85.2) | 8 | 93.4% |
| 50%+ white ethnicity | 37.6 (1.7 to 95.3) | 16 | 100.0% |
| **Smoking status** | | | |
| <10% current smoker | 55.7 (11.4 to 92.4) | 21 | 99.9% |
| 10%+ current smoker | 50.5 (12.4 to 88.0) | 32 | 99.0% |
| **Pre-existing obesity** | | | |
| <20% pre-existing obesity | 37.5 (3.7 to 90.3) | 18 | 100.0% |
| 20%+ pre-existing obesity | 48.7 (5.5 to 94.0) | 19 | 99.9% |
| **Pre-existing diabetes** | | | |
| <10% pre-existing diabetes | 35.4 (5.7 to 83.2) | 34 | 99.8% |
| 10%+ pre-existing diabetes | 51.9 (8.3 to 92.8) | 41 | 99.8% |
| **Pre-existing hypertension** | | | |
| <30% pre-existing hypertension | 37.3 (7.0 to 82.5) | 42 | 99.8% |
| 30%+ pre-existing hypertension | 58.5 (16.9 to 90.7) | 30 | 99.7% |
| **Pre-existing cholesterol disorder** | | | |
| 5%+ pre-existing cholesterol disorder | 52.6 (18.3 to 84.6) | 13 | 96.7% |
| **Pre-existing CVD** | | | |
| <10% pre-existing CVD | 38.2 (5.9 to 85.9) | 39 | 99.8% |
| 10%+ pre-existing CVD | 54.7 (9.4 to 93.4) | 30 | 99.8% |
| **Pre-existing cancer** | | | |
| <5% pre-existing cancer | 44.9 (6.0 to 91.2) | 34 | 99.9% |
| 5%+ pre-existing cancer | 43.9 (11.1 to 83.1) | 13 | 99.8% |
| **Pre-existing respiratory disease** | | | |
| <10% pre-existing respiratory disease | 40.1 (6.4 to 86.6) | 40 | 99.6% |
| 10%+ pre-existing respiratory disease | 49.3 (7.4 to 92.2) | 29 | 99.9% |
| **Pre-existing liver disease** | | | |
| <5% pre-existing liver disease | 39.3 (3.4 to 92.2) | 17 | 100.0% |
| 5%+ pre-existing liver disease | 64.3 (12.3 to 95.9) | 5 | 96.2% |
| **Pre-existing kidney disease** | | | |
| <5% pre-existing kidney disease | 42.3 (7.8 to 86.4) | 23 | 99.6% |
| 5%+ pre-existing kidney disease | 61.2 (5.6 to 97.7) | 13 | 100.0% |
| **Pre-existing thyroid disorder** | | | |
| 5%+ pre-existing thyroid disorder | 36.2 (2.3 to 93.1) | 4 | 95.6% |
| **Pre-existing neurological disorder** | | | |
| <5% pre-existing neurological disorder | 31.2 (1.4 to 93.7) | 5 | 99.8% |
| 5%+ pre-existing neurological disorder | 34.5 (2.2 to 92.4) | 7 | 99.3% |
| **Pre-existing immunological disorder or allergy** | | | |
| <5% pre-existing immunological disorder or allergy | 27.8 (2.9 to 83.1) | 9 | 99.7% |
| 5%+ pre-existing immunological disorder or allergy | 52.0 (3.8 to 96.7) | 15 | 99.6% |
| **Pre-existing anxiety or depression** | | | |
| <10% pre-existing anxiety or depression | 43.2 (0.1 to 99.8) | 5 | 99.8% |

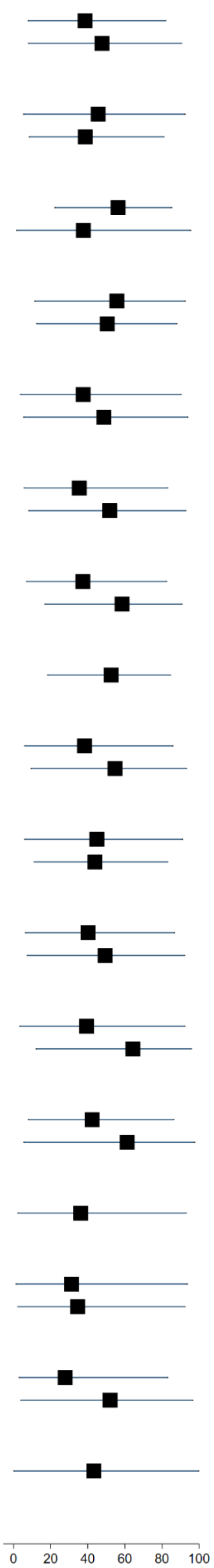

Supplementary Figure 44: Forest plot of prevalence of Long Covid in all included studies by domains of outcome assessment with 95% prediction intervals

| study | Dominant source of participants | Nature of outcome | Mean days since infection | cases | total | % persistent symptoms | |
|---|---|---|---|---|---|---|---|
| **single domain** | | | | | | | |
| Al-Aly et al (hospitalized) | hospitalised | outcomes from health record linkage | 150 | 1091 | 11800 | 9.2 | (8.7, 9.8) |
| Al-Aly et al (non-hospitalized) | community | outcomes from health record linkage | 126 | 1718 | 60255 | 2.9 | (2.7, 3.0) |
| Ayoubkhani et al | hospitalised | outcomes from health record linkage | 154 | 6085 | 28335 | 21.5 | (21.0, 22.0) |
| Baricich et al | hospitalised | symptoms & function discovered at follow-up | 139 | 66 | 204 | 32.4 | (26.0, 39.2) |
| Becker et al | outpatients | symptoms & function discovered at follow-up | 231 | 178 | 740 | 24.1 | (21.0, 27.3) |
| Blanco et al | hospitalised | pathology discovered at follow-up | 104 | 52 | 100 | 52.0 | (41.8, 62.1) |
| Caspersen et al | community | outcomes from health record linkage | 350 | 28 | 170 | 16.5 | (11.2, 22.9) |
| Castro et al | hospitalised | outcomes from health record linkage | 128 | 721 | 6619 | 10.9 | (10.2, 11.7) |
| Clavario et al | hospitalised | symptoms & function discovered at follow-up | 121 | 160 | 200 | 80.0 | (73.8, 85.3) |
| Cristillo et al | hospitalised | symptoms & function discovered at follow-up | 196 | 50 | 101 | 49.5 | (39.4, 59.6) |
| Fernandez-de-Las-Penas et al | hospitalised | symptoms & function discovered at follow-up | 227 | 930 | 1142 | 81.4 | (79.1, 83.7) |
| Frija-Masson et al | hospitalised | pathology discovered at follow-up | 91 | 103 | 137 | 75.2 | (67.1, 82.2) |
| Froidure et al | hospitalised | pathology discovered at follow-up | 95 | 73 | 107 | 68.2 | (58.5, 76.9) |
| Fu et al | hospitalised | symptoms & function discovered at follow-up | 196 | 20 | 199 | 10.1 | (6.2, 15.1) |
| Han et al | hospitalised | pathology discovered at follow-up | 175 | 71 | 114 | 62.3 | (52.7, 71.2) |
| Kashif et al | . | symptoms & function discovered at follow-up | 105 | 101 | 242 | 41.7 | (35.5, 48.2) |
| Li et al | . | pathology discovered at follow-up | 120 | 173 | 289 | 59.9 | (54.0, 65.6) |
| Liao et al | hospitalised | pathology discovered at follow-up | 104 | 122 | 142 | 85.9 | (79.1, 91.2) |
| Liao et al | healthcare workers | pathology discovered at follow-up | 409 | 113 | 303 | 37.3 | (31.8, 43.0) |
| Liu et al | hospitalised | symptoms & function discovered at follow-up | 196 | 373 | 1301 | 28.7 | (26.2, 31.2) |
| Lucidi et al | . | symptoms & function discovered at follow-up | 186 | 40 | 110 | 36.4 | (27.4, 46.1) |
| Mazza et al | hospitalised | symptoms & function discovered at follow-up | 104 | 81 | 226 | 35.8 | (29.6, 42.5) |
| Nabahati et al | hospitalised | pathology discovered at follow-up | 104 | 90 | 173 | 52.0 | (44.3, 59.7) |
| Nguyen et al | hospitalised | symptoms & function discovered at follow-up | 213 | 30 | 125 | 24.0 | (16.8, 32.5) |
| Nunez-Fernandez et al | hospitalised | pathology discovered at follow-up | 98 | 58 | 200 | 29.0 | (22.8, 35.8) |
| Orru et al | social media | symptoms & function discovered at follow-up | 91 | 113 | 152 | 74.3 | (66.6, 81.1) |
| Peluso et al | social media | symptoms & function discovered at follow-up | 112 | 89 | 143 | 62.2 | (53.8, 70.2) |
| Qin et al | hospitalised | pathology discovered at follow-up | 105 | 87 | 647 | 13.4 | (10.9, 16.3) |
| Qu et al | hospitalised | symptoms & function discovered at follow-up | 105 | 176 | 540 | 32.6 | (28.7, 36.7) |
| Rass et al | hospitalised | symptoms & function discovered at follow-up | 90 | 82 | 135 | 60.7 | (52.0, 69.0) |
| Riestra-Ayora et al | healthcare workers | symptoms & function discovered at follow-up | 182 | 52 | 195 | 26.7 | (20.6, 33.5) |
| Sibila et al | hospitalised | pathology discovered at follow-up | 116 | 98 | 172 | 57.0 | (49.2, 64.5) |
| Simani et al | hospitalised | symptoms & function discovered at follow-up | 197 | 12 | 120 | 10.0 | (5.3, 16.8) |
| Skjorten et al | hospitalised | symptoms & function discovered at follow-up | 105 | 59 | 126 | 46.8 | (37.9, 55.9) |
| Sonnweber et al | community | pathology discovered at follow-up | 103 | 73 | 133 | 54.9 | (46.0, 63.5) |
| Sykes et al | hospitalised | symptoms & function discovered at follow-up | 127 | 75 | 127 | 59.1 | (50.0, 67.7) |
| Tarsitani et al | hospitalised | symptoms & function discovered at follow-up | 105 | 34 | 115 | 29.6 | (21.4, 38.8) |
| Tawfik et al | healthcare workers | symptoms & function discovered at follow-up | 91 | 40 | 120 | 33.3 | (25.0, 42.5) |
| Taylor et al | hospitalised | symptoms & function discovered at follow-up | 126 | 261 | 545 | 47.9 | (43.6, 52.2) |
| Todt et al | hospitalised | symptoms & function discovered at follow-up | 105 | 96 | 239 | 40.2 | (33.9, 46.7) |
| Tohamy et al | outpatients | pathology discovered at follow-up | 105 | 5 | 100 | 5.0 | (1.6, 11.3) |
| Ursini et al | social media | symptoms & function discovered at follow-up | 182 | 270 | 616 | 43.8 | (39.9, 47.9) |
| Walle-Hansen et al | hospitalised | symptoms & function discovered at follow-up | 200 | 57 | 106 | 53.8 | (43.8, 63.5) |
| Weng et al | hospitalised | symptoms & function discovered at follow-up | 104 | 52 | 117 | 44.4 | (35.3, 53.9) |
| Xiong et al | healthcare workers | symptoms & function discovered at follow-up | 167 | 114 | 162 | 70.4 | (62.7, 77.3) |
| Xiong et al | hospitalised | symptoms & function discovered at follow-up | 111 | 267 | 538 | 49.6 | (45.3, 53.9) |
| Yan et al | hospitalised | pathology discovered at follow-up | 379 | 47 | 119 | 39.5 | (30.7, 48.9) |
| Yin et al | hospitalised | pathology discovered at follow-up | 203 | 176 | 316 | 55.7 | (50.0, 61.3) |
| Zhang et al | hospitalised | pathology discovered at follow-up | 106 | 67 | 122 | 54.9 | (45.7, 63.9) |
| Zhang et al | hospitalised | pathology discovered at follow-up | 105 | 178 | 245 | 72.7 | (66.6, 78.1) |
| Zhou et al | hospitalised | pathology discovered at follow-up | 127 | 114 | 164 | 69.5 | (61.9, 76.5) |
| | | | | | | **41.4** | **(6.9, 87.1)** |
| **multiple domains** | | | | | | | |
| Abdelrahman et al | outpatients | symptoms & function discovered at follow-up | 270 | 105 | 172 | 61.0 | (53.3, 68.4) |
| Aminian et al | outpatients | outcomes from health record linkage | 243 | 1255 | 2839 | 44.2 | (42.4, 46.1) |
| Arnold et al | hospitalised | symptoms & function discovered at follow-up | 90 | 81 | 110 | 73.6 | (64.4, 81.6) |
| Augustin et al | community | symptoms & function discovered at follow-up | 131 | 123 | 442 | 27.8 | (23.7, 32.3) |
| Bellan et al | hospitalised | pathology discovered at follow-up | 120 | 128 | 238 | 53.8 | (47.2, 60.2) |
| Bliddal et al | community | symptoms & function discovered at follow-up | 84 | 52 | 129 | 40.3 | (31.8, 49.3) |
| Blomberg et al | outpatients | symptoms & function discovered at follow-up | 183 | 189 | 312 | 60.6 | (54.9, 66.0) |
| Boscolo-Rizzo et al | community | symptoms & function discovered at follow-up | 365 | 161 | 304 | 53.0 | (47.2, 58.7) |
| COMEBAC study | hospitalised | symptoms & function discovered at follow-up | 127 | 244 | 478 | 51.0 | (46.5, 55.6) |
| Carrillo-Garcia et al | hospitalised | symptoms & function discovered at follow-up | 105 | 100 | 151 | 66.2 | (58.1, 73.7) |
| Caruso et al | hospitalised | symptoms & function discovered at follow-up | 182 | 91 | 118 | 77.1 | (68.5, 84.3) |
| Chai et al | hospitalised | symptoms & function discovered at follow-up | 378 | 156 | 546 | 28.6 | (24.8, 32.6) |
| Cirulli et al | community | symptoms & function discovered at follow-up | 90 | 18 | 122 | 14.8 | (9.0, 22.3) |
| Diaz-Fuentes et al | hospitalised | symptoms & function discovered at follow-up | 84 | 88 | 111 | 79.3 | (70.5, 86.4) |
| Domenech-Montoliu et al | community | symptoms & function discovered at follow-up | 213 | 258 | 483 | 53.4 | (48.9, 57.9) |
| Erol et al | hospitalised | symptoms & function discovered at follow-up | 170 | 45 | 121 | 37.2 | (28.6, 46.4) |
| Gaber et al | healthcare workers | symptoms & function discovered at follow-up | 122 | 61 | 138 | 44.2 | (35.8, 52.9) |
| Garcia-Abellan et al | hospitalised | symptoms & function discovered at follow-up | 182 | 28 | 116 | 24.1 | (16.7, 33.0) |
| Gonzalez-Hermosillo et al | hospitalised | symptoms & function discovered at follow-up | 105 | 119 | 130 | 91.5 | (85.4, 95.7) |
| Havervall et al | healthcare workers | symptoms & function discovered at follow-up | 122 | 69 | 323 | 21.4 | (17.0, 26.2) |
| Huang et al | hospitalised | symptoms & function discovered at follow-up | 186 | 1265 | 1655 | 76.4 | (74.3, 78.5) |
| Jacobson et al | outpatients | symptoms & function discovered at follow-up | 119 | 79 | 118 | 66.9 | (57.7, 75.3) |
| Kim et al | outpatients | symptoms & function discovered at follow-up | 195 | 591 | 900 | 65.7 | (62.5, 68.8) |
| Lemhofer et al | community | symptoms & function discovered at follow-up | 91 | 226 | 365 | 61.9 | (56.7, 66.9) |
| Liyanage-Don et al | hospitalised | symptoms & function discovered at follow-up | 113 | 99 | 153 | 64.7 | (56.6, 72.3) |
| Logue et al | outpatients | symptoms & function discovered at follow-up | 169 | 53 | 177 | 30.0 | (23.3, 37.3) |
| Lui et al | hospitalised | symptoms & function discovered at follow-up | 96 | 41 | 204 | 20.1 | (14.8, 26.3) |
| Maestre-Muniz et al | hospitalised | symptoms & function discovered at follow-up | 379 | 309 | 543 | 56.9 | (52.6, 61.1) |
| Martinez et al | healthcare workers | symptoms & function discovered at follow-up | 168 | 69 | 260 | 26.5 | (21.3, 32.3) |
| Matteudi et al | outpatients | symptoms & function discovered at follow-up | 180 | 23 | 137 | 16.8 | (11.0, 24.1) |
| Mechi et al | outpatients | symptoms & function discovered at follow-up | 274 | 92 | 112 | 82.1 | (73.8, 88.7) |
| Mei et al | hospitalised | symptoms & function discovered at follow-up | 158 | 976 | 3677 | 26.5 | (25.1, 28.0) |
| Menges et al | community | symptoms & function discovered at follow-up | 219 | 106 | 431 | 24.6 | (20.6, 28.9) |
| Milanese et al | hospitalised | pathology discovered at follow-up | 196 | 64 | 135 | 47.4 | (38.8, 56.2) |
| Millet et al | hospitalised | symptoms & function discovered at follow-up | 365 | 83 | 173 | 48.0 | (40.3, 55.7) |
| Mohiuddin et al | outpatients | symptoms & function discovered at follow-up | 161 | 67 | 313 | 21.4 | (17.0, 26.4) |
| Munblit et al | hospitalised | symptoms & function discovered at follow-up | 232 | 1534 | 2649 | 57.9 | (56.0, 59.8) |
| Nehme et al | . | symptoms & function discovered at follow-up | 243 | 160 | 410 | 39.0 | (34.3, 43.9) |
| O'Keefe et al | outpatients | symptoms & function discovered at follow-up | 119 | 79 | 198 | 39.9 | (33.0, 47.1) |
| ONS study August 2021 | community | symptoms & function discovered at follow-up | 84 | 2501 | 21374 | 11.7 | (11.3, 12.1) |
| Ong et al | hospitalised | symptoms & function discovered at follow-up | 104 | 13 | 175 | 7.4 | (4.0, 12.4) |
| Osmanov et al | hospitalised | symptoms & function discovered at follow-up | 270 | 126 | 519 | 24.3 | (20.6, 28.2) |
| PHOSP-COVID study | hospitalised | symptoms & function discovered at follow-up | 174 | 797 | 861 | 92.6 | (90.6, 94.2) |
| Peghin et al | outpatients | symptoms & function discovered at follow-up | 191 | 241 | 599 | 40.2 | (36.3, 44.3) |
| Petersen et al | community | symptoms & function discovered at follow-up | 125 | 95 | 180 | 52.8 | (45.2, 60.2) |
| Radtke et al | community | symptoms & function discovered at follow-up | 84 | 4 | 109 | 3.7 | (1.0, 9.1) |
| Righi et al | hospitalised | symptoms & function discovered at follow-up | 84 | 83 | 421 | 19.7 | (16.0, 23.8) |
| Roessler et al (adults) | community | outcomes from health record linkage | 91 | 13396 | 145184 | 9.2 | (9.1, 9.4) |
| Roessler et al (children) | community | outcomes from health record linkage | 91 | 734 | 11950 | 6.1 | (5.7, 6.6) |
| Romero-Duarte et al | hospitalised | symptoms & function discovered at follow-up | 196 | 509 | 797 | 63.9 | (60.4, 67.2) |
| Sathyamurthy et al | hospitalised | symptoms & function discovered at follow-up | 104 | 66 | 279 | 23.7 | (18.8, 29.1) |
| Seeßle et al | outpatients | symptoms & function discovered at follow-up | 147 | 107 | 146 | 73.3 | (65.3, 80.3) |
| Shang et al | hospitalised | symptoms & function discovered at follow-up | 196 | 441 | 796 | 55.4 | (51.9, 58.9) |
| Sigfrid et al | hospitalised | symptoms & function discovered at follow-up | 222 | 305 | 327 | 93.3 | (90.0, 95.7) |
| Skala et al | outpatients | symptoms & function discovered at follow-up | 91 | 56 | 102 | 54.9 | (44.7, 64.8) |
| Soraas et al | community | symptoms & function discovered at follow-up | 126 | 380 | 676 | 56.2 | (52.4, 60.0) |
| Stavem et al | community | symptoms & function discovered at follow-up | 117 | 185 | 451 | 41.0 | (36.4, 45.7) |
| Stephenson et al | community | symptoms & function discovered at follow-up | 104 | 2038 | 3065 | 66.5 | (64.8, 68.2) |
| Sudre et al | community | symptoms & function discovered at follow-up | 84 | 108 | 4182 | 2.6 | (2.1, 3.1) |
| Taboada et al | hospitalised | symptoms & function discovered at follow-up | 189 | 87 | 183 | 47.5 | (40.1, 55.0) |
| Taquet et al | community | outcomes from health record linkage | 90 | 100007 | 273618 | 36.5 | (36.4, 36.7) |
| Tempany et al | healthcare workers | symptoms & function discovered at follow-up | 84 | 116 | 217 | 53.5 | (46.6, 60.2) |
| Tleyjeh et al | hospitalised | symptoms & function discovered at follow-up | 136 | 125 | 222 | 56.3 | (49.5, 62.9) |
| Townsend et al | outpatients | symptoms & function discovered at follow-up | 86 | 74 | 128 | 57.8 | (48.8, 66.5) |
| Trunfio et al | hospitalised | symptoms & function discovered at follow-up | 194 | 41 | 168 | 24.4 | (18.1, 31.6) |
| Venturelli et al | outpatients | symptoms & function discovered at follow-up | 105 | 394 | 767 | 51.4 | (47.8, 55.0) |
| Whitaker et al | community | symptoms & function discovered at follow-up | 84 | 28713 | 76155 | 37.7 | (37.4, 38.0) |
| Yan et al | hospitalised | symptoms & function discovered at follow-up | 98 | 0 | 125 | 0.0 | (0.0, 2.9) |
| Zayet et al | hospitalised | symptoms & function discovered at follow-up | 289 | 127 | 354 | 35.9 | (30.9, 41.1) |
| Zhan et al | hospitalised | symptoms & function discovered at follow-up | 348 | 36 | 121 | 29.8 | (21.8, 38.7) |
| Zhang et al | hospitalised | symptoms & function discovered at follow-up | 378 | 1095 | 2433 | 45.0 | (43.0, 47.0) |
| | | | | | | **42.6** | **(6.3, 89.2)** |

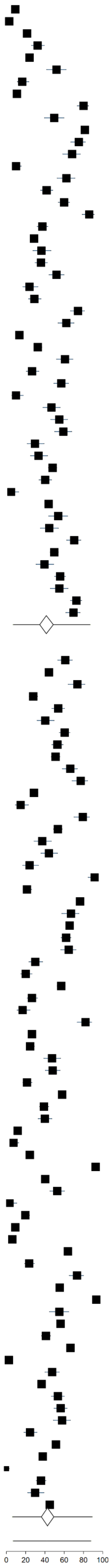

Supplementary Figure 45: Forest plot of risk difference of Long Covid in all included studies with controls with 95% prediction intervals

| study | Dominant source of participants | Type of control | Mean days since infection | Absolute risk difference (%) (95% CI) |
|---|---|---|---|---|
| Al-Aly et al (hospitalized) | hospitalised | influenza patients | 150 | 7.0 ( 6.5, 7.6) |
| Al-Aly et al (non-hospitalized) | community | no positive test result | 126 | 2.9 ( 2.6, 3.1) |
| Ayoubkhani et al | hospitalised | negative test result | 154 | 20.6 ( 20.1, 21.1) |
| Blomberg et al | outpatients | negative test result | 183 | 42.0 ( 29.1, 54.8) |
| Caspersen et al | community | no positive test result | 350 | 16.1 ( 10.6, 21.7) |
| Castro et al | hospitalised | negative test result | 128 | -1.1 ( -1.9, -0.3) |
| Chai et al | hospitalised | other | 378 | 2.5 ( -2.9, 7.9) |
| Cirulli et al | community | negative test result | 90 | 7.7 ( 1.4, 14.1) |
| Havervall et al | healthcare workers | negative test result | 122 | 14.2 ( 9.4, 18.9) |
| Huang et al | hospitalised | no positive test result | 185 | 34.8 ( 31.1, 38.6) |
| Liu et al | hospitalised | negative test result | 196 | 7.2 ( 2.7, 11.7) |
| Logue et al | outpatients | negative test result | 169 | 20.4 ( 6.2, 34.7) |
| Mei et al | hospitalised | no positive test result | 158 | 2.4 ( 0.5, 4.3) |
| ONS study August 2021 | community | negative test result | 84 | 1.6 ( 1.1, 2.1) |
| Radtke et al | community | negative test result | 84 | 1.4 ( -2.2, 5.0) |
| Riestra-Ayora et al | healthcare workers | negative test result | 182 | 26.7 ( 20.5, 32.9) |
| Roessler et al (adults) | community | no positive test result | 91 | 9.2 ( 8.9, 9.6) |
| Roessler et al (children) | community | no positive test result | 91 | 6.1 ( 5.1, 7.2) |
| Soraas et al | community | negative test result | 126 | -0.7 ( -4.6, 3.3) |
| Stephenson et al | community | negative test result | 104 | 13.2 ( 10.9, 15.5) |
| Sudre et al | community | negative test result | 84 | 2.3 ( 1.8, 2.8) |
| Taquet et al | community | influenza patients | 90 | 6.8 ( 6.5, 7.2) |
| Tohamy et al | outpatients | negative test result | 105 | 3.0 ( -2.1, 8.1) |
| Whitaker et al | community | unknown | 84 | 34.6 ( 34.3, 35.0) |
| Xiong et al | hospitalised | no positive test result | 111 | 37.7 ( 31.4, 44.0) |
| Zhou et al | hospitalised | no positive test result | 127 | 55.2 ( 42.5, 67.9) |
| **95% Prediction Interval** | | | | 13.9 ( -16.2, 43.9) |

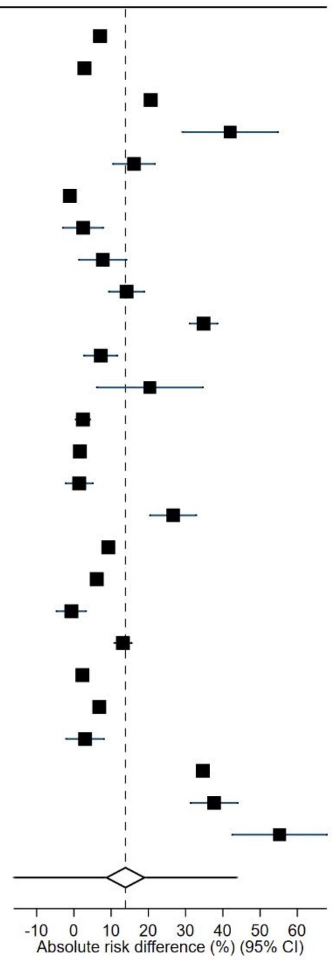

Supplementary Figure 46: Forest plot of relative risk of Long Covid in included studies with community-based samples and controls with 95% prediction intervals

| study | region | type of control | mean days since infection | relative risk (95% CI) |
|---|---|---|---|---|
| Al-Aly et al (non-hospitalized) | N America | no positive test result | 126 | 1.9 ( 1.8, 2.0) |
| Caspersen et al | Europe | no positive test result | 350 | 51.4 ( 36.0, 73.4) |
| Cirulli et al | N America | negative test result | 90 | 2.1 ( 1.3, 3.3) |
| Havervall et al | Europe | negative test result | 122 | 3.0 ( 2.2, 4.0) |
| ONS study August 2021 | Europe | negative test result | 84 | 1.5 ( 1.3, 1.7) |
| Radtke et al | Europe | negative test result | 84 | 1.6 ( 0.6, 4.6) |
| Roessler et al (adults) | Europe | no positive test result | 91 | 1.3 ( 1.3, 1.3) |
| Roessler et al (children) | Europe | no positive test result | 91 | 1.3 ( 1.3, 1.4) |
| Soraas et al | Europe | negative test result | 126 | 1.0 ( 0.9, 1.1) |
| Stephenson et al | Europe | negative test result | 104 | 1.2 ( 1.2, 1.3) |
| Sudre et al | Other | negative test result | 84 | 9.8 ( 5.3, 18.2) |
| Taquet et al | N America | influenza patients | 90 | 1.2 ( 1.2, 1.2) |
| Whitaker et al | Europe | unknown | 84 | 12.3 ( 12.2, 12.5) |
| **95% Prediction Interval** | | | | 2.7 ( 0.2, 39.4) |

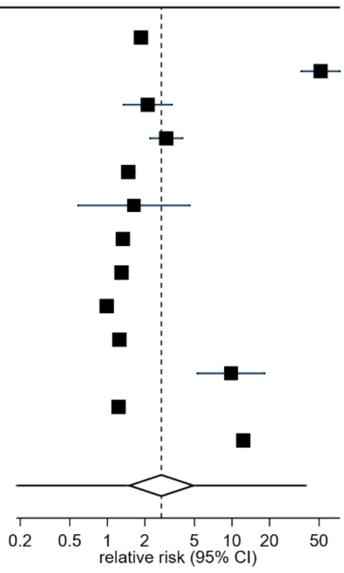

Supplementary Figure 47: Forest plot of risk difference of Long Covid in included studies with community-based samples and controls with 95% prediction intervals

| study | region | type of control | mean days since infection | absolute risk difference (%) (95% CI) |
|---|---|---|---|---|
| Al-Aly et al (non-hospitalized) | N America | no positive test result | 126 | 2.9 ( 2.6, 3.1) |
| Caspersen et al | Europe | no positive test result | 350 | 16.1 ( 10.6, 21.7) |
| Cirulli et al | N America | negative test result | 90 | 7.7 ( 1.4, 14.1) |
| Havervall et al | Europe | negative test result | 122 | 14.2 ( 9.4, 18.9) |
| ONS study August 2021 | Europe | negative test result | 84 | 1.6 ( 1.1, 2.1) |
| Radtke et al | Europe | negative test result | 84 | 1.4 ( -2.2, 5.0) |
| Riestra-Ayora et al | Europe | negative test result | 182 | 26.7 ( 20.5, 32.9) |
| Roessler et al (adults) | Europe | no positive test result | 91 | 9.2 ( 8.9, 9.6) |
| Roessler et al (children) | Europe | no positive test result | 91 | 6.1 ( 5.1, 7.2) |
| Soraas et al | Europe | negative test result | 126 | -0.7 ( -4.6, 3.3) |
| Stephenson et al | Europe | negative test result | 104 | 13.2 ( 10.9, 15.5) |
| Sudre et al | Other | negative test result | 84 | 2.3 ( 1.8, 2.8) |
| Taquet et al | N America | influenza patients | 90 | 6.8 ( 6.5, 7.2) |
| Whitaker et al | Europe | unknown | 84 | 34.6 ( 34.3, 35.0) |
| **95% Prediction Interval** | | | | 10.1 ( -12.7, 32.8) |

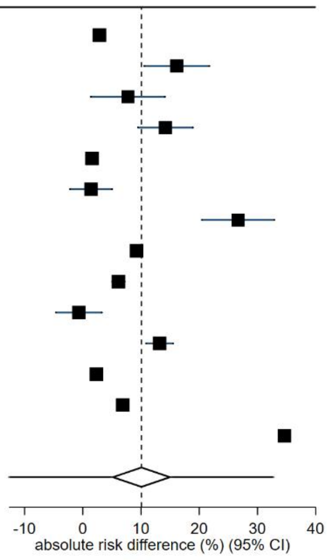

Supplementary Figure 48: Forest plot of risk difference of Long Covid in included studies with community-based samples and controls assessed as having low risk of bias with 95% prediction intervals

| study | region | type of control | mean days since infection | absolute risk difference (%) (95% CI) |
|---|---|---|---|---|
| ONS study August 2021 | Europe | negative test result | 84 | 1.6 ( 1.1, 2.1) |
| Radtke et al | Europe | negative test result | 84 | 1.4 ( -2.2, 5.0) |
| Roessler et al (adults) | Europe | no positive test result | 91 | 9.2 ( 8.9, 9.6) |
| Roessler et al (children) | Europe | no positive test result | 91 | 6.1 ( 5.1, 7.2) |
| **95% Prediction Interval** | | | | 4.8 ( -13.2, 22.7) |

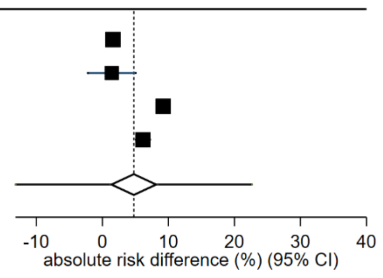

Supplementary Figure 49: Contour-enhanced funnel plot

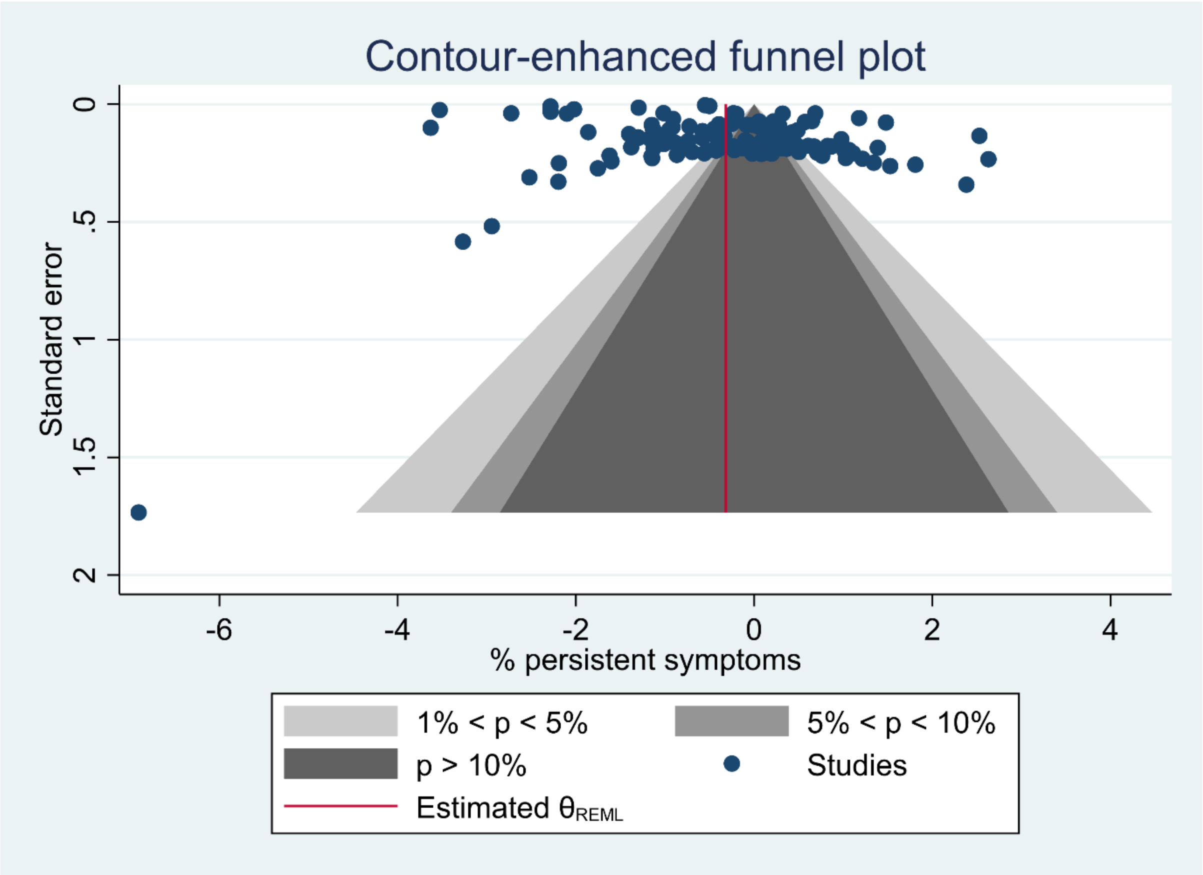

Supplementary Figure 50: Forest plot of prevalence of Long Covid in the included studies by study design with 95% prediction intervals

| study | region | mean days since infection | cases | total | % persistent symptoms |
|---|---|---|---|---|---|
| **cohort** | | | | | |
| Abdelrahman et al | Other | 270 | 105 | 172 | 61.0 (53.3, 68.4) |
| Al-Aly et al (hospitalized) | N America | 150 | 1091 | 11800 | 9.2 (8.7, 9.8) |
| Al-Aly et al (non-hospitalized) | N America | 126 | 1718 | 60255 | 2.9 (2.7, 3.0) |
| Aminian et al | N America | 243 | 1255 | 2839 | 44.2 (42.4, 46.1) |
| Arnold et al | Europe | 90 | 81 | 110 | 73.6 (64.4, 81.6) |
| Augustin et al | Europe | 131 | 123 | 442 | 27.8 (23.7, 32.3) |
| Ayoubkhani et al | Europe | 154 | 6085 | 28335 | 21.5 (21.0, 22.0) |
| Bellan et al | Europe | 120 | 128 | 238 | 53.8 (47.2, 60.2) |
| Blanco et al | Europe | 104 | 52 | 100 | 52.0 (41.8, 62.1) |
| Bliddal et al | Europe | 84 | 52 | 129 | 40.3 (31.8, 49.3) |
| Blomberg et al | Europe | 183 | 189 | 312 | 60.6 (54.9, 66.0) |
| Boscolo-Rizzo et al | Europe | 365 | 161 | 304 | 53.0 (47.2, 58.7) |
| COMEBAC study | Europe | 127 | 244 | 478 | 51.0 (46.5, 55.6) |
| Carrillo-Garcia et al | Europe | 105 | 100 | 151 | 66.2 (58.1, 73.7) |
| Caruso et al | Europe | 182 | 91 | 118 | 77.1 (68.5, 84.3) |
| Caspersen et al | Europe | 350 | 28 | 170 | 16.5 (11.2, 22.9) |
| Castro et al | N America | 128 | 721 | 6619 | 10.9 (10.2, 11.7) |
| Chai et al | China | 378 | 156 | 546 | 28.6 (24.8, 32.6) |
| Cirulli et al | N America | 90 | 18 | 122 | 14.8 (9.0, 22.3) |
| Clavario et al | Europe | 121 | 160 | 200 | 80.0 (73.8, 85.3) |
| Cristillo et al | Europe | 196 | 50 | 101 | 49.5 (39.4, 59.6) |
| Diaz-Fuentes et al | N America | 84 | 88 | 111 | 79.3 (70.5, 86.4) |
| Domenech-Montoliu et al | Europe | 213 | 258 | 483 | 53.4 (48.9, 57.9) |
| Erol et al | Other | 170 | 45 | 121 | 37.2 (28.6, 46.4) |
| Fernandez-de-Las-Penas et al | Europe | 227 | 930 | 1142 | 81.4 (79.1, 83.7) |
| Froidure et al | Europe | 95 | 73 | 107 | 68.2 (58.5, 76.9) |
| Garcia-Abellan et al | Europe | 182 | 28 | 116 | 24.1 (16.7, 33.0) |
| Gonzalez-Hermosillo et al | Other | 105 | 119 | 130 | 91.5 (85.4, 95.7) |
| Han et al | China | 175 | 71 | 114 | 62.3 (52.7, 71.2) |
| Havervall et al | Europe | 122 | 69 | 323 | 21.4 (17.0, 26.2) |
| Huang et al | China | 186 | 1265 | 1655 | 76.4 (74.3, 78.5) |
| Jacobson et al | N America | 119 | 79 | 118 | 66.9 (57.7, 75.3) |
| Kashif et al | Other | 105 | 101 | 242 | 41.7 (35.5, 48.2) |
| Li et al | China | 120 | 173 | 289 | 59.9 (54.0, 65.6) |
| Liao et al | China | 104 | 122 | 142 | 85.9 (79.1, 91.2) |
| Liao et al | China | 409 | 113 | 303 | 37.3 (31.8, 43.0) |
| Logue et al | N America | 169 | 53 | 177 | 30.0 (23.3, 37.3) |
| Lucidi et al | Europe | 186 | 40 | 110 | 36.4 (27.4, 46.1) |
| Lui et al | China | 96 | 41 | 204 | 20.1 (14.8, 26.3) |
| Martinez et al | Europe | 168 | 69 | 260 | 26.5 (21.3, 32.3) |
| Matteudi et al | Europe | 180 | 23 | 137 | 16.8 (11.0, 24.1) |
| Mazza et al | Europe | 104 | 81 | 226 | 35.8 (29.6, 42.5) |
| Mei et al | China | 158 | 976 | 3677 | 26.5 (25.1, 28.0) |
| Menges et al | Europe | 219 | 106 | 431 | 24.6 (20.6, 28.9) |
| Milanese et al | Europe | 196 | 64 | 135 | 47.4 (38.8, 56.2) |
| Millet et al | N America | 365 | 83 | 173 | 48.0 (40.3, 55.7) |
| Mohiuddin et al | Other | 161 | 67 | 313 | 21.4 (17.0, 26.4) |
| Munblit et al | Europe | 232 | 1534 | 2649 | 57.9 (56.0, 59.8) |
| Nabahati et al | Other | 104 | 90 | 173 | 52.0 (44.3, 59.7) |
| Nehme et al | Europe | 243 | 160 | 410 | 39.0 (34.3, 43.9) |
| Nunez-Fernandez et al | Europe | 98 | 58 | 200 | 29.0 (22.8, 35.8) |
| ONS study August 2021 | Europe | 84 | 2501 | 21374 | 11.7 (11.3, 12.1) |
| Ong et al | Other | 104 | 13 | 175 | 7.4 (4.0, 12.4) |
| Osmanov et al | Europe | 270 | 126 | 519 | 24.3 (20.6, 28.2) |
| PHOSP-COVID study | Europe | 174 | 797 | 861 | 92.6 (90.6, 94.2) |
| Peghin et al | Europe | 191 | 241 | 599 | 40.2 (36.3, 44.3) |
| Peluso et al | N America | 112 | 89 | 143 | 62.2 (53.8, 70.2) |
| Petersen et al | Europe | 125 | 95 | 180 | 52.8 (45.2, 60.2) |
| Qin et al | China | 105 | 87 | 647 | 13.4 (10.9, 16.3) |
| Qu et al | China | 105 | 176 | 540 | 32.6 (28.7, 36.7) |
| Radtke et al | Europe | 84 | 4 | 109 | 3.7 (1.0, 9.1) |
| Rass et al | Europe | 90 | 82 | 135 | 60.7 (52.0, 69.0) |
| Riestra-Ayora et al | Europe | 182 | 52 | 195 | 26.7 (20.6, 33.5) |
| Righi et al | Europe | 84 | 83 | 421 | 19.7 (16.0, 23.8) |
| Roessler et al (adults) | Europe | 91 | 13396 | 145184 | 9.2 (9.1, 9.4) |
| Roessler et al (children) | Europe | 91 | 734 | 11950 | 6.1 (5.7, 6.6) |
| Romero-Duarte et al | Europe | 196 | 509 | 797 | 63.9 (60.4, 67.2) |
| Sathyamurthy et al | Other | 104 | 66 | 279 | 23.7 (18.8, 29.1) |
| Seeßle et al | Europe | 147 | 107 | 146 | 73.3 (65.3, 80.3) |
| Shang et al | China | 196 | 441 | 796 | 55.4 (51.9, 58.9) |
| Sibila et al | Europe | 116 | 98 | 172 | 57.0 (49.2, 64.5) |
| Sigfrid et al | Europe | 222 | 305 | 327 | 93.3 (90.0, 95.7) |
| Simani et al | Other | 197 | 12 | 120 | 10.0 (5.3, 16.8) |
| Skala et al | Europe | 91 | 56 | 102 | 54.9 (44.7, 64.8) |
| Skjorten et al | Europe | 105 | 59 | 126 | 46.8 (37.9, 55.9) |
| Sonnweber et al | Europe | 103 | 73 | 133 | 54.9 (46.0, 63.5) |
| Soraas et al | Europe | 126 | 380 | 676 | 56.2 (52.4, 60.0) |
| Stephenson et al | Europe | 104 | 2038 | 3065 | 66.5 (64.8, 68.2) |
| Sudre et al | Other | 84 | 108 | 4182 | 2.6 (2.1, 3.1) |
| Sykes et al | Europe | 127 | 75 | 127 | 59.1 (50.0, 67.7) |
| Taquet et al | N America | 90 | 100007 | 273618 | 36.5 (36.4, 36.7) |
| Tarsitani et al | Europe | 105 | 34 | 115 | 29.6 (21.4, 38.8) |
| Tawfik et al | Other | 91 | 40 | 120 | 33.3 (25.0, 42.5) |
| Taylor et al | Europe | 126 | 261 | 545 | 47.9 (43.6, 52.2) |
| Tleyjeh et al | Other | 136 | 125 | 222 | 56.3 (49.5, 62.9) |
| Todt et al | Other | 105 | 96 | 239 | 40.2 (33.9, 46.7) |
| Tohamy et al | Other | 105 | 5 | 100 | 5.0 (1.6, 11.3) |
| Townsend et al | Europe | 86 | 74 | 128 | 57.8 (48.8, 66.5) |
| Venturelli et al | Europe | 105 | 394 | 767 | 51.4 (47.8, 55.0) |
| Walle-Hansen et al | Europe | 200 | 57 | 106 | 53.8 (43.8, 63.5) |
| Weng et al | China | 104 | 52 | 117 | 44.4 (35.3, 53.9) |
| Xiong et al | China | 111 | 267 | 538 | 49.6 (45.3, 53.9) |
| Xiong et al | China | 167 | 114 | 162 | 70.4 (62.7, 77.3) |
| Yan et al | China | 379 | 47 | 119 | 39.5 (30.7, 48.9) |
| Yan et al | China | 98 | 0 | 125 | 0.0 (0.0, 2.9) |
| Yin et al | China | 203 | 176 | 316 | 55.7 (50.0, 61.3) |
| Zayet et al | Europe | 289 | 127 | 354 | 35.9 (30.9, 41.1) |
| Zhan et al | China | 348 | 36 | 121 | 29.8 (21.8, 38.7) |
| Zhang et al | China | 106 | 67 | 122 | 54.9 (45.7, 63.9) |
| Zhang et al | China | 105 | 178 | 245 | 72.7 (66.6, 78.1) |
| Zhang et al | China | 378 | 1095 | 2433 | 45.0 (43.0, 47.0) |
| Zhou et al | China | 127 | 114 | 164 | 69.5 (61.9, 76.5) |
| | | | | | **41.3 (6.0, 88.6)** |
| **cross-sectional** | | | | | |
| Baricich et al | Europe | 139 | 66 | 204 | 32.4 (26.0, 39.2) |
| Becker et al | N America | 231 | 178 | 740 | 24.1 (21.0, 27.3) |
| Frija-Masson et al | Europe | 91 | 103 | 137 | 75.2 (67.1, 82.2) |
| Fu et al | China | 196 | 20 | 199 | 10.1 (6.2, 15.1) |
| Gaber et al | Europe | 122 | 61 | 138 | 44.2 (35.8, 52.9) |
| Kim et al | Other | 195 | 591 | 900 | 65.7 (62.5, 68.8) |
| Lemhofer et al | Europe | 91 | 226 | 365 | 61.9 (56.7, 66.9) |
| Liu et al | China | 196 | 373 | 1301 | 28.7 (26.2, 31.2) |
| Liyanage-Don et al | N America | 113 | 99 | 153 | 64.7 (56.6, 72.3) |
| Maestre-Muniz et al | Europe | 379 | 309 | 543 | 56.9 (52.6, 61.1) |
| Mechi et al | Other | 274 | 92 | 112 | 82.1 (73.8, 88.7) |
| Nguyen et al | Europe | 213 | 30 | 125 | 24.0 (16.8, 32.5) |
| O'Keefe et al | N America | 119 | 79 | 198 | 39.9 (33.0, 47.1) |
| Orru et al | Europe | 91 | 113 | 152 | 74.3 (66.6, 81.1) |
| Stavem et al | Europe | 117 | 185 | 451 | 41.0 (36.4, 45.7) |
| Taboada et al | Europe | 189 | 87 | 183 | 47.5 (40.1, 55.0) |
| Tempany et al | Europe | 84 | 116 | 217 | 53.5 (46.6, 60.2) |
| Trunfio et al | Europe | 194 | 41 | 168 | 24.4 (18.1, 31.6) |
| Ursini et al | Europe | 182 | 270 | 616 | 43.8 (39.9, 47.9) |
| Whitaker et al | Europe | 84 | 28713 | 76155 | 37.7 (37.4, 38.0) |
| | | | | | **45.9 (11.2, 85.1)** |

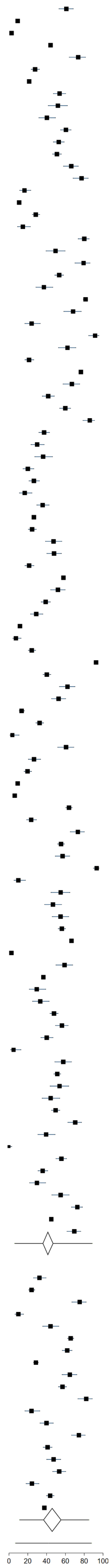
